# Socioeconomic inequities within and between cities in objectively measured green space qualities at small geographical scales: Evidence from Australia

**DOI:** 10.1101/2025.04.09.25325554

**Authors:** Lauren Del Rosario, Thomas Astell-Burt, Michael Navakatikyan, Jonathan R Olsen, Fiona Caryl, Brenda Lin, Bin Jalaludin, Evelyne de Leeuw, Richard Mitchell, Xiaoqi Feng

**Affiliations:** School of Architecture, Design and Planning, University of Sydney, Sydney, Australia; School of Population Health, Faculty of Medicine and Health, University of New South Wales, Sydney, Australia; Population Wellbeing and Environment Research Lab (PowerLab), Sydney, NSW, Australia; School of Health and Society, Faculty of Arts, Social Sciences, and Humanities, University of Wollongong, Wollongong, Australia; Westmead Applied Research Centre, Sydney Medical School, Faculty of Medicine and Health, The University of Sydney, 176 Hawkesbury Road, Westmead, NSW 2145, Australia; Charles Perkins Centre, The University of Sydney, Johns Hopkins Drive, Camperdown, NSW 2050, Australia; MRC/CSO Social and Public Health Sciences Unit, University of Glasgow, Clarice Pears Building, 90 Byres Road, UK; Institute for Social Science Research, The University of Queensland, Brisbane, Australia; CSIRO, Land & Water Ecosystem Sciences Precinct 41 Boggo Road Dutton Park, QLD, AU 4102; École de santé publique de l’Université de Montréal, Montreal, QC, Canada; Cities Institute, Faculty of Arts, Design and Architecture, University of New South Wales, Sydney, Australia; The George Institute for Global Health, Level 18, International Towers 3, 300 Barangaroo Ave, Sydney, NSW 2000, Australia

**Keywords:** Green space, Parks, Inequity, Health, Tree cover, Biodiversity

## Abstract

**Objective:** To determine the extent of inequitable distributions in green space qualities in urban areas of Australia.

**Method:** Existing data from the cities of Sydney, Newcastle, and Wollongong in Australia was used to define green space qualities relating to accessibility, amenities/activities, beaches/coastline, biodiversity, incivilities, landcover and land use. Green space qualities were measured within multiple-scale network distance buffers for residential mesh blocks and linked with the Australian Bureau of Statistics Index of Relative Socio-economic Disadvantage (IRSD). Correlations were analysed using Spearman’s rank correlation coefficient between IRSD score (reversed; higher scores are more disadvantaged) and green space qualities aggregated over mesh blocks. Influence of IRSD, population density and random effects of population structures were examined using single-level and multilevel models. Spatial patterns and clusters were identified through choropleth maps and hot spot analyses.

**Results:** At the 1600m scale, more disadvantaged areas tended to have green spaces with lower percentages of nearby street trees to roads (Rho=-0.52, p≤0.001), lower percentages of slope >6° (Rho=-0.49), lower likelihood of threatened mammal species/habitat occurrences (Rho=-0.47), and lower percentages of tree canopy (Rho=-0.46). More disadvantaged areas tended to have green spaces with higher percentages of open grass (Rho=0.38, p≤0.001) and bare earth (Rho=0.33, p≤0.001) and higher densities of robberies (Rho=0.34, p≤0.001). For selected qualities, multilevel models tended to support the relationships that were found using Spearman’s rank correlation.

**Discussion:** Socioeconomic inequities in tree canopy, biodiversity and incivilities are present for green spaces in large and mid-sized Australian cities.

## Background

The health benefits of green space are clearly evident (Hartig et al., 2014, Markevych et al., 2017, Astell-Burt et al., 2022, Zhou et al., 2024, Zhang and Luo, 2025). Inequitable distribution of qualities such as those pertaining to access, amenities, biodiversity, incivilities, landcover and land use, that make green spaces attractive places to visit and live near could lead to poorer health outcomes for disadvantaged populations.

Prior research has employed direct observation audits to evaluate the quality of green spaces (e.g. Crawford et al., 2008, Vaughan et al., 2013, Bruton and Floyd, 2014, Hoffimann et al., 2017, Vidal et al., 2021, Zhang et al., 2021). Studies from the US, Australia and Europe have revealed that more disadvantaged areas typically have poorer quality green spaces, e.g. fewer playgrounds per park (Vaughan et al., 2013), fewer sporting activities, (Vidal et al., 2021), dirtier sitting areas (Bruton and Floyd, 2014), fewer amenities (Crawford et al., 2008, Hoffimann et al., 2017), and fewer water features, paths and lighting (Crawford et al., 2008). However, in Hong Kong, amenities, safety and aesthetics were not related with disadvantage or ethnic minorities, but more active facilities were present in areas with moderate to high disadvantage (Zhang et al., 2021).

Since direct observation audits are time intensive, studies have also analysed green space qualities using objective data (You, 2016, Zhang, 2023, Lin et al., 2015, Li et al., 2025), with some studies examining results across several cities/regions (Rigolon et al., 2018, Fossa et al., 2023, Viinikka et al., 2023, Heo and Bell, 2023). Relying on a composite index of park quality, more disadvantaged areas across 99 U.S. cities had poorer quality green spaces (Rigolon et al., 2018). Using landcover data across three U.S. cities, Fossa et al. (2023) found higher tree coverage in higher socioeconomic areas. In Nanjing, raster and vector databases from government agencies and street-view imagery have been used to assess green space quality (Zhang, 2023). Using objective data in Shenzhen, You (2016) found that green space quality was lower in higher socioeconomic areas but access was lower in disadvantaged areas. In Finland, areas of higher socioeconomic status were closer to green spaces larger than 10ha and forests, whereas lower socioeconomic areas had access to green spaces with more recreational facilities and routes for jogging, skiing, walking, hiking and biking (Viinikka et al., 2023). Studies that did not employ direct observation audits tended to analyse fewer qualities.

Knobel et al.’s (2019) systematic review found that quality domains such as surroundings, land cover, and biodiversity are underrepresented in existing multi-dimensional, direct observation, urban green space quality assessment tools. The review led to the development of the RECITAL tool (Knobel et al., 2021), from which we derived most of our quality domains, or groups of related green space qualities. The qualities reviewed by Knobel et al. (2019) are derived from in-person surveys and often only at city or council-level within cities. Little research exists in quantifying the quality of green spaces objectively over large geographic extents. There are no studies that have comprehensively combined approximately 100 objectively measured green space qualities across a large geographical area, and explored variation by area-level socioeconomic status. We aim to fill this gap by using objective, mapped data with which we find associations with socioeconomic disadvantage. This is important as the varying qualities may mitigate against increasing temperatures and provide health benefits within urban areas. The research has practical applications in terms of mapping gaps in provision to address inequalities.

## Methods

### Setting

The study area includes the three coastal cities of Sydney, Newcastle, and Wollongong, in New South Wales (NSW), Australia. Australian Bureau of Statistics (ABS) Urban Centre and Localities (UCLs) were used as the boundary for the three cities. There are diverse experiences within these cities because of differences in size and structure. Sydney is a large city with approximately 5 million people. Newcastle and Wollongong have populations less than 1 million.

### ABS data

Analysis was undertaken using ABS mesh blocks for 2021 – the smallest geographic units of the Australian Statistical Geography Standard (ABS, 2021). Sydney has 41,318 residential mesh blocks, Newcastle has 3443 and Wollongong has 2820. Mesh blocks contain approximately 30-60 dwellings (ABS, 2021). Multiple-scale network buffers (100m, 200m, 300m, 400m, 800m and 1600m) were created around residential mesh block centroids which were joined with intersecting green spaces. See Supplementary Material S2 for more detail on network buffers.

Socio-economic Indexes for Areas (SEIFA) Index of Relative Socio-economic Disadvantage (IRSD)(ABS, 2023b) was used to determine area-level socioeconomic status. The finest scale at which IRSD data is released is the Statistical Area Level 1 (SA1), which is one level higher than the mesh block. SA1s typically have a population between 200-800 people (ABS, 2021b). Mesh blocks were joined to IRSD data based on their respective SA1. Residential mesh blocks that had no IRSD data (n=43) due to nil/nominal population, for example, were excluded from further analysis. Reversed quintiles for maps and IRSD scores for correlation analysis were calculated based on the respective SA1’s decile and score respectively against the rest of NSW. The IRSD indices were reversed so that higher scores denoted more disadvantaged areas.

For multilevel modelling, mesh blocks, Statistical Area Level 2 (SA2), Statistical Area Level 3 (SA3) and Statistical Area Level 4 (SA4) were used to control for population density effects and nested geographies. Mesh blocks are nested in SA1s, which are nested in SA2s and so on up to SA4s (ABS 2021a). SA2s tend to have populations of 3,000–25,000 and SA4s tend to have populations >100,000 (ABS 2021a).

### Green space data

For Sydney, the NSW Department of Planning, Housing and Infrastructure’s Existing Green Assets dataset was used and included green spaces from various categories: bushland, cemeteries, civic, community purpose, golf courses, heritage and cultural, operational, parks and gardens, Office of Strategic Lands areas, special purpose areas, sporting areas, undeveloped or unspecified, and waterfront areas. A 5km buffer was created around the Sydney UCL to reduce the impact of edge effects. Multipart polygons were split into singlepart, due to the presence of some multiple separate green spaces extending over 20km (Figure S1). Large green spaces (>10km^2^) located outside of the UCL^1^ and classified as bushland were removed since access is likely to be more difficult than other green spaces. Small green spaces (<5m^2^) were removed.

The coverage of the Existing Green Assets dataset did not extend to Newcastle and Wollongong. ABS 2021 mesh blocks categorised as “Parkland” were used for green space locations within a 5km buffer of Newcastle and Wollongong UCLs. Large green spaces (>10km^2^) located outside of the UCL were removed. As mesh blocks are categorised based on majority land uses, there may be areas within “Parkland” mesh blocks that are not green spaces and vice versa.

Green space qualities data is summarised in Table S1. Table S2 summarises the qualities we considered. These qualities were defined in broad alignment to existing tools for subjectively rating green spaces (e.g., Knobel et al., 2021) where robust objective spatial data were available. Access, amenities, incivilities, land covers, animal biodiversity and birds biodiversity domains were included in Knobel et al. (2021). Due to the coastal nature of the three cities included in this study, we decided to also include a beaches/coastline domain since blue spaces may also be beneficial for health and wellbeing (White et al., 2020) and thermal cooling (Liu and Li, 2025, Yang et al., 2025). Knobel et al. (2021) also included a domain for surroundings, however because of data availability, we chose to include land use data and tree canopy data rather than characteristics such as building facades.

### Quality domain 1: Access

Pathways (PSMA, Geoscape Australia) and bikeways (Transport for NSW) were intersected with green space locations to determine length of paths and bikeways per green space. The GeoVision trees dataset (Precisely) was intersected with pathways and bikeways to determine length of paths and bikeways with trees overhead. A 5m Digital Elevation Model (Geoscience Australia) was used to determine the length of pathways with gradient >6° (steep hills; Trail Hiking Australia, 2024). Public transport access qualities were determined from the number of stops (Transport for NSW) within a 100m radius of green spaces.^2^ Length of main roads (PSMA, Geoscape Australia) within a 100m radius of green spaces was determined.

### Quality domain 2: Amenities/Activities

The number of public toilets (The National Public Toilet Map, Department of Health and Aged Care), cafés, hotels/bars, restaurants/takeaways, and supermarkets/greengrocers (Sensis) were counted within 100m radii around green spaces. The presence of free 5km parkrun events were also included (http://www.parkrun.com.au/events/).

### Quality domain 3: Beaches/Coastline

The area of beaches (Geofabrik and OpenStreetMap and contributors) that intersect with green spaces and coastline presence (ABS 2021) were evaluated.

### Quality domain 4: Biodiversity

Likely occurrences of threatened flora and fauna species/habitat (Species of National Environmental Significance Distributions, Australian Government Department of Climate Change, Energy, the Environment and Water) were intersected with green spaces. Areas of biodiversity value (Biodiversity Values Map version 16.4, NSW Government Department of Climate Change, Energy, the Environment, Water) that intersect with green spaces were evaluated. See Table S2 for more information.

We attempted to approximate taxonomic diversity of broad categories (i.e. birds, fish, flora, frogs, mammals, reptiles and other animals) using the Shannon Diversity Index and Simpson Diversity Index. While these indices typically rely on data for individuals, in the absence of data for individuals, the index was applied using the number of species^3^ within each category. For formulae see Table S2.

### Quality domain 5: Incivilities

Density of alcohol-related assaults, malicious damage, non-domestic violence assault, robbery, and stealing from persons (Bureau of Crime Statistics and Research (BoCSAR)) were intersected with green spaces. Crime density for each type were coded from zero to three based on the coding in the BoCSAR data (0 = no value, 1 = “Low Density”, 2 = “Medium Density”, 3 = “High Density”). It is noted that incivility data is not green space-specific and individual green spaces may or may not have had the particular incivilities occurring within them. See Table S2 for more information.

### Quality domain 6: Land cover

GeoVision land cover data at 2m resolution (Precisely) was analysed for summer (December 2020). Green space land cover areas were identified for buildings, built-up areas, bare earth, grass, other vegetation, roads, swimming pools, trees, water, and shadows.

### Quality domain 7: Land use

Surrounding land uses within 100m of green space were determined (ABS 2021 Mesh blocks). Area and percentage of tree canopy (GeoVision, Precisely) surrounding green spaces was also calculated, including separate analysis for tree heights ≤4m and >4m, assuming that trees >4m would provide more shade. Percentage of tree canopy to roads surrounding green space was also calculated by creating a 10m buffer around all roads (PSMA, Geoscape Australia). Trees that intersected with the buffered roads dataset were used to determine percentage of tree canopy to roads. This was used as a proxy for street trees.

### Aggregating green space qualities to mesh blocks

An “intersect area” is the area that the green space intersects with the mesh block’s buffer. We aggregated green space qualities of intersected green spaces to mesh blocks. We consider a set of intersects within mesh blocks as an “aggregated green space”. Various methods of aggregation were developed with variables presented either as a weighted sum (“w0 Sum”), a weighted mean (“w1 Mean”), a maximum (“Max”), presence/absence (“Quality”) or were derived using qualities that have already been aggregated as weighted sums or weighted means (“Formula”). See Supplementary Material S4 for greater detail on aggregation methods.

Some green spaces had inadequate tree cover data (qualities using tree cover data in access and land use sections), slope data and/or land use data. For tree cover and slope data, this was coded based on observations with other datasets.^4^ For land cover data, if the total of all land cover categories used in this research was below 80% of the green space area, land cover data was considered inadequate. If there was inadequate data for one of the intersect areas, the whole mesh block was considered to have missing data and was not included in statistical analysis for the respective qualities. Mesh blocks containing “NA” for any variable accounted for less than 10% of the sample at 1600m.

### Statistical Analysis

Associations between socioeconomic status (IRSD quintiles) and aggregated green space qualities at mesh block level were examined using Spearman’s rank correlation coefficient.

Population effects and nested geographies were controlled for. For 1600m buffers, single-level and multilevel models with up to four levels for the random intercepts associated with geographical areas (mesh block, Statistical Area Level 2 (SA2), Statistical Area Level 3 (SA3) and Statistical Area Level 4 (SA4)) were fitted with a Markov Chain Monte Carlo (MCMC)(Browne, 2023) estimation in MLwiN v3.05 (Rasbash et al., 2023).

Eight models were developed. Model 1 (intercept only) and Model 2 (intercept + IRSD) were without higher random effects. Model 3 (intercept + RE(SA2)), Model 4 (intercept + IRSD + RE(SA2)), and Model 5 (intercept + IRSD + Pop(SA2) + RE(SA2)) were two-level models testing random effects with SA2. Model 5 included population density of SA2s. Model 6 (intercept + RE(SA2, SA3, SA4)), Model 7 (intercept + IRSD + RE(SA2, SA3, SA4)) and Model 8 (intercept + IRSD + population(SA2, SA3, SA4) + RE(SA2, SA3, SA4)) were four-level models, testing random effects with SA2, SA3 and SA4. Model 8 also included population density of SA2s, SA3s and SA4s.

Models were built covering ten qualities: percentage of tree canopy to roads surrounding green space; percentage of green space >6°; percentage of tree canopy; area of open grass; percentage of open grass; percentage of bare earth; mean level of incivilities; mammal species/habitat; total species/habitat; and Shannon Diversity Index. For qualities with a negative Rho, the threshold was ≤-0.39. Since qualities with a positive Rho were weaker, the threshold was ≥0.30. However, some qualities that met the threshold requirements were excluded as they were very similar to other variables – they were highly correlated with an included variable (rho >=0.80)(Table S13). Excluded qualities included percentage of tree canopy >4m to roads surrounding green space, percentage of tree canopy >4m surrounding green space, percentage of tree canopy surrounding green space, density of malicious damage and density of robberies.

The Deviance Information Criterion (DIC) was used to compare the relative fits of the models, as it is the main index of fit for MCMC modelling in MLwiN software. The smaller the DIC, the better the model fit with all other criteria.

### Spatial analysis

Hotspot analysis using the Getis Ord Gi* Statistic was undertaken using ArcMap 10.8.2 (Esri, Redlands, CA) to determine if green space qualities were spatially clustered, for example, if mesh blocks with high/low values of qualities were locally concentrated. Spatial weights matrices were used with the eight nearest neighbours chosen to conceptualise the spatial relationships (Singh, 2018). A correction for a false discovery rate was applied. The hot spot analysis produces the following thresholds for ‘hot’ and ‘cold’ spots: 90%, 95% and 99% confidence. Choropleth maps were also created for various green space qualities using QGIS 3.30.3 (QGIS Development Team, Gossau, Zürich).

## Results

### Disadvantage

In Sydney, there was a general trend of low disadvantage in Sydney’s northern, north-west, east, Sutherland Shire and Blue Mountains (Figure 1). Areas of higher disadvantage included western and south-western Sydney. In Newcastle, areas of high disadvantage were scattered across the city. In Wollongong, areas of low disadvantage tended to be in the north-west.

**Figure 1.**
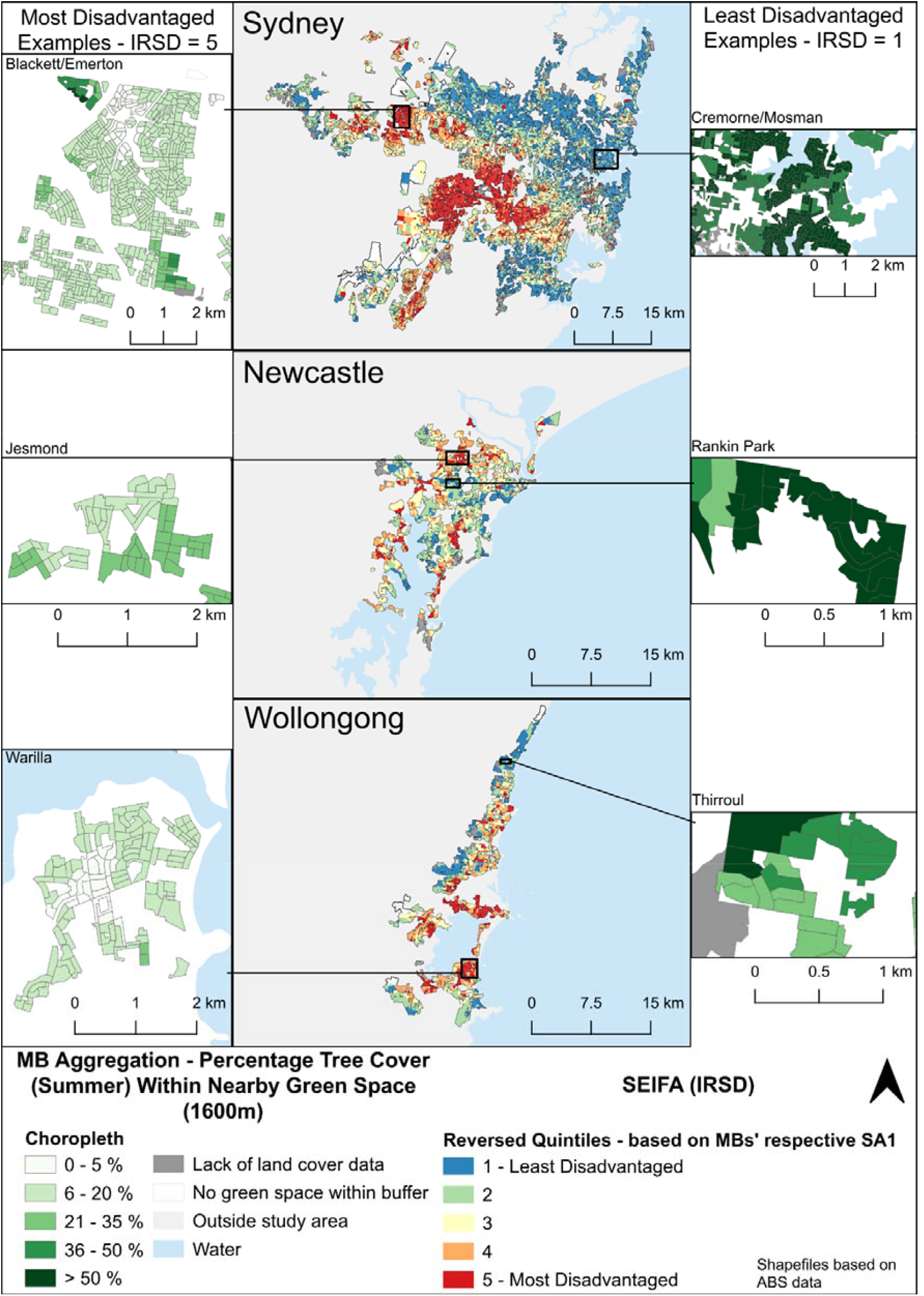
SEIFA (IRSD – reversed quintiles) with inset maps of examples of least disadvantaged mesh blocks and most disadvantaged mesh blocks showing Percentage Tree Cover Within Nearby Green Space (1600m network distance) – Aggregation for residential mesh blocks with SEIFA (IRSD) data. Shapefiles based on ABS data.

### Choropleth and hot spot mapping

In Sydney, mesh blocks with the highest percentage of tree cover in neighbouring green spaces were in the northern and southern areas and the Blue Mountains (Figure 2, Figure S8). These areas tend to be less disadvantaged when compared with IRSD reversed quintiles (Figure 1). Cold spots, with statistically significant clusters of low percentage tree cover, mainly occurred in Western and South-Western Sydney and Sydney’s centre^5^. In Newcastle, cold spots of percentage tree cover were most apparent in parts of the north. In Wollongong, cold spots were largely along the coast.

**Figure 2.**
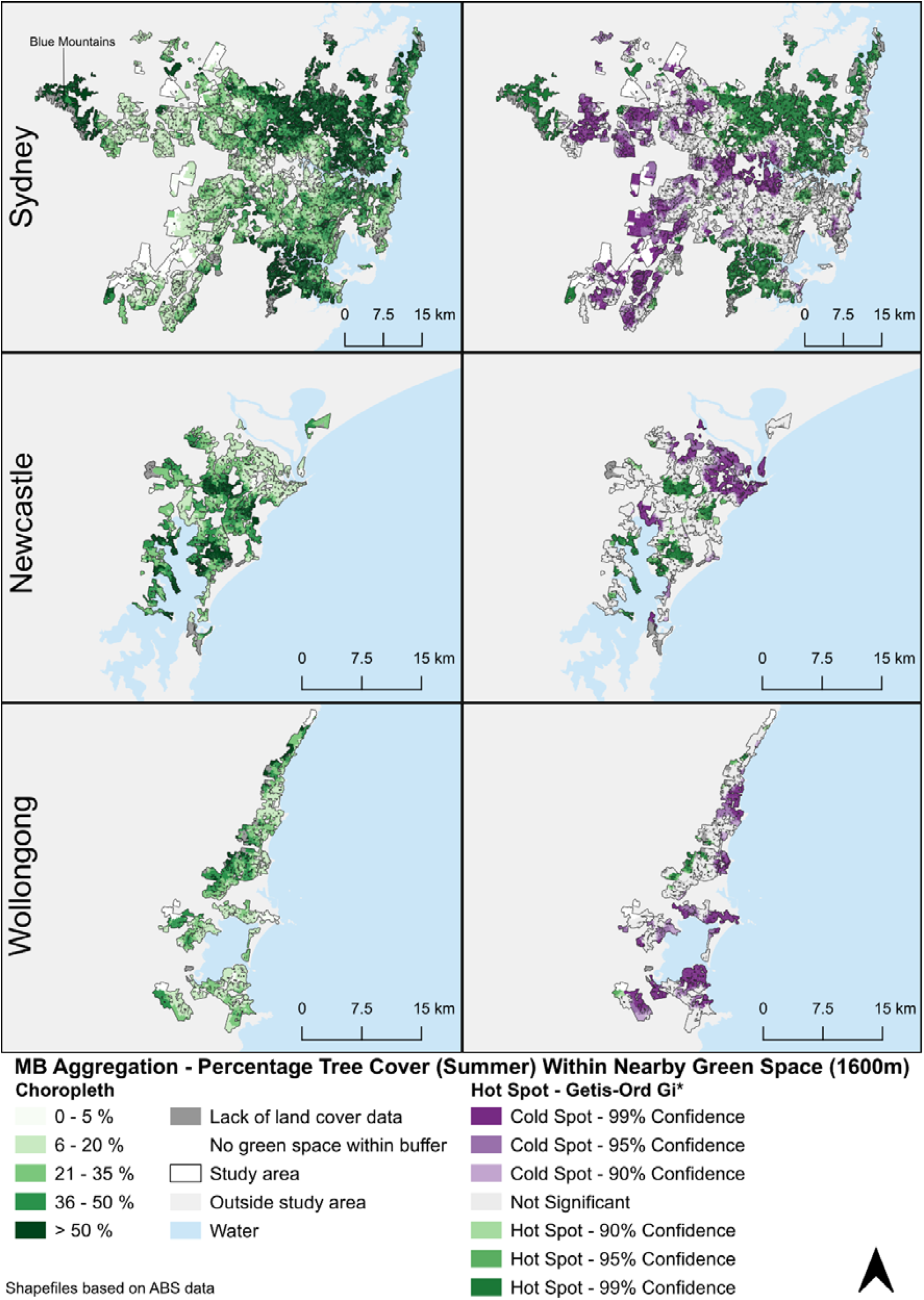
Percentage Tree Cover Within Nearby Green Space (1600m network distance) – Aggregation for residential mesh blocks that contain SEIFA data. Shapefiles based on ABS data.

Examples of mesh blocks that were most disadvantaged (reversed IRSD quintile = 5) and least disadvantaged (reversed IRSD quintile = 1) are illustrated in Figure 1 for 1600m buffers. At 1600m, least disadvantaged mesh block examples shown for Sydney and Newcastle had a majority of example mesh blocks >36% tree cover and the presence of example mesh blocks >36% for Thirroul in Wollongong. Percentage tree cover of green spaces for the most disadvantaged mesh block examples in Sydney’s West ranged from 0-5% to >50%, with most 6-20%. Percentage tree cover ranged from 6-20% to 21-35% for the most disadvantaged mesh blocks in Jesmond in Newcastle. The majority of most disadvantaged mesh blocks in Warilla had a percentage tree cover aggregation of 6-20%. Other mesh blocks that are most disadvantaged or least disadvantaged across the three cities may not have the same patterns for percentage tree cover aggregation as these examples.

### Negative correlations between green space qualities and socioeconomic disadvantage

For the full set (all three cities together) using 400m buffers, more disadvantaged areas tended to have lower likelihood of threatened mammal species/habitat (Rho=-0.51, p-value≤0.001) and total threatened species/habitat (Rho=-0.48, p-value≤0.001), lower percentages of nearby street trees to roads (Rho=-0.44, p-value≤0.001), nearby street trees >4m to roads (Rho=-0.43, p-value≤0.001), and lower percentages of green spaces with slopes >6° (Rho=-0.42, p-value≤0.001) (Table 1; for p-values in full see Table S6-S8; for averages and standard errors see Table S9-S11.).

**Table 1.** Spearman’s rank coefficient of correlation between SEIFA IRSD score (reversed) and aggregated over mesh block green space qualities. Only variables with a correlation rho≤0.40 or rho≤-0.40 for any geography at either 400m or 1600m are displayed here.

| Index description | Method | Buffer 400m (Rho and P-value) |  |  |  | Buffer 1600m (Rho and P-value) |  |  |  |
| --- | --- | --- | --- | --- | --- | --- | --- | --- | --- |
|  |  | Full set | Syd | New | Wol | Full set | Syd | New | Wol |
| Length of Main Roads within 100 metres of the GS | w0 Sum | -0.04* | -0.04* | 0.08* | 0.18* | 0 | 0 | 0.18* | 0.43* |
| Percentage of GS which is greater than 6 degrees slope | Formula | -0.42* | -0.44* | -0.32* | -0.42* | -0.49* | -0.52* | -0.34* | -0.44* |
| Presence of coastline | Quality | -0.26* | -0.31* | -0.06× | -0.02 | -0.35* | -0.41* | -0.16* | 0.21* |
| Number of mammal species/species habitat likely to occur within GS | Max | -0.51* | -0.56* | -0.21* | -0.29* | -0.47* | -0.53* | -0.21* | -0.23* |
| Total number of species/species habitat likely to occur within GS | Formula | -0.48* | -0.54* | -0.07* | 0.19* | -0.39* | -0.45* | 0 | 0.18* |
| Shannon diversity index | Formula | -0.41* | -0.46* | -0.18* | -0.29* | -0.39* | -0.44* | -0.23* | -0.28* |
| Simpson diversity index (range 0 to 1=most diverse) | Formula | -0.30* | -0.33* | -0.11* | -0.40* | -0.28* | -0.30* | -0.23* | -0.49* |
| Area of open grass within GS | w0 Sum | 0.16* | 0.14* | 0.11* | 0.27* | 0.30* | 0.29* | 0.17* | 0.45* |
| Area of roads/paths within GS | w0 Sum | 0.06* | 0.02* | 0.03 | 0.22* | 0.19* | 0.16* | 0.03+ | 0.41* |
| Percentage of open grass within GS | Formula | 0.30* | 0.29* | 0.27* | 0.34* | 0.38* | 0.38* | 0.29* | 0.44* |
| Percentage of tree canopy within GS | Formula | -0.40* | -0.41* | -0.21* | -0.32* | -0.46* | -0.47* | -0.23* | -0.44* |
| Percentage of tree canopy to roads surrounding GS (within 100m doughnut buffer) | Formula | -0.44* | -0.45* | -0.12* | -0.30* | -0.52* | -0.54* | -0.20* | -0.40* |
| Percentage of tree canopy $\leq 4$m to roads surrounding GS (within 100m doughnut buffer) | Formula | -0.20* | -0.19* | -0.01 | -0.34* | -0.25* | -0.24* | 0.06× | -0.41* |
| Percentage of tree canopy $>4$m to roads surrounding GS (within 100m doughnut buffer) | Formula | -0.43* | -0.44* | -0.13* | -0.26* | -0.51* | -0.53* | -0.20* | -0.37* |
| Percentage of tree canopy surrounding GS (within 100m doughnut buffer) | Formula | -0.35* | -0.35* | -0.12* | -0.19* | -0.44* | -0.45* | -0.16* | -0.31* |
| Percentage of tree canopy $>4$m surrounding GS (within 100m doughnut buffer) | Formula | -0.36* | -0.36* | -0.13* | -0.16* | -0.45* | -0.46* | -0.16* | -0.29* |

P-value: \* $\leq 0.001$ , × $\leq 0.01$ , + $\leq 0.05$ . For more table notes, see Supplementary Material S6.

For the full set using 1600m buffers, more disadvantaged areas tended to have lower percentages of nearby street trees to roads (Rho=-0.52, p≤0.001) and nearby street trees >4m to roads (Rho=-0.51, p≤0.001), lower percentages of slope >6° (Rho=-0.49, p≤0.001), lower likelihood of threatened mammal species/habitat occurrences (Rho=-0.47, p≤0.001), lower percentages of tree canopy (Rho=-0.46, p≤0.001) and percentage of surrounding tree canopy >4m nearby (Rho=-0.45, p≤0.001)(Table 1).

### Positive correlations between green space qualities and socioeconomic disadvantage

For the full set using 400m buffers, more disadvantaged areas tended to have a higher percentage of open grass (Rho=0.30, p-value≤0.001)(Table 1).

For the full set using 1600m buffers, more disadvantaged areas tended to have green spaces with higher percentages of open grass (Rho=0.38, p≤0.001) and bare earth (Rho=0.33, p≤0.001), area of grass (Rho=0.3, p≤0.001) and higher densities of robberies (Rho=0.34, p≤0.001), malicious damage (Rho=0.33, p≤0.001) and incivilities (Rho=0.3, p≤0.001)(Table 1).

### Qualities not correlated with socioeconomics

At 400m for the full set, there were no correlations with socioeconomics for number of takeaway outlets within 100m and built up area within green space (each with Rho=0). At 1600m for the full set, there were no correlations for pathway length within green spaces or length of nearby main roads (each with Rho=0).

### Buffer size differences

Differences between buffer sizes tended to be small (Figure 3). For select qualities with large Rho values, percentage of street trees to roads had the largest difference across buffer sizes, with the Rho value for the 100m buffer being -0.4 which continued a modest decrease to -0.52 at the 1600m buffer (Figure 3). Between the 400m and 1600m buffer sizes, the largest difference overall in Rho values was 0.18 for area of industrial land use and percentage of industrial land use (Tables S4-S5). The direction of correlation differed for 16/104 qualities (15.4%)^6^, yet at best these were only weakly correlated (i.e. the Rho values for both 400m and 1600m buffers were ≤0.15 if positive or ≥-0.15 if negative).

**Figure 3.**
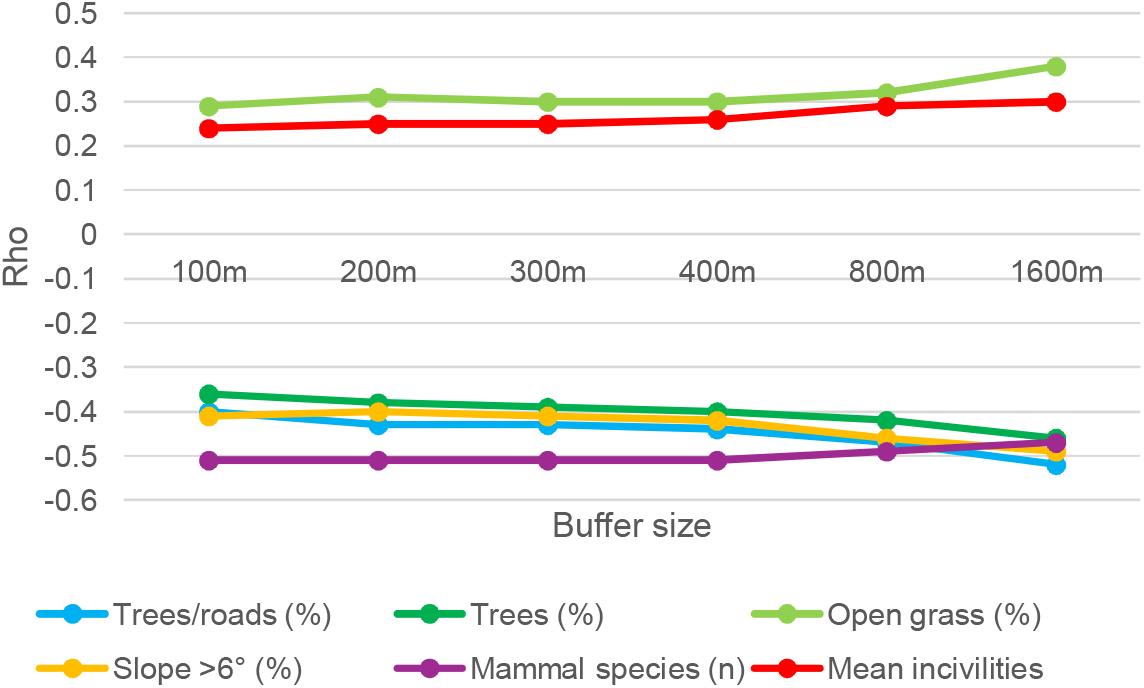
Differences in correlations (Rho) between buffer sizes for selected green space qualities.

### Between-city analysis

The direction of correlation differed for 59/104 qualities (56.7%) between the three cities (400m). The largest differences for the 400m analysis were for total threatened species/habitat (Sydney: Rho = -0.54, p-value ≤0.001, Newcastle: Rho = -0.07, p-value ≤0.001, Wollongong: Rho = 0.19, p-value ≤0.001) and threatened flora species/habitat (Sydney: Rho = -0.35, p-value ≤0.001, Newcastle: Rho = 0.06, p-value ≤0.01, Wollongong: Rho = 0.32, p-value ≤0.001).

The direction of correlation differed for 55/104 qualities (52.9%) between the three cities (1600m). The largest differences were for total threatened species/habitat (Sydney: Rho = -0.45, p-value ≤0.001, Newcastle: Rho = 0, p-value = 0.881, Wollongong: Rho = 0.18, p-value ≤0.001) and coastline presence (Sydney: Rho = -0.41, p-value ≤0.001, Newcastle: Rho = -0.16, p-value ≤0.001, Wollongong: Rho = 0.21, p-value ≤0.001).

### Multilevel modelling

When controlling for population factors and nested geographies, multilevel modelling tended to confirm that the associations from rank correlations persist. The larger the IRSD quintile, the larger the absolute value of the regression coefficient (Table 2, Table S12). For the four qualities that have a positive Spearman’s rank correlation, the multilevel models tended to reflect the positive association. For the six qualities that had a negative Spearman’s rank correlation, the multilevel models tended to also support the negative association (Table 2, Table S12). However, mammal species was an exception. Small/no effects were observed for the biodiversity indices, almost without trend in Models 4 and 5. This can be attributed to the biodiversity qualities having a large granularity, so that within the SA2 level, the associations were not clearly discernible.

**Table 2.** Summary of green space qualities associations with IRSD adjusted for population density and random effects of geographic areas (buffer size=1600m). Models 2, 4 and 5 are shown here. For all models, see Table S12.

| Quality | Model | IRSD quintile (ref=1, least disadvantaged) | | | | SA2 population density (deciles) | ΔDIC |
| --- | --- | --- | --- | --- | --- | --- | --- |
|  |  | 2 | 3 | 4 | 5 |  |  |
| Trees/roads (%) | 2 | -4.8 (-5.0, -4.6)* | -8.1 (-8.3, -7.9)* | -10.0 (-10.2, -9.8)* | -11.3 (-11.6, -11.1)* | - | 81,399 |
|  | 4 | -0.8 (-0.9, -0.7)* | -1.3 (-1.4, -1.2)* | -1.2 (-1.4, -1.1)* | -1.2 (-1.3, -1.0)* | - | 3 |
|  | 5 | -0.8 (-0.9, -0.7)* | -1.3 (-1.4, -1.2)* | -1.2 (-1.4, -1.1)* | -1.2 (-1.3, -1.0)* | 0.9 (0.7, 1.2)* | 3 |
| Trees (%) | 2 | -7.9 (-8.3, -7.5)* | -15.0 (-15.4, -14.5)* | -19.0 (-19.5, -18.5)* | -22.1 (-22.5, -21.6)* | - | 54,473 |
|  | 4 | -2.0 (-2.2, -1.7)* | -4.6 (-4.9, -4.3)* | -6.1 (-6.4, -5.7)* | -8.2 (-8.6, -7.7)* | - | 2 |
|  | 5 | -2.0 (-2.2, -1.7)* | -4.6 (-4.9, -4.3)* | -6.1 (-6.4, -5.7)* | -8.2 (-8.6, -7.7)* | -0.1 (-0.6, 0.4) | 2 |
| Slope >6° (%) | 2 | -11.5 (-12.0, -11.1)* | -19.1 (-19.6, -18.6)* | -22.4 (-22.9, -21.9)* | -26.3 (-26.8, -25.9)* | - | 43,471 |
|  | 4 | -4.5 (-4.8, -4.2)* | -8.0 (-8.4, -7.7)* | -9.7 (-10.1, -9.3)* | -11.4 (-11.9, -10.9)* | - | 3 |
|  | 5 | -4.5 (-4.8, -4.2)* | -8.0 (-8.4, -7.7)* | -9.7 (-10.1, -9.3)* | -11.4 (-11.9, -10.9)* | -1.3 (-1.8, -0.8)* | 3 |
| Mammal species, n | 2 | -0.8 (-0.9, -0.7)* | -1.4 (-1.4, -1.3)* | -1.8 (-1.9, -1.8)* | -2.4 (-2.5, -2.4)* | - | 109,992 |
|  | 4 | 0.0 (-0.0, 0.0) | 0.0 (-0.0, 0.0) | -0.0 (-0.0, 0.0) | -0.0 (-0.1, 0.0) | - | 4 |
|  | 5 | 0.0 (-0.0, 0.0) | 0.0 (-0.0, 0.0) | -0.0 (-0.0, 0.0) | -0.0 (-0.1, 0.0) | 0.2 (0.1, 0.2)* | 4 |
| All species, n | 2 | -6.8 (-7.3, -6.2)* | -12.5 (-13.1, -11.9)* | -16.3 (-16.9, -15.6)* | -23.7 (-24.3, -23.1)* | - | 94,603 |
|  | 4 | -0.4 (-0.6, -0.2)x | -0.3 (-0.6, -0.0)+ | 0.1 (-0.2, 0.5) | 0.3 (-0.1, 0.7)^ | - | 1 |
|  | 5 | -0.4 (-0.6, -0.2)* | -0.3 (-0.6, -0.0)+ | 0.1 (-0.2, 0.4) | 0.3 (-0.1, 0.7) | 1.9 (1.2, 2.5)* | 1 |
| Shannon index | 2 | -0.059 (-0.062, -0.056)* | -0.091 (-0.094, -0.088)* | -0.109 (-0.112, -0.105)* | -0.137 (-0.140, -0.133)* | - | 80,266 |
|  | 4 | -0.005 (-0.006, -0.003)* | -0.006 (-0.007, -0.004)* | -0.009 (-0.011, -0.007)* | -0.005 (-0.008, -0.003)* | - | 2 |
|  | 5 | -0.005 (-0.006, -0.003)* | -0.006 (-0.008, -0.004)* | -0.009 (-0.011, -0.007)* | -0.005 (-0.008, -0.003)* | 0.002 (-0.002, 0.005) | 2 |
| Mean incivilities | 2 | 0.234 (0.207, 0.260)* | 0.396 (0.368, 0.424)* | 0.563 (0.534, 0.591)* | 0.828 (0.800, 0.856)* | - | 62,455 |
|  | 4 | 0.196 (0.180, 0.211)* | 0.368 (0.350, 0.386)* | 0.530 (0.509, 0.551)* | 0.715 (0.690, 0.739)* | - | 2 |
|  | 5 | 0.196 (0.180, 0.211)* | 0.368 (0.350, 0.386)* | 0.530 (0.509, 0.551)* | 0.713 (0.689, 0.738)* | 0.208 (0.185, 0.229)* | 1 |
| Open grass (sq m) | 2 | 12373 (10834, 13893)* | 22746 (21142, 24339)* | 27118 (25473, 28754)* | 31135 (29534, 32721)* | - | 32,941 |
|  | 4 | 5765 (4571, 6954)* | 14520 (13101, 15937)* | 18199 (16586, 19804)* | 24911 (23021, 26785)* | - | 4 |
|  | 5 | 5761 (4558, 6954)* | 14515 (13086, 15939)* | 18175 (16573, 19802)* | 24832 (22937, 26730)* | 3678 (2274, 5008)* | 3 |
| Open grass (%) | 2 | 4.5 (4.1, 4.9)* | 8.8 (8.4, 9.2)* | 11.8 (11.4, 12.2)* | 15.5 (15.1, 15.9)* | - | 45,671 |
|  | 4 | 1.7 (1.4, 1.9)* | 4.0 (3.7, 4.3)* | 4.7 (4.3, 5.0)* | 6.2 (5.7, 6.6)* | - | 1 |
|  | 5 | 1.7 (1.4, 1.9)* | 4.0 (3.7, 4.3)* | 4.7 (4.3, 5.0)* | 6.1 (5.7, 6.6)* | 0.3 (-0.1, 0.7) | 1 |
| Bare earth (%) | 2 | 1.4 (1.1, 1.7)* | 3.4 (3.0, 3.7)* | 4.4 (4.1, 4.7)* | 6.7 (6.4, 7.0)* | - | 65,347 |
|  | 4 | 0.2 (0.1, 0.4)x | 0.8 (0.6, 1.0)* | 1.4 (1.2, 1.6)* | 1.9 (1.6, 2.1)* | - | 1 |
|  | 5 | 0.2 (0.1, 0.4)x | 0.8 (0.6, 1.0)* | 1.4 (1.2, 1.6)* | 1.9 (1.6, 2.1)* | -0.0 (-0.4, 0.3) | 1 |

\* P≤0.001, x P≤0.01, + P≤0.05, ^ P≤0.1. For more table notes, see Supplementary Material S6.

The absolute value of the regression coefficients uniformly decreases from Model 2 to Models 4 and 5. This is attributed to a strong geographical trend that exceeds the SA2 level. Nevertheless, within models at the SA2 level, the direction of association remains the same. The ΔDIC indicates that Models 4 and 5 are better than Model 2.

## Discussion

### Main findings

There was variation in certain qualities of green spaces by socioeconomic status. For example, more socioeconomically disadvantaged areas had higher percentages of grass and bare earth, and higher densities of robberies and malicious damage. The least disadvantaged areas had higher likely occurrences of threatened mammal species, larger percentages of trees, nearby street trees to roads, and slope >6°.

The city affects the direction for certain correlations of green space qualities with socioeconomic advantage, highlighting that context is vital. However, the extent to which this matters varies depending on the quality. For the 1600m analysis, biodiversity qualities and coastline presence were highly influenced by the city. For total threatened species/habitat, Sydney was negatively correlated, Newcastle had no correlation and Wollongong had a weak positive correlation. For coastline presence, Sydney was negatively correlated, Newcastle was weakly negatively correlated and Wollongong was weakly positively correlated. Correlation differences could be due to differing urban structure and development patterns, with outer suburbs in Australian cities not uniformly linked with socioeconomic disadvantage, especially with differences between metropolitan cities (e.g. Sydney) and regional cities (e.g. Newcastle, Wollongong)(Giles-Corti et al., 2022). For example, Sydney has greater disadvantage in the outer and inland Western suburbs, while in Wollongong, more disadvantaged areas are located close to the CBD and nearer to the coast (Giles-Corti et al., 2022). Despite the differences in correlation directions, however, key qualities with large Rho values tended to be consistent across cities. These included area and percentage of slope >6°, number of mammal species, mean level of incivilities, area and percentage of open grass, percentage of tree canopy, percentage of nearby street trees to roads and percentage of surrounding tree canopy.

### Comparisons with prior literature

Previous research has linked tree canopy cover and tree density with mental, physical, and social health benefits (Astell-Burt et al., 2023, Reid et al., 2017, Astell-Burt and Feng, 2019, Feng et al., 2023, Jiang et al., 2020, Feng et al., 2022, Kardan et al., 2015, Astell-Burt and Feng, 2020, Feng et al., 2021). This study has demonstrated for the first time in Sydney, Newcastle and Wollongong that percentage tree cover is lower in disadvantaged areas. This supports Fossa et al.’s (2023) finding that parks in more socioeconomically deprived areas had less tree canopy. However, it is different to a study from Sydney that has found that public park tree cover had a slight negative association with socioeconomic advantage (Lin et al., 2015). It is noted that Lin et al. (2015) conducted analyses for the Index of Relative Socio-economic Advantage and Disadvantage (IRSAD) rather than IRSD at the SA1 level and then aggregate up to the Statistical Area 2 (SA2) level, whereas in our analysis, IRSD values for mesh blocks were assumed to be the same as their respective SA1. As there are differences between the IRSD (focus on disadvantage) and IRSAD (focus on both advantage and disadvantage), this may make a slight difference to results, for example if a place has both high levels of advantage and disadvantage. Since different scales of aggregation can influence results (Flowerdew et al., 2008), our aggregated data was at a finer scale which can decrease the effect of the modifiable areal unit problem given that SA2s tend to range from 3,000–25,000 people (ABS, 2021c), while SA1s tend to range from 200–800 people (ABS, 2021b). Whereas we aggregate percentage tree cover based on green spaces that are within specific buffers from mesh block centroids, Lin et al. (2015) do not use this method of aggregation. Their analysis “did not consider access to parks beyond the suburb boundary”^7^ though residents could potentially access nearby parks that are in different suburbs (p. 956). While most green spaces had public land status, our study did not distinguish between green spaces based on land status and included unspecified land status.

Less tree cover in more disadvantaged areas could have important health implications. Larger percentages of tree canopy have been associated with lower odds of heart disease, hypertension and diabetes (Astell-Burt and Feng, 2019). Not only do residents at most risk of developing preventable diseases live in areas that contain the smallest amounts of green space (Astell-Burt et al., 2014) but qualities associated with good health such as tree cover are also smaller.

We found that larger likelihood of total and mammal, species/habitat occurrences are correlated with areas of lower disadvantage overall. However, only mammal species was consistent across all three study areas. There are mental health benefits with increased green space biodiversity (Fuller et al., 2007). Measures to increase biodiversity in areas of higher disadvantage may benefit the mental health and wellbeing of residents in these areas. We also tested correlations between percentage tree canopy in green spaces and assigned biodiversity values which had positive correlations (Table S13).

This research has found that slope >6° is correlated with less disadvantaged areas. Although there is a scarcity of research examining the relationship between slope of green spaces and area-level socioeconomic status, an American study has found a link between uneven terrain and wealth of urban communities (Ye and Becker, 2018). Using green spaces with steeper slopes may elicit increased physical activity benefits. For example, increased slope engages more leg muscle activity than walking on flat surfaces (Psurny et al., 2018). While Villanueva et al. (2013) have found that living in hillier areas is associated with lower odds of diabetes which could be explained by increased intensity of physical activity due to steeper slopes, Sun et al. (2022) suggest that “participation in physical activity may not be the main mechanism behind the associations between green space and health status” in hilly areas (p. 14). Rather, perceived greenery and social cohesion and safety may be more important (Sun et al., 2022). More research examining the health effects and socio-economic relationships with slope of green spaces is warranted, as well as the degree to which results are generalisable to other contexts. For example, the favelas in Brazil would have vastly different patterns to Australian cities.

This study has found that areas of grass or bare earth are more likely to be found in areas of higher disadvantage. This supports Fossa et al.’s (2023) finding that areas of socioeconomic deprivation had more grass/impervious/soil surfaces. Large grassed areas however can support physical activities such as ball sports (Fossa et al., 2023). We are unable to distinguish whether grassed areas are manicured lawns for golf and other sports, or grass growing on derelict land for example. Upkeep and usage of grassed fields is therefore an important consideration, but has not been considered in our study.

Buffer size matters for geographical differences, for example in hotspot analyses for percentage tree cover with typically stronger patterns for larger buffer sizes. However, in terms of correlations with socioeconomic disparities, buffer size made only minor differences, with somewhat stronger correlations for percentage tree cover for larger buffer distances (Figure 3). In terms of health benefits, other studies have found larger buffer sizes to be more important, for example, there were fewer cases of anxiety/mood disorder treatments with greater proportion of and shorter distances to green space within 3km, but not within 300m (Nutsford et al., 2013). Although this provides some assurances that the socioeconomic inequities observed appear to be reasonably consistent regardless of buffer size, this does not mean the identification, magnitudes, and directions of associations between green space qualities and various health and behavioural outcomes are not contingent upon buffer size and these warrant testing.

### Spatial planning and policy implications

It is important that policy makers and planners do not just measure access and the availability of green spaces, but assess the qualities of those green spaces to understand what they provide local residents.

How a city plans and manages its green space strategically will have large scale consequences on the quality of green spaces. Street trees (Wang et al., 2022) as well as trees within green spaces should be planted and grassed/bare earth areas minimised in low socioeconomic areas to reduce disparities in tree canopy cover. Although tree cover in parks/yards tends to be higher in areas of socioeconomic advantage in Brisbane, Australia, high quality remnant vegetation is more equally distributed (Shanahan et al., 2014). Thus, councils should ensure proper management of remnant vegetation (Shanahan et al., 2014).

To encourage animal diversity in these areas, locally native plant species should be planted to provide habitat for native animals (Ikin et al., 2015, Berthon et al., 2021) and understorey vegetation volume should be increased (Threlfall et al., 2017). Education initiatives for local residents and visitors to green spaces to protect biodiversity, such as properly containing cats could also be implemented (Ikin et al., 2015).

The Existing Green Assets dataset did not extend to Newcastle and Wollongong. It would be beneficial to have comprehensive green space data in these areas included in future for greater consistency when comparing results across cities.

### Strengths

A main strength of this article is the use of a rich green space dataset with qualities spanning access, amenities/activities, beaches/coastline, biodiversity, incivilities, land cover, and neighbouring land uses. This dataset can be used by researchers to assess associations with mental, physical, and social health outcomes. Spatial data from government and other agencies provides high quality, robust and consistent data across large geographical areas and reduces requirement for costly and timely in-situ audits. We analysed the qualities across three Australian cities of different population sizes, with coastal and/or inland communities. We used eight different sized road network buffers and strengthened findings from correlation analysis through conducting multilevel regression models. By demonstrating which qualities may matter most for protecting and promoting public health and levelling up health inequities, this evidence can inform significant investment in urban planning strategies designed to realise those improvements with and for communities.

### Limitations

We do not empirically estimate the health impacts of green spaces in the current study, but we extrapolate from other evidence that green space and socioeconomic disadvantage are likely to have a downstream impact on health inequities that we are inferring. For example, while causal evidence for green space quality and health is lacking, Ai et al. (2023) have found a relationship between increased green space and reduced air pollution, with air pollution influencing Disability-Adjusted Life Years (DALYs) (Hu et al., 2023). We show that more disadvantaged areas have poorer quality of green space (e.g. more likely to have open grass and crime and less likely to have tree cover) which we infer will have an impact on health in addition to the impact on health of being more disadvantaged. Testing associations between these quality indicators and various health outcomes is a focus for future research and beyond the scope of the current study.

There were some data quality issues with the datasets used. For example, with land cover data, there was some potential for image/pixel misclassification. Certain areas of tree cover and open grass appeared to be misclassified, for example as bare earth. We found a clear misclassification near Bicentennial Park/Homebush (Figure S2-S3). There were also examples of missing pathways in green spaces in the Geoscape roads data (Figure S4).

Since green spaces in Sydney were split into singlepart features, there may be some instances where single green spaces are divided into multiple smaller green spaces. There may also be double counting of certain qualities when 100m buffers/doughnut buffers are used around green space polygons.

Residents may use short cuts that deviate from the street network used for the buffers. Some green spaces may have been excluded from buffers around mesh block centroids, even though it may be feasible for a resident to access these in reality.

This study focuses on provision of green space qualities and not on their use (Astell-Burt et al., 2014). Availability of green space qualities does not mean that those at most risk of developing preventable diseases or in need of controlling them will use parks and benefit from the qualities offered.

Buffer sizes may have different meanings across the three cities examined here. The variation of green space in the city is related to the size, planning, and climatology of the cities. In areas that are drier or more densely built (due to planning requirement, e.g. around train stations), there will be a much different expectation and ability to maintain green space. The size of a city will have different population pressures that impact on the need to do these things, so an urban centre in a smaller city may be under fewer constraints and thus the results of these analyses at the same buffer size may be impacted differentially.

Edge effects relating to the large bushland/green space areas being removed from analysis^8^ may occur. It was assumed, however, that access to these areas would be more difficult than other smaller green spaces. Excluding motorways in the network analysis may also influence access issues for those living close to motorways by underestimating travel-time if going by car. However, these may represent barriers to access by pedestrians for example due to a general lack of safe crossing points, and may be damaging to restorative experiences due to excessive noise.

The finest level that SEIFA IRSD data was available was the SA1 level. We therefore had to assign the SA1 value to mesh blocks within it.

### Future work

Other qualities that could be considered in future include children’s playgrounds. For example, more children used and actively played in playground facilities that were more varied (Reimers and Knapp, 2017). However, as this research was related to older people it was beyond the scope of the present study. Nevertheless, children’s playgrounds offer intergenerational interactions for example through grandparents bringing their grandchildren. Picnic and barbecue facilities are also positive for social interaction in older adults (Veitch et al., 2020).

Future work could include a longitudinal analysis of the health effects of access to specific green space qualities and how this varies among populations of differing socioeconomic status. This could provide causal evidence linking green space qualities with varied health outcomes.

We used the IRSD so that we could focus on disadvantage, since the IRSAD could mask some areas with high disadvantage if these areas also have large proportions of advantage. The IRSD contains variables relating to low income, low rent, unemployment, low education, disabilities, one parent families, overcrowding of dwellings, lack of cars, separated/divorced, poor English, employment as “Labourers”, “Machinery Operators and Drivers”, or “Low Skill Community and Personal Service Workers” (ABS, 2023a). Future work could explore other indices such as IRSAD, or indicators of education or income specifically to see if there are differences in correlations with green space qualities.

## Conclusion

Socioeconomic disparities in green space qualities of tree cover, total threatened species, slope and incivilities have been identified. Previous studies have found tree cover and bird diversity to be good for health and/or wellbeing (Astell-Burt and Feng, 2019, Methorst et al., 2021, Aerts et al., 2018, Fuller et al., 2007). A lack of these qualities in more disadvantaged areas could contribute to higher rates of chronic disease and poorer mental wellbeing. Buffer size influences tended to be small for green space correlations with socioeconomic inequality. This is the first study to analyse a comprehensive set of access, amenity/activity, beaches/coastline, biodiversity, incivility, land cover and land use qualities in Sydney, Newcastle and Wollongong. This could help planners and policy makers to improve health and wellbeing in more disadvantaged populations through the planting of native plants/trees in and around urban parks and implementation of education initiatives for conserving biodiversity.

## Data Availability

The authors do not have permission to share confidential data.

## Acknowledgements

The authors would like to acknowledge Philip Kosiak for preparing and documenting GIS data in the initial phases of the project. The authors would like to acknowledge OpenStreetMap for map data (copyrighted OpenStreetMap contributors and available from https://www.openstreetmap.org). This research used the NCRIS-enabled Australian Urban Research Infrastructure Network (AURIN) e-Infrastructure to access Australian Government Department of Social Services “DSS - National Public Toilets (Point) 2017” on 5 February 2024.

## Footnotes

1 Parts of these excluded green spaces may be located within the UCL boundary too.

2 Whenever 100m buffers/100m doughnut buffers are used to calculate various qualities around green spaces, barriers such as rivers, train tracks or roadways prohibiting pedestrians were not considered.

3 Specifically, likely occurrences of threatened species/species habitat.

4 Tree data was checked against a data set derived from GeoVision December 2020 2m landcover data (Precisely) and World Imagery basemap (Esri, Maxar, Earthstar Geographics, and the GIS User Community). There were many minor occurrences where there was no instance of trees in the GeoVision November 2021 data but the data set derived from GeoVision December 2020 had trees. These cases were not necessarily coded as inadequate data unless the extent of the data likely did not properly overlap the green spaces/study area.

5 The cold spot near the centre of Sydney may be due to a misclassification in the land cover data where areas of trees or grass are misclassified in original data, for example as bare earth e.g. see Figure S2 and Figure S3.

6 Including qualities for which one Rho value was zero.

7 Lin et al. (2015) define suburbs as the SA2 boundary.

8 For example in Sydney, parts of the Blue Mountains, parts of Sutherland, near Berowra, and near Ku-ring-gai Chase National Parks.

## References

Aerts, R., Honnay, O. & Van Nieuwenhuyse, A. 2018. Biodiversity and human health: mechanisms and evidence of the positive health effects of diversity in nature and green spaces. Br Med Bull, 127, 5–22.

Ai, H., Zhang, X. & Zhou, Z. 2023. The impact of greenspace on air pollution: Empirical evidence from China. Ecological Indicators, 146.

Astell-Burt, T. & Feng, X. 2019. Urban green space, tree canopy, and prevention of heart disease, hypertension, and diabetes: a longitudinal study. The Lancet Planetary Health, 3.

Astell-Burt, T. & Feng, X. 2020. Urban green space, tree canopy and prevention of cardiometabolic diseases: a multilevel longitudinal study of 46 786 Australians. International journal of epidemiology, 49, 926–933.

Astell-Burt, T., Feng, X., Mavoa, S., Badland, H. M. & Giles-Corti, B. 2014. Do low-income neighbourhoods have the least green space? A cross-sectional study of Australia’s most populous cities. BMC Public Health, 14, 292.

Astell-Burt, T., Hartig, T., Putra, I., Walsan, R., Dendup, T. & Feng, X. 2022. Green space and loneliness: A systematic review with theoretical and methodological guidance for future research. Sci Total Environ, 847, 157521.

Astell-Burt, T., Walsan, R., Davis, W. & Feng, X. 2023. What types of green space disrupt a lonelygenic environment? A cohort study. Soc Psychiatry Psychiatr Epidemiol, 58, 745–755.

Australian Bureau Of Statistics. 2021a. Australian Statistical Geography Standard (ASGS) Edition 3 [Online]. Available: https://www.abs.gov.au/statistics/standards/australian-statistical-geography-standard-asgs-edition-3/latest-release [Accessed 12 September 2024].

Australian Bureau Of Statistics. 2021b. Statistical Area Level 1: Australian Statistical Geography Standard (ASGS) Edition 3 [Online]. Available: https://www.abs.gov.au/statistics/standards/australian-statistical-geography-standard-asgs-edition-3/jul2021-jun2026/main-structure-and-greater-capital-city-statistical-areas/statistical-area-level-1 [Accessed 3 June 2024].

Australian Bureau Of Statistics. 2021c. Statistical Area Level 2: Australian Statistical Geography Standard (ASGS) Edition 3 [Online]. Available: https://www.abs.gov.au/statistics/standards/australian-statistical-geography-standard-asgs-edition-3/jul2021-jun2026/main-structure-and-greater-capital-city-statistical-areas/statistical-area-level-2 [Accessed 3 June 2024].

Australian Bureau Of Statistics. 2023a. Construction of the indexes: Socio-Economic Indexes for Areas (SEIFA): Technical Paper [Online]. Available: https://www.abs.gov.au/statistics/detailed-methodology-information/concepts-sources-methods/socio-economic-indexes-areas-seifa-technical-paper/2021/construction-indexes#technical-details-of-each-index-variables-and-loadings [Accessed 30 October 2025].

Australian Bureau Of Statistics. 2023b. Socio-Economic Indexes for Areas (SEIFA), Australia. Index data cubes. [Online]. Available: https://www.abs.gov.au/statistics/people/people-and-communities/socio-economic-indexes-areas-seifa-australia/latest-release [Accessed 31 July 2023].

Australian Bureau Of Statistics [ABS]. 2021. Mesh Blocks: Australian Statistical Geography Standard (ASGS) Edition 3 [Online]. Australian Bureau of Statistics. Available: https://www.abs.gov.au/statistics/standards/australian-statistical-geography-standard-asgs-edition-3/jul2021-jun2026/main-structure-and-greater-capital-city-statistical-areas/mesh-blocks [Accessed 14 August 2023].

Berthon, K., Thomas, F. & Bekessy, S. 2021. The role of ‘nativeness’ in urban greening to support animal biodiversity. Landscape and Urban Planning, 205.

Browne, W. J. 2023. MCMC Estimation in MLwiN, v3.07. Centre for Multilevel Modelling, University of Bristol.

Bruton, C. M. & Floyd, M. F. 2014. Disparities in built and natural features of urban parks: comparisons by neighborhood level race/ethnicity and income. J Urban Health, 91, 894–907.

Crawford, D., Timperio, A., Giles-Corti, B., Ball, K., Hume, C., Roberts, R., Andrianopoulos, N. & Salmon, J. 2008. Do features of public open spaces vary according to neighbourhood socio-economic status? Health Place, 14, 889–93.

Feng, X., Navakatikyan, M. A., Toms, R. & Astell-Burt, T. 2023. Leafier communities, healthier hearts: an Australian cohort study of 104,725 adults tracking cardiovascular events and mortality across 10 years of linked health data. Heart, Lung and Circulation, 32, 105–113.

Feng, X., Toms, R. & Astell-Burt, T. 2021. Association between green space, outdoor leisure time and physical activity. Urban Forestry & Urban Greening, 66, 127349.

Feng, X., Toms, R. & Astell-Burt, T. 2022. The nexus between urban green space, housing type, and mental health. Social Psychiatry and Psychiatric Epidemiology, 57, 1917–1923.

Flowerdew, R., Manley, D. J. & Sabel, C. E. 2008. Neighbourhood effects on health: does it matter where you draw the boundaries? Soc Sci Med, 66, 1241–55.

Fossa, A. J., Zelner, J., Bergmans, R., Zivin, K. & Adar, S. D. 2023. Sociodemographic correlates of greenness within public parks in three U.S. cities. Wellbeing, Space and Society, 5.

Fuller, R. A., Irvine, K. N., Devine-Wright, P., Warren, P. H. & Gaston, K. J. 2007. Psychological benefits of greenspace increase with biodiversity. Biology letters, 3, 390–394.

Giles-Corti, B., Saghapour, T., Turrell, G., Gunn, L., Both, A., Lowe, M., Rozek, J., Roberts, R., Hooper, P., Butt, A. & Higgs, C. 2022. Spatial and socioeconomic inequities in liveability in Australia’s 21 largest cities: Does city size matter? Health Place, 78, 102899.

Hartig, T., Mitchell, R., De Vries, S. & Frumkin, H. 2014. Nature and health. Annu Rev Public Health, 35, 207–28.

Heo, S. & Bell, M. L. 2023. Investigation on urban greenspace in relation to sociodemographic factors and health inequity based on different greenspace metrics in 3 US urban communities. J Expo Sci Environ Epidemiol, 33, 218–228.

Hoffimann, E., Barros, H. & Ribeiro, A. I. 2017. Socioeconomic Inequalities in Green Space Quality and Accessibility-Evidence from a Southern European City. Int J Environ Res Public Health, 14.

Hu, W., Fang, L., Zhang, H., Ni, R. & Pan, G. 2023. Changing trends in the air pollution-related disease burden from 1990 to 2019 and its predicted level in 25 years. Environ Sci Pollut Res Int, 30, 1761–1773.

Ikin, K., Le Roux, D. S., Rayner, L., Villaseñor, N. R., Eyles, K., Gibbons, P., Manning, A. D. & Lindenmayer, D. B. 2015. Key lessons for achieving biodiversity-sensitive cities and towns. Ecological Management & Restoration, 16, 206–214.

Jiang, X., Larsen, L. & Sullivan, W. 2020. Connections between daily greenness exposure and health outcomes. International journal of environmental research and public health, 17, 3965.

Kardan, O., Gozdyra, P., Misic, B., Moola, F., Palmer, L. J., Paus, T. & Berman, M. G. 2015. Neighborhood greenspace and health in a large urban center. Scientific reports, 5, 11610.

Knobel, P., Dadvand, P., Alonso, L., Costa, L., Español, M. & Maneja, R. 2021. Development of the urban green space quality assessment tool (RECITAL). Urban Forestry & Urban Greening, 57.

Knobel, P., Dadvand, P. & Maneja-Zaragoza, R. 2019. A systematic review of multi-dimensional quality assessment tools for urban green spaces. Health Place, 59, 102198.

Li, X., Liu, Q., Liu, X. & Ren, Y. 2025. Greener is healthier: Unraveling the distinction of health impacts of urban green indicators in Wuhan, China. Sustainable Cities and Society, 125.

Lin, B., Meyers, J. & Barnett, G. 2015. Understanding the potential loss and inequities of green space distribution with urban densification. Urban Forestry & Urban Greening, 14, 952–958.

Liu, Y. & Li, G. 2025. Inequities in thermal comfort and urban blue-green spaces cooling: An explainable machine learning study across residents of different socioeconomic statuses in Hangzhou, China. Sustainable Cities and Society, 127.

Markevych, I., Schoierer, J., Hartig, T., Chudnovsky, A., Hystad, P., Dzhambov, A. M., De Vries, S., Triguero-Mas, M., Brauer, M., Nieuwenhuijsen, M. J., Lupp, G., Richardson, E. A., Astell-Burt, T., Dimitrova, D., Feng, X., Sadeh, M., Standl, M., Heinrich, J. & Fuertes, E. 2017. Exploring pathways linking greenspace to health: Theoretical and methodological guidance. Environ Res, 158, 301–317.

Methorst, J., Bonn, A., Marselle, M., Böhning-Gaese, K. & Rehdanz, K. 2021. Species richness is positively related to mental health – A study for Germany. Landscape and Urban Planning, 211.

Nutsford, D., Pearson, A. L. & Kingham, S. 2013. An ecological study investigating the association between access to urban green space and mental health. Public Health, 127, 1005–11.

Psurny, M., Svoboda, Z., Janura, M., Kubonova, E., Bizovska, L., Martinez Lemos, R. I. & Abrantes, J. 2018. The Effects of Nordic Walking and Slope of the Ground on Lower Limb Muscle Activity. Journal of Strength and Conditioning Research, 32, 217–222.

Rasbash, J., Steele, F., Browne, W. J. & Goldstein, H. 2023. A User’s Guide to MLwiN, v3.07. Centre for Multilevel Modelling, University of Bristol; 2023.

Reid, C. E., Clougherty, J. E., Shmool, J. L. C. & Kubzansky, L. D. 2017. Is All Urban Green Space the Same? A Comparison of the Health Benefits of Trees and Grass in New York City. Int J Environ Res Public Health, 14.

Reimers, A. K. & Knapp, G. 2017. Playground usage and physical activity levels of children based on playground spatial features. Z Gesundh Wiss, 25, 661–669.

Rigolon, A., Browning, M. & Jennings, V. 2018. Inequities in the quality of urban park systems: An environmental justice investigation of cities in the United States. Landscape and Urban Planning, 178, 156–169.

Shanahan, D. F., Lin, B. B., Gaston, K. J., Bush, R. & Fuller, R. A. 2014. Socio-economic inequalities in access to nature on public and private lands: A case study from Brisbane, Australia. Landscape and Urban Planning, 130, 14–23.

Singh, H. 2018. Spatial Epidemiological Investigation of Sport and Leisure Injuries in Victoria, Australia. Doctor of Philosophy, Federation University Australia.

Sun, P., Song, Y. & Lu, W. 2022. Effect of Urban Green Space in the Hilly Environment on Physical Activity and Health Outcomes: Mediation Analysis on Multiple Greenery Measures. Land, 11.

Threlfall, C. G., Mata, L., Mackie, J. A., Hahs, A. K., Stork, N. E., Williams, N. S. G., Livesley, S. J. & Beggs, J. 2017. Increasing biodiversity in urban green spaces through simple vegetation interventions. Journal of Applied Ecology, 54, 1874–1883.

TRAIL HIKING AUSTRALIA. 2024. Australian Walking Track Grading System (AWTGS) [Online]. Available: https://www.trailhiking.com.au/preparation/australian-walking-track-grading-system-awtgs/ [Accessed 16 September 2024].

Vaughan, K. B., Kaczynski, A. T., Wilhelm Stanis, S. A., Besenyi, G. M., Bergstrom, R. & Heinrich, K. M. 2013. Exploring the distribution of park availability, features, and quality across Kansas City, Missouri by income and race/ethnicity: an environmental justice investigation. Ann Behav Med, 45 Suppl 1, S28–38.

Veitch, J., Flowers, E., Ball, K., Deforche, B. & Timperio, A. 2020. Designing parks for older adults: A qualitative study using walk-along interviews. Urban Forestry & Urban Greening, 54.

Vidal, D. G., Fernandes, C. O., Viterbo, L. M. F., Vilaça, H., Barros, N. & Maia, R. L. 2021. Combining an evaluation grid application to assess ecosystem services of urban green spaces and a socioeconomic spatial analysis. International Journal of Sustainable Development & World Ecology, 28, 291–302.

Viinikka, A., Tiitu, M., Heikinheimo, V., Halonen, J. I., Nyberg, E. & Vierikko, K. 2023. Associations of neighborhood-level socioeconomic status, accessibility, and quality of green spaces in Finnish urban regions. Applied Geography, 157.

Villanueva, K., Knuiman, M., Koohsari, M. J., Hickey, S., Foster, S., Badland, H., Nathan, A., Bull, F. & Giles-Corti, B. 2013. People living in hilly residential areas in metropolitan Perth have less diabetes: spurious association or important environmental determinant? International Journal of Health Geographics, 12.

Wang, R., Feng, Z. & Pearce, J. 2022. Neighbourhood greenspace quantity, quality and socioeconomic inequalities in mental health. Cities, 129.

White, M. P., Elliott, L. R., Gascon, M., Roberts, B. & Fleming, L. E. 2020. Blue space, health and well-being: A narrative overview and synthesis of potential benefits. Environ Res, 191, 110169.

Yang, M., Ye, P. & He, J. 2025. Green and Blue Infrastructure for Urban Cooling: Multi-Scale Mechanisms, Spatial Optimization, and Methodological Integration. Sustainable Cities and Society.

Ye, V. Y. & Becker, C. M. 2018. The Z-axis: Elevation gradient effects in Urban America. Regional Science and Urban Economics, 70, 312–329.

You, H. 2016. Characterizing the inequalities in urban public green space provision in Shenzhen, China. Habitat International, 56, 176–180.

Zhang, J. 2023. Inequalities in the quality and proximity of green space exposure are more pronounced than in quantity aspect: Evidence from a rapidly urbanizing Chinese city. Urban Forestry & Urban Greening, 79.

Zhang, R., Zhang, C.-Q., Cheng, W., Lai, P. C. & Schüz, B. 2021. The neighborhood socioeconomic inequalities in urban parks in a High-density City: An environmental justice perspective. Landscape and Urban Planning, 211.

Zhang, Y. & Luo, F. 2025. How are green spaces associated with chronic disease incidence in Australia? Direct health benefits and interactive effects with socioeconomic status based on multiple green space indicators. Sustainable Cities and Society, 121.

Zhou, X., Sho, K., Qiu, H., Chang, S. & Cen, Q. 2024. Longitudinal association between urban blue-green space exposure and mortality: A systematic review and meta-analysis of exposure types and buffers. Sustainable Cities and Society, 116.

