## Supplementary Material for "Socioeconomic inequities within and between cities in objectively measured green space qualities at small geographical scales: Evidence from Australia"

### S1. Green space data

Data used to create the green space dataset is summarised in Table S1.

Table S1. Green space data

| **Data** | **Source** |
| --- | --- |
| Green space locations | NSW Department of Planning, Housing and Infrastructure 2019, “Existing Green Assets”, <https://datasets.seed.nsw.gov.au/dataset/c79c0f66-845b-4926-b750-1752f245f62b/resource/f98874cd-5d86-445d-bf0c-72944dd6740c/download/planning_green_assets.zip>, accessed 24 January 2024 (for Sydney). © State Government of NSW and NSW Department of Planning, Housing and Infrastructure 2019  ABS 2021 Mesh Blocks categorised as “Parkland” (for Newcastle and Wollongong) - ABS 2021, “Digital boundary files: Australian Statistical Geography Standard (ASGS) Edition 3”, <https://www.abs.gov.au/statistics/standards/australian-statistical-geography-standard-asgs-edition-3/jul2021-jun2026/access-and-downloads/digital-boundary-files> (accessed 31 July 2023). |
| Pathways | Roads February 2020 – “NSW_STREET_LINE_shp.shp” – PSMA (Geoscape Australia). |
| Roads | Roads February 2020 – “NSW_STREET_LINE_shp.shp” – PSMA (Geoscape Australia). |
| Bikeways | Transport for NSW 2018, “NSWBicycleNetwork5March2018.shp”, <https://opendata.transport.nsw.gov.au/dataset/cycleway-data/resource/c29501ba-9b7f-4857-8d7f-8bdf2265201f>, accessed 19 February 2019. |
| Elevation | Geoscience Australia 2015, “Digital Elevation Model (DEM) of Australia derived from LiDAR 5 Metre Grid”, Geoscience Australia, Canberra, <https://doi.org/10.26186/89644> (accessed 25 January 2024). |
| Railway station (including light rail stops), bus stop and ferry wharf locations | Transport for NSW 2023, “Timetables Complete GTFS” <https://opendata.transport.nsw.gov.au/dataset/timetables-complete-gtfs>, accessed 25 January 2024. |
| Trees (note separate to tree cover used within green spaces from land cover data) | GeoVision (Precisely), Trees, November 2021. |
| Public toilets | Department of Social Services, 2017. “DSS - National Public Toilets (Point) 2017”, <https://data.aurin.org.au/dataset/au-govt-dss-national-public-toilets-2017-na> – Accessed from AURIN. Accessed on 5 February 2024. |
| Café, hotel/bar, restaurant, takeaway, supermarkets/greengrocers locations | Sensis (Thryv Australia). |
| parkrun event course | parkrun 2024, <http://www.parkrun.com.au/events/>, accessed 14/18 March 2024. |
| Beaches | “gis_osm_natural_a_free_1.shp” – Geofabrik GmbH and OpenStreetMap Contributors 2022, “Australia.shp.zip [australia-latest-free.shp.zip]” downloaded from <https://download.geofabrik.de/australia-oceania/australia-latest-free.shp.zip>, accessed 12 December 2022. |
| Coastline | Created from States and Territories shapefile, “STE_2021_AUST_2020MGA56.shp” – ABS 2021, “Digital boundary files: Australian Statistical Geography Standard (ASGS) Edition 3”, <https://www.abs.gov.au/statistics/standards/australian-statistical-geography-standard-asgs-edition-3/jul2021-jun2026/access-and-downloads/digital-boundary-files> (accessed 31 July 2023). |
| Biodiversity data | NSW Department of Climate Change, Energy, the Environment, Water (NSW), 2024, “Biodiversity Values Map”, <https://datasets.seed.nsw.gov.au/dataset/biodiversity-values-map>, accessed 7 February 2024.  Australian Government Department of Climate Change, Energy, the Environment and Water, 2024, “Australia - Species of National Environmental Significance Distributions (public grids)”, Canberra, Copyright: Commonwealth of Australia, <https://fed.dcceew.gov.au/datasets/erin::australia-species-of-national-environmental-significance-distributions-public-grids/about>, accessed 7 February 2024. |
| Crime density | NSW Bureau of Crime Statistics and Research, 2023, “Crime Tool Hotspots”, <https://www.bocsar.nsw.gov.au/Documents/Datasets/CrimeToolHotspots_JanToDec2022.zip>, downloaded on 5 February 2024. |
| Land cover | GeoVision (Precisely), Surface Cover, 2m, December 2020. Assessed accuracy was “from +/-0.5m CE90 to +/-2.5m CE90” (Geoscape, “Surface Cover: Product Description Version 1.6”, p. 15). |
| Land use data | ABS 2021, “Digital boundary files: Australian Statistical Geography Standard (ASGS) Edition 3”, <https://www.abs.gov.au/statistics/standards/australian-statistical-geography-standard-asgs-edition-3/jul2021-jun2026/access-and-downloads/digital-boundary-files> (accessed 31 July 2023). |

### S2. Service areas

Network buffers around ABS residential mesh block centroids were created using a network analysis with data from PSMA (Geoscape Australia). Motorways (but not entry and exit ramps) and bus-only transit areas were excluded from the network. Occasionally, ferry routes such as the Mortlake Ferry were included in the network. Output polygons were not trimmed.

Note in output service area polygons, perpendicular distance from roads is often smaller than the input service area size. For example, for 400m buffers, the perpendicular distance from roads may only be 20-40m, whereas the distance following the roads is 400m. This may underestimate the intersect area of mesh block network buffers with green spaces.

There may also be cases where smaller buffer sizes may have slightly larger intersect areas than respective larger buffer sizes. Due to the formation of output polygons, some parts of a 100m service area polygon for example may be slightly wider than the respective 200m service area polygon, meaning that in some cases, when green spaces are intersected, the intersect area at 200m is underestimated relative to the 100m intersect area.

### S3. Green space data screenshots

The highlighted green spaces in Figure S1 represent one single record in the Existing Green Assets data. For this reason, multipart features were split into singlepart features.

Figure S1. Presence of multiple separate green space examples in the Existing Green Assets dataset (NSW Department of Planning, Housing and Infrastructure 2019). ArcMap 10.8.2. (Esri, Redlands, CA).

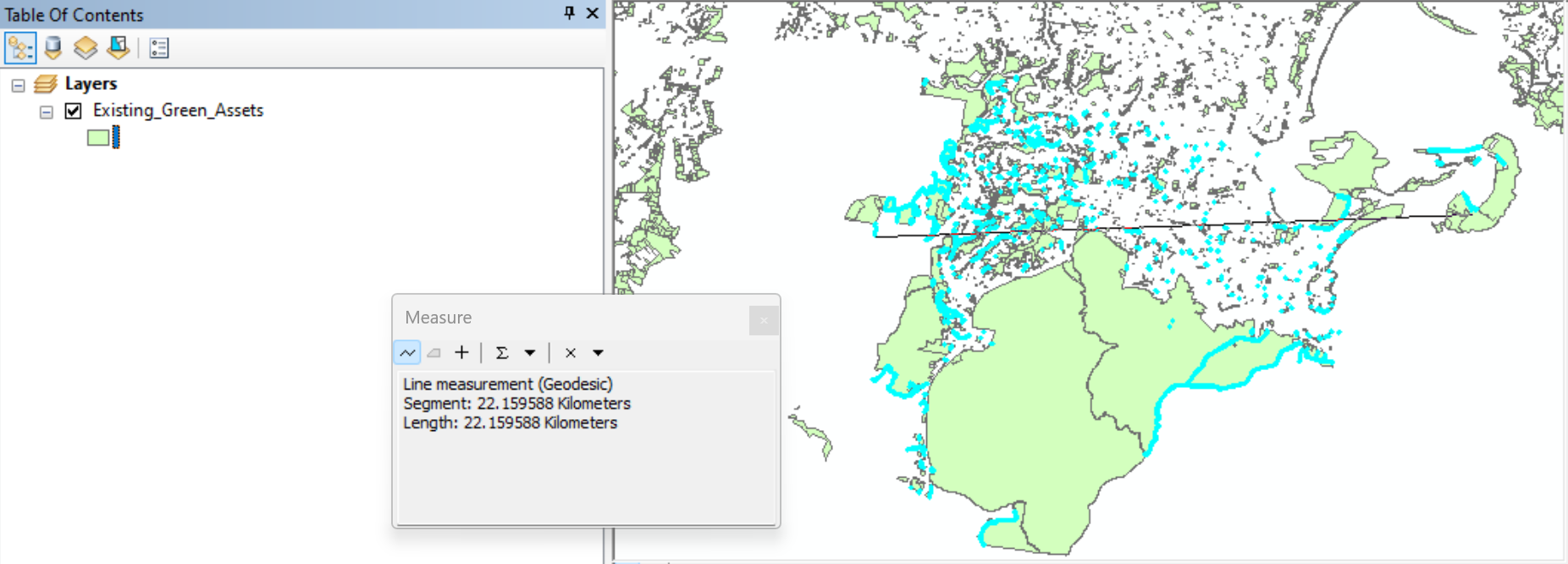

Figure S2.Image/pixel misclassification in land cover data near Bicentennial Park/Homebush. Highlighted = Bare Earth. Source: GeoVision/Precisely 2020.

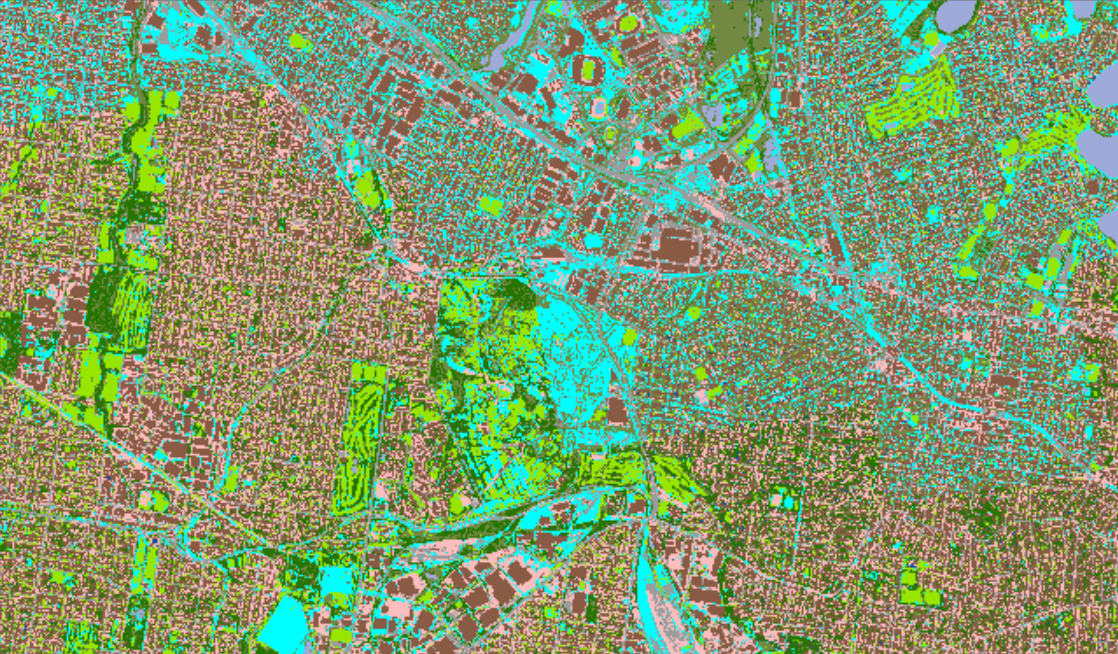

Figure S3. Image/pixel misclassification in land cover data near Bicentennial Park/Homebush. Selected Bare Earth data from Figure S2 has been converted to polygons. Source: GeoVision/Precisely 2020.

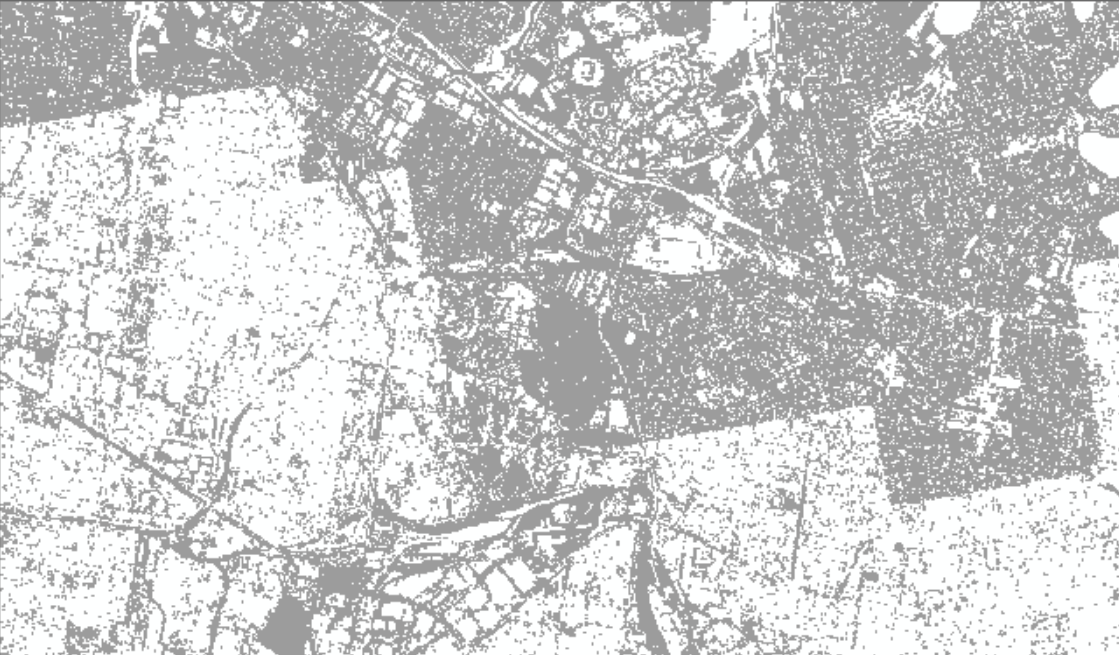

Figure S4. Example of paths lacking within green spaces in the PSMA (Geoscape) roads data set. Basemap: World Imagery basemap (Esri, Maxar, Earthstar Geographics, and the GIS User Community).

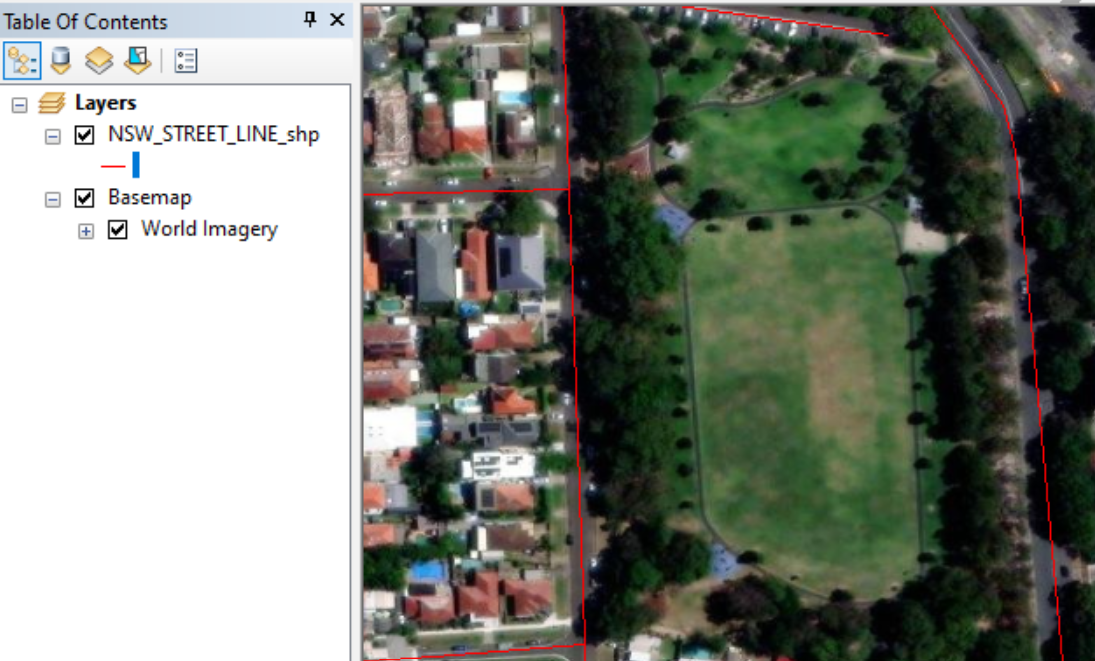

### S4. Qualities and calculations

A summary of qualities is contained in Table S2. For the aggregation method, “Count” = unweighted count, “Sum” = unweighted sum, “w0 Sum” = sum weighted by the ratio of the green space intersect with the buffer area to the green space area, “w1 Mean” = mean weighted by the ratio of the intersect area to the sum of all intersect areas, “Max” = maximum value over the intersects within the buffer, “Quality” = Value 0/1, aggregated to 1 if any intersects equal 1 (technically Quality = Max), and “Formula” = calculation of the index from other qualities that are already aggregated using a formula.

#### Aggregation

The main idea behind the aggregation can be called “an aggregated green space” which consists of disjoint parts of green spaces (intersects) located fully or partially within a mesh block buffer. The qualities of the aggregated green space should imitate a single isolated green space. There are originally sourced qualities and qualities calculated from the originals.

#### Originally sourced qualities

This approach to aggregation assumes that in the absence of additional information, the quality of a green space is spread homogenously over the area of the green space.

The first of the weighted methods of aggregation is a weighted sum over qualities (“w0 Sum”), which is a sum of qualities. For example, pathway length, which is summed over segments of pathways. If a green space was segmented, that quality would be smaller for these segments. Qualities of this type are aggregated by summing over intersects with weights equal to the ratio of a green space intersect area to the green space area. The sum of these weights does not equal one, and the maximum value of a weight can be one if the whole green space lies within the buffer.

*Example of calculation*

*Assuming that for a certain mesh block there are two intersects with green spaces.*

*Qualities of type “sum” (e.g. no. of stations) within green spaces are 10 and 20, respectively.*

*The areas of green spaces (AG) are	500 sq m and 2000 sq m.*

*The areas of intersects (AI) are	100 sq m and 200 sq m.*

*Number of stations within the intersects would be proportional to weights=AI/AG, i.e.:*

*100/500 and 200/2000.*

*Finally, the number of stations over all intersects is*

*(100/500)*10 + (200/2000)*20=*

*(1/5)*10 + (1/10)*20=*

*2 + 2= 4.*

The second method is a weighted mean over qualities (“w1 Mean”), which are means of qualities, for example, area to perimeter ratio. If a green space was segmented, that quality would be the same for all segments. Qualities of this type are aggregated by summing over intersects with weights equal to the ratio of a green space intersect area to the sum of all intersects’ areas. The sum of these weights equals one, and the maximum value of a weight can be one, only if there is a single green space intersecting the buffer.

*Example of calculation*

*Assuming as in the previous example that for a certain mesh block there are two intersects with green spaces.*

*Qualities of type “mean” (e.g., mean level of hypothetical noise^[1]^) within green spaces are 10 and 20.*

*The areas of green spaces (AG) are	500 sq m and 2000 sq m.*

*The areas of intersects (AI) are	100 sq m and 200 sq m.*

*Mean level of noise within the intersects would be the same as in the green spaces, 10 and 20.*

*But the contribution of each intersect into aggregated noise would be proportional to the area of each intersect (100 and 200), scaled by the total area of intersects equal 300:*

*100/300 and 200/300.*

*Finally, the mean over all intersects is*

*(100/300)*10 + (200/300)*20=*

*(1/3)*10 + (2/3)*20=*

*10/3 + 40/3=*

*50/3=16.7.*

A non-weighted aggregation method is applied to biodiversity and incivility qualities. These features would not be diminished if the area of the green space is segmented because fauna species for example are mobile, thus being of type “mean”. But if one intersect has a higher count of species in the aggregated green space, this count defines biodiversity. Thus, biodiversity qualities have been aggregated by taking the maximum count (“Max”) over the intersecting green spaces.

“Quality” refers to qualities for which presence/absence was noted and these are aggregated similarly to those categorised as “Max”.

There are also two trivial aggregations; number (N) of intersects within a mesh block and a simple sum of intersect areas and the respective green space areas (Sum) within a mesh block.

#### Calculated qualities

The final aggregation method (“Formula”) denoted a derivation using qualities that have already been aggregated as weighted sums or weighted means (and a simple sum for the area of intersects).

#### Exclusions

For certain green spaces there was inadequate tree cover data (qualities using tree cover data in access and land use sections), slope data and/or land use data. For tree cover and slope data, this was coded based on observations with other datasets.^[2]^ For land cover data, if the total of all land cover categories used in this research was below 80% of the green space area, land cover data was considered inadequate. If there was inadequate data for one of the intersect areas, the whole mesh block was considered to have missing data and was not included in statistical analysis for the respective qualities.

There were two calculated variables that were used as denominators for calculating percentages, *Sum of Land Use Areas within 100m of a green space* (“LU100_Area0_eq”) and *buffered roads area* (“Can100_Rds0_eq”). In these cases, if the denominators were equal to zero, the percentages were set as having a missing value. There were very few cases of this at the aggregated level, two for Can100_Rds0_eq and none for LU100_Area0_eq. These exclusions at the level of green spaces were not propagated to mesh blocks.

Table S2. Summary of qualities

| **Qualities** | **Aggregation method** | **Calculation** |
| --- | --- | --- |
| N green spaces in MB buffer | N | Count of green spaces within the network distance buffers around MB centroids. |
| Intersect of buffer w green spaces | Sum | MBs were selected from 2021 ABS Census data. X and Y centroids that are located inside a polygon were calculated. 100m, 200m, 300m, 400m, 800m and 1600m Service Area analyses were conducted using ArcMap’s Network Analyst.  The service areas for each of the MBs were intersected with the green space data. The sum of the service areas (network buffers) in square metres that intersect with green spaces were evaluated. |
| Green space area (A) | Sum | Area of green spaces (NSW Department of Planning, Housing and Infrastructure 2019; ABS 2021). |
| Green space perimeter (P) | w0 Sum | Perimeter from green space datasets (NSW Department of Planning, Housing and Infrastructure 2019; ABS 2021). Note, some green spaces are represented by multiple polygons, and we also split multipart polygons into singlepart polygons. In these cases, calculated perimeters may be larger than actual, based on these polygons. |
| Length of pathways within green space | w0 Sum | Length of PSMA Roads data coded as pathways. |
| Length of bikeways within green space | w0 Sum | Length of bikeway data from Transport for NSW (NSWBicycleNetwork5March2018.gdb). |
| Length of pathways covered by trees within green space; Length of bikeways covered by trees within green space | w0 Sum | Pathways/Bikeways intersected with the November 2021 GeoVision Trees dataset.  Green spaces that had no or partial tree canopy data covering them were also excluded from statistical analysis for these qualities. This was manually evaluated by checking against the *World Imagery* basemap (Sources: Esri, Maxar, Earthstar Geographics, and the GIS User Community) and from a dataset derived from GeoVision December 2020 2m landcover data, especially on the outskirts of the UCL boundaries. Note there are many minor occurrences where there was no data in the tree canopy dataset but the derived GeoVision landcover data had trees. This was not coded as no or partial data unless the extent of the data likely did not properly overlap the green spaces/study area.  During the aggregation of MBs, for a certain MB that contains intersects with green spaces that had no or partial data, the exclusion also extended to that MB’s intersects with green spaces that did have adequate data. Thus, the exclusion is propagated to the level of a mesh block. |
| Length of pathways greater than 6 degrees gradient within green space; Length of bikeways greater than 6 degrees gradient within green space | w0 Sum | To determine gradient, a 5m DEM raster from Geoscience Australia was used. Esri’s ArcMap’s Slope, Reclassify and Raster to Polygon conversion tools were used. Length of pathways/bikeways greater than six degrees were evaluated.  Green spaces that had no or partial data for gradient were excluded from statistical analysis for these qualities. Records with “No data” were evaluated through data selections. Records with “Partial data” were manually evaluated. During the aggregation of MBs, for a certain MB that contains intersects with green spaces that had no or partial data, the exclusion also extended to that MB’s intersects with green spaces that did have adequate data. Thus, the exclusion is propagated to the level of a mesh block. |
| Number of bus stops within and surrounding a green space (within 100m buffer); Number of train or light rail stations within and surrounding a green space (within 100m buffer); Number of ferry wharves within and surrounding a green space (within 100m buffer) | w0 Sum | The number of stop locations (Transport for NSW) that are within a 100m radius (including input polygon) of each green space. |
| Area of green space greater than 6 degrees gradient | w0 Sum | To determine gradient, a 5m DEM raster from Geoscience Australia was used. Esri’s ArcMap’s Slope, Reclassify and Raster to Polygon conversion tools were used. Area of green space greater than six degrees was calculated.  Green spaces that had no or partial data for gradient were excluded from statistical analysis for these qualities. Records with “No data” were evaluated through data selections. Records with “Partial data” were manually evaluated. During the aggregation of MBs, for a certain MB that contains intersects with green spaces that had no or partial data, the exclusion also extended to that MB’s intersects with green spaces that did have adequate data. Thus, the exclusion is propagated to the level of a mesh block. |
| Length of main roads surrounding a green space (within 100m doughnut buffer) | w0 Sum | Length of main roads (PSMA, Geoscape Australia) that are within a 100m radius (excluding input polygon) of each green space. |
| Ratio of Area(A) to Perimeter (P) | w1 Mean | Green space area divided by Perimeter. |
| Ratio of Perimeter (P) to perimeter standardised by area (A) | w1 Mean | To assess the shape of the green space we propose to calculate a perimeter of the perfect circle given area of the green space. We call it a standardized perimeter (Pstd). The index of shape is ratio of P/Pstd. Its minimum value = 1 when it is a perfect circle. The higher the index the more the shape differs from a perfect circle. |
| Total number of transport stops/stations/wharves within and surrounding a GS (within 100m buffer) | Formula | Total number of all stop/station/wharf locations (Transport for NSW) that are within a 100m radius (including input polygon) of each green space. |
| Pathway Length per Area of green space; Bikeway Length per Area of green space | Formula | Length of pathways/bikeways within green space divided by green space area. |
| Trees over Pathway per Area of green space; Trees over Bikeway per Area of GS | Formula | Length of pathways/bikeways covered by trees within green space divided by Area of green space.  Green spaces that had no or partial tree canopy data covering them were also excluded from statistical analysis for these qualities. This was manually evaluated by checking against the *World Imagery* basemap (Sources: Esri, Maxar, Earthstar Geographics, and the GIS User Community) and from a dataset derived from GeoVision December 2020 2m landcover data, especially on the outskirts of the UCL boundaries. Note there are many minor occurrences where there was no data in the tree canopy dataset but the derived GeoVision landcover data had trees. This was not coded as no or partial data unless the extent of the data likely did not properly overlap the green spaces/study area.  During the aggregation of MBs, for a certain MB that contains intersects with green spaces that had no or partial data, the exclusion also extended to that MB’s intersects with green spaces that did have adequate data. Thus, the exclusion is propagated to the level of a mesh block. |
| Percentage of green space greater than 6 degrees gradient | Formula | Area of green space greater than 6 degrees gradient divided by Green Space Area, multiplied by 100.  Green spaces that had no or partial data for gradient were excluded from statistical analysis for these qualities. Records with “No data” were evaluated through data selections. Records with “Partial data” were manually evaluated. During the aggregation of MBs, for a certain MB that contains intersects with green spaces that had no or partial data, the exclusion also extended to that MB’s intersects with green spaces that did have adequate data. Thus, the exclusion is propagated to the level of a mesh block. |
| Percentage of pathways greater than 6 degrees gradient within green space; Percentage of bikeways greater than 6 degrees gradient within green space | Formula | Length of pathways greater than 6 degrees gradient within green space divided by Green Space Area.  Green spaces that had no or partial data for gradient were excluded from statistical analysis for these qualities. Records with “No data” were evaluated through data selections. Records with “Partial data” were manually evaluated. During the aggregation of MBs, for a certain MB that contains intersects with green spaces that had no or partial data, the exclusion also extended to that MB’s intersects with green spaces that did have adequate data. Thus, the exclusion is propagated to the level of a mesh block. |
| Number of public toilet facilities within and surrounding a GS (within 100m buffer) | w0 Sum | Data for public toilets was from The National Public Toilet Map, Department of Health and Aged Care. Counts within 100m radii (including input polygon) around green spaces were calculated. |
| Number of cafes within and surrounding a green space (within 100m buffer); Number of hotels/bars within and surrounding a green space (within 100m buffer); Number of restaurants/takeaways within and surrounding a green space (within 100m buffer); Number of restaurants within and surrounding a green space (within 100m buffer); Number of takeaways within and surrounding a green space (within 100m buffer); Number of supermarkets/ greengrocers within and surrounding a green space (within 100m buffer) | w0 Sum | Locations were from Sensis data (Thryv Australia). Counts within 100m radii (including input polygon) around green spaces were calculated.  Records were not considered if the ABN Status was listed as “Cancelled.”  Pizza shops and fish and seafoods retail were both considered here to be takeaways, though there may be pizza shops that were restaurants or fish and seafoods retail that sell fresh seafood.  Some fast-food chains were listed as restaurants or cafes. This classification was changed to takeaways using the following selection (example selection for Sydney):  "Business_N" LIKE 'DONUT KING%' OR ("Business_N" LIKE 'GRILL%' AND "Business_N" NOT LIKE 'GRILLS%' AND "Business_N" NOT LIKE 'GRILL %' AND "Business_N" NOT LIKE 'GRILLI%' AND "Business_N" NOT LIKE 'GRILLE%') OR "Business_N" LIKE 'HUNGRY JACK%' OR "Business_N" LIKE 'KFC%' OR ("Business_N" LIKE 'MCDONALD%' AND "Business_N" NOT LIKE 'MCDONALD L%' AND "Business_N" NOT LIKE 'MCDONALD J%' AND "Business_N" NOT LIKE 'MCDONALD,%') OR ("Business_N" LIKE 'NANDO%' AND "Business_N" NOT LIKE '%QUALITY MEATS') OR "Business_N" LIKE 'PIZZA HUT%' OR "Business_N" LIKE 'DOMINO%' OR "Business_N" LIKE 'RED ROOSTER%' OR "Business_N" LIKE 'SUBWAY%' OR ("Business_N" LIKE 'SUSHI%' AND "Business_N" NOT LIKE '%TRAIN%' AND "Business_N" NOT LIKE '%RESTAURANT%') OR "Business_N" LIKE 'OPORTO%' OR "Business_N" LIKE 'GUZMAN%' OR "Business_N" LIKE 'ZAMBRERO%' |
| Presence of parkrun event | Quality | The parkrun website, <https://www.parkrun.com.au/events/events/> was checked for the presence of a parkrun event. Course maps were checked for each event and if the course went through a green space polygon, that polygon was coded as “Y”. Remaining polygons were coded as “N”.  The parkrun course information pages use data from: Google MyMaps. Imagery © 2024, TerraMetrics, AEROmetrex & Jacobs, Airbus, CNES / Airbus, Landsat / Copernicus, Maxar Technologies, Map data © 2024 Google.  Note the *World Street Map* (Sources: Esri, HERE, Garmin, USGS, Intermap, INCREMENT P, NRCan, Esri Japan, METI, Esri China (Hong Kong), Esri Korea, Esri (Thailand), NGCC, © OpenStreetMap contributors, and the GIS User Community.), *World Imagery* (Sources: Esri, Maxar, Earthstar Geographics, and the GIS User Community), and *World Boundaries and Places* (Sources: Esri, HERE, Garmin, (c) OpenStreetMap contributors, and the GIS user community) were used in ArcMap. |
| Presence of coastline | Quality | A coastline dataset was created from the States and Territories shapefile, “STE_2021_AUST_2020MGA56.shp” – ABS 2021, “Digital boundary files: Australian Statistical Geography Standard (ASGS) Edition 3”, <https://www.abs.gov.au/statistics/standards/australian-statistical-geography-standard-asgs-edition-3/jul2021-jun2026/access-and-downloads/digital-boundary-files> (accessed 31 July 2023).  Using Esri’s ArcMap 10.8.2, a polygon shapefile which was approximately a rectangle shape that extended over the coastline of NSW was manually created. The Erase tool was used with the rectangle-shaped polygon as input and the STE_2021_AUST_2020MGA56.shp as the Erase Feature to create the coastline shapefile.  The Select Layer by Location tool (Intersect) was used for the green spaces as input and the coastline shapefile as the selecting features to determine if the green space had the presence of a coastline. |
| Area of beach within and surrounding a green space (within 100m buffer) | w0 Sum | Data used was from: “gis_osm_natural_a_free_1.shp” – Geofabrik GmbH and OpenStreetMap Contributors 2022, “Australia.shp.zip [australia-latest-free.shp.zip]” downloaded from https://download.geofabrik.de/australia-oceania/australia-latest-free.shp.zip, accessed 12 December 2022  Records that had "fclass" LIKE 'beach' were selected from gis_osm_natural_a_free_1.shp.  A 100m buffer around green spaces (including input polygon) was intersected with beaches to calculate the area of beaches within 100m of green spaces. |
| Number of bird species/species habitat likely to occur within green space; Number of fish species/species habitat likely to occur within green space; Number of flora species/species habitat likely to occur within green space; Number of frog species/species habitat likely to occur within green space; Number of mammal species/species habitat likely to occur within green space; Number of other animal species/species habitat likely to occur within green space; Number of reptile species/species habitat likely to occur within green space | Max | Number of species/species habitat likely to occur was sourced from “Species of National Environmental Significance Distributions” (Australian Government Department of Climate Change, Energy, the Environment and Water).  Data relates to species protected under the *Environment Protection and Biodiversity Conservation Act 1999* and may not include other species not listed under this Act. Note that the counts data were not green space-specific but were for either 1km grid cells or 10km grid cells. Data show indicative presence that a “Species or species habitat likely to occur” or a “Species or species habitat may occur” and individual green spaces within a given grid cell may or may not have that species/taxonomic group actually present. For this research, only records with presence coded as “Species or species habitat likely to occur” were considered.  It is noted that biodiversity counts for grid cells are not likely randomly distributed but are assumed to be clustered within green spaces.  The biodiversity counts data was spatially joined with the green space locations data. Python scripts were then run to check for duplicate species records and for counts per green space based on taxonomic groups. |
| Area of land with biodiversity value within green space; Area of land with biodiversity value outside of green space within 100m | w0 Sum | Data was from NSW Department of Climate Change, Energy, the Environment, Water (NSW), 2024, “Biodiversity Values Map”, https://datasets.seed.nsw.gov.au/dataset/biodiversity-values-map, accessed 7 February 2024.  This data includes land considered to have high biodiversity value. This includes classes such as 'Biodiverse riparian land', 'Coastal Management Act - Littoral Rainforest', 'Coastal Management Act - Wetlands', 'Core Habitat within an approved Koala Plan of Management (Koala SEPP)', 'Declared Area of Outstanding Biodiversity Value', 'Ramsar Wetlands', and 'Threatened species or communities with potential for serious and irreversible impacts'. These areas of high biodiversity value are considered to be “particularly sensitive to impacts from development” (NSW Department of Climate Change, Energy, the Environment, Water (NSW), “The Biodiversity Values Map and Threshold Tool”, <https://www2.environment.nsw.gov.au/topics/animals-and-plants/biodiversity-offsets-scheme/clear-and-develop-land/biodiversity-values-map-and-threshold-tool>, accessed 26 March 2024).  The Biodiversity Values dataset was intersected with green space locations and the intersecting area within green spaces were identified.  Additionally, area of high biodiversity value within 100m radii (excluding input polygon) around green spaces were calculated. |
| Total number of species/species habitat likely to occur within green space | Formula | Sum of bird, fish, flora, frog, mammal, other animal and reptile species/species habitat likely to occur. |
| Shannon diversity index; Simpson diversity index | Formula | Shannon diversity index:  $H=-\sum_{i=1}^{s} p_{i}{\ln p}_{i}$  Simpson diversity index:  $D=1-\sum_{i=1}^{s} {p_{i}}^{2}$  Where *p*_i_ is the proportion *n*_i_/*N* of the species within one group divided by the total number of species in all groups, *ln* is the natural logarithms, *s* is the number of the species’ groups, equal to 7 here (birds, fishes, flora, frogs, mammals, reptiles and other animals). |
| Percentage of land with biodiversity value within green space; Percentage of land with biodiversity value outside of green space within 100m | Formula | Biodiversity values land within a green space as a percentage of green space area was evaluated.  Percentage of land with biodiversity value outside of green space within 100m was calculated by dividing by the 100m doughnut buffer area and multiplying by 100. |
| Density of alcohol-related assaults; Density of malicious damage; Density of non-domestic violence assaults; Density of robberies; Density of steal from persons | Max | Data for density of incivilities were obtained from the Bureau of Crime Statistics and each intersected with green spaces. Note that the incivility data is not green space-specific data and individual green spaces may or may not have had the particular incivilities occurring within them. Crime density for each type were coded from zero to three (0 = no value, 1 = “Low Density”, 2 = “Medium Density”, 3 = “High Density”). The maximum value was chosen for a green space if there were multiple intersecting crime values.  It is noted that crime hotspot data do not show counts but the density of incivilities. This means that hotspots can occur in areas with relatively low incivilities if these are concentrated in a particular location. Conversely, there will not be hotspots in areas with high counts of incivilities if these are more geographically dispersed across the area (Melissa Burgess 2011, “Understanding crime hotspot maps”, NSW BOCSAR, <https://bocsar.nsw.gov.au/content/dam/dcj/bocsar/documents/publications/bb/bb01-100/bb60.pdf>, accessed 3 June 2025). |
| Mean level of incivilities | Formula | The mean of all incivility types was evaluated. |
| Area of bare earth within green space; Area of buildings within green space; Area of built up area within green space; Area of open grass within green space; Area of other vegetation within green space; Area of roads/paths within green space; Area of shadow within green space; Area of swimming pools within green space; Area of tree canopy within green space; Area of water within green space | w0 Sum | GeoVision (Precisely), Surface Cover, 2m, December 2020 was analysed for the three study areas. Areas of each land cover type for green spaces were identified, including bare earth, buildings, built up areas, open grass, other vegetation, roads/paths, shadows, swimming pools, tree canopy and water.  Firstly, the Mosaic to New Raster tool was used in ArcMap 10.8.2 for images that intersect with a 10km buffer around the respective city’s Urban Centre and Locality boundary. Each of the land cover types except for cloud cover were then extracted using the Raster to Polygon conversion tool.  Each of these land cover type polygons were then intersected with the green space locations to determine the area of each land cover type.  When the areas of each of the land cover types are added, they often to do not exactly equal the green space area. In some circumstances, there was only partial or no land cover data for certain green spaces. As such, if the sum of each of the percentages of land cover classes was smaller than 80%, then the land cover data was considered to be partial or no data for that particular green space. The green space and its associated mesh block intersect areas were excluded from statistical analysis for these qualities. During the aggregation of MBs, for a certain MB that contains intersects with green spaces that had no or partial data, the exclusion also extended to that MB’s intersects with green spaces that did have adequate data. Thus, the exclusion is propagated to the level of a mesh block. |
| Percentage of bare earth within green space; Percentage of buildings within green space; Percentage of built up area within green space; Percentage of open grass within green space; Percentage of other vegetation within green space; Percentage of roads/paths within green space; Percentage of shadow within green space; Percentage of swimming pools within green space; Percentage of tree canopy within green space; Percentage of water within green space | Formula | Land Cover class divided by the Green Space Area, multiplied by 100.  If the sum of each of the percentages of land cover classes was smaller than 80%, then the land cover data was considered to be partial or no data for that particular green space. The green space and its associated mesh block intersect areas were excluded from statistical analysis for these qualities. During the aggregation of MBs, for a certain MB that contains intersects with green spaces that had no or partial data, the exclusion also extended to that MB’s intersects with green spaces that did have adequate data. Thus, the exclusion is propagated to the level of a mesh block. |
| Area of commercial land use surrounding green space (within 100m doughnut buffer); Area of education land use surrounding green space (within 100m doughnut buffer); Area of hospital/medical land use surrounding green space (within 100m doughnut buffer); Area of industrial land use surrounding green space (within 100m doughnut buffer); Area of other land use surrounding green space (within 100m doughnut buffer); Area of parkland land use surrounding green space (within 100m doughnut buffer); Area of primary production land use surrounding green space (within 100m doughnut buffer); Area of residential land use surrounding green space (within 100m doughnut buffer); Area of transport land use surrounding green space (within 100m doughnut buffer); Area of water land use surrounding green space (within 100m doughnut buffer) | w0 Sum | ABS Census 2021 (mesh block category) data was used to determine area of land uses within 100m of green spaces (excluding input polygon). |
| Area of tree canopy surrounding green space (within 100m doughnut buffer); Area of tree canopy ≤4m surrounding green space (within 100m doughnut buffer); Area of tree canopy >4m surrounding green space (within 100m doughnut buffer) | w0 Sum | Area of tree canopy from GeoVision (Precisely) November 2021 data that was within 100m of a green space (excluding input polygon) was calculated. Areas of tree canopy ≤4m and >4m were separately calculated.  Green spaces that had no or partial tree canopy data covering them were excluded from statistical analysis for these qualities as it was likely that areas within 100m radii also had no or partial data. This was manually evaluated by checking against the *World Imagery* basemap (Sources: Esri, Maxar, Earthstar Geographics, and the GIS User Community) and from a dataset derived from GeoVision December 2020 2m landcover data, especially on the outskirts of the UCL boundaries. Note there are many minor occurrences where there was no data in the tree canopy dataset but the derived GeoVision landcover data had trees. This was not coded as no or partial data unless the extent of the data likely did not properly overlap the green spaces/study area.  During the aggregation of MBs, for a certain MB that contains intersects with green spaces that had no or partial data, the exclusion also extended to that MB’s intersects with green spaces that did have adequate data. Thus, the exclusion is propagated to the level of a mesh block. |
| Percentage of commercial land use surrounding green space (within 100m doughnut buffer); Percentage of education land use surrounding green space (within 100m doughnut buffer); Percentage of hospital/medical land use surrounding green space (within 100m doughnut buffer); Percentage of industrial land use surrounding green space (within 100m doughnut buffer); Percentage of other land use surrounding green space (within 100m doughnut buffer); Percentage of parkland land use surrounding green space (within 100m doughnut buffer); Percentage of primary production land use surrounding green space (within 100m doughnut buffer); Percentage of residential land use surrounding green space (within 100m doughnut buffer); Percentage of transport land use surrounding green space (within 100m doughnut buffer); Percentage of water land use surrounding green space (within 100m doughnut buffer) | Formula | Respective Land Use Area divided by the Sum of Land Use Areas within 100m of a green space (excluding input polygon).  If the denominators were equal to zero, the percentages were set as having a missing value. There were no cases at the aggregated level. These exclusions at the level of green spaces were not propagated to mesh blocks. |
| Percentage of tree canopy to roads surrounding green space (within 100m doughnut buffer); Percentage of tree canopy ≤4m to roads surrounding green space (within 100m doughnut buffer); Percentage of tree canopy >4m to roads surrounding green space (within 100m doughnut buffer) | Formula | A 10m buffer around all roads from PSMA (Geoscape Australia) was created. Trees that intersected with the buffered roads dataset were used to determine tree canopy coverage of areas within 100m of green spaces (excluding input polygon). This was used as a proxy for street trees but may also include trees on private property. This was calculated by dividing the area of tree canopy by the buffered roads area and multiplying by 100.  Percentages of tree canopy that was ≤4m and >4m were separately calculated.  If the denominator (buffered roads area) was equal to zero, the percentages were set as having a missing value. There were two cases at the aggregated level. These exclusions at the level of green spaces were not propagated to mesh blocks.  Green spaces that had no or partial tree canopy data covering them were excluded from statistical analysis for these qualities as it was likely that areas within 100m radii also had no or partial data. This was manually evaluated by checking against the *World Imagery* basemap (Sources: Esri, Maxar, Earthstar Geographics, and the GIS User Community) and from a dataset derived from GeoVision December 2020 2m landcover data, especially on the outskirts of the UCL boundaries. Note there are many minor occurrences where there was no data in the tree canopy dataset but the derived GeoVision landcover data had trees. This was not coded as no or partial data unless the extent of the data likely did not properly overlap the green spaces/study area.  During the aggregation of MBs, for a certain MB that contains intersects with green spaces that had no or partial data, the exclusion also extended to that MB’s intersects with green spaces that did have adequate data. Thus, the exclusion is propagated to the level of a mesh block. |
| Percentage of tree canopy surrounding green space (within 100m doughnut buffer); Percentage of tree canopy ≤4m surrounding green space (within 100m doughnut buffer); Percentage of tree canopy >4m surrounding green space (within 100m doughnut buffer) | Formula | Area of tree canopy divided by the doughnut buffer area, multiplied by 100.  Green spaces that had no or partial tree canopy data covering them were excluded from statistical analysis for these qualities as it was likely that areas within 100m radii also had no or partial data. This was manually evaluated by checking against the *World Imagery* basemap (Sources: Esri, Maxar, Earthstar Geographics, and the GIS User Community) and from a dataset derived from GeoVision December 2020 2m landcover data, especially on the outskirts of the UCL boundaries. Note there are many minor occurrences where there was no data in the tree canopy dataset but the derived GeoVision landcover data had trees. This was not coded as no or partial data unless the extent of the data likely did not properly overlap the green spaces/study area.  During the aggregation of MBs, for a certain MB that contains intersects with green spaces that had no or partial data, the exclusion also extended to that MB’s intersects with green spaces that did have adequate data. Thus, the exclusion is propagated to the level of a mesh block. |

### S4. Summary results tables

Table S3. Spearman's rank coefficient of correlation between SEIFA IRSD score (reversed) and aggregated over MB green space features – n, Rho and compressed P-values – 100m and 200m buffers.

|  |  | **Buffer 100m (n)** | | | | **Buffer 200m (n)** | | | | **Buffer 100m (Rho and P-value)** | | | | **Buffer 200m (Rho and P-value)** | | | |
| --- | --- | --- | --- | --- | --- | --- | --- | --- | --- | --- | --- | --- | --- | --- | --- | --- | --- |
| **Index description** | **Method** | **Full set** | **Syd** | **New** | **Wol** | **Full set** | **Syd** | **New** | **Wol** | **Full set** | **Syd** | **New** | **Wol** | **Full set** | **Syd** | **New** | **Wol** |
| **General indices** | | | | | | | | | | | | | | | | | |
| N green spaces in MB | Count | 11030 | 9945 | 612 | 473 | 23449 | 20933 | 1419 | 1097 | -0.09* | -0.09* | -0.01 | -0.04 | -0.10* | -0.09* | -0.05+ | 0.05 |
| Intersect of buffer with green space | Sum | 11030 | 9945 | 612 | 473 | 23449 | 20933 | 1419 | 1097 | -0.02× | -0.03× | -0.10+ | 0.03 | -0.02+ | -0.03* | -0.06+ | 0.04 |
| Green space area (A) | Sum | 11030 | 9945 | 612 | 473 | 23449 | 20933 | 1419 | 1097 | 0.01 | -0.01 | -0.03 | -0.03 | 0.01 | -0.02× | -0.06+ | 0.01 |
| Green space perimeter (P) | w0 Sum | 11030 | 9945 | 612 | 473 | 23449 | 20933 | 1419 | 1097 | -0.07* | -0.06* | -0.09+ | 0.04 | -0.07* | -0.06* | -0.05+ | 0.04 |
| **Access indices – original sourced** | | | | | | | | | | | | | | | | | |
| Length of pathways within green space | w0 Sum | 11030 | 9945 | 612 | 473 | 23449 | 20933 | 1419 | 1097 | 0 | -0.01 | -0.12× | 0.10+ | 0 | 0 | -0.15* | 0.12* |
| Length of bikeways within green space | w0 Sum | 11030 | 9945 | 612 | 473 | 23449 | 20933 | 1419 | 1097 | 0.02+ | 0 | -0.05 | 0.04 | 0.02* | 0 | -0.05 | 0.07+ |
| Length of pathways covered by trees within green space | w0 Sum | 10976 | 9911 | 601 | 464 | 23301 | 20841 | 1388 | 1072 | -0.03× | -0.03+ | -0.14* | 0.04 | -0.03* | -0.02* | -0.18* | 0.08× |
| Length of bikeways covered by trees within green space | w0 Sum | 10976 | 9911 | 601 | 464 | 23301 | 20841 | 1388 | 1072 | 0 | -0.01 | -0.09+ | 0.05 | 0 | -0.01 | -0.06+ | 0.06 |
| Length of pathways greater than 6 degrees gradient within green space | w0 Sum | 10997 | 9913 | 612 | 472 | 23390 | 20879 | 1418 | 1093 | -0.13* | -0.14* | -0.19* | -0.03 | -0.13* | -0.14* | -0.22* | -0.01 |
| Length of bikeways greater than 6 degrees gradient within green space | w0 Sum | 10997 | 9913 | 612 | 472 | 23390 | 20879 | 1418 | 1093 | -0.03× | -0.04* | -0.12× | -0.09 | -0.04* | -0.05* | -0.12* | -0.04 |
| Number of Railway Stations within and surrounding a green space (within 100m buffer) | w0 Sum | 11030 | 9945 | 612 | 473 | 23449 | 20933 | 1419 | 1097 | -0.01 | -0.01 | 0 | -0.01 | -0.01 | 0 | -0.02 | -0.05 |
| Number of Bus Stops within and surrounding a green space (within 100m buffer) | w0 Sum | 11030 | 9945 | 612 | 473 | 23449 | 20933 | 1419 | 1097 | 0 | 0 | -0.04 | 0.07 | -0.01+ | -0.02+ | -0.06+ | 0.12* |
| Number of ferry wharves within and surrounding a green space (within 100m buffer) | w0 Sum | 11030 | 9945 | 612 | 473 | 23449 | 20933 | 1419 | 1097 | -0.09* | -0.09* | 0.08+ |  | -0.09* | -0.09* | 0.05+ |  |
| Area of GS which is greater than 6 degrees of Slope | w0 Sum | 10997 | 9913 | 612 | 472 | 23390 | 20879 | 1418 | 1093 | -0.31* | -0.33* | -0.26* | -0.28* | -0.29* | -0.31* | -0.24* | -0.25* |
| Length of Main Roads within 100 metres of the Green Space | w0 Sum | 11030 | 9945 | 612 | 473 | 23449 | 20933 | 1419 | 1097 | -0.02+ | -0.02 | 0.09+ | 0.02 | -0.03* | -0.03* | 0.07× | 0.10* |
| **Access indices – calculated** | | | | | | | | | | | | | | | | | |
| Ratio of Area(A) to Perimeter (P) | w1 Mean | 11030 | 9945 | 612 | 473 | 23449 | 20933 | 1419 | 1097 | 0.08* | 0.05* | 0 | -0.02 | 0.08* | 0.05* | -0.02 | 0 |
| Ratio of Perimeter (P) to perimeter standardised by area (A) | w1 Mean | 11030 | 9945 | 612 | 473 | 23449 | 20933 | 1419 | 1097 | -0.13* | -0.11* | -0.16* | -0.10+ | -0.13* | -0.12* | -0.15* | -0.08+ |
| Total number of transport stops/stations/wharves within and surrounding a green space (within 100m buffer) | Formula | 11030 | 9945 | 612 | 473 | 23449 | 20933 | 1419 | 1097 | 0 | 0 | -0.05 | 0.07 | -0.02× | -0.02× | -0.07+ | 0.12* |
| Pathway Length per Area of Green Space | Formula | 11030 | 9945 | 612 | 473 | 23449 | 20933 | 1419 | 1097 | 0 | 0 | -0.12× | 0.10+ | 0 | 0 | -0.16* | 0.12* |
| Bikeway Length per Area of Green Space | Formula | 11030 | 9945 | 612 | 473 | 23449 | 20933 | 1419 | 1097 | 0.02+ | 0 | -0.04 | 0.03 | 0.03* | 0.01 | -0.02 | 0.06+ |
| Trees over Pathway per Area of Green Space | Formula | 10976 | 9911 | 601 | 464 | 23301 | 20841 | 1388 | 1072 | -0.03× | -0.03+ | -0.13× | 0.03 | -0.03* | -0.02* | -0.19* | 0.07+ |
| Trees over Bikeway per Area of Green Space | Formula | 10976 | 9911 | 601 | 464 | 23301 | 20841 | 1388 | 1072 | 0 | -0.01 | -0.07 | 0.03 | 0 | -0.01 | -0.06+ | 0.04 |
| Percentage of GS which is greater than 6 degrees of Slope | Formula | 10997 | 9913 | 612 | 472 | 23390 | 20879 | 1418 | 1093 | -0.41* | -0.42* | -0.35* | -0.41* | -0.40* | -0.42* | -0.33* | -0.41* |
| Pathway Length > 6 degrees per Area of GS | Formula | 10997 | 9913 | 612 | 472 | 23390 | 20879 | 1418 | 1093 | -0.14* | -0.15* | -0.21* | -0.03 | -0.14* | -0.14* | -0.23* | -0.02 |
| Bikeway Length > 6 degrees slope per Area of GS | Formula | 10997 | 9913 | 612 | 472 | 23390 | 20879 | 1418 | 1093 | -0.03× | -0.04* | -0.13* | -0.10+ | -0.04* | -0.05* | -0.12* | -0.05 |
| **Amenities/Activities indices – original sourced** | | | | | | | | | | | | | | | | | |
| Number of public toilet facilities within and surrounding a green space (within 100m buffer) | w0 Sum | 11030 | 9945 | 612 | 473 | 23449 | 20933 | 1419 | 1097 | -0.05* | -0.07* | 0.04 | 0.07 | -0.05* | -0.06* | -0.03 | 0.07+ |
| Number of cafes within and surrounding a green space (within 100m buffer) | w0 Sum | 11030 | 9945 | 612 | 473 | 23449 | 20933 | 1419 | 1097 | -0.09* | -0.09* | -0.12× | 0.02 | -0.11* | -0.10* | -0.14* | 0.03 |
| Number of hotels/bars within and surrounding a green space (within 100m buffer) | w0 Sum | 11030 | 9945 | 612 | 473 | 23449 | 20933 | 1419 | 1097 | -0.03× | -0.02+ | -0.04 |  | -0.04* | -0.04* | -0.06+ | 0.03 |
| Number of restaurants/takeaways within and surrounding a green space (within 100m buffer) | w0 Sum | 11030 | 9945 | 612 | 473 | 23449 | 20933 | 1419 | 1097 | -0.06* | -0.06* | -0.04 | -0.04 | -0.06* | -0.05* | -0.02 | -0.04 |
| Number of restaurants within and surrounding a green space (within 100m buffer) | w0 Sum | 11030 | 9945 | 612 | 473 | 23449 | 20933 | 1419 | 1097 | -0.11* | -0.11* | -0.10+ | -0.10+ | -0.11* | -0.10* | -0.09* | -0.09× |
| Number of takeaways within and surrounding a green space (within 100m buffer) | w0 Sum | 11030 | 9945 | 612 | 473 | 23449 | 20933 | 1419 | 1097 | 0.01 | 0.02 | 0.04 | 0.06 | 0.01 | 0.01+ | 0.02 | 0.05 |
| Number of supermarkets/greengrocers within and surrounding a green space (within 100m buffer) | w0 Sum | 11030 | 9945 | 612 | 473 | 23449 | 20933 | 1419 | 1097 | 0.02+ | 0.02 | 0.04 | 0.12× | 0.02+ | 0.02× | 0.01 | 0.10* |
| Presence of parkrun event | Quality | 11030 | 9945 | 612 | 473 | 23449 | 20933 | 1419 | 1097 | -0.01 | -0.01 | -0.10+ | -0.08 | -0.02* | -0.02× | -0.08× | -0.11* |
| **Beach/Coast indices – original sourced** | | | | | | | | | | | | | | | | | |
| Presence of coastline | Quality | 11030 | 9945 | 612 | 473 | 23449 | 20933 | 1419 | 1097 | -0.25* | -0.29* | -0.04 | -0.07 | -0.25* | -0.28* | -0.07× | -0.07+ |
| Area of beach within and surrounding a green space (within 100m buffer) | w0 Sum | 11030 | 9945 | 612 | 473 | 23449 | 20933 | 1419 | 1097 | -0.16* | -0.18* | -0.14* | -0.23* | -0.15* | -0.17* | -0.11* | -0.20* |
| **Biodiversity indices – original sourced** | | | | | | | | | | | | | | | | | |
| Number of bird species/species habitat likely to occur within green space | Max | 11030 | 9945 | 612 | 473 | 23449 | 20933 | 1419 | 1097 | -0.30* | -0.34* | 0.02 | 0.15× | -0.29* | -0.34* | 0 | 0.13* |
| Number of fish species/species habitat likely to occur within green space | Max | 11030 | 9945 | 612 | 473 | 23449 | 20933 | 1419 | 1097 | -0.32* | -0.36* | -0.09+ | 0.02 | -0.31* | -0.35* | -0.11* | 0 |
| Number of flora species/species habitat likely to occur within green space | Max | 11030 | 9945 | 612 | 473 | 23449 | 20933 | 1419 | 1097 | -0.28* | -0.36* | 0.05 | 0.28* | -0.27* | -0.35* | 0.04 | 0.29* |
| Number of frog species/species habitat likely to occur within green space | Max | 11030 | 9945 | 612 | 473 | 23449 | 20933 | 1419 | 1097 | -0.27* | -0.31* | 0.08 | -0.14× | -0.26* | -0.29* | 0.06+ | -0.20* |
| Number of mammal species/species habitat likely to occur within green space | Max | 11030 | 9945 | 612 | 473 | 23449 | 20933 | 1419 | 1097 | -0.51* | -0.55* | -0.18* | -0.30* | -0.51* | -0.55* | -0.18* | -0.30* |
| Number of other animal species/species habitat likely to occur within green space | Max | 11030 | 9945 | 612 | 473 | 23449 | 20933 | 1419 | 1097 | -0.26* | -0.26* |  |  | -0.28* | -0.28* |  |  |
| Number of reptile species/species habitat likely to occur within green space | Max | 11030 | 9945 | 612 | 473 | 23449 | 20933 | 1419 | 1097 | -0.35* | -0.39* | -0.09+ | -0.02 | -0.34* | -0.39* | -0.12* | -0.06+ |
| Area of land with biodiversity value within green space | w0 Sum | 11030 | 9945 | 612 | 473 | 23449 | 20933 | 1419 | 1097 | -0.01 | -0.02 | 0.07 | -0.01 | 0 | 0 | 0.06+ | -0.01 |
| Area of land with biodiversity value outside of green space within 100m | w0 Sum | 11030 | 9945 | 612 | 473 | 23449 | 20933 | 1419 | 1097 | -0.03* | -0.04* | 0.08+ | -0.01 | -0.02× | -0.02* | 0.07× | -0.02 |
| **Biodiversity indices – calculated** | | | | | | | | | | | | | | | | | |
| Total number of species/species habitat likely to occur within green space | Formula | 11030 | 9945 | 612 | 473 | 23449 | 20933 | 1419 | 1097 | -0.48* | -0.54* | -0.05 | 0.13× | -0.48* | -0.54* | -0.05+ | 0.11* |
| Shannon diversity index | Formula | 11030 | 9945 | 612 | 473 | 23449 | 20933 | 1419 | 1097 | -0.43* | -0.47* | -0.18* | -0.28* | -0.42* | -0.46* | -0.21* | -0.29* |
| Simpson diversity index (range 0 to 1=most diverse) | Formula | 11030 | 9945 | 612 | 473 | 23449 | 20933 | 1419 | 1097 | -0.32* | -0.35* | -0.15* | -0.38* | -0.31* | -0.35* | -0.13* | -0.36* |
| Percentage of land with biodiversity value within green space | Formula | 11030 | 9945 | 612 | 473 | 23449 | 20933 | 1419 | 1097 | -0.01 | -0.02 | 0.10+ | 0 | 0 | -0.01 | 0.08× | -0.01 |
| Percentage of land with biodiversity value outside of green space within 100m | Formula | 11030 | 9945 | 612 | 473 | 23449 | 20933 | 1419 | 1097 | -0.03* | -0.04* | 0.12× | 0 | -0.02* | -0.03* | 0.10* | -0.02 |
| **Incivilities indices – original sourced** | | | | | | | | | | | | | | | | | |
| Density of alcohol-related assaults | Max | 11030 | 9945 | 612 | 473 | 23449 | 20933 | 1419 | 1097 | 0.06* | 0.08* | 0.05 | 0.02 | 0.07* | 0.08* | 0.05+ | 0.08× |
| Density of malicious damage | Max | 11030 | 9945 | 612 | 473 | 23449 | 20933 | 1419 | 1097 | 0.25* | 0.26* | 0.17* | 0.30* | 0.25* | 0.26* | 0.23* | 0.37* |
| Density of non-domestic violence assaults | Max | 11030 | 9945 | 612 | 473 | 23449 | 20933 | 1419 | 1097 | 0.13* | 0.14* | 0.06 | 0.11+ | 0.13* | 0.14* | 0.06+ | 0.17* |
| Density of robberies | Max | 11030 | 9945 | 612 | 473 | 23449 | 20933 | 1419 | 1097 | 0.18* | 0.19* | 0.02 | 0.16* | 0.18* | 0.19* | 0.01 | 0.18* |
| Density of steal from persons | Max | 11030 | 9945 | 612 | 473 | 23449 | 20933 | 1419 | 1097 | 0.07* | 0.08* | 0.01 | 0.06 | 0.07* | 0.09* | 0.01 | 0.12* |
| **Incivilities indices – calculated** | | | | | | | | | | | | | | | | | |
| Mean level of incivilities | Formula | 11030 | 9945 | 612 | 473 | 23449 | 20933 | 1419 | 1097 | 0.24* | 0.25* | 0.16* | 0.32* | 0.25* | 0.25* | 0.20* | 0.39* |
| **Land cover indices – original sourced** | | | | | | | | | | | | | | | | | |
| Area of bare earth within green space | w0 Sum | 11011 | 9931 | 609 | 471 | 23402 | 20900 | 1411 | 1091 | 0.19* | 0.19* | -0.03 | 0.11+ | 0.18* | 0.19* | -0.01 | 0.10* |
| Area of buildings within green space | w0 Sum | 11011 | 9931 | 609 | 471 | 23402 | 20900 | 1411 | 1091 | 0.02 | 0.01 | 0.01 | 0.05 | 0.02× | 0.01 | 0.03 | 0.12* |
| Area of built up area within green space | w0 Sum | 11011 | 9931 | 609 | 471 | 23402 | 20900 | 1411 | 1091 | 0.01 | 0 | -0.04 | 0.01 | 0 | -0.01 | -0.01 | 0.08+ |
| Area of open grass within green space | w0 Sum | 11011 | 9931 | 609 | 471 | 23402 | 20900 | 1411 | 1091 | 0.17* | 0.16* | 0.05 | 0.18* | 0.18* | 0.17* | 0.08× | 0.21* |
| Area of other vegetation within green space | w0 Sum | 11011 | 9931 | 609 | 471 | 23402 | 20900 | 1411 | 1091 | 0.03× | 0.03+ | -0.13× | -0.03 | 0.03* | 0.02+ | -0.10* | -0.02 |
| Area of roads/paths within green space | w0 Sum | 11011 | 9931 | 609 | 471 | 23402 | 20900 | 1411 | 1091 | 0.06* | 0.03× | -0.02 | 0.14× | 0.06* | 0.03* | 0 | 0.13* |
| Area of shadow within green space | w0 Sum | 11011 | 9931 | 609 | 471 | 23402 | 20900 | 1411 | 1091 | -0.11* | -0.09* | -0.07 | -0.17* | -0.12* | -0.09* | -0.12* | -0.19* |
| Area of swimming pools within green space | w0 Sum | 11011 | 9931 | 609 | 471 | 23402 | 20900 | 1411 | 1091 | -0.09* | -0.08* | -0.17* | 0.06 | -0.09* | -0.08* | -0.19* | 0.06+ |
| Area of tree canopy within green space | w0 Sum | 11011 | 9931 | 609 | 471 | 23402 | 20900 | 1411 | 1091 | -0.22* | -0.22* | -0.15* | -0.12+ | -0.22* | -0.23* | -0.13* | -0.13* |
| Area of water within green space | w0 Sum | 11011 | 9931 | 609 | 471 | 23402 | 20900 | 1411 | 1091 | -0.09* | -0.12* | 0.05 | -0.07 | -0.09* | -0.13* | -0.01 | -0.05 |
| **Land cover indices - calculated** | | | | | | | | | | | | | | | | | |
| Percentage of bare earth within green space | Formula | 11011 | 9931 | 609 | 471 | 23402 | 20900 | 1411 | 1091 | 0.27* | 0.28* | 0.04 | 0.12+ | 0.27* | 0.29* | 0.03 | 0.09× |
| Percentage of buildings within green space | Formula | 11011 | 9931 | 609 | 471 | 23402 | 20900 | 1411 | 1091 | 0.03× | 0.03× | 0.05 | 0.08 | 0.04* | 0.03* | 0.08× | 0.15* |
| Percentage of built up area within green space | Formula | 11011 | 9931 | 609 | 471 | 23402 | 20900 | 1411 | 1091 | 0.02+ | 0.02+ | 0 | 0 | 0.02+ | 0.02× | 0.03 | 0.08× |
| Percentage of open grass within green space | Formula | 11011 | 9931 | 609 | 471 | 23402 | 20900 | 1411 | 1091 | 0.29* | 0.28* | 0.28* | 0.34* | 0.31* | 0.30* | 0.27* | 0.38* |
| Percentage of other vegetation within green space | Formula | 11011 | 9931 | 609 | 471 | 23402 | 20900 | 1411 | 1091 | 0.08* | 0.07* | -0.09+ | -0.17* | 0.07* | 0.07* | -0.13* | -0.16* |
| Percentage of roads/paths within green space | Formula | 11011 | 9931 | 609 | 471 | 23402 | 20900 | 1411 | 1091 | 0.08* | 0.04* | 0.04 | 0.22* | 0.09* | 0.05* | 0.06+ | 0.24* |
| Percentage of shadow within green space | Formula | 11011 | 9931 | 609 | 471 | 23402 | 20900 | 1411 | 1091 | -0.11* | -0.09* | -0.08 | -0.19* | -0.12* | -0.10* | -0.12* | -0.20* |
| Percentage of swimming pools within green space | Formula | 11011 | 9931 | 609 | 471 | 23402 | 20900 | 1411 | 1091 | -0.08* | -0.08* | -0.16* | 0.06 | -0.09* | -0.08* | -0.18* | 0.06+ |
| Percentage of tree canopy within green space | Formula | 11011 | 9931 | 609 | 471 | 23402 | 20900 | 1411 | 1091 | -0.36* | -0.36* | -0.16* | -0.24* | -0.38* | -0.38* | -0.18* | -0.30* |
| Percentage of water within green space | Formula | 11011 | 9931 | 609 | 471 | 23402 | 20900 | 1411 | 1091 | -0.08* | -0.12* | 0.06 | -0.08 | -0.09* | -0.13* | 0 | -0.05 |
| **Land use indices – original sourced** | | | | | | | | | | | | | | | | | |
| Area of commercial land use surrounding green space (within 100m doughnut buffer) | w0 Sum | 11030 | 9945 | 612 | 473 | 23449 | 20933 | 1419 | 1097 | 0.05* | 0.06* | 0.06 | 0.17* | 0.05* | 0.05* | 0.04 | 0.17* |
| Area of education land use surrounding green space (within 100m doughnut buffer) | w0 Sum | 11030 | 9945 | 612 | 473 | 23449 | 20933 | 1419 | 1097 | 0.06* | 0.05* | 0.13× | 0.09+ | 0.04* | 0.04* | 0.11* | 0.14* |
| Area of hospital/medical land use surrounding green space (within 100m doughnut buffer) | w0 Sum | 11030 | 9945 | 612 | 473 | 23449 | 20933 | 1419 | 1097 | 0.02+ | 0.03× | -0.08+ | 0.01 | 0 | 0 | -0.08× | 0 |
| Area of industrial land use surrounding green space (within 100m doughnut buffer) | w0 Sum | 11030 | 9945 | 612 | 473 | 23449 | 20933 | 1419 | 1097 | 0.09* | 0.08* | 0.10+ | 0.19* | 0.09* | 0.08* | 0.10* | 0.14* |
| Area of other land use surrounding green space (within 100m doughnut buffer) | w0 Sum | 11030 | 9945 | 612 | 473 | 23449 | 20933 | 1419 | 1097 | 0.01 | 0 | 0.12× | -0.07 | 0.01 | 0 | 0.10* | -0.11* |
| Area of parkland land use surrounding green space (within 100m doughnut buffer) | w0 Sum | 11030 | 9945 | 612 | 473 | 23449 | 20933 | 1419 | 1097 | -0.06* | -0.06* | -0.14* | -0.13× | -0.04* | -0.04* | -0.09× | -0.09× |
| Area of primary production land use surrounding green space (within 100m doughnut buffer) | w0 Sum | 11030 | 9945 | 612 | 473 | 23449 | 20933 | 1419 | 1097 | 0.05* | 0.05* | 0.12× | -0.06 | 0.05* | 0.04* | 0.13* | -0.03 |
| Area of residential land use surrounding green space (within 100m doughnut buffer) | w0 Sum | 11030 | 9945 | 612 | 473 | 23449 | 20933 | 1419 | 1097 | -0.06* | -0.04* | -0.10+ | 0.04 | -0.07* | -0.05* | -0.06+ | 0.05 |
| Area of transport land use surrounding green space (within 100m doughnut buffer) | w0 Sum | 11030 | 9945 | 612 | 473 | 23449 | 20933 | 1419 | 1097 | 0 | 0.01 | 0.03 | 0.02 | 0 | 0.02+ | 0 | 0.04 |
| Area of water land use surrounding green space (within 100m doughnut buffer) | w0 Sum | 11030 | 9945 | 612 | 473 | 23449 | 20933 | 1419 | 1097 | 0 | 0 | 0.09+ |  | 0 | 0 | 0.07× |  |
| Area of tree canopy surrounding green space (within 100m doughnut buffer) | w0 Sum | 10976 | 9911 | 601 | 464 | 23301 | 20841 | 1388 | 1072 | -0.16* | -0.14* | -0.11× | -0.02 | -0.18* | -0.16* | -0.09* | -0.02 |
| Area of tree canopy ≤4m surrounding green space (within 100m doughnut buffer) | w0 Sum | 10976 | 9911 | 601 | 464 | 23301 | 20841 | 1388 | 1072 | -0.10* | -0.08* | -0.09+ | -0.01 | -0.11* | -0.10* | -0.05 | -0.02 |
| Area of tree canopy >4m surrounding green space (within 100m doughnut buffer) | w0 Sum | 10976 | 9911 | 601 | 464 | 23301 | 20841 | 1388 | 1072 | -0.16* | -0.14* | -0.11× | -0.02 | -0.18* | -0.17* | -0.10* | -0.02 |
| **Land use indices – calculated** | | | | | | | | | | | | | | | | | |
| Percentage of commercial land use surrounding green space (within 100m doughnut buffer) | Formula | 11030 | 9945 | 612 | 473 | 23449 | 20933 | 1419 | 1097 | 0.06* | 0.06* | 0.07 | 0.17* | 0.06* | 0.06* | 0.05 | 0.17* |
| Percentage of education land use surrounding green space (within 100m doughnut buffer) | Formula | 11030 | 9945 | 612 | 473 | 23449 | 20933 | 1419 | 1097 | 0.06* | 0.06* | 0.13× | 0.09+ | 0.05* | 0.04* | 0.11* | 0.14* |
| Percentage of hospital/medical land use surrounding green space (within 100m doughnut buffer) | Formula | 11030 | 9945 | 612 | 473 | 23449 | 20933 | 1419 | 1097 | 0.02+ | 0.03× | -0.08+ | 0.01 | 0 | 0 | -0.08× | 0 |
| Percentage of industrial land use surrounding green space (within 100m doughnut buffer) | Formula | 11030 | 9945 | 612 | 473 | 23449 | 20933 | 1419 | 1097 | 0.09* | 0.08* | 0.10+ | 0.18* | 0.09* | 0.08* | 0.10* | 0.14* |
| Percentage of other land use surrounding green space (within 100m doughnut buffer) | Formula | 11030 | 9945 | 612 | 473 | 23449 | 20933 | 1419 | 1097 | 0.01 | 0 | 0.12× | -0.07 | 0.01 | 0 | 0.10* | -0.11* |
| Percentage of parkland land use surrounding green space (within 100m doughnut buffer) | Formula | 11030 | 9945 | 612 | 473 | 23449 | 20933 | 1419 | 1097 | -0.05* | -0.06* | -0.03 | -0.14× | -0.04* | -0.04* | -0.04 | -0.12* |
| Percentage of primary production land use surrounding green space (within 100m doughnut buffer) | Formula | 11030 | 9945 | 612 | 473 | 23449 | 20933 | 1419 | 1097 | 0.05* | 0.05* | 0.12× | -0.06 | 0.05* | 0.04* | 0.13* | -0.03 |
| Percentage of residential land use surrounding green space (within 100m doughnut buffer) | Formula | 11030 | 9945 | 612 | 473 | 23449 | 20933 | 1419 | 1097 | 0 | 0.01 | -0.10+ | 0 | -0.02+ | 0 | -0.08× | 0 |
| Percentage of transport land use surrounding green space (within 100m doughnut buffer) | Formula | 11030 | 9945 | 612 | 473 | 23449 | 20933 | 1419 | 1097 | 0 | 0.01 | 0.03 | 0.02 | 0 | 0.02+ | 0 | 0.04 |
| Percentage of water land use surrounding green space (within 100m doughnut buffer) | Formula | 11030 | 9945 | 612 | 473 | 23449 | 20933 | 1419 | 1097 | 0 | 0 | 0.09+ |  | 0 | 0 | 0.07× |  |
| Percentage of tree canopy to roads surrounding green space (within 100m doughnut buffer) | Formula | 10976 | 9911 | 601 | 464 | 23301 | 20841 | 1388 | 1072 | -0.40* | -0.41* | -0.05 | -0.22* | -0.43* | -0.43* | -0.08× | -0.25* |
| Percentage of tree canopy ≤4m to roads surrounding green space (within 100m doughnut buffer) | Formula | 10976 | 9911 | 601 | 464 | 23301 | 20841 | 1388 | 1072 | -0.18* | -0.17* | 0.04 | -0.23* | -0.19* | -0.18* | 0.04 | -0.30* |
| Percentage of tree canopy >4m to roads surrounding green space (within 100m doughnut buffer) | Formula | 10976 | 9911 | 601 | 464 | 23301 | 20841 | 1388 | 1072 | -0.40* | -0.40* | -0.05 | -0.19* | -0.42* | -0.42* | -0.09* | -0.21* |
| Percentage of tree canopy surrounding green space (within 100m doughnut buffer) | Formula | 10976 | 9911 | 601 | 464 | 23301 | 20841 | 1388 | 1072 | -0.31* | -0.30* | -0.05 | -0.10+ | -0.34* | -0.33* | -0.07× | -0.14* |
| Percentage of tree canopy ≤4m surrounding green space (within 100m doughnut buffer) | Formula | 10976 | 9911 | 601 | 464 | 23301 | 20841 | 1388 | 1072 | -0.12* | -0.10* | 0.03 | -0.10+ | -0.14* | -0.12* | 0.04 | -0.15* |
| Percentage of tree canopy >4m surrounding green space (within 100m doughnut buffer) | Formula | 10976 | 9911 | 601 | 464 | 23301 | 20841 | 1388 | 1072 | -0.31* | -0.30* | -0.04 | -0.09 | -0.34* | -0.33* | -0.08× | -0.12* |

Method of aggregating intersects over mesh blocks: Count=unweighted count | Sum=unweighted sum | w0 Sum = sum weighted by the ratio of the green space intersect with the buffer area to the green space area | w1 Mean = mean weighted by the ratio of the intersect area to the sum of all intersect areas | Max = maximum value over the intersects within the buffer | Quality = Value 0/1, aggregated to 1 if any intersects equal 1 (technically Quality = Max) | Formula = calculation of the index from other qualities that have already been aggregated as weighted sums or weighted means.

P-value signs: * ≤0.001, × ≤0.01, + ≤0.05

GS = green space

Table S4. Spearman's rank coefficient of correlation between SEIFA IRSD score (reversed) and aggregated over MB green space features – n, Rho and compressed P-values – 300m and 400m buffers.

|  |  | **Buffer 300m (n)** | | | | **Buffer 400m (n)** | | | | **Buffer 300m (Rho and P-value)** | | | | **Buffer 400m (Rho and P-value)** | | | |
| --- | --- | --- | --- | --- | --- | --- | --- | --- | --- | --- | --- | --- | --- | --- | --- | --- | --- |
| **Index description** | **Method** | **Full set** | **Syd** | **New** | **Wol** | **Full set** | **Syd** | **New** | **Wol** | **Full set** | **Syd** | **New** | **Wol** | **Full set** | **Syd** | **New** | **Wol** |
| **General indices** | | | | | | | | | | | | | | | | | |
| N green spaces in MB buffer | Count | 33290 | 29481 | 2153 | 1656 | 39570 | 34846 | 2664 | 2060 | -0.11* | -0.11* | -0.02 | 0.06+ | -0.11* | -0.11* | -0.02 | 0.07* |
| Intersect of buffer with green space | Sum | 33290 | 29481 | 2153 | 1656 | 39570 | 34846 | 2664 | 2060 | -0.02* | -0.04* | -0.04 | 0.11* | -0.02* | -0.04* | -0.03 | 0.14* |
| Green space area (A) | Sum | 33290 | 29481 | 2153 | 1656 | 39570 | 34846 | 2664 | 2060 | -0.02* | -0.05* | -0.05+ | 0.07× | -0.04* | -0.07* | -0.03 | 0.08* |
| Green space perimeter (P) | w0 Sum | 33290 | 29481 | 2153 | 1656 | 39570 | 34846 | 2664 | 2060 | -0.08* | -0.08* | -0.03 | 0.11* | -0.07* | -0.07* | -0.02 | 0.14* |
| **Access indices – original sourced** | | | | | | | | | | | | | | | | | |
| Length of pathways within green space | w0 Sum | 33290 | 29481 | 2153 | 1656 | 39570 | 34846 | 2664 | 2060 | -0.02* | -0.02× | -0.14* | 0.14* | -0.02* | -0.02* | -0.12* | 0.13* |
| Length of bikeways within green space | w0 Sum | 33290 | 29481 | 2153 | 1656 | 39570 | 34846 | 2664 | 2060 | 0.01+ | -0.01 | -0.05+ | 0.12* | 0.01 | -0.01+ | -0.03 | 0.13* |
| Length of pathways covered by trees within green space | w0 Sum | 33018 | 29306 | 2102 | 1610 | 39187 | 34604 | 2593 | 1990 | -0.05* | -0.05* | -0.19* | 0.11* | -0.06* | -0.06* | -0.17* | 0.10* |
| Length of bikeways covered by trees within green space | w0 Sum | 33018 | 29306 | 2102 | 1610 | 39187 | 34604 | 2593 | 1990 | -0.02* | -0.03* | -0.06× | 0.10* | -0.03* | -0.04* | -0.04+ | 0.10* |
| Length of pathways greater than 6 degrees gradient within green space | w0 Sum | 33207 | 29404 | 2151 | 1652 | 39462 | 34745 | 2662 | 2055 | -0.16* | -0.16* | -0.21* | 0.02 | -0.17* | -0.18* | -0.19* | 0 |
| Length of bikeways greater than 6 degrees gradient within green space | w0 Sum | 33207 | 29404 | 2151 | 1652 | 39462 | 34745 | 2662 | 2055 | -0.06* | -0.08* | -0.11* | -0.02 | -0.07* | -0.09* | -0.09* | -0.03 |
| Number of Railway Stations within and surrounding a green space (within 100m buffer) | w0 Sum | 33290 | 29481 | 2153 | 1656 | 39570 | 34846 | 2664 | 2060 | 0 | 0.01 | -0.02 | -0.04 | 0.01 | 0.02* | -0.01 | -0.05+ |
| Number of Bus Stops within and surrounding a green space (within 100m buffer) | w0 Sum | 33290 | 29481 | 2153 | 1656 | 39570 | 34846 | 2664 | 2060 | -0.02* | -0.02* | -0.04 | 0.17* | -0.02* | -0.02× | -0.04+ | 0.20* |
| Number of ferry wharves within and surrounding a green space (within 100m buffer) | w0 Sum | 33290 | 29481 | 2153 | 1656 | 39570 | 34846 | 2664 | 2060 | -0.10* | -0.10* | 0.05+ |  | -0.11* | -0.11* | 0.05× |  |
| Area of GS which is greater than 6 degrees of Slope | w0 Sum | 33207 | 29404 | 2151 | 1652 | 39462 | 34745 | 2662 | 2055 | -0.29* | -0.32* | -0.22* | -0.17* | -0.30* | -0.33* | -0.21* | -0.17* |
| Length of Main Roads within 100 metres of the Green Space | w0 Sum | 33290 | 29481 | 2153 | 1656 | 39570 | 34846 | 2664 | 2060 | -0.04* | -0.04* | 0.08* | 0.14* | -0.04* | -0.04* | 0.08* | 0.18* |
| **Access indices – calculated** | | | | | | | | | | | | | | | | | |
| Ratio of Area(A) to Perimeter (P) | w1 Mean | 33290 | 29481 | 2153 | 1656 | 39570 | 34846 | 2664 | 2060 | 0.06* | 0.03* | 0 | 0.05 | 0.05* | 0.02× | 0 | 0.06× |
| Ratio of Perimeter (P) to perimeter standardised by area (A) | w1 Mean | 33290 | 29481 | 2153 | 1656 | 39570 | 34846 | 2664 | 2060 | -0.15* | -0.14* | -0.16* | -0.06+ | -0.16* | -0.15* | -0.15* | -0.09* |
| Total number of transport stops/stations/wharves within and surrounding a green space (within 100m buffer) | Formula | 33290 | 29481 | 2153 | 1656 | 39570 | 34846 | 2664 | 2060 | -0.03* | -0.03* | -0.05+ | 0.17* | -0.02* | -0.02* | -0.05+ | 0.20* |
| Pathway Length per Area of Green Space | Formula | 33290 | 29481 | 2153 | 1656 | 39570 | 34846 | 2664 | 2060 | -0.01× | -0.01+ | -0.15* | 0.13* | -0.02× | -0.01 | -0.12* | 0.11* |
| Bikeway Length per Area of Green Space | Formula | 33290 | 29481 | 2153 | 1656 | 39570 | 34846 | 2664 | 2060 | 0.02* | 0 | -0.04 | 0.10* | 0.01× | 0 | -0.02 | 0.09* |
| Trees over Pathway per Area of Green Space | Formula | 33018 | 29306 | 2102 | 1610 | 39187 | 34604 | 2593 | 1990 | -0.05* | -0.05* | -0.19* | 0.10* | -0.06* | -0.06* | -0.17* | 0.07* |
| Trees over Bikeway per Area of Green Space | Formula | 33018 | 29306 | 2102 | 1610 | 39187 | 34604 | 2593 | 1990 | -0.02* | -0.02* | -0.06× | 0.07× | -0.03* | -0.04* | -0.04 | 0.06× |
| Percentage of GS which is greater than 6 degrees of Slope | Formula | 33207 | 29404 | 2151 | 1652 | 39462 | 34745 | 2662 | 2055 | -0.41* | -0.43* | -0.32* | -0.40* | -0.42* | -0.44* | -0.32* | -0.42* |
| Pathway Length > 6 degrees per Area of GS | Formula | 33207 | 29404 | 2151 | 1652 | 39462 | 34745 | 2662 | 2055 | -0.16* | -0.17* | -0.22* | 0.01 | -0.17* | -0.18* | -0.19* | -0.02 |
| Bikeway Length > 6 degrees slope per Area of GS | Formula | 33207 | 29404 | 2151 | 1652 | 39462 | 34745 | 2662 | 2055 | -0.06* | -0.08* | -0.11* | -0.04 | -0.07* | -0.08* | -0.08* | -0.06× |
| **Amenities/Activities indices – original sourced** | | | | | | | | | | | | | | | | | |
| Number of public toilet facilities within and surrounding a green space (within 100m buffer) | w0 Sum | 33290 | 29481 | 2153 | 1656 | 39570 | 34846 | 2664 | 2060 | -0.05* | -0.06* | -0.03 | 0.13* | -0.06* | -0.07* | -0.01 | 0.15* |
| Number of cafes within and surrounding a green space (within 100m buffer) | w0 Sum | 33290 | 29481 | 2153 | 1656 | 39570 | 34846 | 2664 | 2060 | -0.12* | -0.11* | -0.16* | 0.05+ | -0.13* | -0.12* | -0.17* | 0.06+ |
| Number of hotels/bars within and surrounding a green space (within 100m buffer) | w0 Sum | 33290 | 29481 | 2153 | 1656 | 39570 | 34846 | 2664 | 2060 | -0.05* | -0.05* | -0.06× | 0.02 | -0.06* | -0.05* | -0.05× | 0.02 |
| Number of restaurants/takeaways within and surrounding a green space (within 100m buffer) | w0 Sum | 33290 | 29481 | 2153 | 1656 | 39570 | 34846 | 2664 | 2060 | -0.06* | -0.05* | -0.01 | -0.02 | -0.06* | -0.05* | 0.01 | 0 |
| Number of restaurants within and surrounding a green space (within 100m buffer) | w0 Sum | 33290 | 29481 | 2153 | 1656 | 39570 | 34846 | 2664 | 2060 | -0.11* | -0.10* | -0.09* | -0.06× | -0.11* | -0.10* | -0.09* | -0.06+ |
| Number of takeaways within and surrounding a green space (within 100m buffer) | w0 Sum | 33290 | 29481 | 2153 | 1656 | 39570 | 34846 | 2664 | 2060 | 0 | 0.01+ | 0.03 | 0.05+ | 0 | 0.01 | 0.04+ | 0.06× |
| Number of supermarkets/greengrocers within and surrounding a green space (within 100m buffer) | w0 Sum | 33290 | 29481 | 2153 | 1656 | 39570 | 34846 | 2664 | 2060 | 0.01 | 0.01+ | 0.02 | 0.12* | 0.01× | 0.02* | 0.02 | 0.12* |
| Presence of parkrun event | Quality | 33290 | 29481 | 2153 | 1656 | 39570 | 34846 | 2664 | 2060 | -0.03* | -0.02* | -0.07* | -0.10* | -0.04* | -0.03* | -0.09* | -0.13* |
| **Beach/Coast indices – original sourced** | | | | | | | | | | | | | | | | | |
| Presence of coastline | Quality | 33290 | 29481 | 2153 | 1656 | 39570 | 34846 | 2664 | 2060 | -0.25* | -0.29* | -0.06× | -0.02 | -0.26* | -0.31* | -0.06× | -0.02 |
| Area of beach within and surrounding a green space (within 100m buffer) | w0 Sum | 33290 | 29481 | 2153 | 1656 | 39570 | 34846 | 2664 | 2060 | -0.16* | -0.18* | -0.11* | -0.17* | -0.17* | -0.19* | -0.11* | -0.17* |
| **Biodiversity indices – original sourced** | | | | | | | | | | | | | | | | | |
| Number of bird species/species habitat likely to occur within green space | Max | 33290 | 29481 | 2153 | 1656 | 39570 | 34846 | 2664 | 2060 | -0.29* | -0.34* | 0 | 0.17* | -0.29* | -0.34* | 0 | 0.20* |
| Number of fish species/species habitat likely to occur within green space | Max | 33290 | 29481 | 2153 | 1656 | 39570 | 34846 | 2664 | 2060 | -0.31* | -0.35* | -0.11* | 0.04 | -0.32* | -0.36* | -0.11* | 0.05+ |
| Number of flora species/species habitat likely to occur within green space | Max | 33290 | 29481 | 2153 | 1656 | 39570 | 34846 | 2664 | 2060 | -0.27* | -0.35* | 0.06× | 0.32* | -0.26* | -0.35* | 0.06× | 0.32* |
| Number of frog species/species habitat likely to occur within green space | Max | 33290 | 29481 | 2153 | 1656 | 39570 | 34846 | 2664 | 2060 | -0.24* | -0.28* | 0.09* | -0.18* | -0.22* | -0.26* | 0.09* | -0.19* |
| Number of mammal species/species habitat likely to occur within green space | Max | 33290 | 29481 | 2153 | 1656 | 39570 | 34846 | 2664 | 2060 | -0.51* | -0.56* | -0.20* | -0.30* | -0.51* | -0.56* | -0.21* | -0.29* |
| Number of other animal species/species habitat likely to occur within green space | Max | 33290 | 29481 | 2153 | 1656 | 39570 | 34846 | 2664 | 2060 | -0.29* | -0.29* |  |  | -0.31* | -0.31* |  |  |
| Number of reptile species/species habitat likely to occur within green space | Max | 33290 | 29481 | 2153 | 1656 | 39570 | 34846 | 2664 | 2060 | -0.34* | -0.39* | -0.12* | -0.05+ | -0.35* | -0.39* | -0.12* | -0.03 |
| Area of land with biodiversity value within green space | w0 Sum | 33290 | 29481 | 2153 | 1656 | 39570 | 34846 | 2664 | 2060 | -0.01+ | -0.02× | 0.06× | 0.01 | -0.01× | -0.02* | 0.07* | 0.01 |
| Area of land with biodiversity value outside of green space within 100m | w0 Sum | 33290 | 29481 | 2153 | 1656 | 39570 | 34846 | 2664 | 2060 | -0.04* | -0.05* | 0.08* | 0 | -0.05* | -0.05* | 0.08* | 0 |
| **Biodiversity indices – calculated** | | | | | | | | | | | | | | | | | |
| Total number of species/species habitat likely to occur within green space | Formula | 33290 | 29481 | 2153 | 1656 | 39570 | 34846 | 2664 | 2060 | -0.48* | -0.54* | -0.06× | 0.17* | -0.48* | -0.54* | -0.07* | 0.19* |
| Shannon biodiversity index | Formula | 33290 | 29481 | 2153 | 1656 | 39570 | 34846 | 2664 | 2060 | -0.41* | -0.46* | -0.18* | -0.28* | -0.41* | -0.46* | -0.18* | -0.29* |
| Simpson biodiversity index (range 0 to 1=most diverse) | Formula | 33290 | 29481 | 2153 | 1656 | 39570 | 34846 | 2664 | 2060 | -0.30* | -0.34* | -0.10* | -0.38* | -0.30* | -0.33* | -0.11* | -0.40* |
| Percentage of land with biodiversity value within green space | Formula | 33290 | 29481 | 2153 | 1656 | 39570 | 34846 | 2664 | 2060 | -0.01× | -0.02* | 0.08* | 0.01 | -0.02* | -0.02* | 0.08* | 0 |
| Percentage of land with biodiversity value outside of green space within 100m | Formula | 33290 | 29481 | 2153 | 1656 | 39570 | 34846 | 2664 | 2060 | -0.04* | -0.05* | 0.10* | -0.01 | -0.05* | -0.06* | 0.10* | -0.01 |
| **Incivilities indices – original sourced** | | | | | | | | | | | | | | | | | |
| Density of alcohol-related assaults | Max | 33290 | 29481 | 2153 | 1656 | 39570 | 34846 | 2664 | 2060 | 0.06* | 0.07* | 0.07× | 0.14* | 0.06* | 0.07* | 0.08* | 0.16* |
| Density of malicious damage | Max | 33290 | 29481 | 2153 | 1656 | 39570 | 34846 | 2664 | 2060 | 0.26* | 0.26* | 0.23* | 0.35* | 0.26* | 0.27* | 0.25* | 0.34* |
| Density of non-domestic violence assaults | Max | 33290 | 29481 | 2153 | 1656 | 39570 | 34846 | 2664 | 2060 | 0.13* | 0.14* | 0.09* | 0.20* | 0.14* | 0.15* | 0.10* | 0.24* |
| Density of robberies | Max | 33290 | 29481 | 2153 | 1656 | 39570 | 34846 | 2664 | 2060 | 0.18* | 0.20* | -0.01 | 0.19* | 0.19* | 0.21* | -0.03 | 0.20* |
| Density of steal from persons | Max | 33290 | 29481 | 2153 | 1656 | 39570 | 34846 | 2664 | 2060 | 0.07* | 0.08* | -0.01 | 0.12* | 0.08* | 0.09* | -0.01 | 0.14* |
| **Incivilities indices – calculated** | | | | | | | | | | | | | | | | | |
| Mean level of incivilities | Formula | 33290 | 29481 | 2153 | 1656 | 39570 | 34846 | 2664 | 2060 | 0.25* | 0.26* | 0.20* | 0.38* | 0.26* | 0.27* | 0.21* | 0.37* |
| **Land cover indices – original sourced** | | | | | | | | | | | | | | | | | |
| Area of bare earth within green space | w0 Sum | 33198 | 29408 | 2142 | 1648 | 39408 | 34710 | 2648 | 2050 | 0.17* | 0.17* | 0.01 | 0.16* | 0.17* | 0.18* | 0.02 | 0.17* |
| Area of buildings within green space | w0 Sum | 33198 | 29408 | 2142 | 1648 | 39408 | 34710 | 2648 | 2050 | 0.01 | 0 | 0.04 | 0.19* | 0.01 | 0 | 0.03 | 0.20* |
| Area of built up area within green space | w0 Sum | 33198 | 29408 | 2142 | 1648 | 39408 | 34710 | 2648 | 2050 | 0 | -0.01+ | -0.01 | 0.14* | 0 | -0.01 | 0 | 0.16* |
| Area of open grass within green space | w0 Sum | 33198 | 29408 | 2142 | 1648 | 39408 | 34710 | 2648 | 2050 | 0.16* | 0.15* | 0.10* | 0.27* | 0.16* | 0.14* | 0.11* | 0.27* |
| Area of other vegetation within green space | w0 Sum | 33198 | 29408 | 2142 | 1648 | 39408 | 34710 | 2648 | 2050 | 0.02× | 0 | -0.07* | 0.06× | 0.01+ | 0 | -0.07* | 0.09* |
| Area of roads/paths within green space | w0 Sum | 33198 | 29408 | 2142 | 1648 | 39408 | 34710 | 2648 | 2050 | 0.05* | 0.02× | 0.02 | 0.19* | 0.06* | 0.02* | 0.03 | 0.22* |
| Area of shadow within green space | w0 Sum | 33198 | 29408 | 2142 | 1648 | 39408 | 34710 | 2648 | 2050 | -0.14* | -0.12* | -0.12* | -0.18* | -0.15* | -0.13* | -0.14* | -0.18* |
| Area of swimming pools within green space | w0 Sum | 33198 | 29408 | 2142 | 1648 | 39408 | 34710 | 2648 | 2050 | -0.11* | -0.10* | -0.17* | 0.08× | -0.12* | -0.11* | -0.17* | 0.08* |
| Area of tree canopy within green space | w0 Sum | 33198 | 29408 | 2142 | 1648 | 39408 | 34710 | 2648 | 2050 | -0.23* | -0.24* | -0.11* | -0.06× | -0.23* | -0.25* | -0.11* | -0.05+ |
| Area of water within green space | w0 Sum | 33198 | 29408 | 2142 | 1648 | 39408 | 34710 | 2648 | 2050 | -0.11* | -0.14* | -0.03 | 0.01 | -0.11* | -0.15* | -0.01 | 0.03 |
| **Land cover indices - calculated** | | | | | | | | | | | | | | | | | |
| Percentage of bare earth within green space | Formula | 33198 | 29408 | 2142 | 1648 | 39408 | 34710 | 2648 | 2050 | 0.27* | 0.29* | 0.05+ | 0.13* | 0.28* | 0.30* | 0.07* | 0.12* |
| Percentage of buildings within green space | Formula | 33198 | 29408 | 2142 | 1648 | 39408 | 34710 | 2648 | 2050 | 0.02* | 0.02* | 0.07× | 0.16* | 0.03* | 0.03* | 0.07* | 0.17* |
| Percentage of built up area within green space | Formula | 33198 | 29408 | 2142 | 1648 | 39408 | 34710 | 2648 | 2050 | 0.02* | 0.02* | 0.01 | 0.11* | 0.02* | 0.03* | 0.03 | 0.11* |
| Percentage of open grass within green space | Formula | 33198 | 29408 | 2142 | 1648 | 39408 | 34710 | 2648 | 2050 | 0.30* | 0.29* | 0.27* | 0.34* | 0.30* | 0.29* | 0.27* | 0.34* |
| Percentage of other vegetation within green space | Formula | 33198 | 29408 | 2142 | 1648 | 39408 | 34710 | 2648 | 2050 | 0.06* | 0.06* | -0.11* | -0.16* | 0.06* | 0.07* | -0.11* | -0.16* |
| Percentage of roads/paths within green space | Formula | 33198 | 29408 | 2142 | 1648 | 39408 | 34710 | 2648 | 2050 | 0.09* | 0.05* | 0.07× | 0.23* | 0.10* | 0.06* | 0.07* | 0.26* |
| Percentage of shadow within green space | Formula | 33198 | 29408 | 2142 | 1648 | 39408 | 34710 | 2648 | 2050 | -0.15* | -0.12* | -0.12* | -0.20* | -0.16* | -0.13* | -0.15* | -0.21* |
| Percentage of swimming pools within green space | Formula | 33198 | 29408 | 2142 | 1648 | 39408 | 34710 | 2648 | 2050 | -0.10* | -0.10* | -0.16* | 0.07× | -0.12* | -0.11* | -0.16* | 0.07* |
| Percentage of tree canopy within green space | Formula | 33198 | 29408 | 2142 | 1648 | 39408 | 34710 | 2648 | 2050 | -0.39* | -0.40* | -0.19* | -0.31* | -0.40* | -0.41* | -0.21* | -0.32* |
| Percentage of water within green space | Formula | 33198 | 29408 | 2142 | 1648 | 39408 | 34710 | 2648 | 2050 | -0.11* | -0.14* | -0.02 | 0 | -0.11* | -0.15* | -0.01 | 0.02 |
| **Land use indices – original sourced** | | | | | | | | | | | | | | | | | |
| Area of commercial land use surrounding green space (within 100m doughnut buffer) | w0 Sum | 33290 | 29481 | 2153 | 1656 | 39570 | 34846 | 2664 | 2060 | 0.04* | 0.05* | 0.03 | 0.19* | 0.05* | 0.06* | 0.05× | 0.20* |
| Area of education land use surrounding green space (within 100m doughnut buffer) | w0 Sum | 33290 | 29481 | 2153 | 1656 | 39570 | 34846 | 2664 | 2060 | 0.03* | 0.02* | 0.08* | 0.14* | 0.03* | 0.02* | 0.06× | 0.15* |
| Area of hospital/medical land use surrounding green space (within 100m doughnut buffer) | w0 Sum | 33290 | 29481 | 2153 | 1656 | 39570 | 34846 | 2664 | 2060 | -0.01 | 0 | -0.09* | 0 | -0.02* | -0.01+ | -0.09* | 0.03 |
| Area of industrial land use surrounding green space (within 100m doughnut buffer) | w0 Sum | 33290 | 29481 | 2153 | 1656 | 39570 | 34846 | 2664 | 2060 | 0.08* | 0.07* | 0.11* | 0.14* | 0.09* | 0.09* | 0.12* | 0.14* |
| Area of other land use surrounding green space (within 100m doughnut buffer) | w0 Sum | 33290 | 29481 | 2153 | 1656 | 39570 | 34846 | 2664 | 2060 | 0.01 | 0 | 0.09* | -0.07× | 0.01 | 0 | 0.09* | -0.07* |
| Area of parkland land use surrounding green space (within 100m doughnut buffer) | w0 Sum | 33290 | 29481 | 2153 | 1656 | 39570 | 34846 | 2664 | 2060 | -0.06* | -0.06* | -0.04 | -0.02 | -0.07* | -0.07* | -0.03 | 0 |
| Area of primary production land use surrounding green space (within 100m doughnut buffer) | w0 Sum | 33290 | 29481 | 2153 | 1656 | 39570 | 34846 | 2664 | 2060 | 0.05* | 0.04* | 0.13* | -0.05+ | 0.05* | 0.04* | 0.14* | -0.04 |
| Area of residential land use surrounding green space (within 100m doughnut buffer) | w0 Sum | 33290 | 29481 | 2153 | 1656 | 39570 | 34846 | 2664 | 2060 | -0.08* | -0.07* | -0.03 | 0.11* | -0.07* | -0.06* | -0.02 | 0.13* |
| Area of transport land use surrounding green space (within 100m doughnut buffer) | w0 Sum | 33290 | 29481 | 2153 | 1656 | 39570 | 34846 | 2664 | 2060 | 0.01+ | 0.02* | 0 | 0.05 | 0.02× | 0.03* | 0 | 0.05+ |
| Area of water land use surrounding green space (within 100m doughnut buffer) | w0 Sum | 33290 | 29481 | 2153 | 1656 | 39570 | 34846 | 2664 | 2060 | -0.01 | -0.01 | 0.09* |  | -0.01× | -0.01× | 0.09* |  |
| Area of tree canopy surrounding green space (within 100m doughnut buffer) | w0 Sum | 33018 | 29306 | 2102 | 1610 | 39187 | 34604 | 2593 | 1990 | -0.19* | -0.18* | -0.07× | 0.04 | -0.19* | -0.19* | -0.07* | 0.05+ |
| Area of tree canopy ≤4m surrounding green space (within 100m doughnut buffer) | w0 Sum | 33018 | 29306 | 2102 | 1610 | 39187 | 34604 | 2593 | 1990 | -0.13* | -0.11* | -0.03 | 0.04 | -0.12* | -0.11* | -0.02 | 0.06× |
| Area of tree canopy >4m surrounding green space (within 100m doughnut buffer) | w0 Sum | 33018 | 29306 | 2102 | 1610 | 39187 | 34604 | 2593 | 1990 | -0.20* | -0.19* | -0.08* | 0.04 | -0.20* | -0.19* | -0.07* | 0.05+ |
| **Land use indices – calculated** | | | | | | | | | | | | | | | | | |
| Percentage of commercial land use surrounding green space (within 100m doughnut buffer) | Formula | 33290 | 29481 | 2153 | 1656 | 39570 | 34846 | 2664 | 2060 | 0.06* | 0.06* | 0.04 | 0.19* | 0.07* | 0.07* | 0.06× | 0.20* |
| Percentage of education land use surrounding green space (within 100m doughnut buffer) | Formula | 33290 | 29481 | 2153 | 1656 | 39570 | 34846 | 2664 | 2060 | 0.04* | 0.04* | 0.08* | 0.13* | 0.04* | 0.04* | 0.07* | 0.14* |
| Percentage of hospital/medical land use surrounding green space (within 100m doughnut buffer) | Formula | 33290 | 29481 | 2153 | 1656 | 39570 | 34846 | 2664 | 2060 | -0.01 | 0 | -0.09* | 0 | -0.02× | -0.01+ | -0.09* | 0.03 |
| Percentage of industrial land use surrounding green space (within 100m doughnut buffer) | Formula | 33290 | 29481 | 2153 | 1656 | 39570 | 34846 | 2664 | 2060 | 0.09* | 0.08* | 0.11* | 0.13* | 0.10* | 0.09* | 0.12* | 0.14* |
| Percentage of other land use surrounding green space (within 100m doughnut buffer) | Formula | 33290 | 29481 | 2153 | 1656 | 39570 | 34846 | 2664 | 2060 | 0.01 | 0 | 0.09* | -0.07× | 0.01 | 0 | 0.09* | -0.08* |
| Percentage of parkland land use surrounding green space (within 100m doughnut buffer) | Formula | 33290 | 29481 | 2153 | 1656 | 39570 | 34846 | 2664 | 2060 | -0.05* | -0.05* | -0.02 | -0.08× | -0.05* | -0.06* | 0 | -0.09* |
| Percentage of primary production land use surrounding green space (within 100m doughnut buffer) | Formula | 33290 | 29481 | 2153 | 1656 | 39570 | 34846 | 2664 | 2060 | 0.05* | 0.04* | 0.13* | -0.05+ | 0.05* | 0.04* | 0.14* | -0.04 |
| Percentage of residential land use surrounding green space (within 100m doughnut buffer) | Formula | 33290 | 29481 | 2153 | 1656 | 39570 | 34846 | 2664 | 2060 | -0.02* | -0.01 | -0.06× | -0.03 | -0.02* | -0.01 | -0.09* | -0.03 |
| Percentage of transport land use surrounding green space (within 100m doughnut buffer) | Formula | 33290 | 29481 | 2153 | 1656 | 39570 | 34846 | 2664 | 2060 | 0.01+ | 0.03* | 0 | 0.05 | 0.02* | 0.03* | 0 | 0.05+ |
| Percentage of water land use surrounding green space (within 100m doughnut buffer) | Formula | 33290 | 29481 | 2153 | 1656 | 39570 | 34846 | 2664 | 2060 | -0.01 | -0.01 | 0.09* |  | -0.01× | -0.01× | 0.09* |  |
| Percentage of tree canopy to roads surrounding green space (within 100m doughnut buffer) | Formula | 33018 | 29306 | 2102 | 1610 | 39187 | 34604 | 2593 | 1990 | -0.43* | -0.44* | -0.11* | -0.28* | -0.44* | -0.45* | -0.12* | -0.30* |
| Percentage of tree canopy ≤4m to roads surrounding green space (within 100m doughnut buffer) | Formula | 33018 | 29306 | 2102 | 1610 | 39187 | 34604 | 2593 | 1990 | -0.19* | -0.18* | 0.01 | -0.33* | -0.20* | -0.19* | -0.01 | -0.34* |
| Percentage of tree canopy >4m to roads surrounding green space (within 100m doughnut buffer) | Formula | 33018 | 29306 | 2102 | 1610 | 39187 | 34604 | 2593 | 1990 | -0.42* | -0.43* | -0.12* | -0.24* | -0.43* | -0.44* | -0.13* | -0.26* |
| Percentage of tree canopy surrounding green space (within 100m doughnut buffer) | Formula | 33018 | 29306 | 2102 | 1610 | 39187 | 34604 | 2593 | 1990 | -0.35* | -0.34* | -0.10* | -0.17* | -0.35* | -0.35* | -0.12* | -0.19* |
| Percentage of tree canopy ≤4m surrounding green space (within 100m doughnut buffer) | Formula | 33018 | 29306 | 2102 | 1610 | 39187 | 34604 | 2593 | 1990 | -0.14* | -0.12* | 0.01 | -0.19* | -0.14* | -0.12* | -0.01 | -0.19* |
| Percentage of tree canopy >4m surrounding green space (within 100m doughnut buffer) | Formula | 33018 | 29306 | 2102 | 1610 | 39187 | 34604 | 2593 | 1990 | -0.35* | -0.35* | -0.11* | -0.15* | -0.36* | -0.36* | -0.13* | -0.16* |

Method of aggregating intersects over mesh blocks: Count=unweighted count | Sum=unweighted sum | w0 Sum = sum weighted by the ratio of the green space intersect with the buffer area to the green space area | w1 Mean = mean weighted by the ratio of the intersect area to the sum of all intersect areas | Max = maximum value over the intersects within the buffer | Quality = Value 0/1, aggregated to 1 if any intersects equal 1 (technically Quality = Max) | Formula = calculation of the index from other qualities that have already been aggregated as weighted sums or weighted means.

P-value signs: * ≤0.001, × ≤0.01, + ≤0.05

GS = green space

Table S5. Spearman's rank coefficient of correlation between SEIFA IRSD score (reversed) and aggregated over MB green space features – n, Rho and compressed P-values – 800m and 1600m buffers.

|  |  | **Buffer 800m (n)** | | | | **Buffer 1600m (n)** | | | | **Buffer 800m (Rho and P-value)** | | | | **Buffer 1600m (Rho and P-value)** | | | |
| --- | --- | --- | --- | --- | --- | --- | --- | --- | --- | --- | --- | --- | --- | --- | --- | --- | --- |
| **Index description** | **Method** | **Full set** | **Syd** | **New** | **Wol** | **Full set** | **Syd** | **New** | **Wol** | **Full set** | **Syd** | **New** | **Wol** | **Full set** | **Syd** | **New** | **Wol** |
| **General indices** | | | | | | | | | | | | | | | | | |
| N green spaces in MB | Count | 46245 | 40198 | 3345 | 2702 | 47079 | 40852 | 3441 | 2786 | -0.09* | -0.08* | 0.03 | 0.18* | -0.03* | -0.01+ | 0.07* | 0.26* |
| Intersect of buffer with green space | Sum | 46245 | 40198 | 3345 | 2702 | 47079 | 40852 | 3441 | 2786 | 0.01 | -0.01 | -0.03 | 0.23* | 0.09* | 0.08* | -0.06* | 0.34* |
| Green space area (A) | Sum | 46245 | 40198 | 3345 | 2702 | 47079 | 40852 | 3441 | 2786 | -0.05* | -0.08* | -0.02 | 0.12* | -0.04* | -0.07* | -0.02 | 0.02 |
| GS perimeter (P) | w0 Sum | 46245 | 40198 | 3345 | 2702 | 47079 | 40852 | 3441 | 2786 | -0.04* | -0.04* | -0.01 | 0.23* | 0.02* | 0.04* | -0.04+ | 0.32* |
| **Access indices – original sourced** | | | | | | | | | | | | | | | | | |
| Length of pathways within green space | w0 Sum | 46245 | 40198 | 3345 | 2702 | 47079 | 40852 | 3441 | 2786 | -0.01+ | 0 | -0.11* | 0.20* | 0 | 0 | -0.13* | 0.28* |
| Length of bikeways within green space | w0 Sum | 46245 | 40198 | 3345 | 2702 | 47079 | 40852 | 3441 | 2786 | 0.04* | 0.02* | 0.02 | 0.20* | 0.10* | 0.09* | 0.04+ | 0.29* |
| Length of pathways covered by trees within green space | w0 Sum | 45318 | 39614 | 3167 | 2537 | 44620 | 39386 | 2894 | 2340 | -0.08* | -0.07* | -0.16* | 0.11* | -0.11* | -0.09* | -0.12* | 0.11* |
| Length of bikeways covered by trees within green space | w0 Sum | 45318 | 39614 | 3167 | 2537 | 44620 | 39386 | 2894 | 2340 | -0.03* | -0.03* | 0 | 0.14* | -0.01× | -0.01 | 0.09* | 0.15* |
| Length of pathways greater than 6 degrees gradient within green space | w0 Sum | 46098 | 40074 | 3332 | 2692 | 46833 | 40711 | 3395 | 2727 | -0.21* | -0.22* | -0.18* | 0.03 | -0.24* | -0.25* | -0.21* | 0.08* |
| Length of bikeways greater than 6 degrees gradient within green space | w0 Sum | 46098 | 40074 | 3332 | 2692 | 46833 | 40711 | 3395 | 2727 | -0.09* | -0.10* | -0.08* | 0.04 | -0.07* | -0.08* | -0.08* | 0.11* |
| Number of Railway Stations within and surrounding a green space (within 100m buffer) | w0 Sum | 46245 | 40198 | 3345 | 2702 | 47079 | 40852 | 3441 | 2786 | 0.04* | 0.05* | 0.02 | -0.05+ | 0.06* | 0.09* | 0.01 | 0.01 |
| Number of Bus Stops within and surrounding a green space (within 100m buffer) | w0 Sum | 46245 | 40198 | 3345 | 2702 | 47079 | 40852 | 3441 | 2786 | 0.02* | 0.03* | -0.03 | 0.27* | 0.08* | 0.11* | -0.04+ | 0.34* |
| Number of ferry wharves within and surrounding a green space (within 100m buffer) | w0 Sum | 46245 | 40198 | 3345 | 2702 | 47079 | 40852 | 3441 | 2786 | -0.15* | -0.15* | -0.01 |  | -0.19* | -0.19* | -0.09* |  |
| Area of GS which is greater than 6 degrees of Slope | w0 Sum | 46098 | 40074 | 3332 | 2692 | 46833 | 40711 | 3395 | 2727 | -0.30* | -0.33* | -0.22* | -0.08* | -0.29* | -0.32* | -0.24* | -0.05× |
| Length of Main Roads within 100 metres of the Green Space | w0 Sum | 46245 | 40198 | 3345 | 2702 | 47079 | 40852 | 3441 | 2786 | -0.03* | -0.03* | 0.14* | 0.29* | 0 | 0 | 0.18* | 0.43* |
| **Access indices – calculated** | | | | | | | | | | | | | | | | | |
| Ratio of Area(A) to Perimeter (P) | w1 Mean | 46245 | 40198 | 3345 | 2702 | 47079 | 40852 | 3441 | 2786 | 0.04* | 0.01+ | -0.01 | 0.06× | 0.05* | 0.03* | -0.07* | 0.03 |
| Ratio of Perimeter (P) to perimeter standardised by area (A) | w1 Mean | 46245 | 40198 | 3345 | 2702 | 47079 | 40852 | 3441 | 2786 | -0.19* | -0.18* | -0.14* | -0.11* | -0.22* | -0.20* | -0.09* | -0.15* |
| Total number of transport stops/stations/wharves within and surrounding a green space (within 100m buffer) | Formula | 46245 | 40198 | 3345 | 2702 | 47079 | 40852 | 3441 | 2786 | 0.01× | 0.02* | -0.03 | 0.27* | 0.08* | 0.10* | -0.05× | 0.34* |
| Pathway Length per Area of Green Space | Formula | 46245 | 40198 | 3345 | 2702 | 47079 | 40852 | 3441 | 2786 | 0 | 0.02* | -0.12* | 0.13* | -0.04* | -0.02* | -0.14* | 0.16* |
| Bikeway Length per Area of Green Space | Formula | 46245 | 40198 | 3345 | 2702 | 47079 | 40852 | 3441 | 2786 | 0.05* | 0.04* | 0.04+ | 0.10* | 0.09* | 0.08* | 0.07* | 0.11* |
| Trees over Pathway per Area of Green Space | Formula | 45318 | 39614 | 3167 | 2537 | 44620 | 39386 | 2894 | 2340 | -0.09* | -0.08* | -0.17* | 0.06× | -0.15* | -0.13* | -0.15* | 0.03 |
| Trees over Bikeway per Area of Green Space | Formula | 45318 | 39614 | 3167 | 2537 | 44620 | 39386 | 2894 | 2340 | -0.04* | -0.04* | 0 | 0.07* | -0.06* | -0.04* | 0.08* | 0.04+ |
| Percentage of GS which is greater than 6 degrees of Slope | Formula | 46098 | 40074 | 3332 | 2692 | 46833 | 40711 | 3395 | 2727 | -0.46* | -0.49* | -0.34* | -0.40* | -0.49* | -0.52* | -0.34* | -0.44* |
| Pathway Length > 6 degrees per Area of GS | Formula | 46098 | 40074 | 3332 | 2692 | 46833 | 40711 | 3395 | 2727 | -0.23* | -0.24* | -0.20* | -0.02 | -0.30* | -0.31* | -0.25* | -0.03 |
| Bikeway Length > 6 degrees slope per Area of GS | Formula | 46098 | 40074 | 3332 | 2692 | 46833 | 40711 | 3395 | 2727 | -0.09* | -0.10* | -0.09* | -0.03 | -0.10* | -0.10* | -0.09* | -0.04+ |
| **Amenities/Activities indices – original sourced** | | | | | | | | | | | | | | | | | |
| Number of public toilet facilities within and surrounding a green space (within 100m buffer) | w0 Sum | 46245 | 40198 | 3345 | 2702 | 47079 | 40852 | 3441 | 2786 | -0.05* | -0.06* | 0.03 | 0.25* | -0.07* | -0.08* | 0.06* | 0.29* |
| Number of cafes within and surrounding a green space (within 100m buffer) | w0 Sum | 46245 | 40198 | 3345 | 2702 | 47079 | 40852 | 3441 | 2786 | -0.15* | -0.14* | -0.15* | 0.12* | -0.18* | -0.17* | -0.14* | 0.16* |
| Number of hotels/bars within and surrounding a green space (within 100m buffer) | w0 Sum | 46245 | 40198 | 3345 | 2702 | 47079 | 40852 | 3441 | 2786 | -0.10* | -0.09* | -0.06* | 0.04+ | -0.12* | -0.11* | -0.08* | 0.11* |
| Number of restaurants/takeaways within and surrounding a green space (within 100m buffer) | w0 Sum | 46245 | 40198 | 3345 | 2702 | 47079 | 40852 | 3441 | 2786 | -0.03* | -0.02* | 0.07* | 0.06* | -0.05* | -0.04* | 0.04+ | 0.16* |
| Number of restaurants within and surrounding a green space (within 100m buffer) | w0 Sum | 46245 | 40198 | 3345 | 2702 | 47079 | 40852 | 3441 | 2786 | -0.10* | -0.09* | -0.05× | 0.01 | -0.12* | -0.11* | -0.05× | 0.11* |
| Number of takeaways within and surrounding a green space (within 100m buffer) | w0 Sum | 46245 | 40198 | 3345 | 2702 | 47079 | 40852 | 3441 | 2786 | 0.03* | 0.04* | 0.11* | 0.09* | 0.03* | 0.04* | 0.12* | 0.18* |
| Number of supermarkets/greengrocers within and surrounding a green space (within 100m buffer) | w0 Sum | 46245 | 40198 | 3345 | 2702 | 47079 | 40852 | 3441 | 2786 | 0.03* | 0.04* | 0.05× | 0.11* | 0.04* | 0.06* | 0 | 0.13* |
| Presence of parkrun event | Quality | 46245 | 40198 | 3345 | 2702 | 47079 | 40852 | 3441 | 2786 | -0.07* | -0.06* | -0.13* | -0.15* | -0.11* | -0.10* | -0.21* | -0.16* |
| **Beach/Coast indices – original sourced** | | | | | | | | | | | | | | | | | |
| Presence of coastline | Quality | 46245 | 40198 | 3345 | 2702 | 47079 | 40852 | 3441 | 2786 | -0.31* | -0.37* | -0.10* | 0.11* | -0.35* | -0.41* | -0.16* | 0.21* |
| Area of beach within and surrounding a green space (within 100m buffer) | w0 Sum | 46245 | 40198 | 3345 | 2702 | 47079 | 40852 | 3441 | 2786 | -0.23* | -0.27* | -0.12* | -0.10* | -0.31* | -0.35* | -0.17* | -0.05+ |
| **Biodiversity indices – original sourced** | | | | | | | | | | | | | | | | | |
| Number of bird species/species habitat likely to occur within green space | Max | 46245 | 40198 | 3345 | 2702 | 47079 | 40852 | 3441 | 2786 | -0.28* | -0.34* | 0.05× | 0.23* | -0.26* | -0.32* | 0.10* | 0.23* |
| Number of fish species/species habitat likely to occur within green space | Max | 46245 | 40198 | 3345 | 2702 | 47079 | 40852 | 3441 | 2786 | -0.34* | -0.38* | -0.11* | 0.12* | -0.33* | -0.37* | -0.14* | 0.12* |
| Number of flora species/species habitat likely to occur within green space | Max | 46245 | 40198 | 3345 | 2702 | 47079 | 40852 | 3441 | 2786 | -0.25* | -0.33* | 0.12* | 0.32* | -0.22* | -0.30* | 0.15* | 0.20* |
| Number of frog species/species habitat likely to occur within green space | Max | 46245 | 40198 | 3345 | 2702 | 47079 | 40852 | 3441 | 2786 | -0.18* | -0.21* | 0.09* | -0.27* | -0.13* | -0.17* | 0.11* | -0.35* |
| Number of mammal species/species habitat likely to occur within green space | Max | 46245 | 40198 | 3345 | 2702 | 47079 | 40852 | 3441 | 2786 | -0.49* | -0.55* | -0.22* | -0.25* | -0.47* | -0.53* | -0.21* | -0.23* |
| Number of other animal species/species habitat likely to occur within green space | Max | 46245 | 40198 | 3345 | 2702 | 47079 | 40852 | 3441 | 2786 | -0.33* | -0.33* |  |  | -0.34* | -0.34* |  |  |
| Number of reptile species/species habitat likely to occur within green space | Max | 46245 | 40198 | 3345 | 2702 | 47079 | 40852 | 3441 | 2786 | -0.35* | -0.41* | -0.12* | 0.07* | -0.33* | -0.39* | -0.15* | 0.14* |
| Area of land with biodiversity value within green space | w0 Sum | 46245 | 40198 | 3345 | 2702 | 47079 | 40852 | 3441 | 2786 | 0.01× | 0.01+ | 0.12* | 0.06× | 0.09* | 0.11* | 0.14* | 0.10* |
| Area of land with biodiversity value outside of green space within 100m | w0 Sum | 46245 | 40198 | 3345 | 2702 | 47079 | 40852 | 3441 | 2786 | -0.04* | -0.05* | 0.14* | 0.03 | 0.02* | 0.03* | 0.17* | 0.07* |
| **Biodiversity indices – calculated** | | | | | | | | | | | | | | | | | |
| Total number of species/species habitat likely to occur within green space | Formula | 46245 | 40198 | 3345 | 2702 | 47079 | 40852 | 3441 | 2786 | -0.45* | -0.52* | -0.03 | 0.22* | -0.39* | -0.45* | 0 | 0.18* |
| Shannon biodiversity index | Formula | 46245 | 40198 | 3345 | 2702 | 47079 | 40852 | 3441 | 2786 | -0.41* | -0.46* | -0.21* | -0.22* | -0.39* | -0.44* | -0.23* | -0.28* |
| Simpson biodiversity index (range 0 to 1=most diverse) | Formula | 46245 | 40198 | 3345 | 2702 | 47079 | 40852 | 3441 | 2786 | -0.30* | -0.33* | -0.18* | -0.42* | -0.28* | -0.30* | -0.23* | -0.49* |
| Percentage of land with biodiversity value within green space | Formula | 46245 | 40198 | 3345 | 2702 | 47079 | 40852 | 3441 | 2786 | -0.01 | -0.01 | 0.14* | 0.04+ | 0.05* | 0.05* | 0.14* | 0.04+ |
| Percentage of land with biodiversity value outside of green space within 100m | Formula | 46245 | 40198 | 3345 | 2702 | 47079 | 40852 | 3441 | 2786 | -0.06* | -0.07* | 0.15* | 0.01 | -0.02* | -0.03* | 0.16* | 0.01 |
| **Incivilities indices – original sourced** | | | | | | | | | | | | | | | | | |
| Density of alcohol-related assaults | Max | 46245 | 40198 | 3345 | 2702 | 47079 | 40852 | 3441 | 2786 | 0.09* | 0.10* | 0.14* | 0.20* | 0.13* | 0.14* | 0.17* | 0.16* |
| Density of malicious damage | Max | 46245 | 40198 | 3345 | 2702 | 47079 | 40852 | 3441 | 2786 | 0.30* | 0.31* | 0.24* | 0.36* | 0.33* | 0.35* | 0.20* | 0.35* |
| Density of non-domestic violence assaults | Max | 46245 | 40198 | 3345 | 2702 | 47079 | 40852 | 3441 | 2786 | 0.19* | 0.20* | 0.13* | 0.32* | 0.28* | 0.30* | 0.16* | 0.39* |
| Density of robberies | Max | 46245 | 40198 | 3345 | 2702 | 47079 | 40852 | 3441 | 2786 | 0.25* | 0.28* | -0.01 | 0.27* | 0.34* | 0.38* | -0.04+ | 0.25* |
| Density of steal from persons | Max | 46245 | 40198 | 3345 | 2702 | 47079 | 40852 | 3441 | 2786 | 0.11* | 0.12* | -0.01 | 0.19* | 0.16* | 0.18* | -0.05× | 0.27* |
| **Incivilities indices – calculated** | | | | | | | | | | | | | | | | | |
| Mean level of incivilities | Formula | 46245 | 40198 | 3345 | 2702 | 47079 | 40852 | 3441 | 2786 | 0.29* | 0.31* | 0.19* | 0.40* | 0.30* | 0.32* | 0.13* | 0.39* |
| **Land cover indices – original sourced** | | | | | | | | | | | | | | | | | |
| Area of bare earth within green space | w0 Sum | 45800 | 39823 | 3293 | 2684 | 45462 | 39469 | 3284 | 2709 | 0.21* | 0.23* | 0.05× | 0.24* | 0.29* | 0.32* | 0.05× | 0.27* |
| Area of buildings within green space | w0 Sum | 45800 | 39823 | 3293 | 2684 | 45462 | 39469 | 3284 | 2709 | 0.04* | 0.04* | 0.05× | 0.31* | 0.11* | 0.11* | 0.07* | 0.34* |
| Area of built up area within green space | w0 Sum | 45800 | 39823 | 3293 | 2684 | 45462 | 39469 | 3284 | 2709 | 0.02* | 0.01+ | 0.06* | 0.23* | 0.04* | 0.03* | 0.05× | 0.30* |
| Area of open grass within green space | w0 Sum | 45800 | 39823 | 3293 | 2684 | 45462 | 39469 | 3284 | 2709 | 0.19* | 0.18* | 0.13* | 0.36* | 0.30* | 0.29* | 0.17* | 0.45* |
| Area of other vegetation within green space | w0 Sum | 45800 | 39823 | 3293 | 2684 | 45462 | 39469 | 3284 | 2709 | 0.05* | 0.04* | -0.06* | 0.17* | 0.10* | 0.09* | -0.06× | 0.25* |
| Area of roads/paths within green space | w0 Sum | 45800 | 39823 | 3293 | 2684 | 45462 | 39469 | 3284 | 2709 | 0.11* | 0.07* | 0.05× | 0.30* | 0.19* | 0.16* | 0.03+ | 0.41* |
| Area of shadow within green space | w0 Sum | 45800 | 39823 | 3293 | 2684 | 45462 | 39469 | 3284 | 2709 | -0.17* | -0.13* | -0.15* | -0.15* | -0.16* | -0.12* | -0.13* | -0.12* |
| Area of swimming pools within green space | w0 Sum | 45800 | 39823 | 3293 | 2684 | 45462 | 39469 | 3284 | 2709 | -0.11* | -0.09* | -0.15* | 0.11* | 0.02× | 0.04* | -0.06* | 0.15* |
| Area of tree canopy within green space | w0 Sum | 45800 | 39823 | 3293 | 2684 | 45462 | 39469 | 3284 | 2709 | -0.24* | -0.25* | -0.13* | -0.02 | -0.24* | -0.25* | -0.15* | -0.05+ |
| Area of water within green space | w0 Sum | 45800 | 39823 | 3293 | 2684 | 45462 | 39469 | 3284 | 2709 | -0.12* | -0.17* | 0.03 | 0.15* | -0.09* | -0.13* | 0.10* | 0.23* |
| **Land cover indices - calculated** | | | | | | | | | | | | | | | | | |
| Percentage of bare earth within green space | Formula | 45800 | 39823 | 3293 | 2684 | 45462 | 39469 | 3284 | 2709 | 0.30* | 0.33* | 0.10* | 0.14* | 0.33* | 0.37* | 0.09* | 0.12* |
| Percentage of buildings within green space | Formula | 45800 | 39823 | 3293 | 2684 | 45462 | 39469 | 3284 | 2709 | 0.06* | 0.06* | 0.07* | 0.21* | 0.08* | 0.09* | 0.09* | 0.22* |
| Percentage of built up area within green space | Formula | 45800 | 39823 | 3293 | 2684 | 45462 | 39469 | 3284 | 2709 | 0.03* | 0.03* | 0.08* | 0.13* | -0.01 | -0.01 | 0.06* | 0.14* |
| Percentage of open grass within green space | Formula | 45800 | 39823 | 3293 | 2684 | 45462 | 39469 | 3284 | 2709 | 0.32* | 0.31* | 0.27* | 0.34* | 0.38* | 0.38* | 0.29* | 0.44* |
| Percentage of other vegetation within green space | Formula | 45800 | 39823 | 3293 | 2684 | 45462 | 39469 | 3284 | 2709 | 0.08* | 0.08* | -0.11* | -0.15* | 0.03* | 0.03* | -0.10* | -0.18* |
| Percentage of roads/paths within green space | Formula | 45800 | 39823 | 3293 | 2684 | 45462 | 39469 | 3284 | 2709 | 0.14* | 0.10* | 0.10* | 0.26* | 0.19* | 0.16* | 0.06* | 0.26* |
| Percentage of shadow within green space | Formula | 45800 | 39823 | 3293 | 2684 | 45462 | 39469 | 3284 | 2709 | -0.18* | -0.15* | -0.16* | -0.23* | -0.19* | -0.16* | -0.14* | -0.24* |
| Percentage of swimming pools within green space | Formula | 45800 | 39823 | 3293 | 2684 | 45462 | 39469 | 3284 | 2709 | -0.11* | -0.10* | -0.14* | 0.10* | -0.01 | 0.01× | -0.05× | 0.13* |
| Percentage of tree canopy within green space | Formula | 45800 | 39823 | 3293 | 2684 | 45462 | 39469 | 3284 | 2709 | -0.42* | -0.43* | -0.23* | -0.36* | -0.46* | -0.47* | -0.23* | -0.44* |
| Percentage of water within green space | Formula | 45800 | 39823 | 3293 | 2684 | 45462 | 39469 | 3284 | 2709 | -0.13* | -0.17* | 0.04+ | 0.12* | -0.11* | -0.16* | 0.12* | 0.17* |
| **Land use indices – original sourced** | | | | | | | | | | | | | | | | | |
| Area of commercial land use surrounding green space (within 100m doughnut buffer) | w0 Sum | 46245 | 40198 | 3345 | 2702 | 47079 | 40852 | 3441 | 2786 | 0.09* | 0.10* | 0.12* | 0.27* | 0.14* | 0.16* | 0.12* | 0.33* |
| Area of education land use surrounding green space (within 100m doughnut buffer) | w0 Sum | 46245 | 40198 | 3345 | 2702 | 47079 | 40852 | 3441 | 2786 | 0.07* | 0.07* | 0.08* | 0.23* | 0.12* | 0.13* | 0.13* | 0.21* |
| Area of hospital/medical land use surrounding green space (within 100m doughnut buffer) | w0 Sum | 46245 | 40198 | 3345 | 2702 | 47079 | 40852 | 3441 | 2786 | -0.02* | -0.01 | -0.06* | 0.05+ | 0.01× | 0.03* | -0.03 | -0.03 |
| Area of industrial land use surrounding green space (within 100m doughnut buffer) | w0 Sum | 46245 | 40198 | 3345 | 2702 | 47079 | 40852 | 3441 | 2786 | 0.15* | 0.14* | 0.22* | 0.20* | 0.27* | 0.27* | 0.31* | 0.28* |
| Area of other land use surrounding green space (within 100m doughnut buffer) | w0 Sum | 46245 | 40198 | 3345 | 2702 | 47079 | 40852 | 3441 | 2786 | 0.02* | 0 | 0.10* | -0.05+ | 0.04* | 0.02* | 0.17* | -0.15* |
| Area of parkland land use surrounding green space (within 100m doughnut buffer) | w0 Sum | 46245 | 40198 | 3345 | 2702 | 47079 | 40852 | 3441 | 2786 | -0.08* | -0.08* | 0.02 | 0.11* | -0.02* | -0.01 | 0.05× | 0.22* |
| Area of primary production land use surrounding green space (within 100m doughnut buffer) | w0 Sum | 46245 | 40198 | 3345 | 2702 | 47079 | 40852 | 3441 | 2786 | 0.08* | 0.06* | 0.17* | 0.05+ | 0.12* | 0.10* | 0.20* | 0.14* |
| Area of residential land use surrounding green space (within 100m doughnut buffer) | w0 Sum | 46245 | 40198 | 3345 | 2702 | 47079 | 40852 | 3441 | 2786 | -0.04* | -0.02* | -0.01 | 0.21* | 0.03* | 0.06* | -0.04+ | 0.26* |
| Area of transport land use surrounding green space (within 100m doughnut buffer) | w0 Sum | 46245 | 40198 | 3345 | 2702 | 47079 | 40852 | 3441 | 2786 | 0.03* | 0.05* | 0 | 0.08* | 0.02* | 0.05* | 0.02 | 0.07* |
| Area of water land use surrounding green space (within 100m doughnut buffer) | w0 Sum | 46245 | 40198 | 3345 | 2702 | 47079 | 40852 | 3441 | 2786 | -0.03* | -0.03* | 0.12* |  | -0.04* | -0.04* | 0.15* |  |
| Area of tree canopy surrounding green space (within 100m doughnut buffer) | w0 Sum | 45318 | 39614 | 3167 | 2537 | 44620 | 39386 | 2894 | 2340 | -0.22* | -0.21* | -0.06* | 0.09* | -0.23* | -0.22* | -0.07* | 0.09* |
| Area of tree canopy ≤4m surrounding green space (within 100m doughnut buffer) | w0 Sum | 45318 | 39614 | 3167 | 2537 | 44620 | 39386 | 2894 | 2340 | -0.11* | -0.09* | 0 | 0.12* | -0.08* | -0.06* | 0.07* | 0.11* |
| Area of tree canopy >4m surrounding green space (within 100m doughnut buffer) | w0 Sum | 45318 | 39614 | 3167 | 2537 | 44620 | 39386 | 2894 | 2340 | -0.23* | -0.22* | -0.07* | 0.09* | -0.25* | -0.24* | -0.08* | 0.08* |
| **Land use indices – calculated** | | | | | | | | | | | | | | | | | |
| Percentage of commercial land use surrounding green space (within 100m doughnut buffer) | Formula | 46245 | 40198 | 3345 | 2702 | 47079 | 40852 | 3441 | 2786 | 0.12* | 0.12* | 0.12* | 0.25* | 0.17* | 0.17* | 0.12* | 0.29* |
| Percentage of education land use surrounding green space (within 100m doughnut buffer) | Formula | 46245 | 40198 | 3345 | 2702 | 47079 | 40852 | 3441 | 2786 | 0.09* | 0.09* | 0.10* | 0.18* | 0.15* | 0.14* | 0.15* | 0.11* |
| Percentage of hospital/medical land use surrounding green space (within 100m doughnut buffer) | Formula | 46245 | 40198 | 3345 | 2702 | 47079 | 40852 | 3441 | 2786 | -0.01× | -0.01 | -0.06* | 0.05+ | 0.01× | 0.03* | -0.03+ | -0.04+ |
| Percentage of industrial land use surrounding green space (within 100m doughnut buffer) | Formula | 46245 | 40198 | 3345 | 2702 | 47079 | 40852 | 3441 | 2786 | 0.16* | 0.15* | 0.21* | 0.19* | 0.28* | 0.28* | 0.30* | 0.23* |
| Percentage of other land use surrounding green space (within 100m doughnut buffer) | Formula | 46245 | 40198 | 3345 | 2702 | 47079 | 40852 | 3441 | 2786 | 0.02* | 0 | 0.10* | -0.06× | 0.04* | 0.02* | 0.16* | -0.18* |
| Percentage of parkland land use surrounding green space (within 100m doughnut buffer) | Formula | 46245 | 40198 | 3345 | 2702 | 47079 | 40852 | 3441 | 2786 | -0.08* | -0.10* | 0.06× | -0.05× | -0.07* | -0.09* | 0.10* | 0.02 |
| Percentage of primary production land use surrounding green space (within 100m doughnut buffer) | Formula | 46245 | 40198 | 3345 | 2702 | 47079 | 40852 | 3441 | 2786 | 0.07* | 0.06* | 0.17* | 0.05+ | 0.11* | 0.10* | 0.20* | 0.14* |
| Percentage of residential land use surrounding green space (within 100m doughnut buffer) | Formula | 46245 | 40198 | 3345 | 2702 | 47079 | 40852 | 3441 | 2786 | -0.02* | 0 | -0.12* | -0.09* | -0.04* | -0.01× | -0.12* | -0.15* |
| Percentage of transport land use surrounding green space (within 100m doughnut buffer) | Formula | 46245 | 40198 | 3345 | 2702 | 47079 | 40852 | 3441 | 2786 | 0.03* | 0.05* | 0 | 0.08* | 0.03* | 0.06* | 0.02 | 0.07* |
| Percentage of water land use surrounding green space (within 100m doughnut buffer) | Formula | 46245 | 40198 | 3345 | 2702 | 47079 | 40852 | 3441 | 2786 | -0.03* | -0.03* | 0.12* |  | -0.04* | -0.04* | 0.15* |  |
| Percentage of tree canopy to roads surrounding green space (within 100m doughnut buffer) | Formula | 45318 | 39614 | 3167 | 2537 | 44620 | 39386 | 2894 | 2340 | -0.47* | -0.49* | -0.17* | -0.36* | -0.52* | -0.54* | -0.20* | -0.40* |
| Percentage of tree canopy ≤4m to roads surrounding green space (within 100m doughnut buffer) | Formula | 45318 | 39614 | 3167 | 2537 | 44620 | 39386 | 2894 | 2340 | -0.21* | -0.20* | -0.03 | -0.38* | -0.25* | -0.24* | 0.06× | -0.41* |
| Percentage of tree canopy >4m to roads surrounding green space (within 100m doughnut buffer) | Formula | 45318 | 39614 | 3167 | 2537 | 44620 | 39386 | 2894 | 2340 | -0.47* | -0.49* | -0.18* | -0.32* | -0.51* | -0.53* | -0.20* | -0.37* |
| Percentage of tree canopy surrounding green space (within 100m doughnut buffer) | Formula | 45318 | 39614 | 3167 | 2537 | 44620 | 39386 | 2894 | 2340 | -0.39* | -0.39* | -0.16* | -0.25* | -0.44* | -0.45* | -0.16* | -0.31* |
| Percentage of tree canopy ≤4m surrounding green space (within 100m doughnut buffer) | Formula | 45318 | 39614 | 3167 | 2537 | 44620 | 39386 | 2894 | 2340 | -0.15* | -0.13* | -0.03 | -0.23* | -0.20* | -0.17* | 0.03 | -0.30* |
| Percentage of tree canopy >4m surrounding green space (within 100m doughnut buffer) | Formula | 45318 | 39614 | 3167 | 2537 | 44620 | 39386 | 2894 | 2340 | -0.39* | -0.40* | -0.17* | -0.24* | -0.45* | -0.46* | -0.16* | -0.29* |

Method of aggregating intersects over mesh blocks: Count=unweighted count | Sum=unweighted sum | w0 Sum = sum weighted by the ratio of the green space intersect with the buffer area to the green space area | w1 Mean = mean weighted by the ratio of the intersect area to the sum of all intersect areas | Max = maximum value over the intersects within the buffer | Quality = Value 0/1, aggregated to 1 if any intersects equal 1 (technically Quality = Max) | Formula = calculation of the index from other qualities that have already been aggregated as weighted sums or weighted means.

P-value signs: * ≤0.001, × ≤0.01, + ≤0.05

GS = green space

Table S6. Spearman's rank coefficient of correlation between SEIFA IRSD score (reversed) and aggregated over MB green space features with full p-values – 100m and 200m buffers.

|  |  | **Buffer 100m** | | | | | | | | **Buffer 200m** | | | | | | | |
| --- | --- | --- | --- | --- | --- | --- | --- | --- | --- | --- | --- | --- | --- | --- | --- | --- | --- |
|  |  | **Full set** | | **Sydney** | | **Newcastle** | | **Wollongong** | | **Full set** | | **Sydney** | | **Newcastle** | | **Wollongong** | |
| **Index description** | **Method** | **Rho** | **P** | **Rho** | **P** | **Rho** | **P** | **Rho** | **P** | **Rho** | **P** | **Rho** | **P** | **Rho** | **P** | **Rho** | **P** |
| **General indices** | | | | | | | | | | | | | | | | | |
| N green spaces in MB buffer | Count | -0.09 | ≤0.001 | -0.09 | ≤0.001 | -0.01 | 0.78 | -0.04 | 0.383 | -0.1 | ≤0.001 | -0.09 | ≤0.001 | -0.05 | ≤0.05 | 0.05 | 0.091 |
| Intersect of buffer with green space | Sum | -0.02 | ≤0.01 | -0.03 | ≤0.01 | -0.1 | ≤0.05 | 0.03 | 0.48 | -0.02 | ≤0.05 | -0.03 | ≤0.001 | -0.06 | ≤0.05 | 0.04 | 0.233 |
| Green space area (A) | Sum | 0.01 | 0.15 | -0.01 | 0.147 | -0.03 | 0.423 | -0.03 | 0.567 | 0.01 | 0.315 | -0.02 | ≤0.01 | -0.06 | ≤0.05 | 0.01 | 0.625 |
| GS perimeter (P) | w0 Sum | -0.07 | ≤0.001 | -0.06 | ≤0.001 | -0.09 | ≤0.05 | 0.04 | 0.437 | -0.07 | ≤0.001 | -0.06 | ≤0.001 | -0.05 | ≤0.05 | 0.04 | 0.173 |
| **Access indices – original sourced** | | | | | | | | | | | | | | | | | |
| Length of pathways within green space | w0 Sum | 0 | 0.754 | -0.01 | 0.525 | -0.12 | ≤0.01 | 0.1 | ≤0.05 | 0 | 0.733 | 0 | 0.672 | -0.15 | ≤0.001 | 0.12 | ≤0.001 |
| Length of bikeways within green space | w0 Sum | 0.02 | ≤0.05 | 0 | 0.909 | -0.05 | 0.225 | 0.04 | 0.406 | 0.02 | ≤0.001 | 0 | 0.884 | -0.05 | 0.085 | 0.07 | ≤0.05 |
| Length of pathways covered by trees within green space | w0 Sum | -0.03 | ≤0.01 | -0.03 | ≤0.05 | -0.14 | ≤0.001 | 0.04 | 0.394 | -0.03 | ≤0.001 | -0.02 | ≤0.001 | -0.18 | ≤0.001 | 0.08 | ≤0.01 |
| Length of bikeways covered by trees within green space | w0 Sum | 0 | 0.639 | -0.01 | 0.194 | -0.09 | ≤0.05 | 0.05 | 0.314 | 0 | 0.546 | -0.01 | 0.112 | -0.06 | ≤0.05 | 0.06 | 0.055 |
| Length of pathways greater than 6 degrees gradient within green space | w0 Sum | -0.13 | ≤0.001 | -0.14 | ≤0.001 | -0.19 | ≤0.001 | -0.03 | 0.567 | -0.13 | ≤0.001 | -0.14 | ≤0.001 | -0.22 | ≤0.001 | -0.01 | 0.698 |
| Length of bikeways greater than 6 degrees gradient within green space | w0 Sum | -0.03 | ≤0.01 | -0.04 | ≤0.001 | -0.12 | ≤0.01 | -0.09 | 0.057 | -0.04 | ≤0.001 | -0.05 | ≤0.001 | -0.12 | ≤0.001 | -0.04 | 0.144 |
| Number of Railway Stations within and surrounding a green space (within 100m buffer) | w0 Sum | -0.01 | 0.36 | -0.01 | 0.429 | 0 | 0.942 | -0.01 | 0.803 | -0.01 | 0.437 | 0 | 0.991 | -0.02 | 0.426 | -0.05 | 0.108 |
| Number of Bus Stops within and surrounding a green space (within 100m buffer) | w0 Sum | 0 | 0.966 | 0 | 0.981 | -0.04 | 0.32 | 0.07 | 0.104 | -0.01 | ≤0.05 | -0.02 | ≤0.05 | -0.06 | ≤0.05 | 0.12 | ≤0.001 |
| Number of ferry wharves within and surrounding a green space (within 100m buffer) | w0 Sum | -0.09 | ≤0.001 | -0.09 | ≤0.001 | 0.08 | ≤0.05 |  |  | -0.09 | ≤0.001 | -0.09 | ≤0.001 | 0.05 | ≤0.05 |  |  |
| Area of GS which is greater than 6 degrees of Slope | w0 Sum | -0.31 | ≤0.001 | -0.33 | ≤0.001 | -0.26 | ≤0.001 | -0.28 | ≤0.001 | -0.29 | ≤0.001 | -0.31 | ≤0.001 | -0.24 | ≤0.001 | -0.25 | ≤0.001 |
| Length of Main Roads within 100 metres of the Green Space | w0 Sum | -0.02 | ≤0.05 | -0.02 | 0.07 | 0.09 | ≤0.05 | 0.02 | 0.717 | -0.03 | ≤0.001 | -0.03 | ≤0.001 | 0.07 | ≤0.01 | 0.1 | ≤0.001 |
| **Access indices - calculated** | | | | | | | | | | | | | | | | | |
| Ratio of Area(A) to Perimeter (P) | w1 Mean | 0.08 | ≤0.001 | 0.05 | ≤0.001 | 0 | 0.922 | -0.02 | 0.632 | 0.08 | ≤0.001 | 0.05 | ≤0.001 | -0.02 | 0.57 | 0 | 0.962 |
| Ratio of Perimeter (P) to perimeter standardised by area (A) | w1 Mean | -0.13 | ≤0.001 | -0.11 | ≤0.001 | -0.16 | ≤0.001 | -0.1 | ≤0.05 | -0.13 | ≤0.001 | -0.12 | ≤0.001 | -0.15 | ≤0.001 | -0.08 | ≤0.05 |
| Total number of transport stops/stations/wharves within and surrounding a green space (within 100m buffer) | Formula | 0 | 0.625 | 0 | 0.62 | -0.05 | 0.177 | 0.07 | 0.11 | -0.02 | ≤0.01 | -0.02 | ≤0.01 | -0.07 | ≤0.05 | 0.12 | ≤0.001 |
| Pathway Length per Area of Green Space | Formula | 0 | 0.972 | 0 | 0.925 | -0.12 | ≤0.01 | 0.1 | ≤0.05 | 0 | 0.91 | 0 | 0.712 | -0.16 | ≤0.001 | 0.12 | ≤0.001 |
| Bikeway Length per Area of Green Space | Formula | 0.02 | ≤0.05 | 0 | 0.707 | -0.04 | 0.326 | 0.03 | 0.494 | 0.03 | ≤0.001 | 0.01 | 0.33 | -0.02 | 0.386 | 0.06 | ≤0.05 |
| Trees over Pathway per Area of Green Space | Formula | -0.03 | ≤0.01 | -0.03 | ≤0.05 | -0.13 | ≤0.01 | 0.03 | 0.499 | -0.03 | ≤0.001 | -0.02 | ≤0.001 | -0.19 | ≤0.001 | 0.07 | ≤0.05 |
| Trees over Bikeway per Area of Green Space | Formula | 0 | 0.701 | -0.01 | 0.27 | -0.07 | 0.08 | 0.03 | 0.584 | 0 | 0.538 | -0.01 | 0.169 | -0.06 | ≤0.05 | 0.04 | 0.176 |
| Percentage of GS which is greater than 6 degrees of Slope | Formula | -0.41 | ≤0.001 | -0.42 | ≤0.001 | -0.35 | ≤0.001 | -0.41 | ≤0.001 | -0.4 | ≤0.001 | -0.42 | ≤0.001 | -0.33 | ≤0.001 | -0.41 | ≤0.001 |
| Pathway Length > 6 degrees per Area of GS | Formula | -0.14 | ≤0.001 | -0.15 | ≤0.001 | -0.21 | ≤0.001 | -0.03 | 0.471 | -0.14 | ≤0.001 | -0.14 | ≤0.001 | -0.23 | ≤0.001 | -0.02 | 0.517 |
| Bikeway Length > 6 degrees slope per Area of GS | Formula | -0.03 | ≤0.01 | -0.04 | ≤0.001 | -0.13 | ≤0.001 | -0.1 | ≤0.05 | -0.04 | ≤0.001 | -0.05 | ≤0.001 | -0.12 | ≤0.001 | -0.05 | 0.079 |
| **Amenities/Activities indices – original sourced** | | | | | | | | | | | | | | | | | |
| Number of public toilet facilities within and surrounding a green space (within 100m buffer) | w0 Sum | -0.05 | ≤0.001 | -0.07 | ≤0.001 | 0.04 | 0.378 | 0.07 | 0.116 | -0.05 | ≤0.001 | -0.06 | ≤0.001 | -0.03 | 0.303 | 0.07 | ≤0.05 |
| Number of cafes within and surrounding a green space (within 100m buffer) | w0 Sum | -0.09 | ≤0.001 | -0.09 | ≤0.001 | -0.12 | ≤0.01 | 0.02 | 0.589 | -0.11 | ≤0.001 | -0.1 | ≤0.001 | -0.14 | ≤0.001 | 0.03 | 0.327 |
| Number of hotels/bars within and surrounding a green space (within 100m buffer) | w0 Sum | -0.03 | ≤0.01 | -0.02 | ≤0.05 | -0.04 | 0.267 |  |  | -0.04 | ≤0.001 | -0.04 | ≤0.001 | -0.06 | ≤0.05 | 0.03 | 0.282 |
| Number of restaurants/takeaways within and surrounding a green space (within 100m buffer) | w0 Sum | -0.06 | ≤0.001 | -0.06 | ≤0.001 | -0.04 | 0.349 | -0.04 | 0.438 | -0.06 | ≤0.001 | -0.05 | ≤0.001 | -0.02 | 0.439 | -0.04 | 0.242 |
| Number of restaurants within and surrounding a green space (within 100m buffer) | w0 Sum | -0.11 | ≤0.001 | -0.11 | ≤0.001 | -0.1 | ≤0.05 | -0.1 | ≤0.05 | -0.11 | ≤0.001 | -0.1 | ≤0.001 | -0.09 | ≤0.001 | -0.09 | ≤0.01 |
| Number of takeaways within and surrounding a green space (within 100m buffer) | w0 Sum | 0.01 | 0.174 | 0.02 | 0.072 | 0.04 | 0.358 | 0.06 | 0.18 | 0.01 | 0.279 | 0.01 | ≤0.05 | 0.02 | 0.371 | 0.05 | 0.07 |
| Number of supermarkets/greengrocers within and surrounding a green space (within 100m buffer) | w0 Sum | 0.02 | ≤0.05 | 0.02 | 0.051 | 0.04 | 0.282 | 0.12 | ≤0.01 | 0.02 | ≤0.05 | 0.02 | ≤0.01 | 0.01 | 0.715 | 0.1 | ≤0.001 |
| Presence of parkrun event | Quality | -0.01 | 0.237 | -0.01 | 0.417 | -0.1 | ≤0.05 | -0.08 | 0.073 | -0.02 | ≤0.001 | -0.02 | ≤0.01 | -0.08 | ≤0.01 | -0.11 | ≤0.001 |
| **Beach/Coast – original sourced** | | | | | | | | | | | | | | | | | |
| Presence of coastline | Quality | -0.25 | ≤0.001 | -0.29 | ≤0.001 | -0.04 | 0.34 | -0.07 | 0.137 | -0.25 | ≤0.001 | -0.28 | ≤0.001 | -0.07 | ≤0.01 | -0.07 | ≤0.05 |
| Area of beach within and surrounding a green space (within 100m buffer) | w0 Sum | -0.16 | ≤0.001 | -0.18 | ≤0.001 | -0.14 | ≤0.001 | -0.23 | ≤0.001 | -0.15 | ≤0.001 | -0.17 | ≤0.001 | -0.11 | ≤0.001 | -0.2 | ≤0.001 |
| **Biodiversity indices – original sourced** | | | | | | | | | | | | | | | | | |
| Number of bird species/species habitat likely to occur within green space | Max | -0.3 | ≤0.001 | -0.34 | ≤0.001 | 0.02 | 0.616 | 0.15 | ≤0.01 | -0.29 | ≤0.001 | -0.34 | ≤0.001 | 0 | 0.977 | 0.13 | ≤0.001 |
| Number of fish species/species habitat likely to occur within green space | Max | -0.32 | ≤0.001 | -0.36 | ≤0.001 | -0.09 | ≤0.05 | 0.02 | 0.668 | -0.31 | ≤0.001 | -0.35 | ≤0.001 | -0.11 | ≤0.001 | 0 | 0.982 |
| Number of flora species/species habitat likely to occur within green space | Max | -0.28 | ≤0.001 | -0.36 | ≤0.001 | 0.05 | 0.261 | 0.28 | ≤0.001 | -0.27 | ≤0.001 | -0.35 | ≤0.001 | 0.04 | 0.115 | 0.29 | ≤0.001 |
| Number of frog species/species habitat likely to occur within green space | Max | -0.27 | ≤0.001 | -0.31 | ≤0.001 | 0.08 | 0.056 | -0.14 | ≤0.01 | -0.26 | ≤0.001 | -0.29 | ≤0.001 | 0.06 | ≤0.05 | -0.2 | ≤0.001 |
| Number of mammal species/species habitat likely to occur within green space | Max | -0.51 | ≤0.001 | -0.55 | ≤0.001 | -0.18 | ≤0.001 | -0.3 | ≤0.001 | -0.51 | ≤0.001 | -0.55 | ≤0.001 | -0.18 | ≤0.001 | -0.3 | ≤0.001 |
| Number of other animal species/species habitat likely to occur within green space | Max | -0.26 | ≤0.001 | -0.26 | ≤0.001 |  |  |  |  | -0.28 | ≤0.001 | -0.28 | ≤0.001 |  |  |  |  |
| Number of reptile species/species habitat likely to occur within green space | Max | -0.35 | ≤0.001 | -0.39 | ≤0.001 | -0.09 | ≤0.05 | -0.02 | 0.616 | -0.34 | ≤0.001 | -0.39 | ≤0.001 | -0.12 | ≤0.001 | -0.06 | ≤0.05 |
| Area of land with biodiversity value within green space | w0 Sum | -0.01 | 0.33 | -0.02 | 0.099 | 0.07 | 0.079 | -0.01 | 0.867 | 0 | 0.933 | 0 | 0.504 | 0.06 | ≤0.05 | -0.01 | 0.805 |
| Area of land with biodiversity value outside of green space within 100m | w0 Sum | -0.03 | ≤0.001 | -0.04 | ≤0.001 | 0.08 | ≤0.05 | -0.01 | 0.864 | -0.02 | ≤0.01 | -0.02 | ≤0.001 | 0.07 | ≤0.01 | -0.02 | 0.552 |
| **Biodiversity indices – calculated** | | | | | | | | | | | | | | | | | |
| Total number of species/species habitat likely to occur within green space | Formula | -0.48 | ≤0.001 | -0.54 | ≤0.001 | -0.05 | 0.244 | 0.13 | ≤0.01 | -0.48 | ≤0.001 | -0.54 | ≤0.001 | -0.05 | ≤0.05 | 0.11 | ≤0.001 |
| Shannon biodiversity index | Formula | -0.43 | ≤0.001 | -0.47 | ≤0.001 | -0.18 | ≤0.001 | -0.28 | ≤0.001 | -0.42 | ≤0.001 | -0.46 | ≤0.001 | -0.21 | ≤0.001 | -0.29 | ≤0.001 |
| Simpson biodiversity index (range 0 to 1=most diverse) | Formula | -0.32 | ≤0.001 | -0.35 | ≤0.001 | -0.15 | ≤0.001 | -0.38 | ≤0.001 | -0.31 | ≤0.001 | -0.35 | ≤0.001 | -0.13 | ≤0.001 | -0.36 | ≤0.001 |
| Percentage of land with biodiversity value within green space | Formula | -0.01 | 0.313 | -0.02 | 0.08 | 0.1 | ≤0.05 | 0 | 0.919 | 0 | 0.813 | -0.01 | 0.4 | 0.08 | ≤0.01 | -0.01 | 0.766 |
| Percentage of land with biodiversity value outside of green space within 100m | Formula | -0.03 | ≤0.001 | -0.04 | ≤0.001 | 0.12 | ≤0.01 | 0 | 0.944 | -0.02 | ≤0.001 | -0.03 | ≤0.001 | 0.1 | ≤0.001 | -0.02 | 0.422 |
| **Incivilities indices – original sourced** | | | | | | | | | | | | | | | | | |
| Density of alcohol-related assaults | Max | 0.06 | ≤0.001 | 0.08 | ≤0.001 | 0.05 | 0.177 | 0.02 | 0.605 | 0.07 | ≤0.001 | 0.08 | ≤0.001 | 0.05 | ≤0.05 | 0.08 | ≤0.01 |
| Density of malicious damage | Max | 0.25 | ≤0.001 | 0.26 | ≤0.001 | 0.17 | ≤0.001 | 0.3 | ≤0.001 | 0.25 | ≤0.001 | 0.26 | ≤0.001 | 0.23 | ≤0.001 | 0.37 | ≤0.001 |
| Density of non-domestic violence assaults | Max | 0.13 | ≤0.001 | 0.14 | ≤0.001 | 0.06 | 0.16 | 0.11 | ≤0.05 | 0.13 | ≤0.001 | 0.14 | ≤0.001 | 0.06 | ≤0.05 | 0.17 | ≤0.001 |
| Density of robberies | Max | 0.18 | ≤0.001 | 0.19 | ≤0.001 | 0.02 | 0.552 | 0.16 | ≤0.001 | 0.18 | ≤0.001 | 0.19 | ≤0.001 | 0.01 | 0.809 | 0.18 | ≤0.001 |
| Density of steal from persons | Max | 0.07 | ≤0.001 | 0.08 | ≤0.001 | 0.01 | 0.769 | 0.06 | 0.173 | 0.07 | ≤0.001 | 0.09 | ≤0.001 | 0.01 | 0.786 | 0.12 | ≤0.001 |
| **Incivilities indices – calculated** | | | | | | | | | | | | | | | | | |
| Mean level of incivilities | Formula | 0.24 | ≤0.001 | 0.25 | ≤0.001 | 0.16 | ≤0.001 | 0.32 | ≤0.001 | 0.25 | ≤0.001 | 0.25 | ≤0.001 | 0.2 | ≤0.001 | 0.39 | ≤0.001 |
| **Land cover indices – original sourced** | | | | | | | | | | | | | | | | | |
| Area of bare earth within green space | w0 Sum | 0.19 | ≤0.001 | 0.19 | ≤0.001 | -0.03 | 0.416 | 0.11 | ≤0.05 | 0.18 | ≤0.001 | 0.19 | ≤0.001 | -0.01 | 0.6 | 0.1 | ≤0.001 |
| Area of buildings within green space | w0 Sum | 0.02 | 0.104 | 0.01 | 0.249 | 0.01 | 0.876 | 0.05 | 0.29 | 0.02 | ≤0.01 | 0.01 | 0.12 | 0.03 | 0.256 | 0.12 | ≤0.001 |
| Area of built up area within green space | w0 Sum | 0.01 | 0.579 | 0 | 0.709 | -0.04 | 0.338 | 0.01 | 0.77 | 0 | 0.942 | -0.01 | 0.369 | -0.01 | 0.844 | 0.08 | ≤0.05 |
| Area of open grass within green space | w0 Sum | 0.17 | ≤0.001 | 0.16 | ≤0.001 | 0.05 | 0.212 | 0.18 | ≤0.001 | 0.18 | ≤0.001 | 0.17 | ≤0.001 | 0.08 | ≤0.01 | 0.21 | ≤0.001 |
| Area of other vegetation within green space | w0 Sum | 0.03 | ≤0.01 | 0.03 | ≤0.05 | -0.13 | ≤0.01 | -0.03 | 0.575 | 0.03 | ≤0.001 | 0.02 | ≤0.05 | -0.1 | ≤0.001 | -0.02 | 0.5 |
| Area of roads/paths within green space | w0 Sum | 0.06 | ≤0.001 | 0.03 | ≤0.01 | -0.02 | 0.588 | 0.14 | ≤0.01 | 0.06 | ≤0.001 | 0.03 | ≤0.001 | 0 | 0.984 | 0.13 | ≤0.001 |
| Area of shadow within green space | w0 Sum | -0.11 | ≤0.001 | -0.09 | ≤0.001 | -0.07 | 0.071 | -0.17 | ≤0.001 | -0.12 | ≤0.001 | -0.09 | ≤0.001 | -0.12 | ≤0.001 | -0.19 | ≤0.001 |
| Area of swimming pools within green space | w0 Sum | -0.09 | ≤0.001 | -0.08 | ≤0.001 | -0.17 | ≤0.001 | 0.06 | 0.199 | -0.09 | ≤0.001 | -0.08 | ≤0.001 | -0.19 | ≤0.001 | 0.06 | ≤0.05 |
| Area of tree canopy within green space | w0 Sum | -0.22 | ≤0.001 | -0.22 | ≤0.001 | -0.15 | ≤0.001 | -0.12 | ≤0.05 | -0.22 | ≤0.001 | -0.23 | ≤0.001 | -0.13 | ≤0.001 | -0.13 | ≤0.001 |
| Area of water within green space | w0 Sum | -0.09 | ≤0.001 | -0.12 | ≤0.001 | 0.05 | 0.265 | -0.07 | 0.122 | -0.09 | ≤0.001 | -0.13 | ≤0.001 | -0.01 | 0.753 | -0.05 | 0.131 |
| **Land cover indices - calculated** | | | | | | | | | | | | | | | | | |
| Percentage of bare earth within green space | Formula | 0.27 | ≤0.001 | 0.28 | ≤0.001 | 0.04 | 0.303 | 0.12 | ≤0.05 | 0.27 | ≤0.001 | 0.29 | ≤0.001 | 0.03 | 0.332 | 0.09 | ≤0.01 |
| Percentage of buildings within green space | Formula | 0.03 | ≤0.01 | 0.03 | ≤0.01 | 0.05 | 0.182 | 0.08 | 0.101 | 0.04 | ≤0.001 | 0.03 | ≤0.001 | 0.08 | ≤0.01 | 0.15 | ≤0.001 |
| Percentage of built up area within green space | Formula | 0.02 | ≤0.05 | 0.02 | ≤0.05 | 0 | 0.931 | 0 | 0.965 | 0.02 | ≤0.05 | 0.02 | ≤0.01 | 0.03 | 0.322 | 0.08 | ≤0.01 |
| Percentage of open grass within green space | Formula | 0.29 | ≤0.001 | 0.28 | ≤0.001 | 0.28 | ≤0.001 | 0.34 | ≤0.001 | 0.31 | ≤0.001 | 0.3 | ≤0.001 | 0.27 | ≤0.001 | 0.38 | ≤0.001 |
| Percentage of other vegetation within green space | Formula | 0.08 | ≤0.001 | 0.07 | ≤0.001 | -0.09 | ≤0.05 | -0.17 | ≤0.001 | 0.07 | ≤0.001 | 0.07 | ≤0.001 | -0.13 | ≤0.001 | -0.16 | ≤0.001 |
| Percentage of roads/paths within green space | Formula | 0.08 | ≤0.001 | 0.04 | ≤0.001 | 0.04 | 0.32 | 0.22 | ≤0.001 | 0.09 | ≤0.001 | 0.05 | ≤0.001 | 0.06 | ≤0.05 | 0.24 | ≤0.001 |
| Percentage of shadow within green space | Formula | -0.11 | ≤0.001 | -0.09 | ≤0.001 | -0.08 | 0.054 | -0.19 | ≤0.001 | -0.12 | ≤0.001 | -0.1 | ≤0.001 | -0.12 | ≤0.001 | -0.2 | ≤0.001 |
| Percentage of swimming pools within green space | Formula | -0.08 | ≤0.001 | -0.08 | ≤0.001 | -0.16 | ≤0.001 | 0.06 | 0.216 | -0.09 | ≤0.001 | -0.08 | ≤0.001 | -0.18 | ≤0.001 | 0.06 | ≤0.05 |
| Percentage of tree canopy within green space | Formula | -0.36 | ≤0.001 | -0.36 | ≤0.001 | -0.16 | ≤0.001 | -0.24 | ≤0.001 | -0.38 | ≤0.001 | -0.38 | ≤0.001 | -0.18 | ≤0.001 | -0.3 | ≤0.001 |
| Percentage of water within green space | Formula | -0.08 | ≤0.001 | -0.12 | ≤0.001 | 0.06 | 0.144 | -0.08 | 0.083 | -0.09 | ≤0.001 | -0.13 | ≤0.001 | 0 | 0.973 | -0.05 | 0.105 |
| **Land use indices – original sourced** | | | | | | | | | | | | | | | | | |
| Area of commercial land use surrounding green space (within 100m doughnut buffer) | w0 Sum | 0.05 | ≤0.001 | 0.06 | ≤0.001 | 0.06 | 0.151 | 0.17 | ≤0.001 | 0.05 | ≤0.001 | 0.05 | ≤0.001 | 0.04 | 0.099 | 0.17 | ≤0.001 |
| Area of education land use surrounding green space (within 100m doughnut buffer) | w0 Sum | 0.06 | ≤0.001 | 0.05 | ≤0.001 | 0.13 | ≤0.01 | 0.09 | ≤0.05 | 0.04 | ≤0.001 | 0.04 | ≤0.001 | 0.11 | ≤0.001 | 0.14 | ≤0.001 |
| Area of hospital/medical land use surrounding green space (within 100m doughnut buffer) | w0 Sum | 0.02 | ≤0.05 | 0.03 | ≤0.01 | -0.08 | ≤0.05 | 0.01 | 0.776 | 0 | 0.505 | 0 | 0.812 | -0.08 | ≤0.01 | 0 | 0.922 |
| Area of industrial land use surrounding green space (within 100m doughnut buffer) | w0 Sum | 0.09 | ≤0.001 | 0.08 | ≤0.001 | 0.1 | ≤0.05 | 0.19 | ≤0.001 | 0.09 | ≤0.001 | 0.08 | ≤0.001 | 0.1 | ≤0.001 | 0.14 | ≤0.001 |
| Area of other land use surrounding green space (within 100m doughnut buffer) | w0 Sum | 0.01 | 0.499 | 0 | 0.669 | 0.12 | ≤0.01 | -0.07 | 0.124 | 0.01 | 0.339 | 0 | 0.613 | 0.1 | ≤0.001 | -0.11 | ≤0.001 |
| Area of parkland land use surrounding green space (within 100m doughnut buffer) | w0 Sum | -0.06 | ≤0.001 | -0.06 | ≤0.001 | -0.14 | ≤0.001 | -0.13 | ≤0.01 | -0.04 | ≤0.001 | -0.04 | ≤0.001 | -0.09 | ≤0.01 | -0.09 | ≤0.01 |
| Area of primary production land use surrounding green space (within 100m doughnut buffer) | w0 Sum | 0.05 | ≤0.001 | 0.05 | ≤0.001 | 0.12 | ≤0.01 | -0.06 | 0.215 | 0.05 | ≤0.001 | 0.04 | ≤0.001 | 0.13 | ≤0.001 | -0.03 | 0.379 |
| Area of residential land use surrounding green space (within 100m doughnut buffer) | w0 Sum | -0.06 | ≤0.001 | -0.04 | ≤0.001 | -0.1 | ≤0.05 | 0.04 | 0.423 | -0.07 | ≤0.001 | -0.05 | ≤0.001 | -0.06 | ≤0.05 | 0.05 | 0.103 |
| Area of transport land use surrounding green space (within 100m doughnut buffer) | w0 Sum | 0 | 0.828 | 0.01 | 0.451 | 0.03 | 0.415 | 0.02 | 0.59 | 0 | 0.58 | 0.02 | ≤0.05 | 0 | 0.954 | 0.04 | 0.236 |
| Area of water land use surrounding green space (within 100m doughnut buffer) | w0 Sum | 0 | 0.678 | 0 | 0.633 | 0.09 | ≤0.05 |  |  | 0 | 0.563 | 0 | 0.662 | 0.07 | ≤0.01 |  |  |
| Area of tree canopy surrounding green space (within 100m doughnut buffer) | w0 Sum | -0.16 | ≤0.001 | -0.14 | ≤0.001 | -0.11 | ≤0.01 | -0.02 | 0.641 | -0.18 | ≤0.001 | -0.16 | ≤0.001 | -0.09 | ≤0.001 | -0.02 | 0.467 |
| Area of tree canopy ≤4m surrounding green space (within 100m doughnut buffer) | w0 Sum | -0.1 | ≤0.001 | -0.08 | ≤0.001 | -0.09 | ≤0.05 | -0.01 | 0.748 | -0.11 | ≤0.001 | -0.1 | ≤0.001 | -0.05 | 0.062 | -0.02 | 0.595 |
| Area of tree canopy >4m surrounding green space (within 100m doughnut buffer) | w0 Sum | -0.16 | ≤0.001 | -0.14 | ≤0.001 | -0.11 | ≤0.01 | -0.02 | 0.693 | -0.18 | ≤0.001 | -0.17 | ≤0.001 | -0.1 | ≤0.001 | -0.02 | 0.558 |
| **Land use indices – calculated** | | | | | | | | | | | | | | | | | |
| Percentage of commercial land use surrounding green space (within 100m doughnut buffer) | Formula | 0.06 | ≤0.001 | 0.06 | ≤0.001 | 0.07 | 0.098 | 0.17 | ≤0.001 | 0.06 | ≤0.001 | 0.06 | ≤0.001 | 0.05 | 0.065 | 0.17 | ≤0.001 |
| Percentage of education land use surrounding green space (within 100m doughnut buffer) | Formula | 0.06 | ≤0.001 | 0.06 | ≤0.001 | 0.13 | ≤0.01 | 0.09 | ≤0.05 | 0.05 | ≤0.001 | 0.04 | ≤0.001 | 0.11 | ≤0.001 | 0.14 | ≤0.001 |
| Percentage of hospital/medical land use surrounding green space (within 100m doughnut buffer) | Formula | 0.02 | ≤0.05 | 0.03 | ≤0.01 | -0.08 | ≤0.05 | 0.01 | 0.79 | 0 | 0.509 | 0 | 0.802 | -0.08 | ≤0.01 | 0 | 0.921 |
| Percentage of industrial land use surrounding green space (within 100m doughnut buffer) | Formula | 0.09 | ≤0.001 | 0.08 | ≤0.001 | 0.1 | ≤0.05 | 0.18 | ≤0.001 | 0.09 | ≤0.001 | 0.08 | ≤0.001 | 0.1 | ≤0.001 | 0.14 | ≤0.001 |
| Percentage of other land use surrounding green space (within 100m doughnut buffer) | Formula | 0.01 | 0.49 | 0 | 0.672 | 0.12 | ≤0.01 | -0.07 | 0.136 | 0.01 | 0.331 | 0 | 0.615 | 0.1 | ≤0.001 | -0.11 | ≤0.001 |
| Percentage of parkland land use surrounding green space (within 100m doughnut buffer) | Formula | -0.05 | ≤0.001 | -0.06 | ≤0.001 | -0.03 | 0.417 | -0.14 | ≤0.01 | -0.04 | ≤0.001 | -0.04 | ≤0.001 | -0.04 | 0.16 | -0.12 | ≤0.001 |
| Percentage of primary production land use surrounding green space (within 100m doughnut buffer) | Formula | 0.05 | ≤0.001 | 0.05 | ≤0.001 | 0.12 | ≤0.01 | -0.06 | 0.199 | 0.05 | ≤0.001 | 0.04 | ≤0.001 | 0.13 | ≤0.001 | -0.03 | 0.38 |
| Percentage of residential land use surrounding green space (within 100m doughnut buffer) | Formula | 0 | 0.714 | 0.01 | 0.216 | -0.1 | ≤0.05 | 0 | 0.974 | -0.02 | ≤0.05 | 0 | 0.627 | -0.08 | ≤0.01 | 0 | 0.902 |
| Percentage of transport land use surrounding green space (within 100m doughnut buffer) | Formula | 0 | 0.853 | 0.01 | 0.431 | 0.03 | 0.415 | 0.02 | 0.59 | 0 | 0.507 | 0.02 | ≤0.05 | 0 | 0.951 | 0.04 | 0.236 |
| Percentage of water land use surrounding green space (within 100m doughnut buffer) | Formula | 0 | 0.679 | 0 | 0.636 | 0.09 | ≤0.05 |  |  | 0 | 0.566 | 0 | 0.664 | 0.07 | ≤0.01 |  |  |
| Percentage of tree canopy to roads surrounding green space (within 100m doughnut buffer) | Formula | -0.4 | ≤0.001 | -0.41 | ≤0.001 | -0.05 | 0.241 | -0.22 | ≤0.001 | -0.43 | ≤0.001 | -0.43 | ≤0.001 | -0.08 | ≤0.01 | -0.25 | ≤0.001 |
| Percentage of tree canopy ≤4m to roads surrounding green space (within 100m doughnut buffer) | Formula | -0.18 | ≤0.001 | -0.17 | ≤0.001 | 0.04 | 0.329 | -0.23 | ≤0.001 | -0.19 | ≤0.001 | -0.18 | ≤0.001 | 0.04 | 0.138 | -0.3 | ≤0.001 |
| Percentage of tree canopy >4m to roads surrounding green space (within 100m doughnut buffer) | Formula | -0.4 | ≤0.001 | -0.4 | ≤0.001 | -0.05 | 0.265 | -0.19 | ≤0.001 | -0.42 | ≤0.001 | -0.42 | ≤0.001 | -0.09 | ≤0.001 | -0.21 | ≤0.001 |
| Percentage of tree canopy surrounding green space (within 100m doughnut buffer) | Formula | -0.31 | ≤0.001 | -0.3 | ≤0.001 | -0.05 | 0.239 | -0.1 | ≤0.05 | -0.34 | ≤0.001 | -0.33 | ≤0.001 | -0.07 | ≤0.01 | -0.14 | ≤0.001 |
| Percentage of tree canopy ≤4m surrounding green space (within 100m doughnut buffer) | Formula | -0.12 | ≤0.001 | -0.1 | ≤0.001 | 0.03 | 0.409 | -0.1 | ≤0.05 | -0.14 | ≤0.001 | -0.12 | ≤0.001 | 0.04 | 0.178 | -0.15 | ≤0.001 |
| Percentage of tree canopy >4m surrounding green space (within 100m doughnut buffer) | Formula | -0.31 | ≤0.001 | -0.3 | ≤0.001 | -0.04 | 0.273 | -0.09 | 0.061 | -0.34 | ≤0.001 | -0.33 | ≤0.001 | -0.08 | ≤0.01 | -0.12 | ≤0.001 |

Method of aggregating intersects over mesh blocks: Count=unweighted count | Sum=unweighted sum | w0 Sum = sum weighted by the ratio of the green space intersect with the buffer area to the green space area | w1 Mean = mean weighted by the ratio of the intersect area to the sum of all intersect areas | Max = maximum value over the intersects within the buffer | Quality = Value 0/1, aggregated to 1 if any intersects equal 1 (technically Quality = Max) | Formula = calculation of the index from other qualities that have already been aggregated as weighted sums or weighted means.

P-value signs: * ≤0.001, × ≤0.01, + ≤0.05

GS = green space

Table S7. Spearman's rank coefficient of correlation between SEIFA IRSD score (reversed) and aggregated over MB green space features with full p-values – 300m and 400m buffers.

|  |  | **Buffer 300m** | | | | | | | | **Buffer 400m** | | | | | | | |
| --- | --- | --- | --- | --- | --- | --- | --- | --- | --- | --- | --- | --- | --- | --- | --- | --- | --- |
|  |  | **Full set** | | **Sydney** | | **Newcastle** | | **Wollongong** | | **Full set** | | **Sydney** | | **Newcastle** | | **Wollongong** | |
| **Index description** | **Method** | **Rho** | **P** | **Rho** | **P** | **Rho** | **P** | **Rho** | **P** | **Rho** | **P** | **Rho** | **P** | **Rho** | **P** | **Rho** | **P** |
| **General indices** | | | | | | | | | | | | | | | | | |
| N green spaces in MB buffer | Count | -0.11 | ≤0.001 | -0.11 | ≤0.001 | -0.02 | 0.343 | 0.06 | ≤0.05 | -0.11 | ≤0.001 | -0.11 | ≤0.001 | -0.02 | 0.415 | 0.07 | ≤0.001 |
| Intersect of buffer with green space | Sum | -0.02 | ≤0.001 | -0.04 | ≤0.001 | -0.04 | 0.063 | 0.11 | ≤0.001 | -0.02 | ≤0.001 | -0.04 | ≤0.001 | -0.03 | 0.083 | 0.14 | ≤0.001 |
| Green space area (A) | Sum | -0.02 | ≤0.001 | -0.05 | ≤0.001 | -0.05 | ≤0.05 | 0.07 | ≤0.01 | -0.04 | ≤0.001 | -0.07 | ≤0.001 | -0.03 | 0.098 | 0.08 | ≤0.001 |
| Green space perimeter (P) | w0 Sum | -0.08 | ≤0.001 | -0.08 | ≤0.001 | -0.03 | 0.197 | 0.11 | ≤0.001 | -0.07 | ≤0.001 | -0.07 | ≤0.001 | -0.02 | 0.284 | 0.14 | ≤0.001 |
| **Access indices – original sourced** | | | | | | | | | | | | | | | | | |
| Length of pathways within green space | w0 Sum | -0.02 | ≤0.001 | -0.02 | ≤0.01 | -0.14 | ≤0.001 | 0.14 | ≤0.001 | -0.02 | ≤0.001 | -0.02 | ≤0.001 | -0.12 | ≤0.001 | 0.13 | ≤0.001 |
| Length of bikeways within green space | w0 Sum | 0.01 | ≤0.05 | -0.01 | 0.096 | -0.05 | ≤0.05 | 0.12 | ≤0.001 | 0.01 | 0.136 | -0.01 | ≤0.05 | -0.03 | 0.101 | 0.13 | ≤0.001 |
| Length of pathways covered by trees within green space | w0 Sum | -0.05 | ≤0.001 | -0.05 | ≤0.001 | -0.19 | ≤0.001 | 0.11 | ≤0.001 | -0.06 | ≤0.001 | -0.06 | ≤0.001 | -0.17 | ≤0.001 | 0.1 | ≤0.001 |
| Length of bikeways covered by trees within green space | w0 Sum | -0.02 | ≤0.001 | -0.03 | ≤0.001 | -0.06 | ≤0.01 | 0.1 | ≤0.001 | -0.03 | ≤0.001 | -0.04 | ≤0.001 | -0.04 | ≤0.05 | 0.1 | ≤0.001 |
| Length of pathways greater than 6 degrees gradient within green space | w0 Sum | -0.16 | ≤0.001 | -0.16 | ≤0.001 | -0.21 | ≤0.001 | 0.02 | 0.539 | -0.17 | ≤0.001 | -0.18 | ≤0.001 | -0.19 | ≤0.001 | 0 | 0.905 |
| Length of bikeways greater than 6 degrees gradient within green space | w0 Sum | -0.06 | ≤0.001 | -0.08 | ≤0.001 | -0.11 | ≤0.001 | -0.02 | 0.477 | -0.07 | ≤0.001 | -0.09 | ≤0.001 | -0.09 | ≤0.001 | -0.03 | 0.146 |
| Number of Railway Stations within and surrounding a green space (within 100m buffer) | w0 Sum | 0 | 0.769 | 0.01 | 0.151 | -0.02 | 0.347 | -0.04 | 0.117 | 0.01 | 0.068 | 0.02 | ≤0.001 | -0.01 | 0.529 | -0.05 | ≤0.05 |
| Number of Bus Stops within and surrounding a green space (within 100m buffer) | w0 Sum | -0.02 | ≤0.001 | -0.02 | ≤0.001 | -0.04 | 0.057 | 0.17 | ≤0.001 | -0.02 | ≤0.001 | -0.02 | ≤0.01 | -0.04 | ≤0.05 | 0.2 | ≤0.001 |
| Number of ferry wharves within and surrounding a green space (within 100m buffer) | w0 Sum | -0.1 | ≤0.001 | -0.1 | ≤0.001 | 0.05 | ≤0.05 |  |  | -0.11 | ≤0.001 | -0.11 | ≤0.001 | 0.05 | ≤0.01 |  |  |
| Area of GS which is greater than 6 degrees of Slope | w0 Sum | -0.29 | ≤0.001 | -0.32 | ≤0.001 | -0.22 | ≤0.001 | -0.17 | ≤0.001 | -0.3 | ≤0.001 | -0.33 | ≤0.001 | -0.21 | ≤0.001 | -0.17 | ≤0.001 |
| Length of Main Roads within 100 metres of the Green Space | w0 Sum | -0.04 | ≤0.001 | -0.04 | ≤0.001 | 0.08 | ≤0.001 | 0.14 | ≤0.001 | -0.04 | ≤0.001 | -0.04 | ≤0.001 | 0.08 | ≤0.001 | 0.18 | ≤0.001 |
| **Access indices - calculated** | | | | | | | | | | | | | | | | | |
| Ratio of Area(A) to Perimeter (P) | w1 Mean | 0.06 | ≤0.001 | 0.03 | ≤0.001 | 0 | 0.853 | 0.05 | 0.059 | 0.05 | ≤0.001 | 0.02 | ≤0.01 | 0 | 0.854 | 0.06 | ≤0.01 |
| Ratio of Perimeter (P) to perimeter standardised by area (A) | w1 Mean | -0.15 | ≤0.001 | -0.14 | ≤0.001 | -0.16 | ≤0.001 | -0.06 | ≤0.05 | -0.16 | ≤0.001 | -0.15 | ≤0.001 | -0.15 | ≤0.001 | -0.09 | ≤0.001 |
| Total number of transport stops/stations/wharves within and surrounding a green space (within 100m buffer) | Formula | -0.03 | ≤0.001 | -0.03 | ≤0.001 | -0.05 | ≤0.05 | 0.17 | ≤0.001 | -0.02 | ≤0.001 | -0.02 | ≤0.001 | -0.05 | ≤0.05 | 0.2 | ≤0.001 |
| Pathway Length per Area of Green Space | Formula | -0.01 | ≤0.01 | -0.01 | ≤0.05 | -0.15 | ≤0.001 | 0.13 | ≤0.001 | -0.02 | ≤0.01 | -0.01 | 0.057 | -0.12 | ≤0.001 | 0.11 | ≤0.001 |
| Bikeway Length per Area of Green Space | Formula | 0.02 | ≤0.001 | 0 | 0.91 | -0.04 | 0.098 | 0.1 | ≤0.001 | 0.01 | ≤0.01 | 0 | 0.779 | -0.02 | 0.219 | 0.09 | ≤0.001 |
| Trees over Pathway per Area of Green Space | Formula | -0.05 | ≤0.001 | -0.05 | ≤0.001 | -0.19 | ≤0.001 | 0.1 | ≤0.001 | -0.06 | ≤0.001 | -0.06 | ≤0.001 | -0.17 | ≤0.001 | 0.07 | ≤0.001 |
| Trees over Bikeway per Area of Green Space | Formula | -0.02 | ≤0.001 | -0.02 | ≤0.001 | -0.06 | ≤0.01 | 0.07 | ≤0.01 | -0.03 | ≤0.001 | -0.04 | ≤0.001 | -0.04 | 0.053 | 0.06 | ≤0.01 |
| Percentage of GS which is greater than 6 degrees of Slope | Formula | -0.41 | ≤0.001 | -0.43 | ≤0.001 | -0.32 | ≤0.001 | -0.4 | ≤0.001 | -0.42 | ≤0.001 | -0.44 | ≤0.001 | -0.32 | ≤0.001 | -0.42 | ≤0.001 |
| Pathway Length > 6 degrees per Area of GS | Formula | -0.16 | ≤0.001 | -0.17 | ≤0.001 | -0.22 | ≤0.001 | 0.01 | 0.784 | -0.17 | ≤0.001 | -0.18 | ≤0.001 | -0.19 | ≤0.001 | -0.02 | 0.345 |
| Bikeway Length > 6 degrees slope per Area of GS | Formula | -0.06 | ≤0.001 | -0.08 | ≤0.001 | -0.11 | ≤0.001 | -0.04 | 0.152 | -0.07 | ≤0.001 | -0.08 | ≤0.001 | -0.08 | ≤0.001 | -0.06 | ≤0.01 |
| **Amenities/Activities indices – original sourced** | | | | | | | | | | | | | | | | | |
| Number of public toilet facilities within and surrounding a green space (within 100m buffer) | w0 Sum | -0.05 | ≤0.001 | -0.06 | ≤0.001 | -0.03 | 0.209 | 0.13 | ≤0.001 | -0.06 | ≤0.001 | -0.07 | ≤0.001 | -0.01 | 0.752 | 0.15 | ≤0.001 |
| Number of cafes within and surrounding a green space (within 100m buffer) | w0 Sum | -0.12 | ≤0.001 | -0.11 | ≤0.001 | -0.16 | ≤0.001 | 0.05 | ≤0.05 | -0.13 | ≤0.001 | -0.12 | ≤0.001 | -0.17 | ≤0.001 | 0.06 | ≤0.05 |
| Number of hotels/bars within and surrounding a green space (within 100m buffer) | w0 Sum | -0.05 | ≤0.001 | -0.05 | ≤0.001 | -0.06 | ≤0.01 | 0.02 | 0.455 | -0.06 | ≤0.001 | -0.05 | ≤0.001 | -0.05 | ≤0.01 | 0.02 | 0.435 |
| Number of restaurants/takeaways within and surrounding a green space (within 100m buffer) | w0 Sum | -0.06 | ≤0.001 | -0.05 | ≤0.001 | -0.01 | 0.525 | -0.02 | 0.382 | -0.06 | ≤0.001 | -0.05 | ≤0.001 | 0.01 | 0.787 | 0 | 0.998 |
| Number of restaurants within and surrounding a green space (within 100m buffer) | w0 Sum | -0.11 | ≤0.001 | -0.1 | ≤0.001 | -0.09 | ≤0.001 | -0.06 | ≤0.01 | -0.11 | ≤0.001 | -0.1 | ≤0.001 | -0.09 | ≤0.001 | -0.06 | ≤0.05 |
| Number of takeaways within and surrounding a green space (within 100m buffer) | w0 Sum | 0 | 0.427 | 0.01 | ≤0.05 | 0.03 | 0.176 | 0.05 | ≤0.05 | 0 | 0.494 | 0.01 | 0.052 | 0.04 | ≤0.05 | 0.06 | ≤0.01 |
| Number of supermarkets/greengrocers within and surrounding a green space (within 100m buffer) | w0 Sum | 0.01 | 0.099 | 0.01 | ≤0.05 | 0.02 | 0.312 | 0.12 | ≤0.001 | 0.01 | ≤0.01 | 0.02 | ≤0.001 | 0.02 | 0.199 | 0.12 | ≤0.001 |
| Presence of parkrun event | Quality | -0.03 | ≤0.001 | -0.02 | ≤0.001 | -0.07 | ≤0.001 | -0.1 | ≤0.001 | -0.04 | ≤0.001 | -0.03 | ≤0.001 | -0.09 | ≤0.001 | -0.13 | ≤0.001 |
| **Beach/Coast – original sourced** | | | | | | | | | | | | | | | | | |
| Presence of coastline | Quality | -0.25 | ≤0.001 | -0.29 | ≤0.001 | -0.06 | ≤0.01 | -0.02 | 0.486 | -0.26 | ≤0.001 | -0.31 | ≤0.001 | -0.06 | ≤0.01 | -0.02 | 0.302 |
| Area of beach within and surrounding a green space (within 100m buffer) | w0 Sum | -0.16 | ≤0.001 | -0.18 | ≤0.001 | -0.11 | ≤0.001 | -0.17 | ≤0.001 | -0.17 | ≤0.001 | -0.19 | ≤0.001 | -0.11 | ≤0.001 | -0.17 | ≤0.001 |
| **Biodiversity indices – original sourced** | | | | | | | | | | | | | | | | | |
| Number of bird species/species habitat likely to occur within green space | Max | -0.29 | ≤0.001 | -0.34 | ≤0.001 | 0 | 0.827 | 0.17 | ≤0.001 | -0.29 | ≤0.001 | -0.34 | ≤0.001 | 0 | 0.889 | 0.2 | ≤0.001 |
| Number of fish species/species habitat likely to occur within green space | Max | -0.31 | ≤0.001 | -0.35 | ≤0.001 | -0.11 | ≤0.001 | 0.04 | 0.145 | -0.32 | ≤0.001 | -0.36 | ≤0.001 | -0.11 | ≤0.001 | 0.05 | ≤0.05 |
| Number of flora species/species habitat likely to occur within green space | Max | -0.27 | ≤0.001 | -0.35 | ≤0.001 | 0.06 | ≤0.01 | 0.32 | ≤0.001 | -0.26 | ≤0.001 | -0.35 | ≤0.001 | 0.06 | ≤0.01 | 0.32 | ≤0.001 |
| Number of frog species/species habitat likely to occur within green space | Max | -0.24 | ≤0.001 | -0.28 | ≤0.001 | 0.09 | ≤0.001 | -0.18 | ≤0.001 | -0.22 | ≤0.001 | -0.26 | ≤0.001 | 0.09 | ≤0.001 | -0.19 | ≤0.001 |
| Number of mammal species/species habitat likely to occur within green space | Max | -0.51 | ≤0.001 | -0.56 | ≤0.001 | -0.2 | ≤0.001 | -0.3 | ≤0.001 | -0.51 | ≤0.001 | -0.56 | ≤0.001 | -0.21 | ≤0.001 | -0.29 | ≤0.001 |
| Number of other animal species/species habitat likely to occur within green space | Max | -0.29 | ≤0.001 | -0.29 | ≤0.001 |  |  |  |  | -0.31 | ≤0.001 | -0.31 | ≤0.001 |  |  |  |  |
| Number of reptile species/species habitat likely to occur within green space | Max | -0.34 | ≤0.001 | -0.39 | ≤0.001 | -0.12 | ≤0.001 | -0.05 | ≤0.05 | -0.35 | ≤0.001 | -0.39 | ≤0.001 | -0.12 | ≤0.001 | -0.03 | 0.178 |
| Area of land with biodiversity value within green space | w0 Sum | -0.01 | ≤0.05 | -0.02 | ≤0.01 | 0.06 | ≤0.01 | 0.01 | 0.604 | -0.01 | ≤0.01 | -0.02 | ≤0.001 | 0.07 | ≤0.001 | 0.01 | 0.661 |
| Area of land with biodiversity value outside of green space within 100m | w0 Sum | -0.04 | ≤0.001 | -0.05 | ≤0.001 | 0.08 | ≤0.001 | 0 | 0.985 | -0.05 | ≤0.001 | -0.05 | ≤0.001 | 0.08 | ≤0.001 | 0 | 0.968 |
| **Biodiversity indices – calculated** | | | | | | | | | | | | | | | | | |
| Total number of species/species habitat likely to occur within green space | Formula | -0.48 | ≤0.001 | -0.54 | ≤0.001 | -0.06 | ≤0.01 | 0.17 | ≤0.001 | -0.48 | ≤0.001 | -0.54 | ≤0.001 | -0.07 | ≤0.001 | 0.19 | ≤0.001 |
| Shannon biodiversity index | Formula | -0.41 | ≤0.001 | -0.46 | ≤0.001 | -0.18 | ≤0.001 | -0.28 | ≤0.001 | -0.41 | ≤0.001 | -0.46 | ≤0.001 | -0.18 | ≤0.001 | -0.29 | ≤0.001 |
| Simpson biodiversity index (range 0 to 1=most diverse) | Formula | -0.3 | ≤0.001 | -0.34 | ≤0.001 | -0.1 | ≤0.001 | -0.38 | ≤0.001 | -0.3 | ≤0.001 | -0.33 | ≤0.001 | -0.11 | ≤0.001 | -0.4 | ≤0.001 |
| Percentage of land with biodiversity value within green space | Formula | -0.01 | ≤0.01 | -0.02 | ≤0.001 | 0.08 | ≤0.001 | 0.01 | 0.723 | -0.02 | ≤0.001 | -0.02 | ≤0.001 | 0.08 | ≤0.001 | 0 | 0.908 |
| Percentage of land with biodiversity value outside of green space within 100m | Formula | -0.04 | ≤0.001 | -0.05 | ≤0.001 | 0.1 | ≤0.001 | -0.01 | 0.771 | -0.05 | ≤0.001 | -0.06 | ≤0.001 | 0.1 | ≤0.001 | -0.01 | 0.719 |
| **Incivilities indices – original sourced** | | | | | | | | | | | | | | | | | |
| Density of alcohol-related assaults | Max | 0.06 | ≤0.001 | 0.07 | ≤0.001 | 0.07 | ≤0.01 | 0.14 | ≤0.001 | 0.06 | ≤0.001 | 0.07 | ≤0.001 | 0.08 | ≤0.001 | 0.16 | ≤0.001 |
| Density of malicious damage | Max | 0.26 | ≤0.001 | 0.26 | ≤0.001 | 0.23 | ≤0.001 | 0.35 | ≤0.001 | 0.26 | ≤0.001 | 0.27 | ≤0.001 | 0.25 | ≤0.001 | 0.34 | ≤0.001 |
| Density of non-domestic violence assaults | Max | 0.13 | ≤0.001 | 0.14 | ≤0.001 | 0.09 | ≤0.001 | 0.2 | ≤0.001 | 0.14 | ≤0.001 | 0.15 | ≤0.001 | 0.1 | ≤0.001 | 0.24 | ≤0.001 |
| Density of robberies | Max | 0.18 | ≤0.001 | 0.2 | ≤0.001 | -0.01 | 0.633 | 0.19 | ≤0.001 | 0.19 | ≤0.001 | 0.21 | ≤0.001 | -0.03 | 0.196 | 0.2 | ≤0.001 |
| Density of steal from persons | Max | 0.07 | ≤0.001 | 0.08 | ≤0.001 | -0.01 | 0.796 | 0.12 | ≤0.001 | 0.08 | ≤0.001 | 0.09 | ≤0.001 | -0.01 | 0.455 | 0.14 | ≤0.001 |
| **Incivilities indices – calculated** | | | | | | | | | | | | | | | | | |
| Mean level of incivilities | Formula | 0.25 | ≤0.001 | 0.26 | ≤0.001 | 0.2 | ≤0.001 | 0.38 | ≤0.001 | 0.26 | ≤0.001 | 0.27 | ≤0.001 | 0.21 | ≤0.001 | 0.37 | ≤0.001 |
| **Land cover indices – original sourced** | | | | | | | | | | | | | | | | | |
| Area of bare earth within green space | w0 Sum | 0.17 | ≤0.001 | 0.17 | ≤0.001 | 0.01 | 0.804 | 0.16 | ≤0.001 | 0.17 | ≤0.001 | 0.18 | ≤0.001 | 0.02 | 0.293 | 0.17 | ≤0.001 |
| Area of buildings within green space | w0 Sum | 0.01 | 0.341 | 0 | 0.419 | 0.04 | 0.099 | 0.19 | ≤0.001 | 0.01 | 0.191 | 0 | 0.689 | 0.03 | 0.074 | 0.2 | ≤0.001 |
| Area of built up area within green space | w0 Sum | 0 | 0.439 | -0.01 | ≤0.05 | -0.01 | 0.709 | 0.14 | ≤0.001 | 0 | 0.609 | -0.01 | 0.055 | 0 | 0.845 | 0.16 | ≤0.001 |
| Area of open grass within green space | w0 Sum | 0.16 | ≤0.001 | 0.15 | ≤0.001 | 0.1 | ≤0.001 | 0.27 | ≤0.001 | 0.16 | ≤0.001 | 0.14 | ≤0.001 | 0.11 | ≤0.001 | 0.27 | ≤0.001 |
| Area of other vegetation within green space | w0 Sum | 0.02 | ≤0.01 | 0 | 0.85 | -0.07 | ≤0.001 | 0.06 | ≤0.01 | 0.01 | ≤0.05 | 0 | 0.953 | -0.07 | ≤0.001 | 0.09 | ≤0.001 |
| Area of roads/paths within green space | w0 Sum | 0.05 | ≤0.001 | 0.02 | ≤0.01 | 0.02 | 0.264 | 0.19 | ≤0.001 | 0.06 | ≤0.001 | 0.02 | ≤0.001 | 0.03 | 0.095 | 0.22 | ≤0.001 |
| Area of shadow within green space | w0 Sum | -0.14 | ≤0.001 | -0.12 | ≤0.001 | -0.12 | ≤0.001 | -0.18 | ≤0.001 | -0.15 | ≤0.001 | -0.13 | ≤0.001 | -0.14 | ≤0.001 | -0.18 | ≤0.001 |
| Area of swimming pools within green space | w0 Sum | -0.11 | ≤0.001 | -0.1 | ≤0.001 | -0.17 | ≤0.001 | 0.08 | ≤0.01 | -0.12 | ≤0.001 | -0.11 | ≤0.001 | -0.17 | ≤0.001 | 0.08 | ≤0.001 |
| Area of tree canopy within green space | w0 Sum | -0.23 | ≤0.001 | -0.24 | ≤0.001 | -0.11 | ≤0.001 | -0.06 | ≤0.01 | -0.23 | ≤0.001 | -0.25 | ≤0.001 | -0.11 | ≤0.001 | -0.05 | ≤0.05 |
| Area of water within green space | w0 Sum | -0.11 | ≤0.001 | -0.14 | ≤0.001 | -0.03 | 0.167 | 0.01 | 0.829 | -0.11 | ≤0.001 | -0.15 | ≤0.001 | -0.01 | 0.524 | 0.03 | 0.212 |
| **Land cover indices - calculated** | | | | | | | | | | | | | | | | | |
| Percentage of bare earth within green space | Formula | 0.27 | ≤0.001 | 0.29 | ≤0.001 | 0.05 | ≤0.05 | 0.13 | ≤0.001 | 0.28 | ≤0.001 | 0.3 | ≤0.001 | 0.07 | ≤0.001 | 0.12 | ≤0.001 |
| Percentage of buildings within green space | Formula | 0.02 | ≤0.001 | 0.02 | ≤0.001 | 0.07 | ≤0.01 | 0.16 | ≤0.001 | 0.03 | ≤0.001 | 0.03 | ≤0.001 | 0.07 | ≤0.001 | 0.17 | ≤0.001 |
| Percentage of built up area within green space | Formula | 0.02 | ≤0.001 | 0.02 | ≤0.001 | 0.01 | 0.632 | 0.11 | ≤0.001 | 0.02 | ≤0.001 | 0.03 | ≤0.001 | 0.03 | 0.152 | 0.11 | ≤0.001 |
| Percentage of open grass within green space | Formula | 0.3 | ≤0.001 | 0.29 | ≤0.001 | 0.27 | ≤0.001 | 0.34 | ≤0.001 | 0.3 | ≤0.001 | 0.29 | ≤0.001 | 0.27 | ≤0.001 | 0.34 | ≤0.001 |
| Percentage of other vegetation within green space | Formula | 0.06 | ≤0.001 | 0.06 | ≤0.001 | -0.11 | ≤0.001 | -0.16 | ≤0.001 | 0.06 | ≤0.001 | 0.07 | ≤0.001 | -0.11 | ≤0.001 | -0.16 | ≤0.001 |
| Percentage of roads/paths within green space | Formula | 0.09 | ≤0.001 | 0.05 | ≤0.001 | 0.07 | ≤0.01 | 0.23 | ≤0.001 | 0.1 | ≤0.001 | 0.06 | ≤0.001 | 0.07 | ≤0.001 | 0.26 | ≤0.001 |
| Percentage of shadow within green space | Formula | -0.15 | ≤0.001 | -0.12 | ≤0.001 | -0.12 | ≤0.001 | -0.2 | ≤0.001 | -0.16 | ≤0.001 | -0.13 | ≤0.001 | -0.15 | ≤0.001 | -0.21 | ≤0.001 |
| Percentage of swimming pools within green space | Formula | -0.1 | ≤0.001 | -0.1 | ≤0.001 | -0.16 | ≤0.001 | 0.07 | ≤0.01 | -0.12 | ≤0.001 | -0.11 | ≤0.001 | -0.16 | ≤0.001 | 0.07 | ≤0.001 |
| Percentage of tree canopy within green space | Formula | -0.39 | ≤0.001 | -0.4 | ≤0.001 | -0.19 | ≤0.001 | -0.31 | ≤0.001 | -0.4 | ≤0.001 | -0.41 | ≤0.001 | -0.21 | ≤0.001 | -0.32 | ≤0.001 |
| Percentage of water within green space | Formula | -0.11 | ≤0.001 | -0.14 | ≤0.001 | -0.02 | 0.248 | 0 | 0.936 | -0.11 | ≤0.001 | -0.15 | ≤0.001 | -0.01 | 0.768 | 0.02 | 0.441 |
| **Land use indices – original sourced** | | | | | | | | | | | | | | | | | |
| Area of commercial land use surrounding green space (within 100m doughnut buffer) | w0 Sum | 0.04 | ≤0.001 | 0.05 | ≤0.001 | 0.03 | 0.109 | 0.19 | ≤0.001 | 0.05 | ≤0.001 | 0.06 | ≤0.001 | 0.05 | ≤0.01 | 0.2 | ≤0.001 |
| Area of education land use surrounding green space (within 100m doughnut buffer) | w0 Sum | 0.03 | ≤0.001 | 0.02 | ≤0.001 | 0.08 | ≤0.001 | 0.14 | ≤0.001 | 0.03 | ≤0.001 | 0.02 | ≤0.001 | 0.06 | ≤0.01 | 0.15 | ≤0.001 |
| Area of hospital/medical land use surrounding green space (within 100m doughnut buffer) | w0 Sum | -0.01 | 0.089 | 0 | 0.629 | -0.09 | ≤0.001 | 0 | 0.859 | -0.02 | ≤0.001 | -0.01 | ≤0.05 | -0.09 | ≤0.001 | 0.03 | 0.234 |
| Area of industrial land use surrounding green space (within 100m doughnut buffer) | w0 Sum | 0.08 | ≤0.001 | 0.07 | ≤0.001 | 0.11 | ≤0.001 | 0.14 | ≤0.001 | 0.09 | ≤0.001 | 0.09 | ≤0.001 | 0.12 | ≤0.001 | 0.14 | ≤0.001 |
| Area of other land use surrounding green space (within 100m doughnut buffer) | w0 Sum | 0.01 | 0.095 | 0 | 0.607 | 0.09 | ≤0.001 | -0.07 | ≤0.01 | 0.01 | 0.106 | 0 | 0.636 | 0.09 | ≤0.001 | -0.07 | ≤0.001 |
| Area of parkland land use surrounding green space (within 100m doughnut buffer) | w0 Sum | -0.06 | ≤0.001 | -0.06 | ≤0.001 | -0.04 | 0.051 | -0.02 | 0.533 | -0.07 | ≤0.001 | -0.07 | ≤0.001 | -0.03 | 0.174 | 0 | 0.899 |
| Area of primary production land use surrounding green space (within 100m doughnut buffer) | w0 Sum | 0.05 | ≤0.001 | 0.04 | ≤0.001 | 0.13 | ≤0.001 | -0.05 | ≤0.05 | 0.05 | ≤0.001 | 0.04 | ≤0.001 | 0.14 | ≤0.001 | -0.04 | 0.052 |
| Area of residential land use surrounding green space (within 100m doughnut buffer) | w0 Sum | -0.08 | ≤0.001 | -0.07 | ≤0.001 | -0.03 | 0.195 | 0.11 | ≤0.001 | -0.07 | ≤0.001 | -0.06 | ≤0.001 | -0.02 | 0.282 | 0.13 | ≤0.001 |
| Area of transport land use surrounding green space (within 100m doughnut buffer) | w0 Sum | 0.01 | ≤0.05 | 0.02 | ≤0.001 | 0 | 0.993 | 0.05 | 0.062 | 0.02 | ≤0.01 | 0.03 | ≤0.001 | 0 | 0.921 | 0.05 | ≤0.05 |
| Area of water land use surrounding green space (within 100m doughnut buffer) | w0 Sum | -0.01 | 0.166 | -0.01 | 0.181 | 0.09 | ≤0.001 |  |  | -0.01 | ≤0.01 | -0.01 | ≤0.01 | 0.09 | ≤0.001 |  |  |
| Area of tree canopy surrounding green space (within 100m doughnut buffer) | w0 Sum | -0.19 | ≤0.001 | -0.18 | ≤0.001 | -0.07 | ≤0.01 | 0.04 | 0.155 | -0.19 | ≤0.001 | -0.19 | ≤0.001 | -0.07 | ≤0.001 | 0.05 | ≤0.05 |
| Area of tree canopy ≤4m surrounding green space (within 100m doughnut buffer) | w0 Sum | -0.13 | ≤0.001 | -0.11 | ≤0.001 | -0.03 | 0.247 | 0.04 | 0.134 | -0.12 | ≤0.001 | -0.11 | ≤0.001 | -0.02 | 0.412 | 0.06 | ≤0.01 |
| Area of tree canopy >4m surrounding green space (within 100m doughnut buffer) | w0 Sum | -0.2 | ≤0.001 | -0.19 | ≤0.001 | -0.08 | ≤0.001 | 0.04 | 0.122 | -0.2 | ≤0.001 | -0.19 | ≤0.001 | -0.07 | ≤0.001 | 0.05 | ≤0.05 |
| **Land use indices – calculated** | | | | | | | | | | | | | | | | | |
| Percentage of commercial land use surrounding green space (within 100m doughnut buffer) | Formula | 0.06 | ≤0.001 | 0.06 | ≤0.001 | 0.04 | 0.094 | 0.19 | ≤0.001 | 0.07 | ≤0.001 | 0.07 | ≤0.001 | 0.06 | ≤0.01 | 0.2 | ≤0.001 |
| Percentage of education land use surrounding green space (within 100m doughnut buffer) | Formula | 0.04 | ≤0.001 | 0.04 | ≤0.001 | 0.08 | ≤0.001 | 0.13 | ≤0.001 | 0.04 | ≤0.001 | 0.04 | ≤0.001 | 0.07 | ≤0.001 | 0.14 | ≤0.001 |
| Percentage of hospital/medical land use surrounding green space (within 100m doughnut buffer) | Formula | -0.01 | 0.106 | 0 | 0.688 | -0.09 | ≤0.001 | 0 | 0.866 | -0.02 | ≤0.01 | -0.01 | ≤0.05 | -0.09 | ≤0.001 | 0.03 | 0.237 |
| Percentage of industrial land use surrounding green space (within 100m doughnut buffer) | Formula | 0.09 | ≤0.001 | 0.08 | ≤0.001 | 0.11 | ≤0.001 | 0.13 | ≤0.001 | 0.1 | ≤0.001 | 0.09 | ≤0.001 | 0.12 | ≤0.001 | 0.14 | ≤0.001 |
| Percentage of other land use surrounding green space (within 100m doughnut buffer) | Formula | 0.01 | 0.096 | 0 | 0.599 | 0.09 | ≤0.001 | -0.07 | ≤0.01 | 0.01 | 0.112 | 0 | 0.617 | 0.09 | ≤0.001 | -0.08 | ≤0.001 |
| Percentage of parkland land use surrounding green space (within 100m doughnut buffer) | Formula | -0.05 | ≤0.001 | -0.05 | ≤0.001 | -0.02 | 0.452 | -0.08 | ≤0.01 | -0.05 | ≤0.001 | -0.06 | ≤0.001 | 0 | 0.943 | -0.09 | ≤0.001 |
| Percentage of primary production land use surrounding green space (within 100m doughnut buffer) | Formula | 0.05 | ≤0.001 | 0.04 | ≤0.001 | 0.13 | ≤0.001 | -0.05 | ≤0.05 | 0.05 | ≤0.001 | 0.04 | ≤0.001 | 0.14 | ≤0.001 | -0.04 | 0.055 |
| Percentage of residential land use surrounding green space (within 100m doughnut buffer) | Formula | -0.02 | ≤0.001 | -0.01 | 0.267 | -0.06 | ≤0.01 | -0.03 | 0.19 | -0.02 | ≤0.001 | -0.01 | 0.174 | -0.09 | ≤0.001 | -0.03 | 0.126 |
| Percentage of transport land use surrounding green space (within 100m doughnut buffer) | Formula | 0.01 | ≤0.05 | 0.03 | ≤0.001 | 0 | 0.992 | 0.05 | 0.061 | 0.02 | ≤0.001 | 0.03 | ≤0.001 | 0 | 0.916 | 0.05 | ≤0.05 |
| Percentage of water land use surrounding green space (within 100m doughnut buffer) | Formula | -0.01 | 0.168 | -0.01 | 0.182 | 0.09 | ≤0.001 |  |  | -0.01 | ≤0.01 | -0.01 | ≤0.01 | 0.09 | ≤0.001 |  |  |
| Percentage of tree canopy to roads surrounding green space (within 100m doughnut buffer) | Formula | -0.43 | ≤0.001 | -0.44 | ≤0.001 | -0.11 | ≤0.001 | -0.28 | ≤0.001 | -0.44 | ≤0.001 | -0.45 | ≤0.001 | -0.12 | ≤0.001 | -0.3 | ≤0.001 |
| Percentage of tree canopy ≤4m to roads surrounding green space (within 100m doughnut buffer) | Formula | -0.19 | ≤0.001 | -0.18 | ≤0.001 | 0.01 | 0.751 | -0.33 | ≤0.001 | -0.2 | ≤0.001 | -0.19 | ≤0.001 | -0.01 | 0.629 | -0.34 | ≤0.001 |
| Percentage of tree canopy >4m to roads surrounding green space (within 100m doughnut buffer) | Formula | -0.42 | ≤0.001 | -0.43 | ≤0.001 | -0.12 | ≤0.001 | -0.24 | ≤0.001 | -0.43 | ≤0.001 | -0.44 | ≤0.001 | -0.13 | ≤0.001 | -0.26 | ≤0.001 |
| Percentage of tree canopy surrounding green space (within 100m doughnut buffer) | Formula | -0.35 | ≤0.001 | -0.34 | ≤0.001 | -0.1 | ≤0.001 | -0.17 | ≤0.001 | -0.35 | ≤0.001 | -0.35 | ≤0.001 | -0.12 | ≤0.001 | -0.19 | ≤0.001 |
| Percentage of tree canopy ≤4m surrounding green space (within 100m doughnut buffer) | Formula | -0.14 | ≤0.001 | -0.12 | ≤0.001 | 0.01 | 0.671 | -0.19 | ≤0.001 | -0.14 | ≤0.001 | -0.12 | ≤0.001 | -0.01 | 0.592 | -0.19 | ≤0.001 |
| Percentage of tree canopy >4m surrounding green space (within 100m doughnut buffer) | Formula | -0.35 | ≤0.001 | -0.35 | ≤0.001 | -0.11 | ≤0.001 | -0.15 | ≤0.001 | -0.36 | ≤0.001 | -0.36 | ≤0.001 | -0.13 | ≤0.001 | -0.16 | ≤0.001 |

Method of aggregating intersects over mesh blocks: Count=unweighted count | Sum=unweighted sum | w0 Sum = sum weighted by the ratio of the green space intersect with the buffer area to the green space area | w1 Mean = mean weighted by the ratio of the intersect area to the sum of all intersect areas | Max = maximum value over the intersects within the buffer | Quality = Value 0/1, aggregated to 1 if any intersects equal 1 (technically Quality = Max) | Formula = calculation of the index from other qualities that have already been aggregated as weighted sums or weighted means.

P-value signs: * ≤0.001, × ≤0.01, + ≤0.05

GS = green space

Table S8. Spearman's rank coefficient of correlation between SEIFA IRSD score (reversed) and aggregated over MB green space features with full p-values – 800m and 1600m buffers.

|  |  | **Buffer 800m** | | | | | | | | **Buffer 1600m** | | | | | | | |
| --- | --- | --- | --- | --- | --- | --- | --- | --- | --- | --- | --- | --- | --- | --- | --- | --- | --- |
|  |  | **Full set** | | **Sydney** | | **Newcastle** | | **Wollongong** | | **Full set** | | **Sydney** | | **Newcastle** | | **Wollongong** | |
| **Index description** | **Method** | **Rho** | **P** | **Rho** | **P** | **Rho** | **P** | **Rho** | **P** | **Rho** | **P** | **Rho** | **P** | **Rho** | **P** | **Rho** | **P** |
| **General indices** | | | | | | | | | | | | | | | | | |
| N green spaces in MB | Count | -0.09 | ≤0.001 | -0.08 | ≤0.001 | 0.03 | 0.102 | 0.18 | ≤0.001 | -0.03 | ≤0.001 | -0.01 | ≤0.05 | 0.07 | ≤0.001 | 0.26 | ≤0.001 |
| Intersect of buffer with green space | Sum | 0.01 | 0.099 | -0.01 | 0.196 | -0.03 | 0.106 | 0.23 | ≤0.001 | 0.09 | ≤0.001 | 0.08 | ≤0.001 | -0.06 | ≤0.001 | 0.34 | ≤0.001 |
| Green space area (A) | Sum | -0.05 | ≤0.001 | -0.08 | ≤0.001 | -0.02 | 0.316 | 0.12 | ≤0.001 | -0.04 | ≤0.001 | -0.07 | ≤0.001 | -0.02 | 0.181 | 0.02 | 0.35 |
| GS perimeter (P) | w0 Sum | -0.04 | ≤0.001 | -0.04 | ≤0.001 | -0.01 | 0.551 | 0.23 | ≤0.001 | 0.02 | ≤0.001 | 0.04 | ≤0.001 | -0.04 | ≤0.05 | 0.32 | ≤0.001 |
| **Access indices – original sourced** | | | | | | | | | | | | | | | | | |
| Length of pathways within green space | w0 Sum | -0.01 | ≤0.05 | 0 | 0.499 | -0.11 | ≤0.001 | 0.2 | ≤0.001 | 0 | 0.319 | 0 | 0.34 | -0.13 | ≤0.001 | 0.28 | ≤0.001 |
| Length of bikeways within green space | w0 Sum | 0.04 | ≤0.001 | 0.02 | ≤0.001 | 0.02 | 0.183 | 0.2 | ≤0.001 | 0.1 | ≤0.001 | 0.09 | ≤0.001 | 0.04 | ≤0.05 | 0.29 | ≤0.001 |
| Length of pathways covered by trees within green space | w0 Sum | -0.08 | ≤0.001 | -0.07 | ≤0.001 | -0.16 | ≤0.001 | 0.11 | ≤0.001 | -0.11 | ≤0.001 | -0.09 | ≤0.001 | -0.12 | ≤0.001 | 0.11 | ≤0.001 |
| Length of bikeways covered by trees within green space | w0 Sum | -0.03 | ≤0.001 | -0.03 | ≤0.001 | 0 | 0.946 | 0.14 | ≤0.001 | -0.01 | ≤0.01 | -0.01 | 0.07 | 0.09 | ≤0.001 | 0.15 | ≤0.001 |
| Length of pathways greater than 6 degrees gradient within green space | w0 Sum | -0.21 | ≤0.001 | -0.22 | ≤0.001 | -0.18 | ≤0.001 | 0.03 | 0.175 | -0.24 | ≤0.001 | -0.25 | ≤0.001 | -0.21 | ≤0.001 | 0.08 | ≤0.001 |
| Length of bikeways greater than 6 degrees gradient within green space | w0 Sum | -0.09 | ≤0.001 | -0.1 | ≤0.001 | -0.08 | ≤0.001 | 0.04 | 0.066 | -0.07 | ≤0.001 | -0.08 | ≤0.001 | -0.08 | ≤0.001 | 0.11 | ≤0.001 |
| Number of Railway Stations within and surrounding a green space (within 100m buffer) | w0 Sum | 0.04 | ≤0.001 | 0.05 | ≤0.001 | 0.02 | 0.277 | -0.05 | ≤0.05 | 0.06 | ≤0.001 | 0.09 | ≤0.001 | 0.01 | 0.384 | 0.01 | 0.674 |
| Number of Bus Stops within and surrounding a green space (within 100m buffer) | w0 Sum | 0.02 | ≤0.001 | 0.03 | ≤0.001 | -0.03 | 0.091 | 0.27 | ≤0.001 | 0.08 | ≤0.001 | 0.11 | ≤0.001 | -0.04 | ≤0.05 | 0.34 | ≤0.001 |
| Number of ferry wharves within and surrounding a green space (within 100m buffer) | w0 Sum | -0.15 | ≤0.001 | -0.15 | ≤0.001 | -0.01 | 0.631 |  |  | -0.19 | ≤0.001 | -0.19 | ≤0.001 | -0.09 | ≤0.001 |  |  |
| Area of GS which is greater than 6 degrees of Slope | w0 Sum | -0.3 | ≤0.001 | -0.33 | ≤0.001 | -0.22 | ≤0.001 | -0.08 | ≤0.001 | -0.29 | ≤0.001 | -0.32 | ≤0.001 | -0.24 | ≤0.001 | -0.05 | ≤0.01 |
| Length of Main Roads within 100 metres of the Green Space | w0 Sum | -0.03 | ≤0.001 | -0.03 | ≤0.001 | 0.14 | ≤0.001 | 0.29 | ≤0.001 | 0 | 0.83 | 0 | 0.585 | 0.18 | ≤0.001 | 0.43 | ≤0.001 |
| **Access indices - calculated** | | | | | | | | | | | | | | | | | |
| Ratio of Area(A) to Perimeter (P) | w1 Mean | 0.04 | ≤0.001 | 0.01 | ≤0.05 | -0.01 | 0.413 | 0.06 | ≤0.01 | 0.05 | ≤0.001 | 0.03 | ≤0.001 | -0.07 | ≤0.001 | 0.03 | 0.081 |
| Ratio of Perimeter (P) to perimeter standardised by area (A) | w1 Mean | -0.19 | ≤0.001 | -0.18 | ≤0.001 | -0.14 | ≤0.001 | -0.11 | ≤0.001 | -0.22 | ≤0.001 | -0.2 | ≤0.001 | -0.09 | ≤0.001 | -0.15 | ≤0.001 |
| Total number of transport stops/stations/wharves within and surrounding a green space (within 100m buffer) | Formula | 0.01 | ≤0.01 | 0.02 | ≤0.001 | -0.03 | 0.102 | 0.27 | ≤0.001 | 0.08 | ≤0.001 | 0.1 | ≤0.001 | -0.05 | ≤0.01 | 0.34 | ≤0.001 |
| Pathway Length per Area of Green Space | Formula | 0 | 0.579 | 0.02 | ≤0.001 | -0.12 | ≤0.001 | 0.13 | ≤0.001 | -0.04 | ≤0.001 | -0.02 | ≤0.001 | -0.14 | ≤0.001 | 0.16 | ≤0.001 |
| Bikeway Length per Area of Green Space | Formula | 0.05 | ≤0.001 | 0.04 | ≤0.001 | 0.04 | ≤0.05 | 0.1 | ≤0.001 | 0.09 | ≤0.001 | 0.08 | ≤0.001 | 0.07 | ≤0.001 | 0.11 | ≤0.001 |
| Trees over Pathway per Area of Green Space | Formula | -0.09 | ≤0.001 | -0.08 | ≤0.001 | -0.17 | ≤0.001 | 0.06 | ≤0.01 | -0.15 | ≤0.001 | -0.13 | ≤0.001 | -0.15 | ≤0.001 | 0.03 | 0.118 |
| Trees over Bikeway per Area of Green Space | Formula | -0.04 | ≤0.001 | -0.04 | ≤0.001 | 0 | 0.935 | 0.07 | ≤0.001 | -0.06 | ≤0.001 | -0.04 | ≤0.001 | 0.08 | ≤0.001 | 0.04 | ≤0.05 |
| Percentage of GS which is greater than 6 degrees of Slope | Formula | -0.46 | ≤0.001 | -0.49 | ≤0.001 | -0.34 | ≤0.001 | -0.4 | ≤0.001 | -0.49 | ≤0.001 | -0.52 | ≤0.001 | -0.34 | ≤0.001 | -0.44 | ≤0.001 |
| Pathway Length > 6 degrees per Area of GS | Formula | -0.23 | ≤0.001 | -0.24 | ≤0.001 | -0.2 | ≤0.001 | -0.02 | 0.257 | -0.3 | ≤0.001 | -0.31 | ≤0.001 | -0.25 | ≤0.001 | -0.03 | 0.109 |
| Bikeway Length > 6 degrees slope per Area of GS | Formula | -0.09 | ≤0.001 | -0.1 | ≤0.001 | -0.09 | ≤0.001 | -0.03 | 0.084 | -0.1 | ≤0.001 | -0.1 | ≤0.001 | -0.09 | ≤0.001 | -0.04 | ≤0.05 |
| **Amenities/Activities indices – original sourced** | | | | | | | | | | | | | | | | | |
| Number of public toilet facilities within and surrounding a green space (within 100m buffer) | w0 Sum | -0.05 | ≤0.001 | -0.06 | ≤0.001 | 0.03 | 0.087 | 0.25 | ≤0.001 | -0.07 | ≤0.001 | -0.08 | ≤0.001 | 0.06 | ≤0.001 | 0.29 | ≤0.001 |
| Number of cafes within and surrounding a green space (within 100m buffer) | w0 Sum | -0.15 | ≤0.001 | -0.14 | ≤0.001 | -0.15 | ≤0.001 | 0.12 | ≤0.001 | -0.18 | ≤0.001 | -0.17 | ≤0.001 | -0.14 | ≤0.001 | 0.16 | ≤0.001 |
| Number of hotels/bars within and surrounding a green space (within 100m buffer) | w0 Sum | -0.1 | ≤0.001 | -0.09 | ≤0.001 | -0.06 | ≤0.001 | 0.04 | ≤0.05 | -0.12 | ≤0.001 | -0.11 | ≤0.001 | -0.08 | ≤0.001 | 0.11 | ≤0.001 |
| Number of restaurants/takeaways within and surrounding a green space (within 100m buffer) | w0 Sum | -0.03 | ≤0.001 | -0.02 | ≤0.001 | 0.07 | ≤0.001 | 0.06 | ≤0.001 | -0.05 | ≤0.001 | -0.04 | ≤0.001 | 0.04 | ≤0.05 | 0.16 | ≤0.001 |
| Number of restaurants within and surrounding a green space (within 100m buffer) | w0 Sum | -0.1 | ≤0.001 | -0.09 | ≤0.001 | -0.05 | ≤0.01 | 0.01 | 0.508 | -0.12 | ≤0.001 | -0.11 | ≤0.001 | -0.05 | ≤0.01 | 0.11 | ≤0.001 |
| Number of takeaways within and surrounding a green space (within 100m buffer) | w0 Sum | 0.03 | ≤0.001 | 0.04 | ≤0.001 | 0.11 | ≤0.001 | 0.09 | ≤0.001 | 0.03 | ≤0.001 | 0.04 | ≤0.001 | 0.12 | ≤0.001 | 0.18 | ≤0.001 |
| Number of supermarkets/greengrocers within and surrounding a green space (within 100m buffer) | w0 Sum | 0.03 | ≤0.001 | 0.04 | ≤0.001 | 0.05 | ≤0.01 | 0.11 | ≤0.001 | 0.04 | ≤0.001 | 0.06 | ≤0.001 | 0 | 0.905 | 0.13 | ≤0.001 |
| Presence of parkrun event | Quality | -0.07 | ≤0.001 | -0.06 | ≤0.001 | -0.13 | ≤0.001 | -0.15 | ≤0.001 | -0.11 | ≤0.001 | -0.1 | ≤0.001 | -0.21 | ≤0.001 | -0.16 | ≤0.001 |
| **Beach/Coast – original sourced** | | | | | | | | | | | | | | | | | |
| Presence of coastline | Quality | -0.31 | ≤0.001 | -0.37 | ≤0.001 | -0.1 | ≤0.001 | 0.11 | ≤0.001 | -0.35 | ≤0.001 | -0.41 | ≤0.001 | -0.16 | ≤0.001 | 0.21 | ≤0.001 |
| Area of beach within and surrounding a green space (within 100m buffer) | w0 Sum | -0.23 | ≤0.001 | -0.27 | ≤0.001 | -0.12 | ≤0.001 | -0.1 | ≤0.001 | -0.31 | ≤0.001 | -0.35 | ≤0.001 | -0.17 | ≤0.001 | -0.05 | ≤0.05 |
| **Biodiversity indices – original sourced** | | | | | | | | | | | | | | | | | |
| Number of bird species/species habitat likely to occur within green space | Max | -0.28 | ≤0.001 | -0.34 | ≤0.001 | 0.05 | ≤0.01 | 0.23 | ≤0.001 | -0.26 | ≤0.001 | -0.32 | ≤0.001 | 0.1 | ≤0.001 | 0.23 | ≤0.001 |
| Number of fish species/species habitat likely to occur within green space | Max | -0.34 | ≤0.001 | -0.38 | ≤0.001 | -0.11 | ≤0.001 | 0.12 | ≤0.001 | -0.33 | ≤0.001 | -0.37 | ≤0.001 | -0.14 | ≤0.001 | 0.12 | ≤0.001 |
| Number of flora species/species habitat likely to occur within green space | Max | -0.25 | ≤0.001 | -0.33 | ≤0.001 | 0.12 | ≤0.001 | 0.32 | ≤0.001 | -0.22 | ≤0.001 | -0.3 | ≤0.001 | 0.15 | ≤0.001 | 0.2 | ≤0.001 |
| Number of frog species/species habitat likely to occur within green space | Max | -0.18 | ≤0.001 | -0.21 | ≤0.001 | 0.09 | ≤0.001 | -0.27 | ≤0.001 | -0.13 | ≤0.001 | -0.17 | ≤0.001 | 0.11 | ≤0.001 | -0.35 | ≤0.001 |
| Number of mammal species/species habitat likely to occur within green space | Max | -0.49 | ≤0.001 | -0.55 | ≤0.001 | -0.22 | ≤0.001 | -0.25 | ≤0.001 | -0.47 | ≤0.001 | -0.53 | ≤0.001 | -0.21 | ≤0.001 | -0.23 | ≤0.001 |
| Number of other animal species/species habitat likely to occur within green space | Max | -0.33 | ≤0.001 | -0.33 | ≤0.001 |  |  |  |  | -0.34 | ≤0.001 | -0.34 | ≤0.001 |  |  |  |  |
| Number of reptile species/species habitat likely to occur within green space | Max | -0.35 | ≤0.001 | -0.41 | ≤0.001 | -0.12 | ≤0.001 | 0.07 | ≤0.001 | -0.33 | ≤0.001 | -0.39 | ≤0.001 | -0.15 | ≤0.001 | 0.14 | ≤0.001 |
| Area of land with biodiversity value within green space | w0 Sum | 0.01 | ≤0.01 | 0.01 | ≤0.05 | 0.12 | ≤0.001 | 0.06 | ≤0.01 | 0.09 | ≤0.001 | 0.11 | ≤0.001 | 0.14 | ≤0.001 | 0.1 | ≤0.001 |
| Area of land with biodiversity value outside of green space within 100m | w0 Sum | -0.04 | ≤0.001 | -0.05 | ≤0.001 | 0.14 | ≤0.001 | 0.03 | 0.086 | 0.02 | ≤0.001 | 0.03 | ≤0.001 | 0.17 | ≤0.001 | 0.07 | ≤0.001 |
| **Biodiversity indices – calculated** | | | | | | | | | | | | | | | | | |
| Total number of species/species habitat likely to occur within green space | Formula | -0.45 | ≤0.001 | -0.52 | ≤0.001 | -0.03 | 0.053 | 0.22 | ≤0.001 | -0.39 | ≤0.001 | -0.45 | ≤0.001 | 0 | 0.881 | 0.18 | ≤0.001 |
| Shannon biodiversity index | Formula | -0.41 | ≤0.001 | -0.46 | ≤0.001 | -0.21 | ≤0.001 | -0.22 | ≤0.001 | -0.39 | ≤0.001 | -0.44 | ≤0.001 | -0.23 | ≤0.001 | -0.28 | ≤0.001 |
| Simpson biodiversity index (range 0 to 1=most diverse) | Formula | -0.3 | ≤0.001 | -0.33 | ≤0.001 | -0.18 | ≤0.001 | -0.42 | ≤0.001 | -0.28 | ≤0.001 | -0.3 | ≤0.001 | -0.23 | ≤0.001 | -0.49 | ≤0.001 |
| Percentage of land with biodiversity value within green space | Formula | -0.01 | 0.262 | -0.01 | 0.094 | 0.14 | ≤0.001 | 0.04 | ≤0.05 | 0.05 | ≤0.001 | 0.05 | ≤0.001 | 0.14 | ≤0.001 | 0.04 | ≤0.05 |
| Percentage of land with biodiversity value outside of green space within 100m | Formula | -0.06 | ≤0.001 | -0.07 | ≤0.001 | 0.15 | ≤0.001 | 0.01 | 0.785 | -0.02 | ≤0.001 | -0.03 | ≤0.001 | 0.16 | ≤0.001 | 0.01 | 0.513 |
| **Incivilities indices – original sourced** | | | | | | | | | | | | | | | | | |
| Density of alcohol-related assaults | Max | 0.09 | ≤0.001 | 0.1 | ≤0.001 | 0.14 | ≤0.001 | 0.2 | ≤0.001 | 0.13 | ≤0.001 | 0.14 | ≤0.001 | 0.17 | ≤0.001 | 0.16 | ≤0.001 |
| Density of malicious damage | Max | 0.3 | ≤0.001 | 0.31 | ≤0.001 | 0.24 | ≤0.001 | 0.36 | ≤0.001 | 0.33 | ≤0.001 | 0.35 | ≤0.001 | 0.2 | ≤0.001 | 0.35 | ≤0.001 |
| Density of non-domestic violence assaults | Max | 0.19 | ≤0.001 | 0.2 | ≤0.001 | 0.13 | ≤0.001 | 0.32 | ≤0.001 | 0.28 | ≤0.001 | 0.3 | ≤0.001 | 0.16 | ≤0.001 | 0.39 | ≤0.001 |
| Density of robberies | Max | 0.25 | ≤0.001 | 0.28 | ≤0.001 | -0.01 | 0.521 | 0.27 | ≤0.001 | 0.34 | ≤0.001 | 0.38 | ≤0.001 | -0.04 | ≤0.05 | 0.25 | ≤0.001 |
| Density of steal from persons | Max | 0.11 | ≤0.001 | 0.12 | ≤0.001 | -0.01 | 0.473 | 0.19 | ≤0.001 | 0.16 | ≤0.001 | 0.18 | ≤0.001 | -0.05 | ≤0.01 | 0.27 | ≤0.001 |
| **Incivilities indices – calculated** | | | | | | | | | | | | | | | | | |
| Mean level of incivilities | Formula | 0.29 | ≤0.001 | 0.31 | ≤0.001 | 0.19 | ≤0.001 | 0.4 | ≤0.001 | 0.3 | ≤0.001 | 0.32 | ≤0.001 | 0.13 | ≤0.001 | 0.39 | ≤0.001 |
| **Land cover indices – original sourced** | | | | | | | | | | | | | | | | | |
| Area of bare earth within green space | w0 Sum | 0.21 | ≤0.001 | 0.23 | ≤0.001 | 0.05 | ≤0.01 | 0.24 | ≤0.001 | 0.29 | ≤0.001 | 0.32 | ≤0.001 | 0.05 | ≤0.01 | 0.27 | ≤0.001 |
| Area of buildings within green space | w0 Sum | 0.04 | ≤0.001 | 0.04 | ≤0.001 | 0.05 | ≤0.01 | 0.31 | ≤0.001 | 0.11 | ≤0.001 | 0.11 | ≤0.001 | 0.07 | ≤0.001 | 0.34 | ≤0.001 |
| Area of built up area within green space | w0 Sum | 0.02 | ≤0.001 | 0.01 | ≤0.05 | 0.06 | ≤0.001 | 0.23 | ≤0.001 | 0.04 | ≤0.001 | 0.03 | ≤0.001 | 0.05 | ≤0.01 | 0.3 | ≤0.001 |
| Area of open grass within green space | w0 Sum | 0.19 | ≤0.001 | 0.18 | ≤0.001 | 0.13 | ≤0.001 | 0.36 | ≤0.001 | 0.3 | ≤0.001 | 0.29 | ≤0.001 | 0.17 | ≤0.001 | 0.45 | ≤0.001 |
| Area of other vegetation within green space | w0 Sum | 0.05 | ≤0.001 | 0.04 | ≤0.001 | -0.06 | ≤0.001 | 0.17 | ≤0.001 | 0.1 | ≤0.001 | 0.09 | ≤0.001 | -0.06 | ≤0.01 | 0.25 | ≤0.001 |
| Area of roads/paths within green space | w0 Sum | 0.11 | ≤0.001 | 0.07 | ≤0.001 | 0.05 | ≤0.01 | 0.3 | ≤0.001 | 0.19 | ≤0.001 | 0.16 | ≤0.001 | 0.03 | ≤0.05 | 0.41 | ≤0.001 |
| Area of shadow within green space | w0 Sum | -0.17 | ≤0.001 | -0.13 | ≤0.001 | -0.15 | ≤0.001 | -0.15 | ≤0.001 | -0.16 | ≤0.001 | -0.12 | ≤0.001 | -0.13 | ≤0.001 | -0.12 | ≤0.001 |
| Area of swimming pools within green space | w0 Sum | -0.11 | ≤0.001 | -0.09 | ≤0.001 | -0.15 | ≤0.001 | 0.11 | ≤0.001 | 0.02 | ≤0.01 | 0.04 | ≤0.001 | -0.06 | ≤0.001 | 0.15 | ≤0.001 |
| Area of tree canopy within green space | w0 Sum | -0.24 | ≤0.001 | -0.25 | ≤0.001 | -0.13 | ≤0.001 | -0.02 | 0.263 | -0.24 | ≤0.001 | -0.25 | ≤0.001 | -0.15 | ≤0.001 | -0.05 | ≤0.05 |
| Area of water within green space | w0 Sum | -0.12 | ≤0.001 | -0.17 | ≤0.001 | 0.03 | 0.079 | 0.15 | ≤0.001 | -0.09 | ≤0.001 | -0.13 | ≤0.001 | 0.1 | ≤0.001 | 0.23 | ≤0.001 |
| **Land cover indices - calculated** | | | | | | | | | | | | | | | | | |
| Percentage of bare earth within green space | Formula | 0.3 | ≤0.001 | 0.33 | ≤0.001 | 0.1 | ≤0.001 | 0.14 | ≤0.001 | 0.33 | ≤0.001 | 0.37 | ≤0.001 | 0.09 | ≤0.001 | 0.12 | ≤0.001 |
| Percentage of buildings within green space | Formula | 0.06 | ≤0.001 | 0.06 | ≤0.001 | 0.07 | ≤0.001 | 0.21 | ≤0.001 | 0.08 | ≤0.001 | 0.09 | ≤0.001 | 0.09 | ≤0.001 | 0.22 | ≤0.001 |
| Percentage of built up area within green space | Formula | 0.03 | ≤0.001 | 0.03 | ≤0.001 | 0.08 | ≤0.001 | 0.13 | ≤0.001 | -0.01 | 0.14 | -0.01 | 0.082 | 0.06 | ≤0.001 | 0.14 | ≤0.001 |
| Percentage of open grass within green space | Formula | 0.32 | ≤0.001 | 0.31 | ≤0.001 | 0.27 | ≤0.001 | 0.34 | ≤0.001 | 0.38 | ≤0.001 | 0.38 | ≤0.001 | 0.29 | ≤0.001 | 0.44 | ≤0.001 |
| Percentage of other vegetation within green space | Formula | 0.08 | ≤0.001 | 0.08 | ≤0.001 | -0.11 | ≤0.001 | -0.15 | ≤0.001 | 0.03 | ≤0.001 | 0.03 | ≤0.001 | -0.1 | ≤0.001 | -0.18 | ≤0.001 |
| Percentage of roads/paths within green space | Formula | 0.14 | ≤0.001 | 0.1 | ≤0.001 | 0.1 | ≤0.001 | 0.26 | ≤0.001 | 0.19 | ≤0.001 | 0.16 | ≤0.001 | 0.06 | ≤0.001 | 0.26 | ≤0.001 |
| Percentage of shadow within green space | Formula | -0.18 | ≤0.001 | -0.15 | ≤0.001 | -0.16 | ≤0.001 | -0.23 | ≤0.001 | -0.19 | ≤0.001 | -0.16 | ≤0.001 | -0.14 | ≤0.001 | -0.24 | ≤0.001 |
| Percentage of swimming pools within green space | Formula | -0.11 | ≤0.001 | -0.1 | ≤0.001 | -0.14 | ≤0.001 | 0.1 | ≤0.001 | -0.01 | 0.106 | 0.01 | ≤0.01 | -0.05 | ≤0.01 | 0.13 | ≤0.001 |
| Percentage of tree canopy within green space | Formula | -0.42 | ≤0.001 | -0.43 | ≤0.001 | -0.23 | ≤0.001 | -0.36 | ≤0.001 | -0.46 | ≤0.001 | -0.47 | ≤0.001 | -0.23 | ≤0.001 | -0.44 | ≤0.001 |
| Percentage of water within green space | Formula | -0.13 | ≤0.001 | -0.17 | ≤0.001 | 0.04 | ≤0.05 | 0.12 | ≤0.001 | -0.11 | ≤0.001 | -0.16 | ≤0.001 | 0.12 | ≤0.001 | 0.17 | ≤0.001 |
| **Land use indices – original sourced** | | | | | | | | | | | | | | | | | |
| Area of commercial land use surrounding green space (within 100m doughnut buffer) | w0 Sum | 0.09 | ≤0.001 | 0.1 | ≤0.001 | 0.12 | ≤0.001 | 0.27 | ≤0.001 | 0.14 | ≤0.001 | 0.16 | ≤0.001 | 0.12 | ≤0.001 | 0.33 | ≤0.001 |
| Area of education land use surrounding green space (within 100m doughnut buffer) | w0 Sum | 0.07 | ≤0.001 | 0.07 | ≤0.001 | 0.08 | ≤0.001 | 0.23 | ≤0.001 | 0.12 | ≤0.001 | 0.13 | ≤0.001 | 0.13 | ≤0.001 | 0.21 | ≤0.001 |
| Area of hospital/medical land use surrounding green space (within 100m doughnut buffer) | w0 Sum | -0.02 | ≤0.001 | -0.01 | 0.083 | -0.06 | ≤0.001 | 0.05 | ≤0.05 | 0.01 | ≤0.01 | 0.03 | ≤0.001 | -0.03 | 0.067 | -0.03 | 0.12 |
| Area of industrial land use surrounding green space (within 100m doughnut buffer) | w0 Sum | 0.15 | ≤0.001 | 0.14 | ≤0.001 | 0.22 | ≤0.001 | 0.2 | ≤0.001 | 0.27 | ≤0.001 | 0.27 | ≤0.001 | 0.31 | ≤0.001 | 0.28 | ≤0.001 |
| Area of other land use surrounding green space (within 100m doughnut buffer) | w0 Sum | 0.02 | ≤0.001 | 0 | 0.791 | 0.1 | ≤0.001 | -0.05 | ≤0.05 | 0.04 | ≤0.001 | 0.02 | ≤0.001 | 0.17 | ≤0.001 | -0.15 | ≤0.001 |
| Area of parkland land use surrounding green space (within 100m doughnut buffer) | w0 Sum | -0.08 | ≤0.001 | -0.08 | ≤0.001 | 0.02 | 0.27 | 0.11 | ≤0.001 | -0.02 | ≤0.001 | -0.01 | 0.058 | 0.05 | ≤0.01 | 0.22 | ≤0.001 |
| Area of primary production land use surrounding green space (within 100m doughnut buffer) | w0 Sum | 0.08 | ≤0.001 | 0.06 | ≤0.001 | 0.17 | ≤0.001 | 0.05 | ≤0.05 | 0.12 | ≤0.001 | 0.1 | ≤0.001 | 0.2 | ≤0.001 | 0.14 | ≤0.001 |
| Area of residential land use surrounding green space (within 100m doughnut buffer) | w0 Sum | -0.04 | ≤0.001 | -0.02 | ≤0.001 | -0.01 | 0.592 | 0.21 | ≤0.001 | 0.03 | ≤0.001 | 0.06 | ≤0.001 | -0.04 | ≤0.05 | 0.26 | ≤0.001 |
| Area of transport land use surrounding green space (within 100m doughnut buffer) | w0 Sum | 0.03 | ≤0.001 | 0.05 | ≤0.001 | 0 | 0.823 | 0.08 | ≤0.001 | 0.02 | ≤0.001 | 0.05 | ≤0.001 | 0.02 | 0.266 | 0.07 | ≤0.001 |
| Area of water land use surrounding green space (within 100m doughnut buffer) | w0 Sum | -0.03 | ≤0.001 | -0.03 | ≤0.001 | 0.12 | ≤0.001 |  |  | -0.04 | ≤0.001 | -0.04 | ≤0.001 | 0.15 | ≤0.001 |  |  |
| Area of tree canopy surrounding green space (within 100m doughnut buffer) | w0 Sum | -0.22 | ≤0.001 | -0.21 | ≤0.001 | -0.06 | ≤0.001 | 0.09 | ≤0.001 | -0.23 | ≤0.001 | -0.22 | ≤0.001 | -0.07 | ≤0.001 | 0.09 | ≤0.001 |
| Area of tree canopy ≤4m surrounding green space (within 100m doughnut buffer) | w0 Sum | -0.11 | ≤0.001 | -0.09 | ≤0.001 | 0 | 0.947 | 0.12 | ≤0.001 | -0.08 | ≤0.001 | -0.06 | ≤0.001 | 0.07 | ≤0.001 | 0.11 | ≤0.001 |
| Area of tree canopy >4m surrounding green space (within 100m doughnut buffer) | w0 Sum | -0.23 | ≤0.001 | -0.22 | ≤0.001 | -0.07 | ≤0.001 | 0.09 | ≤0.001 | -0.25 | ≤0.001 | -0.24 | ≤0.001 | -0.08 | ≤0.001 | 0.08 | ≤0.001 |
| **Land use indices – calculated** | | | | | | | | | | | | | | | | | |
| Percentage of commercial land use surrounding green space (within 100m doughnut buffer) | Formula | 0.12 | ≤0.001 | 0.12 | ≤0.001 | 0.12 | ≤0.001 | 0.25 | ≤0.001 | 0.17 | ≤0.001 | 0.17 | ≤0.001 | 0.12 | ≤0.001 | 0.29 | ≤0.001 |
| Percentage of education land use surrounding green space (within 100m doughnut buffer) | Formula | 0.09 | ≤0.001 | 0.09 | ≤0.001 | 0.1 | ≤0.001 | 0.18 | ≤0.001 | 0.15 | ≤0.001 | 0.14 | ≤0.001 | 0.15 | ≤0.001 | 0.11 | ≤0.001 |
| Percentage of hospital/medical land use surrounding green space (within 100m doughnut buffer) | Formula | -0.01 | ≤0.01 | -0.01 | 0.122 | -0.06 | ≤0.001 | 0.05 | ≤0.05 | 0.01 | ≤0.01 | 0.03 | ≤0.001 | -0.03 | ≤0.05 | -0.04 | ≤0.05 |
| Percentage of industrial land use surrounding green space (within 100m doughnut buffer) | Formula | 0.16 | ≤0.001 | 0.15 | ≤0.001 | 0.21 | ≤0.001 | 0.19 | ≤0.001 | 0.28 | ≤0.001 | 0.28 | ≤0.001 | 0.3 | ≤0.001 | 0.23 | ≤0.001 |
| Percentage of other land use surrounding green space (within 100m doughnut buffer) | Formula | 0.02 | ≤0.001 | 0 | 0.853 | 0.1 | ≤0.001 | -0.06 | ≤0.01 | 0.04 | ≤0.001 | 0.02 | ≤0.001 | 0.16 | ≤0.001 | -0.18 | ≤0.001 |
| Percentage of parkland land use surrounding green space (within 100m doughnut buffer) | Formula | -0.08 | ≤0.001 | -0.1 | ≤0.001 | 0.06 | ≤0.01 | -0.05 | ≤0.01 | -0.07 | ≤0.001 | -0.09 | ≤0.001 | 0.1 | ≤0.001 | 0.02 | 0.378 |
| Percentage of primary production land use surrounding green space (within 100m doughnut buffer) | Formula | 0.07 | ≤0.001 | 0.06 | ≤0.001 | 0.17 | ≤0.001 | 0.05 | ≤0.05 | 0.11 | ≤0.001 | 0.1 | ≤0.001 | 0.2 | ≤0.001 | 0.14 | ≤0.001 |
| Percentage of residential land use surrounding green space (within 100m doughnut buffer) | Formula | -0.02 | ≤0.001 | 0 | 0.998 | -0.12 | ≤0.001 | -0.09 | ≤0.001 | -0.04 | ≤0.001 | -0.01 | ≤0.01 | -0.12 | ≤0.001 | -0.15 | ≤0.001 |
| Percentage of transport land use surrounding green space (within 100m doughnut buffer) | Formula | 0.03 | ≤0.001 | 0.05 | ≤0.001 | 0 | 0.814 | 0.08 | ≤0.001 | 0.03 | ≤0.001 | 0.06 | ≤0.001 | 0.02 | 0.22 | 0.07 | ≤0.001 |
| Percentage of water land use surrounding green space (within 100m doughnut buffer) | Formula | -0.03 | ≤0.001 | -0.03 | ≤0.001 | 0.12 | ≤0.001 |  |  | -0.04 | ≤0.001 | -0.04 | ≤0.001 | 0.15 | ≤0.001 |  |  |
| Percentage of tree canopy to roads surrounding green space (within 100m doughnut buffer) | Formula | -0.47 | ≤0.001 | -0.49 | ≤0.001 | -0.17 | ≤0.001 | -0.36 | ≤0.001 | -0.52 | ≤0.001 | -0.54 | ≤0.001 | -0.2 | ≤0.001 | -0.4 | ≤0.001 |
| Percentage of tree canopy ≤4m to roads surrounding green space (within 100m doughnut buffer) | Formula | -0.21 | ≤0.001 | -0.2 | ≤0.001 | -0.03 | 0.088 | -0.38 | ≤0.001 | -0.25 | ≤0.001 | -0.24 | ≤0.001 | 0.06 | ≤0.01 | -0.41 | ≤0.001 |
| Percentage of tree canopy >4m to roads surrounding green space (within 100m doughnut buffer) | Formula | -0.47 | ≤0.001 | -0.49 | ≤0.001 | -0.18 | ≤0.001 | -0.32 | ≤0.001 | -0.51 | ≤0.001 | -0.53 | ≤0.001 | -0.2 | ≤0.001 | -0.37 | ≤0.001 |
| Percentage of tree canopy surrounding green space (within 100m doughnut buffer) | Formula | -0.39 | ≤0.001 | -0.39 | ≤0.001 | -0.16 | ≤0.001 | -0.25 | ≤0.001 | -0.44 | ≤0.001 | -0.45 | ≤0.001 | -0.16 | ≤0.001 | -0.31 | ≤0.001 |
| Percentage of tree canopy ≤4m surrounding green space (within 100m doughnut buffer) | Formula | -0.15 | ≤0.001 | -0.13 | ≤0.001 | -0.03 | 0.12 | -0.23 | ≤0.001 | -0.2 | ≤0.001 | -0.17 | ≤0.001 | 0.03 | 0.08 | -0.3 | ≤0.001 |
| Percentage of tree canopy >4m surrounding green space (within 100m doughnut buffer) | Formula | -0.39 | ≤0.001 | -0.4 | ≤0.001 | -0.17 | ≤0.001 | -0.24 | ≤0.001 | -0.45 | ≤0.001 | -0.46 | ≤0.001 | -0.16 | ≤0.001 | -0.29 | ≤0.001 |

Method of aggregating intersects over mesh blocks: Count=unweighted count | Sum=unweighted sum | w0 Sum = sum weighted by the ratio of the green space intersect with the buffer area to the green space area | w1 Mean = mean weighted by the ratio of the intersect area to the sum of all intersect areas | Max = maximum value over the intersects within the buffer | Quality = Value 0/1, aggregated to 1 if any intersects equal 1 (technically Quality = Max) | Formula = calculation of the index from other qualities that have already been aggregated as weighted sums or weighted means.

P-value signs: * ≤0.001, × ≤0.01, + ≤0.05

GS = green space

Table S9. Mean values and standard errors of aggregated green space features and SEIFA IRSD score (reversed) – 100m and 200m buffers.

|  |  | **Buffer 100m (Mean)** | | | | **Buffer 200m (Mean)** | | | | **Buffer 100m (Standard Error)** | | | | **Buffer 200m (Standard Error)** | | | |
| --- | --- | --- | --- | --- | --- | --- | --- | --- | --- | --- | --- | --- | --- | --- | --- | --- | --- |
| **Index description** | **Unit** | **Full set** | **Syd** | **New** | **Wol** | **Full set** | **Syd** | **New** | **Wol** | **Full set** | **Syd** | **New** | **Wol** | **Full set** | **Syd** | **New** | **Wol** |
| **SEIFA (IRSD) and general indices** | | | | | | | | | | | | | | | | | |
| Score IRSD reversed | NA | 475.889162 | 473.968919 | 479.498111 | 499.610717 | 475.889162 | 473.968919 | 479.498111 | 499.610717 | 0.49270387 | 0.54087343 | 1.42704435 | 1.74698755 | 0.49270387 | 0.54087343 | 1.42704435 | 1.74698755 |
| Quintile IRSD reversed | NA | 2.77500105 | 2.72218319 | 2.99505958 | 3.27998581 | 2.77500105 | 2.72218319 | 2.99505958 | 3.27998581 | 0.00667168 | 0.00726674 | 0.02026874 | 0.02445657 | 0.00667168 | 0.00726674 | 0.02026874 | 0.02445657 |
| N green spaces in MB | # | 1.27842248 | 1.29391654 | 1.1127451 | 1.16701903 | 1.70770609 | 1.74427937 | 1.36997886 | 1.44667274 | 0.00612647 | 0.00665652 | 0.01437045 | 0.0191421 | 0.00765084 | 0.00836773 | 0.01742234 | 0.02287503 |
| Intersect of buffer with green space | m2 | 653.412278 | 623.316088 | 967.542257 | 879.752793 | 1513.41213 | 1421.94283 | 2431.79644 | 2070.87737 | 9.35200868 | 9.39500602 | 55.0050561 | 55.41634 | 14.7412667 | 14.7470085 | 86.2223763 | 79.0803725 |
| Green space area (A) | m2 | 76415.9038 | 71099.1292 | 156448.397 | 84651.5013 | 98397.458 | 91320.6306 | 196039.749 | 107134.757 | 5512.26928 | 6015.94667 | 15796.6245 | 9394.32579 | 4106.19157 | 4511.57054 | 11607.0711 | 7423.03946 |
| GS perimeter (P) | m | 50.1624892 | 52.4744461 | 29.6007106 | 28.156986 | 101.934275 | 105.990984 | 68.0627207 | 68.3377783 | 0.68347981 | 0.73849737 | 1.99458258 | 1.96877101 | 0.91963065 | 0.99685215 | 2.87709408 | 2.84367615 |
| **Access indices – original sourced** | | | | | | | | | | | | | | | | | |
| Length of pathways within green space | m | 7.04462949 | 7.13602046 | 7.30490436 | 4.78633884 | 16.339834 | 16.5124377 | 17.9714665 | 10.9356416 | 0.21850234 | 0.22989295 | 1.10607475 | 0.73735295 | 0.31977756 | 0.33467009 | 1.71188389 | 1.0051291 |
| Length of bikeways within green space | m | 4.54006054 | 4.22013577 | 7.81151493 | 7.03376395 | 11.4260625 | 10.5816309 | 19.3214112 | 17.3266886 | 0.16445235 | 0.1682315 | 0.89614404 | 0.89868429 | 0.24321547 | 0.24607566 | 1.4075162 | 1.2452045 |
| Length of pathways covered by trees within green space | m | 2.1158957 | 2.25088628 | 0.95035846 | 0.74218069 | 4.95111045 | 5.25343651 | 2.63413514 | 2.07348401 | 0.08081072 | 0.08828771 | 0.1823028 | 0.18332776 | 0.11636037 | 0.12789893 | 0.26162983 | 0.28934591 |
| Length of bikeways covered by trees within green space | m | 1.18217098 | 1.2379176 | 0.45286884 | 0.93606274 | 2.99677501 | 3.12086338 | 1.28036312 | 2.80671352 | 0.05337221 | 0.05796111 | 0.11533232 | 0.19307503 | 0.07776903 | 0.08440321 | 0.17446389 | 0.33181235 |
| Length of pathways greater than 6 degrees gradient within green space | m | 1.92173168 | 1.95023666 | 2.17467734 | 0.99509474 | 4.24378849 | 4.29936946 | 4.72808094 | 2.55375942 | 0.0898593 | 0.09654631 | 0.36493624 | 0.21509855 | 0.11858345 | 0.12682633 | 0.51848309 | 0.33956833 |
| Length of bikeways greater than 6 degrees gradient within green space | m | 0.99250879 | 0.93973011 | 1.83523209 | 1.00828908 | 2.29886387 | 2.16598668 | 3.67204558 | 3.05565344 | 0.05554119 | 0.0576209 | 0.31618211 | 0.20063964 | 0.07340138 | 0.07530017 | 0.4173702 | 0.31987066 |
| Number of Railway Stations within and surrounding a green space (within 100m buffer) | # | 0.004751 | 0.00506874 | 0.00240839 | 0.0011015 | 0.01140372 | 0.01234609 | 0.0057535 | 0.00073013 | 0.00052806 | 0.00058015 | 0.00104193 | 0.00099409 | 0.0008185 | 0.00091072 | 0.0015189 | 0.00029274 |
| Number of Bus Stops within and surrounding a green space (within 100m buffer) | # | 0.27596442 | 0.29573125 | 0.09415905 | 0.09559187 | 0.55419848 | 0.59249043 | 0.22023589 | 0.25549976 | 0.00712792 | 0.00784242 | 0.00981365 | 0.01012726 | 0.00845899 | 0.00936681 | 0.01312778 | 0.01486374 |
| Number of ferry wharves within and surrounding a green space (within 100m buffer) | # | 0.0035263 | 0.00389784 | 0.00021413 | 0 | 0.00535858 | 0.00597828 | 0.00035945 | 0 | 0.00065882 | 0.00073054 | 0.00016419 | 0 | 0.00059302 | 0.00066399 | 0.00022892 | 0 |
| Area of GS which is greater than 6 degrees of Slope | m2 | 180.299682 | 172.819362 | 269.960387 | 221.14726 | 406.312037 | 383.092751 | 660.921727 | 519.540705 | 4.16737295 | 4.28201969 | 22.0128414 | 22.3440674 | 6.110615 | 6.20559029 | 35.8822951 | 28.0098032 |
| Length of Main Roads within 100 metres of the Green Space | m | 31.0559809 | 33.8882773 | 5.98143965 | 3.94907004 | 63.3895359 | 69.6620842 | 12.9743466 | 8.90995542 | 1.32144084 | 1.46054685 | 1.1781889 | 0.78079622 | 1.66649104 | 1.8588869 | 1.42107784 | 0.98327616 |
| **Access indices – calculated** | | | | | | | | | | | | | | | | | |
| Ratio of Area(A) to Perimeter (P) | m2 | 23.248259 | 20.2804807 | 56.1031393 | 43.1369873 | 25.2018031 | 21.9911775 | 58.4312506 | 43.4838824 | 0.28304351 | 0.25821413 | 2.10507408 | 1.58353254 | 0.19949539 | 0.18208951 | 1.38950338 | 1.04782618 |
| Ratio of Perimeter (P) to perimeter standardised by area (A) | m | 1.69090777 | 1.72114972 | 1.42652987 | 1.39712986 | 1.65928173 | 1.68988765 | 1.41764078 | 1.38782753 | 0.00709392 | 0.00776332 | 0.01103829 | 0.01023886 | 0.00452968 | 0.00499863 | 0.0070127 | 0.00651035 |
| Total number of transport stops/stations/wharves within and surrounding a green space (within 100m buffer) | # | 0.28424172 | 0.30469783 | 0.09678158 | 0.09669337 | 0.57096078 | 0.6108148 | 0.22634884 | 0.25622989 | 0.00731721 | 0.0080515 | 0.00983974 | 0.01034707 | 0.00874715 | 0.00968992 | 0.01320728 | 0.01487367 |
| Pathway Length per Area of Green Space | Ratio | 0.00923954 | 0.00976324 | 0.00489541 | 0.00384925 | 0.00794944 | 0.00844774 | 0.00392242 | 0.00364993 | 0.00027181 | 0.00029949 | 0.00041484 | 0.00033736 | 0.00014594 | 0.0001621 | 0.00022162 | 0.0002039 |
| Bikeway Length per Area of Green Space | Ratio | 0.0058789 | 0.00580462 | 0.0061324 | 0.00711275 | 0.00580727 | 0.00575588 | 0.00598027 | 0.00656417 | 0.00014945 | 0.00016285 | 0.00037505 | 0.00042909 | 9.5247E-05 | 0.00010453 | 0.00024569 | 0.00025543 |
| Trees over Pathway per Area of Green Space | Ratio | 0.00290771 | 0.00313837 | 0.00076785 | 0.00075262 | 0.00255978 | 0.00278312 | 0.0006934 | 0.00063426 | 0.00010364 | 0.00011427 | 8.6723E-05 | 0.00012225 | 5.6989E-05 | 6.3368E-05 | 4.8699E-05 | 6.4592E-05 |
| Trees over Bikeway per Area of Green Space | Ratio | 0.00173548 | 0.00184616 | 0.00044481 | 0.00104298 | 0.001751 | 0.00188002 | 0.00047745 | 0.00089162 | 5.953E-05 | 6.5477E-05 | 5.6521E-05 | 0.0001241 | 4.01E-05 | 0.00004454 | 3.7644E-05 | 6.7684E-05 |
| Percentage of GS which is greater than 6 degrees of Slope | % | 28.4507607 | 28.180594 | 31.9731902 | 29.5576163 | 27.912717 | 27.521907 | 32.050199 | 30.0104051 | 0.30751732 | 0.32687874 | 1.20647889 | 1.3131126 | 0.20349554 | 0.21695259 | 0.78474611 | 0.85229957 |
| Pathway Length > 6 degrees per Area of GS | Ratio | 0.00266528 | 0.00282105 | 0.00138512 | 0.00105377 | 0.00227604 | 0.00242229 | 0.0011535 | 0.00093859 | 0.00012892 | 0.00014241 | 0.00014255 | 0.00017573 | 6.94E-05 | 7.7235E-05 | 9.4235E-05 | 0.00010228 |
| Bikeway Length > 6 degrees slope per Area of GS | Ratio | 0.00126939 | 0.00127815 | 0.00115894 | 0.00122871 | 0.00121415 | 0.00122939 | 0.0010024 | 0.00119778 | 5.7788E-05 | 6.3212E-05 | 0.0001258 | 0.00015407 | 3.4426E-05 | 3.7758E-05 | 8.8159E-05 | 9.6789E-05 |
| **Amenities/Activities indices – original sourced** | | | | | | | | | | | | | | | | | |
| Number of public toilet facilities within and surrounding a green space (within 100m buffer) | # | 0.04014107 | 0.04290207 | 0.01256111 | 0.01777479 | 0.0883834 | 0.09512033 | 0.03036856 | 0.03487296 | 0.00213307 | 0.00235584 | 0.00224329 | 0.00298793 | 0.00285845 | 0.00318569 | 0.00336048 | 0.00342718 |
| Number of cafes within and surrounding a green space (within 100m buffer) | # | 0.06941398 | 0.07522951 | 0.0259717 | 0.003349 | 0.14370006 | 0.15687971 | 0.05485806 | 7.13E-03 | 0.00375764 | 0.00414269 | 0.00667734 | 0.0010688 | 0.00523201 | 0.00581991 | 0.00922236 | 1.63E-03 |
| Number of hotels/bars within and surrounding a green space (within 100m buffer) | # | 0.02474502 | 0.02715312 | 0.00473823 | 0 | 0.04817723 | 0.05331335 | 0.00965408 | 3.40E-07 | 0.00215981 | 0.00239264 | 0.00140854 | 0 | 0.00268945 | 0.00300841 | 0.00174987 | 3.40E-07 |
| Number of restaurants/takeaways within and surrounding a green space (within 100m buffer) | # | 0.29389407 | 0.3219642 | 0.04867973 | 0.02098439 | 0.58908202 | 0.65186435 | 0.09277212 | 0.03305762 | 0.01872813 | 0.02073968 | 0.01049353 | 0.00651748 | 0.02521912 | 0.02820278 | 0.01350913 | 0.00564197 |
| Number of restaurants within and surrounding a green space (within 100m buffer) | # | 0.20769442 | 0.22710029 | 0.04073636 | 0.01570055 | 0.41502937 | 0.45856856 | 0.07582142 | 0.02298754 | 0.0136539 | 0.01511665 | 0.00973589 | 0.0056068 | 0.0184289 | 0.02060537 | 0.01217262 | 0.00483396 |
| Number of takeaways within and surrounding a green space (within 100m buffer) | # | 0.08619965 | 0.09486391 | 0.00794337 | 0.00528384 | 0.17405265 | 0.19329579 | 0.0169507 | 0.01007008 | 0.00589627 | 0.00653143 | 0.00262702 | 0.00129423 | 0.00750847 | 0.00839656 | 0.00391016 | 0.00159413 |
| Number of supermarkets/greengrocers within and surrounding a green space (within 100m buffer) | # | 0.03673377 | 0.04033078 | 0.00510554 | 0.00202795 | 0.07649388 | 0.08432829 | 0.01399571 | 0.00784063 | 0.00282108 | 0.00312554 | 0.00133767 | 0.00062596 | 0.0033444 | 0.00373833 | 0.00241069 | 0.001499 |
| Presence of parkrun event | 1/0 | 0.01223935 | 0.00945199 | 0.05065359 | 0.02114165 | 0.01462749 | 0.01275498 | 0.03664553 | 0.02187785 | 0.00104698 | 0.00097033 | 0.0088715 | 0.00662153 | 0.00078403 | 0.00077562 | 0.0049896 | 0.00441869 |
| **Beach/Coast indices – original sourced** | | | | | | | | | | | | | | | | | |
| Presence of coastline | 1/0 | 0.13345422 | 0.12096531 | 0.27124183 | 0.21775899 | 0.13045332 | 0.12014523 | 0.22762509 | 0.20145852 | 0.00323813 | 0.00327004 | 0.01798662 | 0.0189971 | 0.00219949 | 0.00224726 | 0.01113489 | 0.01211536 |
| Area of beach within and surrounding a green space (within 100m buffer) | m2 | 44.1304796 | 41.0598836 | 59.7136602 | 88.5283035 | 109.879946 | 104.857509 | 141.367673 | 164.98804 | 4.50147251 | 4.79807291 | 15.9260128 | 20.3369549 | 7.28906103 | 7.88974598 | 23.5306488 | 26.087094 |
| **Biodiversity indices – original sourced** | | | | | | | | | | | | | | | | | |
| Number of bird species/species habitat likely to occur within green space | # | 30.6978241 | 30.2669683 | 36.9019608 | 31.7293869 | 30.2844045 | 29.8778006 | 35.4601832 | 31.3482224 | 0.13011614 | 0.13455463 | 0.62769613 | 0.65529042 | 0.08763467 | 0.09109567 | 0.40499074 | 0.42165539 |
| Number of fish species/species habitat likely to occur within green space | # | 1.86019946 | 1.81759678 | 2.00490196 | 2.56871036 | 1.72570259 | 1.69158745 | 1.68710359 | 2.42661805 | 0.03200937 | 0.03394534 | 0.11455421 | 0.15715655 | 0.02120665 | 0.02262762 | 0.06996299 | 0.10200607 |
| Number of flora species/species habitat likely to occur within green space | # | 7.50897552 | 7.2860734 | 9.41993464 | 9.7230444 | 7.5593842 | 7.30287106 | 9.64270613 | 9.75934366 | 0.02521827 | 0.02622102 | 0.07658269 | 0.09769256 | 0.01761526 | 0.01837512 | 0.0491251 | 0.06425808 |
| Number of frog species/species habitat likely to occur within green space | # | 1.71514053 | 1.73363499 | 1.09640523 | 2.12684989 | 1.70032837 | 1.71738403 | 1.10782241 | 2.14129444 | 0.0051854 | 0.00528805 | 0.01562672 | 0.0207784 | 0.00361044 | 0.00369676 | 0.01084149 | 0.01368291 |
| Number of mammal species/species habitat likely to occur within green space | # | 7.71196736 | 7.66143791 | 8.20098039 | 8.14164905 | 7.62198814 | 7.57101228 | 7.97533474 | 8.13764813 | 0.01924083 | 0.02009881 | 0.07614849 | 0.10914383 | 0.01284021 | 0.01352072 | 0.04577497 | 0.06951644 |
| Number of other animal species/species habitat likely to occur within green space | # | 0.18748867 | 0.20794369 | 0 | 0 | 0.19002943 | 0.21286963 | 0 | 0 | 0.0039189 | 0.00429699 | 0 | 0 | 0.00268546 | 0.00296939 | 0 | 0 |
| Number of reptile species/species habitat likely to occur within green space | # | 1.46001813 | 1.38471594 | 2.05065359 | 2.27906977 | 1.38223378 | 1.3098457 | 1.82170543 | 2.19507748 | 0.02132209 | 0.02208986 | 0.09949206 | 0.11121015 | 0.01437104 | 0.01494819 | 0.06389958 | 0.07251492 |
| Area of land with biodiversity value within green space | m2 | 40.4381463 | 39.3599038 | 79.187832 | 12.9715808 | 104.425417 | 99.1995747 | 212.806861 | 63.9507387 | 1.99720579 | 2.05041093 | 13.3758542 | 2.40111904 | 3.01229744 | 3.05671429 | 20.0416766 | 7.51227956 |
| Area of land with biodiversity value outside of green space within 100m | m2 | 204.801557 | 211.191795 | 242.5354 | 21.6217917 | 416.266783 | 422.869994 | 566.17171 | 96.3578419 | 12.3359263 | 13.169268 | 59.8787566 | 3.79780068 | 15.3860298 | 16.2209229 | 84.944214 | 13.1387796 |
| **Biodiversity indices – calculated** | | | | | | | | | | | | | | | | | |
| Total number of species/species habitat likely to occur within green space | # | 51.1416138 | 50.358371 | 59.6748366 | 56.5687104 | 50.464071 | 49.6833708 | 57.6948555 | 56.0082042 | 0.19692999 | 0.2055632 | 0.84432799 | 0.96462035 | 0.13242683 | 0.13902122 | 0.53944856 | 0.62367301 |
| Shannon biodiversity index | unit | 1.11467664 | 1.11170461 | 1.0864777 | 1.2136505 | 1.11101027 | 1.10773303 | 1.08210195 | 1.21094054 | 0.00124635 | 0.00131738 | 0.00462755 | 0.00429979 | 0.00085221 | 0.00090558 | 0.0029564 | 0.00289054 |
| Simpson biodiversity index (range 0 to 1=most diverse) | unit | 0.57811559 | 0.57661401 | 0.56747244 | 0.62345768 | 0.57749723 | 0.57560659 | 0.56971232 | 0.62364467 | 0.00051749 | 0.00053896 | 0.00234896 | 0.00179977 | 0.00035726 | 0.0003735 | 0.00155845 | 0.00117817 |
| Percentage of land with biodiversity value within green space | % | 6.57404669 | 6.58289552 | 9.55178778 | 2.53519015 | 6.98573353 | 7.02015903 | 9.03729054 | 3.67507868 | 0.17651905 | 0.18681496 | 0.87950215 | 0.40569386 | 0.12216983 | 0.13011166 | 0.56330522 | 0.33160848 |
| Percentage of land with biodiversity value outside of green space within 100m | % | 2.65810661 | 2.52267583 | 5.67824332 | 1.59792786 | 2.69080897 | 2.57424372 | 5.01478158 | 1.90898897 | 0.06754918 | 0.06733497 | 0.48728033 | 0.22804857 | 0.04660106 | 0.04687694 | 0.30243751 | 0.17900888 |
| **Incivilities indices – original sourced** | | | | | | | | | | | | | | | | | |
| Density of alcohol-related assaults | Score 0-3 | 0.48368087 | 0.50336853 | 0.43954248 | 0.12684989 | 0.47208836 | 0.49228491 | 0.40944327 | 0.16773017 | 0.00948513 | 0.01014 | 0.0394775 | 0.02398073 | 0.00640183 | 0.0068892 | 0.0248557 | 0.01775783 |
| Density of malicious damage | Score 0-3 | 1.18939257 | 1.20120664 | 1.19607843 | 0.93234672 | 1.19638364 | 1.20780586 | 1.18322763 | 0.99544211 | 0.01210835 | 0.01278172 | 0.05213761 | 0.05311435 | 0.00823231 | 0.00873834 | 0.03360542 | 0.03514401 |
| Density of non-domestic violence assaults | Score 0-3 | 0.55466908 | 0.57335344 | 0.54084967 | 0.17970402 | 0.55089769 | 0.57277982 | 0.49682875 | 0.20328168 | 0.00992721 | 0.0105843 | 0.04232863 | 0.02757221 | 0.00676155 | 0.00726558 | 0.02670279 | 0.01881599 |
| Density of robberies | Score 0-3 | 0.35303717 | 0.3635998 | 0.33986928 | 0.14799154 | 0.37225468 | 0.38436918 | 0.35870331 | 0.1586144 | 0.0081404 | 0.00868837 | 0.03427582 | 0.02464866 | 0.00567402 | 0.00608822 | 0.02308624 | 0.01667728 |
| Density of steal from persons | Score 0-3 | 0.3030825 | 0.31654098 | 0.28431373 | 0.04439746 | 0.29941575 | 0.31600822 | 0.24876674 | 0.04831358 | 0.00767604 | 0.00825651 | 0.03096464 | 0.01235292 | 0.00523752 | 0.00568004 | 0.0197037 | 0.00847905 |
| **Incivilities indices – calculated** | | | | | | | | | | | | | | | | | |
| Mean level of incivilities | Score 0-3 | 0.57677244 | 0.59161388 | 0.56013072 | 0.28625793 | 0.57820803 | 0.5946496 | 0.53939394 | 0.31467639 | 0.00792053 | 0.00846877 | 0.03260112 | 0.02109627 | 0.0054092 | 0.00582512 | 0.02095253 | 0.01449161 |
| **Land cover indices – original sourced** | | | | | | | | | | | | | | | | | |
| Area of bare earth within green space | m2 | 77.1878793 | 73.4606738 | 129.49109 | 88.1480133 | 186.535018 | 177.728817 | 304.544171 | 202.610823 | 2.14557602 | 2.14585298 | 14.7815824 | 9.79382098 | 3.45598616 | 3.46191557 | 22.4970601 | 15.2751221 |
| Area of buildings within green space | m2 | 15.5897533 | 15.6603414 | 16.3012247 | 13.1814802 | 33.758653 | 33.6167078 | 40.470861 | 27.7969005 | 0.68190974 | 0.7426727 | 1.66006749 | 2.07900074 | 0.98983915 | 1.08524378 | 2.71019717 | 2.49652244 |
| Area of built up area within green space | m2 | 29.842676 | 29.4176664 | 28.5240967 | 40.5088851 | 64.08812 | 62.5110525 | 68.6324498 | 88.4223657 | 0.88199516 | 0.92993711 | 3.22223354 | 4.81215529 | 1.17368347 | 1.22117501 | 5.12584121 | 6.48481462 |
| Area of open grass within green space | m2 | 164.053899 | 148.449471 | 300.481996 | 316.671445 | 391.820817 | 349.730025 | 743.70199 | 743.051992 | 3.54742876 | 3.46912318 | 22.1699852 | 24.425994 | 5.25058429 | 5.12432502 | 30.6102663 | 34.5242697 |
| Area of other vegetation within green space | m2 | 104.215348 | 99.0641727 | 144.001274 | 161.384546 | 243.750607 | 225.382444 | 395.36993 | 399.534059 | 2.03455221 | 2.09565901 | 8.8867545 | 12.8793233 | 3.07718825 | 3.1025194 | 15.5464428 | 19.1120528 |
| Area of roads/paths within green space | m2 | 27.5459204 | 19.1554661 | 126.178045 | 76.9273186 | 65.6332024 | 44.0937689 | 294.613261 | 182.116517 | 1.02686698 | 0.76572141 | 11.7509427 | 7.08462627 | 1.5258128 | 1.08860204 | 16.6675305 | 9.44751107 |
| Area of shadow within green space | m2 | 7.55808688 | 8.3626185 | 0.07864892 | 0.26546313 | 16.2672662 | 18.1606818 | 0.25001505 | 0.71085576 | 0.35085494 | 0.38812977 | 0.01527141 | 0.09542847 | 0.43811815 | 0.48881199 | 0.03621019 | 0.19996837 |
| Area of swimming pools within green space | m2 | 0.70640277 | 0.68047812 | 0.59775866 | 1.39349817 | 1.70364912 | 1.70706026 | 1.75548683 | 1.57126079 | 0.0658508 | 0.06552224 | 0.19482538 | 0.6306302 | 0.09248319 | 0.09673338 | 0.37554238 | 0.51562 |
| Area of tree canopy within green space | m2 | 213.450679 | 217.541383 | 190.820612 | 156.459023 | 475.966614 | 479.669178 | 507.47947 | 364.281719 | 3.84180509 | 4.08580936 | 14.6406042 | 16.6910587 | 5.71867189 | 6.10452171 | 24.2371177 | 19.4143934 |
| Area of water within green space | m2 | 6.13384557 | 4.39682128 | 24.0863456 | 19.5464055 | 17.2770952 | 13.0146815 | 60.6751804 | 42.8039039 | 0.49616803 | 0.43207696 | 3.67806287 | 5.25557948 | 0.85927338 | 0.79651694 | 6.71193475 | 5.33534419 |
| **Land cover indices - calculated** | | | | | | | | | | | | | | | | | |
| Percentage of bare earth within green space | % | 10.3631429 | 10.4315 | 10.1757816 | 9.16409443 | 10.3572965 | 10.558523 | 8.8470525 | 8.45566527 | 0.15308758 | 0.16394046 | 0.51779572 | 0.64013147 | 0.10130603 | 0.10955692 | 0.29467415 | 0.4068667 |
| Percentage of buildings within green space | % | 2.52797728 | 2.66311638 | 1.3005056 | 1.26569248 | 2.42725393 | 2.55361972 | 1.49529992 | 1.21180222 | 0.06062054 | 0.06658592 | 0.09860798 | 0.11305011 | 0.03866931 | 0.04265294 | 0.09003474 | 0.0644715 |
| Percentage of built up area within green space | % | 5.16413725 | 5.43415404 | 2.05337546 | 3.49304839 | 4.86586896 | 5.13414087 | 1.95996224 | 3.48488945 | 0.09670255 | 0.1060476 | 0.12578323 | 0.22145233 | 0.05978922 | 0.06600409 | 0.07921293 | 0.15003445 |
| Percentage of open grass within green space | % | 23.3879623 | 22.4220943 | 29.9505053 | 35.267893 | 24.5391977 | 23.52783 | 31.392617 | 35.050113 | 0.22665818 | 0.23749579 | 0.91386696 | 1.07248116 | 0.15353192 | 0.16142841 | 0.6074285 | 0.70088045 |
| Percentage of other vegetation within green space | % | 15.5557912 | 15.1934432 | 18.5628132 | 19.30781 | 15.8014416 | 15.4326278 | 18.3699203 | 19.5448735 | 0.13321571 | 0.14158653 | 0.52152713 | 0.52217688 | 0.08834076 | 0.09443547 | 0.31987291 | 0.35211461 |
| Percentage of roads/paths within green space | % | 3.11100311 | 2.50671961 | 9.62202492 | 7.43356618 | 3.19870264 | 2.55991551 | 9.33257001 | 7.50279459 | 0.06741035 | 0.0666486 | 0.38877079 | 0.29013148 | 0.04459294 | 0.04421474 | 0.23649742 | 0.1878842 |
| Percentage of shadow within green space | % | 1.2344506 | 1.36643649 | 0.01331991 | 0.0304521 | 1.13371133 | 1.26676187 | 0.01450737 | 0.03237362 | 0.03831522 | 0.04226988 | 0.00222839 | 0.00636023 | 0.02282611 | 0.02540228 | 0.0014792 | 0.00387532 |
| Percentage of swimming pools within green space | % | 0.10169341 | 0.10531357 | 0.05431095 | 0.0866278 | 0.11288792 | 0.11808053 | 0.06590143 | 0.07418253 | 0.00726805 | 0.00788293 | 0.01142215 | 0.03195849 | 0.00488871 | 0.00535544 | 0.00855986 | 0.01855823 |
| Percentage of tree canopy within green space | % | 36.6271416 | 38.0532074 | 25.2451692 | 21.2754676 | 35.6714739 | 37.047272 | 25.98104 | 21.8483965 | 0.27942417 | 0.29774222 | 0.9016942 | 0.97877966 | 0.18397932 | 0.19714034 | 0.58591934 | 0.64671399 |
| Percentage of water within green space | % | 0.66869353 | 0.51828541 | 2.20500951 | 1.85359084 | 0.67577619 | 0.53500177 | 1.74493188 | 1.9898062 | 0.03563076 | 0.03409236 | 0.24041736 | 0.26516372 | 0.02401462 | 0.02312065 | 0.13310066 | 0.19053317 |
| **Land use indices – original sourced** | | | | | | | | | | | | | | | | | |
| Area of commercial land use surrounding green space (within 100m doughnut buffer) | m2 | 443.72053 | 472.63139 | 241.179947 | 97.9199699 | 984.553187 | 1058.27439 | 493.091995 | 213.52083 | 20.4421551 | 22.4757958 | 41.193653 | 26.2450059 | 29.3264451 | 32.4941395 | 60.398236 | 38.46958 |
| Area of education land use surrounding green space (within 100m doughnut buffer) | m2 | 209.461328 | 222.345305 | 58.5967154 | 133.769971 | 454.346174 | 483.973503 | 157.944369 | 272.400214 | 10.3822955 | 11.3659848 | 10.7286114 | 35.1601073 | 13.1128982 | 14.4627189 | 18.7534888 | 40.8217611 |
| Area of hospital/medical land use surrounding green space (within 100m doughnut buffer) | m2 | 28.7473556 | 30.3617852 | 24.4758359 | 0.330162 | 65.2390439 | 68.8245796 | 57.6617896 | 6.62108911 | 4.71711006 | 5.20458178 | 8.59666023 | 0.21055172 | 5.67070483 | 6.27725277 | 13.992895 | 3.7827219 |
| Area of industrial land use surrounding green space (within 100m doughnut buffer) | m2 | 77.8492198 | 78.8376078 | 49.413547 | 93.8600295 | 189.924662 | 192.574863 | 141.822565 | 201.574817 | 7.366797 | 7.97012695 | 13.1845682 | 33.7496112 | 10.4261881 | 11.2076245 | 24.5046658 | 54.1008262 |
| Area of other land use surrounding green space (within 100m doughnut buffer) | m2 | 5.22332734 | 5.36688425 | 0.60937991 | 8.17483344 | 12.0737344 | 11.5813652 | 7.79071443 | 27.009351 | 1.38262013 | 1.52724431 | 0.29775273 | 2.86646156 | 2.06831573 | 2.27648166 | 2.15668683 | 7.72519472 |
| Area of parkland land use surrounding green space (within 100m doughnut buffer) | m2 | 1040.25899 | 1090.65652 | 574.278642 | 583.54982 | 2368.9376 | 2471.91051 | 1506.89315 | 1519.08278 | 31.3319165 | 34.4402098 | 50.2015484 | 64.3084599 | 42.695081 | 47.1251563 | 86.7183615 | 99.8785806 |
| Area of primary production land use surrounding green space (within 100m doughnut buffer) | m2 | 6.31999778 | 5.56416106 | 6.22431596 | 22.3355442 | 14.7685176 | 13.3382079 | 5.77461213 | 53.6956135 | 1.79152542 | 1.92493696 | 4.95853846 | 8.11037648 | 2.31007291 | 2.43900095 | 2.36556923 | 16.1737847 |
| Area of residential land use surrounding green space (within 100m doughnut buffer) | m2 | 10070.6082 | 10838.752 | 2976.34169 | 3099.15214 | 18729.1813 | 20129.1571 | 6524.85948 | 7801.41455 | 152.773165 | 166.714127 | 221.980389 | 237.437084 | 183.45956 | 201.501169 | 290.512892 | 366.899364 |
| Area of transport land use surrounding green space (within 100m doughnut buffer) | m2 | 108.006187 | 119.771793 | 0.17121197 | 0.15428601 | 225.466371 | 252.507518 | 0.5075792 | 0.45652856 | 10.4935696 | 11.632388 | 0.17055011 | 0.14534445 | 13.0764992 | 14.6370864 | 0.4129458 | 0.32289104 |
| Area of water land use surrounding green space (within 100m doughnut buffer) | m2 | 7.60302597 | 8.42086395 | 0.18935374 | 0 | 14.0831769 | 15.6945688 | 1.19944005 | 0 | 2.56806218 | 2.8481191 | 0.13902083 | 0 | 3.11795218 | 3.49234313 | 0.56276214 | 0 |
| Area of tree canopy surrounding green space (within 100m doughnut buffer) | m2 | 808.955177 | 887.103852 | 90.5830652 | 70.1839063 | 1560.71231 | 1721.97047 | 201.289344 | 185.803513 | 17.0922012 | 18.7472779 | 10.621054 | 5.99048852 | 22.9324115 | 25.3851503 | 14.1372647 | 10.0034585 |
| Area of tree canopy ≤4m surrounding green space (within 100m doughnut buffer) | m2 | 59.9309726 | 64.9576687 | 12.5641859 | 13.9134161 | 116.166824 | 126.073201 | 27.8707299 | 37.8983419 | 1.08527888 | 1.18575244 | 1.47341847 | 1.40430365 | 1.36918134 | 1.50268315 | 2.29186989 | 2.55539131 |
| Area of tree canopy >4m surrounding green space (within 100m doughnut buffer) | m2 | 726.026786 | 797.020885 | 74.8203835 | 53.0796569 | 1399.88178 | 1546.86657 | 166.597649 | 139.143378 | 15.9452256 | 17.4995183 | 9.20212075 | 4.47312071 | 21.4719786 | 23.7846399 | 12.0389112 | 7.30066369 |
| **Land use indices – calculated** | | | | | | | | | | | | | | | | | |
| Percentage of commercial land use surrounding green space (within 100m doughnut buffer) | % | 3.14261832 | 3.10936253 | 4.38603601 | 2.23301417 | 3.22139331 | 3.22074177 | 3.95507134 | 2.2847929 | 0.08587721 | 0.0890808 | 0.46885836 | 0.35948088 | 0.05985174 | 0.06281189 | 0.28694787 | 0.247243 |
| Percentage of education land use surrounding green space (within 100m doughnut buffer) | % | 1.98231039 | 1.92330582 | 2.44830208 | 2.61997098 | 2.17146397 | 2.10859207 | 2.65992636 | 2.73934844 | 0.05339709 | 0.05526864 | 0.24942786 | 0.30778432 | 0.03824263 | 0.03983737 | 0.17017872 | 0.2033114 |
| Percentage of hospital/medical land use surrounding green space (within 100m doughnut buffer) | % | 0.25848201 | 0.24428002 | 0.61680949 | 0.09345533 | 0.28116646 | 0.27324693 | 0.50486192 | 0.14293101 | 0.02053649 | 0.02128595 | 0.12482697 | 0.05087655 | 0.0152161 | 0.01612038 | 0.07285773 | 0.04705823 |
| Percentage of industrial land use surrounding green space (within 100m doughnut buffer) | % | 1.00286177 | 0.92169677 | 1.90249015 | 1.54538461 | 1.16283476 | 1.06789886 | 2.10889772 | 1.75064678 | 0.04919068 | 0.04873882 | 0.31776135 | 0.30658184 | 0.03732476 | 0.03778225 | 0.20871978 | 0.20597914 |
| Percentage of other land use surrounding green space (within 100m doughnut buffer) | % | 0.20860039 | 0.16171346 | 0.38255984 | 0.96933469 | 0.23769035 | 0.16384634 | 0.72294812 | 1.01909042 | 0.0251189 | 0.02336715 | 0.15465645 | 0.24590449 | 0.01868508 | 0.01676211 | 0.13328155 | 0.16311818 |
| Percentage of parkland land use surrounding green space (within 100m doughnut buffer) | % | 12.8883051 | 12.3333988 | 18.9523026 | 16.709397 | 13.2817595 | 12.7919843 | 17.9569189 | 16.5802227 | 0.15757119 | 0.16420711 | 0.7213139 | 0.7537123 | 0.10906434 | 0.11487234 | 0.45181331 | 0.49919841 |
| Percentage of primary production land use surrounding green space (within 100m doughnut buffer) | % | 0.18759458 | 0.1339122 | 0.24255929 | 1.24516933 | 0.19621701 | 0.15145779 | 0.20804418 | 1.03501554 | 0.02305038 | 0.02018908 | 0.10425932 | 0.29706759 | 0.01616289 | 0.01491545 | 0.05975715 | 0.17815086 |
| Percentage of residential land use surrounding green space (within 100m doughnut buffer) | % | 79.5803417 | 80.3507779 | 70.9373976 | 74.5644714 | 78.72479 | 79.4197952 | 71.7839951 | 74.440783 | 0.19114285 | 0.19896774 | 0.85892234 | 0.92292407 | 0.13231792 | 0.13904036 | 0.54658964 | 0.61580495 |
| Percentage of transport land use surrounding green space (within 100m doughnut buffer) | % | 0.6882432 | 0.76184459 | 0.00884237 | 0.01980249 | 0.67182955 | 0.7516262 | 0.00850964 | 0.0071693 | 0.0384866 | 0.04261278 | 0.00723243 | 0.01463589 | 0.02504802 | 0.02800486 | 0.00448081 | 0.00574457 |
| Percentage of water land use surrounding green space (within 100m doughnut buffer) | % | 0.0606426 | 0.0597079 | 0.12270067 | 0 | 0.0508551 | 0.05081059 | 0.09082673 | 0 | 0.01077189 | 0.01083859 | 0.08166641 | 0 | 0.00696209 | 0.00721987 | 0.04348811 | 0 |
| Percentage of tree canopy to roads surrounding green space (within 100m doughnut buffer) | % | 18.4913274 | 19.5730299 | 8.25421277 | 8.64596648 | 17.9920303 | 19.0976605 | 8.46550732 | 8.83193146 | 0.10852595 | 0.11297943 | 0.2720813 | 0.29615111 | 0.07219888 | 0.07553724 | 0.17835121 | 0.20698841 |
| Percentage of tree canopy ≤4m to roads surrounding green space (within 100m doughnut buffer) | % | 1.70548168 | 1.73623716 | 1.36186291 | 1.49362256 | 1.69658845 | 1.7313416 | 1.30587399 | 1.52683222 | 0.00985765 | 0.00945381 | 0.08342514 | 0.03838903 | 0.00624542 | 0.00620557 | 0.04254778 | 0.02465502 |
| Percentage of tree canopy >4m to roads surrounding green space (within 100m doughnut buffer) | % | 16.1772665 | 17.1944169 | 6.63860575 | 6.80605418 | 15.6906486 | 16.724782 | 6.90954796 | 6.95538136 | 0.10445057 | 0.10932052 | 0.22939203 | 0.27089857 | 0.06939708 | 0.0729973 | 0.15633334 | 0.19155533 |
| Percentage of tree canopy surrounding green space (within 100m doughnut buffer) | % | 5.12153502 | 5.47361865 | 1.89189137 | 1.78428287 | 5.03224247 | 5.39914022 | 1.96213541 | 1.87439961 | 0.03734866 | 0.03949085 | 0.06372564 | 0.05835099 | 0.02475163 | 0.02631391 | 0.04193025 | 0.04081032 |
| Percentage of tree canopy ≤4m surrounding green space (within 100m doughnut buffer) | % | 0.44686672 | 0.46255757 | 0.28023837 | 0.32753837 | 0.45299909 | 0.47048425 | 0.27683666 | 0.34115696 | 0.00258302 | 0.00266102 | 0.01247097 | 0.01108907 | 0.00176332 | 0.00183756 | 0.0071322 | 0.00727238 |
| Percentage of tree canopy >4m surrounding green space (within 100m doughnut buffer) | % | 4.5115252 | 4.83757307 | 1.55132089 | 1.38140085 | 4.4140582 | 4.7520012 | 1.62490824 | 1.45535493 | 0.03572234 | 0.03790755 | 0.05750309 | 0.04935158 | 0.02357422 | 0.02516023 | 0.03806578 | 0.03521917 |

IRSD = Index of Relative Social Disadvantage; when reversed, the higher the score the greater the disadvantage.

IRSD scores were reversed calculating Max - X + Min, where Max and Min are scores in the original NSW-wise data, and X is a score to be reversed.

m=metres | sq m=square metres | number = number of items

Table S10. Mean values and standard errors of aggregated green space features and SEIFA IRSD score (reversed) – 300m and 400m buffers.

|  |  | **Buffer 300m (Mean)** | | | | **Buffer 400m (Mean)** | | | | **Buffer 300m (Standard Error)** | | | | **Buffer 400m (Standard Error)** | | | |
| --- | --- | --- | --- | --- | --- | --- | --- | --- | --- | --- | --- | --- | --- | --- | --- | --- | --- |
| **Index description** | **Unit** | **Full set** | **Syd** | **New** | **Wol** | **Full set** | **Syd** | **New** | **Wol** | **Full set** | **Syd** | **New** | **Wol** | **Full set** | **Syd** | **New** | **Wol** |
| **SEIFA (IRSD) and general indices** | | | | | | | | | | | | | | | | | |
| Score IRSD reversed | NA | 475.889162 | 473.968919 | 479.498111 | 499.610717 | 475.889162 | 473.968919 | 479.498111 | 499.610717 | 0.49270387 | 0.54087343 | 1.42704435 | 1.74698755 | 0.49270387 | 0.54087343 | 1.42704435 | 1.74698755 |
| Quintile IRSD reversed | NA | 2.77500105 | 2.72218319 | 2.99505958 | 3.27998581 | 2.77500105 | 2.72218319 | 2.99505958 | 3.27998581 | 0.00667168 | 0.00726674 | 0.02026874 | 0.02445657 | 0.00667168 | 0.00726674 | 0.02026874 | 0.02445657 |
| N green spaces in MB buffer | # | 2.33118053 | 2.4096876 | 1.68276823 | 1.77657005 | 3.2033864 | 3.34836136 | 2.05968468 | 2.23009709 | 0.01013003 | 0.01116089 | 0.02069493 | 0.0261862 | 0.01345441 | 0.01487991 | 0.024417 | 0.03144856 |
| Intersect of buffer with green space | m2 | 2936.20254 | 2750.4154 | 4802.01694 | 3817.90075 | 5285.41055 | 4987.2038 | 8322.70741 | 6401.89271 | 23.4075691 | 23.5208656 | 132.113551 | 114.149834 | 36.8570937 | 37.3461533 | 197.599224 | 172.518692 |
| Green space area (A) | m2 | 116659.739 | 108838.307 | 219879.231 | 121703.259 | 144099.421 | 134214.514 | 263858.68 | 156434.767 | 3528.74564 | 3900.83428 | 9793.7218 | 6200.98323 | 3462.81871 | 3833.37663 | 9878.3356 | 6837.70758 |
| GS perimeter (P) | m | 182.765093 | 190.106613 | 127.975476 | 123.300538 | 312.45593 | 326.415549 | 214.452547 | 203.059862 | 1.32745062 | 1.44735403 | 4.0051384 | 4.00318655 | 1.95325716 | 2.14078232 | 5.48920215 | 5.70824215 |
| **Access indices – original sourced** | | | | | | | | | | | | | | | | | |
| Length of pathways within green space | m | 30.9449113 | 31.2803998 | 34.1306954 | 20.8304612 | 54.2807639 | 55.1773239 | 57.5317606 | 34.9107714 | 0.4770056 | 0.5007517 | 2.48342747 | 1.4105398 | 0.70942776 | 0.75015528 | 3.470433 | 2.08696934 |
| Length of bikeways within green space | m | 22.4558235 | 20.7465376 | 38.3669672 | 32.1990403 | 40.0869118 | 37.1774342 | 66.7365823 | 54.8388224 | 0.36463535 | 0.37121911 | 1.99334973 | 1.74864899 | 0.5429467 | 0.55807502 | 2.73162612 | 2.56170361 |
| Length of pathways covered by trees within green space | m | 9.3918636 | 9.97103052 | 5.36145527 | 4.11164787 | 16.6824537 | 17.7693362 | 9.65287149 | 6.94236471 | 0.17538636 | 0.19355453 | 0.4139842 | 0.44537714 | 0.26640924 | 0.29480885 | 0.63869651 | 0.67767447 |
| Length of bikeways covered by trees within green space | m | 5.89899132 | 6.16185857 | 2.63003149 | 5.38207583 | 10.7119389 | 11.2273268 | 5.00373073 | 9.18776764 | 0.11967688 | 0.13087378 | 0.21116487 | 0.51320322 | 0.185419 | 0.20307506 | 0.33893084 | 0.80154684 |
| Length of pathways greater than 6 degrees gradient within green space | m | 7.82494122 | 7.88625761 | 8.97807551 | 5.232121 | 13.5850795 | 13.7555973 | 15.1581259 | 8.66435456 | 0.16383189 | 0.17378821 | 0.7721639 | 0.51140014 | 0.23869876 | 0.25450552 | 1.07365232 | 0.73973327 |
| Length of bikeways greater than 6 degrees gradient within green space | m | 4.30076572 | 4.03989582 | 6.55355251 | 6.01073786 | 7.68329967 | 7.23473734 | 11.5033494 | 10.3189813 | 0.10101124 | 0.10279957 | 0.5466849 | 0.51180837 | 0.14582384 | 0.1472462 | 0.76551153 | 0.80405524 |
| Number of Railway Stations within and surrounding a green space (within 100m buffer) | # | 0.02063768 | 0.02252404 | 0.00921356 | 0.00190828 | 0.03480102 | 0.03833774 | 0.01227179 | 0.00411046 | 0.00100742 | 0.00112869 | 0.00182884 | 0.00061728 | 0.00123243 | 0.00138796 | 0.00206492 | 0.00109696 |
| Number of Bus Stops within and surrounding a green space (within 100m buffer) | # | 0.98865585 | 1.05741298 | 0.44310869 | 0.47388176 | 1.6525561 | 1.77257388 | 0.75987043 | 0.77681598 | 0.01128652 | 0.01254544 | 0.02043372 | 0.02116705 | 0.01535371 | 0.01711694 | 0.02780054 | 0.02939432 |
| Number of ferry wharves within and surrounding a green space (within 100m buffer) | # | 0.00803023 | 0.00900485 | 0.0008614 | 0 | 0.01187675 | 0.01339301 | 0.00122753 | 0 | 0.00066888 | 0.00075457 | 0.00039113 | 0 | 0.00081552 | 0.00092518 | 0.00043763 | 0 |
| Area of GS which is greater than 6 degrees of Slope | m2 | 765.771495 | 717.222617 | 1278.88895 | 961.78577 | 1354.08694 | 1274.76389 | 2250.64528 | 1533.86372 | 8.9416175 | 9.10778339 | 49.5278973 | 40.1682987 | 13.7519885 | 14.1203459 | 74.081724 | 54.4238392 |
| Length of Main Roads within 100 metres of the Green Space | m | 112.106454 | 123.719855 | 26.0151777 | 17.2875276 | 190.47239 | 211.185541 | 45.1324104 | 28.0526127 | 2.21981952 | 2.49360187 | 1.90005017 | 1.35928902 | 3.14614589 | 3.55130015 | 2.57135511 | 1.70725202 |
| **Access indices – calculated** | | | | | | | | | | | | | | | | | |
| Ratio of Area(A) to Perimeter (P) | m2 | 26.337376 | 23.0539436 | 58.1448286 | 43.4372704 | 27.5452173 | 24.2426951 | 58.316257 | 43.6159162 | 0.16683336 | 0.15204141 | 1.12636016 | 0.83166285 | 0.15159927 | 0.13916098 | 0.98118653 | 0.72118731 |
| Ratio of Perimeter (P) to perimeter standardised by area (A) | m | 1.64155777 | 1.67342964 | 1.40618097 | 1.38017599 | 1.62899655 | 1.66127066 | 1.40018125 | 1.37896754 | 0.00352932 | 0.00391886 | 0.00521731 | 0.00498544 | 0.00302468 | 0.00337093 | 0.00450423 | 0.00430881 |
| Total number of transport stops/stations/wharves within and surrounding a green space (within 100m buffer) | # | 1.01732376 | 1.08894187 | 0.45318365 | 0.47579004 | 1.69923387 | 1.82430462 | 0.77336975 | 0.78092644 | 0.01165057 | 0.01295621 | 0.02064359 | 0.0212248 | 0.01580097 | 0.01762257 | 0.02806582 | 0.02948812 |
| Pathway Length per Area of Green Space | Ratio | 0.0076286 | 0.00814436 | 0.00377799 | 0.00345301 | 0.00726163 | 0.00775758 | 0.00370654 | 0.00346988 | 0.00010503 | 0.00011735 | 0.00016591 | 0.000149 | 7.5793E-05 | 8.4734E-05 | 0.00013715 | 0.00012971 |
| Bikeway Length per Area of Green Space | Ratio | 0.00564622 | 0.00557418 | 0.005999 | 0.00647007 | 0.00554235 | 0.00543538 | 0.00613797 | 0.00658157 | 7.2267E-05 | 7.9582E-05 | 0.00019333 | 0.00019894 | 5.7656E-05 | 6.3255E-05 | 0.00017187 | 0.00017746 |
| Trees over Pathway per Area of Green Space | Ratio | 0.00242948 | 0.00265394 | 0.00069301 | 0.00061085 | 0.00231387 | 0.00253264 | 0.00068662 | 0.00062995 | 4.0335E-05 | 4.5122E-05 | 3.8026E-05 | 4.6183E-05 | 2.9388E-05 | 3.2934E-05 | 3.1821E-05 | 4.0376E-05 |
| Trees over Bikeway per Area of Green Space | Ratio | 0.00167027 | 0.00179708 | 0.00050226 | 0.00088693 | 0.00159981 | 0.00172211 | 0.00050517 | 0.00089933 | 2.8422E-05 | 3.1737E-05 | 3.0402E-05 | 5.2609E-05 | 2.2285E-05 | 2.4954E-05 | 2.4665E-05 | 4.5745E-05 |
| Percentage of GS which is greater than 6 degrees of Slope | % | 27.2020284 | 26.6942396 | 31.9559144 | 30.050341 | 26.5468492 | 25.9685443 | 31.5583805 | 29.8327406 | 0.16359189 | 0.17451261 | 0.63339769 | 0.67833701 | 0.14255536 | 0.15202372 | 0.55232566 | 0.59307257 |
| Pathway Length > 6 degrees per Area of GS | Ratio | 0.00205728 | 0.00219475 | 0.00106732 | 0.00089946 | 0.00190565 | 0.00203526 | 0.00100409 | 0.00088211 | 4.7901E-05 | 5.3693E-05 | 6.357E-05 | 7.1769E-05 | 3.4072E-05 | 3.8301E-05 | 4.7157E-05 | 6.1874E-05 |
| Bikeway Length > 6 degrees slope per Area of GS | Ratio | 0.00114036 | 0.00115493 | 0.00095004 | 0.00112878 | 0.00112958 | 0.00114558 | 0.00089382 | 0.00116443 | 2.5546E-05 | 2.8279E-05 | 5.6996E-05 | 6.9415E-05 | 2.1296E-05 | 2.3686E-05 | 4.1148E-05 | 6.3155E-05 |
| **Amenities/Activities indices – original sourced** | | | | | | | | | | | | | | | | | |
| Number of public toilet facilities within and surrounding a green space (within 100m buffer) | # | 0.1708264 | 0.18465623 | 0.06130181 | 0.06701558 | 0.29633146 | 0.32174984 | 0.10181729 | 0.11791244 | 0.00430746 | 0.00483878 | 0.00460781 | 0.00487078 | 0.00605891 | 0.0068399 | 0.00580603 | 0.00760365 |
| Number of cafes within and surrounding a green space (within 100m buffer) | # | 0.25053355 | 0.27568865 | 0.09010052 | 1.13E-02 | 0.41351056 | 0.45859194 | 0.12724246 | 2.11E-02 | 0.00691648 | 0.00774691 | 0.01209731 | 1.67E-03 | 0.01003749 | 0.01131989 | 0.01469097 | 2.41E-03 |
| Number of hotels/bars within and surrounding a green space (within 100m buffer) | # | 0.08132609 | 0.09047412 | 0.01861232 | 3.46E-06 | 0.13347253 | 0.14942576 | 0.02800377 | 7.76E-06 | 0.00345111 | 0.00388947 | 0.00250657 | 3.43E-06 | 0.00467666 | 0.00529979 | 0.00302752 | 5.86E-06 |
| Number of restaurants/takeaways within and surrounding a green space (within 100m buffer) | # | 1.00825612 | 1.12386163 | 0.16167893 | 0.05083749 | 1.61951209 | 1.8169704 | 0.21505451 | 0.09564932 | 0.0328915 | 0.03706239 | 0.0182748 | 0.00545453 | 0.04393156 | 0.04976622 | 0.02048157 | 0.00819442 |
| Number of restaurants within and surrounding a green space (within 100m buffer) | # | 0.7059039 | 0.78587504 | 0.12762778 | 0.03404355 | 1.12775025 | 1.26411097 | 0.16565435 | 0.06532206 | 0.02402402 | 0.02706594 | 0.0162034 | 0.00432866 | 0.03207603 | 0.03633385 | 0.01836061 | 0.00657929 |
| Number of takeaways within and surrounding a green space (within 100m buffer) | # | 0.30235222 | 0.33798659 | 0.03405115 | 0.01679393 | 0.49176184 | 0.55285943 | 0.04940016 | 0.03032726 | 0.00966692 | 0.01089226 | 0.00487345 | 0.00202208 | 0.01277031 | 0.01446436 | 0.00511832 | 0.00279327 |
| Number of supermarkets/greengrocers within and surrounding a green space (within 100m buffer) | # | 0.12979073 | 0.14367929 | 0.02840177 | 0.01435704 | 0.21284758 | 0.23690889 | 0.04298797 | 0.02550069 | 0.00403199 | 0.00453729 | 0.00361015 | 0.00221148 | 0.00514212 | 0.00581272 | 0.00453726 | 0.00375493 |
| Presence of parkrun event | 1/0 | 0.01856413 | 0.01665479 | 0.04133767 | 0.02294686 | 0.02269396 | 0.02060495 | 0.04391892 | 0.03058252 | 0.00073981 | 0.00074535 | 0.00429126 | 0.00368063 | 0.00074867 | 0.00076102 | 0.00397089 | 0.00379458 |
| **Beach/Coast indices – original sourced** | | | | | | | | | | | | | | | | | |
| Presence of coastline | 1/0 | 0.13832983 | 0.12743801 | 0.22944728 | 0.21376812 | 0.1489765 | 0.13708891 | 0.23423423 | 0.23980583 | 0.00189225 | 0.00194216 | 0.00906403 | 0.01007738 | 0.00179 | 0.00184253 | 0.00820706 | 0.00940944 |
| Area of beach within and surrounding a green space (within 100m buffer) | m2 | 218.379382 | 211.830082 | 246.604652 | 298.277266 | 385.782948 | 377.156111 | 381.994866 | 536.60927 | 11.5880271 | 12.6527052 | 35.0384323 | 38.077518 | 16.7613711 | 18.4467372 | 43.9711346 | 55.2479352 |
| **Biodiversity indices – original sourced** | | | | | | | | | | | | | | | | | |
| Number of bird species/species habitat likely to occur within green space | # | 30.273085 | 29.8524473 | 35.3971203 | 31.0996377 | 30.4630781 | 30.0366183 | 35.0927177 | 31.6898058 | 0.07379952 | 0.0771318 | 0.32509488 | 0.33768265 | 0.06825259 | 0.07164269 | 0.28948801 | 0.30735915 |
| Number of fish species/species habitat likely to occur within green space | # | 1.68879543 | 1.65377701 | 1.66604738 | 2.34178744 | 1.70970432 | 1.67258796 | 1.63400901 | 2.43543689 | 0.01753468 | 0.01879701 | 0.05630556 | 0.0815364 | 0.01607522 | 0.01727501 | 0.05050292 | 0.07366109 |
| Number of flora species/species habitat likely to occur within green space | # | 7.63817963 | 7.37325735 | 9.65536461 | 9.73188406 | 7.74811726 | 7.47592263 | 9.74512012 | 9.76990291 | 0.01486937 | 0.01561373 | 0.03969416 | 0.05318688 | 0.01380102 | 0.01457947 | 0.03522382 | 0.04782883 |
| Number of frog species/species habitat likely to occur within green space | # | 1.69927906 | 1.71737729 | 1.10404087 | 2.15096618 | 1.7026788 | 1.72094358 | 1.11148649 | 2.15825243 | 0.00302915 | 0.00310577 | 0.00852462 | 0.01110952 | 0.00277429 | 0.00283987 | 0.00808638 | 0.01000301 |
| Number of mammal species/species habitat likely to occur within green space | # | 7.58401923 | 7.52443947 | 7.97213191 | 8.14009662 | 7.5794794 | 7.51185215 | 7.96358859 | 8.22669903 | 0.01063888 | 0.01123573 | 0.03691649 | 0.05555755 | 0.00978514 | 0.01034664 | 0.03314885 | 0.05028044 |
| Number of other animal species/species habitat likely to occur within green space | # | 0.19486332 | 0.22004003 | 0 | 0 | 0.20103614 | 0.22829019 | 0 | 0 | 0.00227289 | 0.00252968 | 0 | 0 | 0.0021068 | 0.00235481 | 0 | 0 |
| Number of reptile species/species habitat likely to occur within green space | # | 1.37551817 | 1.29781893 | 1.82071528 | 2.17995169 | 1.40103614 | 1.32195948 | 1.75863363 | 2.27621359 | 0.01203953 | 0.01255952 | 0.05186383 | 0.05856571 | 0.01111792 | 0.01163852 | 0.04626645 | 0.05262355 |
| Area of land with biodiversity value within green space | m2 | 209.58551 | 197.992377 | 430.195828 | 129.152629 | 373.256087 | 354.973436 | 724.012208 | 228.917713 | 4.74378161 | 4.78986443 | 31.2253271 | 11.48586 | 7.25276404 | 7.46922116 | 42.8061902 | 17.1040762 |
| Area of land with biodiversity value outside of green space within 100m | m2 | 746.014404 | 763.464619 | 941.993453 | 180.559869 | 1229.60875 | 1265.23853 | 1478.87254 | 304.56321 | 19.7888739 | 21.0729182 | 100.232833 | 17.6928331 | 26.2766712 | 28.1452092 | 127.191956 | 24.3234372 |
| **Biodiversity indices – calculated** | | | | | | | | | | | | | | | | | |
| Total number of species/species habitat likely to occur within green space | # | 50.4537399 | 49.6391574 | 57.6154203 | 55.6443237 | 50.8051301 | 49.9681743 | 57.3055556 | 56.5563107 | 0.11113475 | 0.11726431 | 0.43431617 | 0.50091846 | 0.10258585 | 0.10861603 | 0.38879575 | 0.45435693 |
| Shannon biodiversity index | unit | 1.1115352 | 1.10805193 | 1.08201219 | 1.21192979 | 1.11367789 | 1.11009193 | 1.08241337 | 1.21476757 | 0.00070803 | 0.00075452 | 0.00239762 | 0.00235555 | 0.0006459 | 0.00068934 | 0.00215403 | 0.00212857 |
| Simpson biodiversity index (range 0 to 1=most diverse) | unit | 0.57782607 | 0.57581465 | 0.56973134 | 0.62415847 | 0.57850689 | 0.57639481 | 0.57106568 | 0.62385686 | 0.00029816 | 0.00031258 | 0.00124922 | 0.0009648 | 0.00027178 | 0.0002857 | 0.00110411 | 0.00088913 |
| Percentage of land with biodiversity value within green space | % | 7.14525304 | 7.19447672 | 9.04729511 | 3.79606229 | 7.28770806 | 7.39605653 | 8.58908958 | 3.77198415 | 0.10116758 | 0.10826881 | 0.44885771 | 0.26645266 | 0.09032784 | 0.09709221 | 0.38805789 | 0.23295726 |
| Percentage of land with biodiversity value outside of green space within 100m | % | 2.70939535 | 2.60200973 | 4.88665588 | 1.79043001 | 2.68619301 | 2.59230407 | 4.62265145 | 1.77013905 | 0.03894246 | 0.03940032 | 0.2397449 | 0.13926829 | 0.03482369 | 0.03534592 | 0.20595457 | 0.12742148 |
| **Incivilities indices – original sourced** | | | | | | | | | | | | | | | | | |
| Density of alcohol-related assaults | Score 0-3 | 0.48690297 | 0.50602083 | 0.44449605 | 0.20169082 | 0.52628254 | 0.54910176 | 0.45457958 | 0.23300971 | 0.00541835 | 0.00584571 | 0.02080563 | 0.01575898 | 0.00510564 | 0.00553163 | 0.01881918 | 0.01488806 |
| Density of malicious damage | Score 0-3 | 1.24505858 | 1.25606323 | 1.25220622 | 1.03985507 | 1.32317412 | 1.3341847 | 1.31531532 | 1.14708738 | 0.0069043 | 0.00735759 | 0.02737474 | 0.02854639 | 0.00637963 | 0.0068091 | 0.02494706 | 0.02635642 |
| Density of non-domestic violence assaults | Score 0-3 | 0.57693001 | 0.5987246 | 0.54342778 | 0.23248792 | 0.62787465 | 0.65255696 | 0.57394895 | 0.28009709 | 0.00576895 | 0.00621786 | 0.02241641 | 0.01642622 | 0.00545322 | 0.00589818 | 0.02051403 | 0.015789 |
| Density of robberies | Score 0-3 | 0.40919195 | 0.42172925 | 0.41058987 | 0.18417874 | 0.46499874 | 0.48060036 | 0.44331832 | 0.22912621 | 0.00492941 | 0.00530028 | 0.01980336 | 0.01487999 | 0.00474656 | 0.0051178 | 0.01834483 | 0.01505887 |
| Density of steal from persons | Score 0-3 | 0.31486933 | 0.33340117 | 0.26103112 | 0.05495169 | 0.34584281 | 0.36669919 | 0.28003003 | 0.07815534 | 0.00448621 | 0.00489118 | 0.01627542 | 0.00754453 | 0.00427778 | 0.00467756 | 0.01519094 | 0.00804348 |
| **Incivilities indices – calculated** | | | | | | | | | | | | | | | | | |
| Mean level of incivilities | Score 0-3 | 0.60659057 | 0.62318782 | 0.58235021 | 0.34263285 | 0.65763457 | 0.67662859 | 0.61343844 | 0.39349515 | 0.00460929 | 0.00498068 | 0.01760718 | 0.01242445 | 0.00436166 | 0.0047304 | 0.0160689 | 0.01212559 |
| **Land cover indices – original sourced** | | | | | | | | | | | | | | | | | |
| Area of bare earth within green space | m2 | 368.089821 | 351.882683 | 573.368618 | 390.488072 | 662.647921 | 639.484519 | 950.489237 | 683.037133 | 5.45161216 | 5.49851541 | 32.9135975 | 23.8253254 | 8.56387771 | 8.85218182 | 44.7517965 | 35.2234012 |
| Area of buildings within green space | m2 | 63.7276531 | 63.0654731 | 80.556294 | 53.6708821 | 115.505584 | 115.106107 | 139.32959 | 91.4957672 | 1.35038777 | 1.46645948 | 4.78807464 | 4.02636668 | 1.88785712 | 2.04696939 | 7.19629135 | 5.36822684 |
| Area of built up area within green space | m2 | 121.989994 | 118.477117 | 134.744147 | 168.098794 | 213.963928 | 208.059276 | 231.678347 | 291.057913 | 1.76292367 | 1.83354233 | 7.65932432 | 9.49153615 | 2.55074283 | 2.62273714 | 11.4149628 | 14.5499316 |
| Area of open grass within green space | m2 | 781.700891 | 699.422605 | 1478.61356 | 1344.11164 | 1435.01279 | 1298.08425 | 2598.6865 | 2250.321 | 8.3115414 | 8.22383018 | 46.5990136 | 46.7271508 | 13.2085953 | 13.069472 | 74.7919376 | 69.5976492 |
| Area of other vegetation within green space | m2 | 470.358293 | 430.802783 | 798.089747 | 750.241582 | 839.725201 | 774.387841 | 1394.83513 | 1228.95869 | 4.83800768 | 4.86400314 | 24.3849798 | 28.492062 | 7.51806391 | 7.62295734 | 36.0858589 | 41.8743409 |
| Area of roads/paths within green space | m2 | 127.796291 | 83.7264275 | 570.323289 | 339.030346 | 222.253941 | 145.761734 | 954.429525 | 571.64105 | 2.36689139 | 1.58339109 | 25.4757282 | 13.2891511 | 3.42428957 | 2.215237 | 35.43175 | 19.6696384 |
| Area of shadow within green space | m2 | 30.5274127 | 34.3569548 | 0.5910065 | 1.10059689 | 54.8105931 | 62.0421787 | 1.0854734 | 1.76463363 | 0.61889251 | 0.69543452 | 0.06289734 | 0.17799523 | 0.97924325 | 1.10597075 | 0.09228709 | 0.24412413 |
| Area of swimming pools within green space | m2 | 3.75581726 | 3.75873953 | 3.50610687 | 4.02823333 | 7.10170589 | 6.95367402 | 6.61786981 | 10.2331128 | 0.16675709 | 0.17925582 | 0.5265019 | 0.76453633 | 0.25851174 | 0.25397793 | 0.81377403 | 2.25744831 |
| Area of tree canopy within green space | m2 | 897.933971 | 903.695564 | 1009.72968 | 649.813036 | 1603.07669 | 1619.80444 | 1794.30584 | 1072.83536 | 8.55974527 | 9.23278336 | 33.4478252 | 25.5287268 | 13.380416 | 14.524737 | 50.2632999 | 36.1155026 |
| Area of water within green space | m2 | 35.8246616 | 27.5821888 | 109.779156 | 86.7858966 | 66.0384047 | 52.0428548 | 186.097919 | 147.925206 | 1.38389937 | 1.32448803 | 9.32352714 | 8.07367148 | 2.11502561 | 2.05194263 | 13.4203909 | 11.4758516 |
| **Land cover indices - calculated** | | | | | | | | | | | | | | | | | |
| Percentage of bare earth within green space | % | 10.588779 | 10.8606629 | 8.52543841 | 8.41894567 | 10.697343 | 11.0012497 | 8.33433412 | 8.60399966 | 0.0832132 | 0.09050809 | 0.22762396 | 0.32707869 | 0.07378037 | 0.08049844 | 0.19579667 | 0.28911176 |
| Percentage of buildings within green space | % | 2.34230574 | 2.47380608 | 1.36782315 | 1.2623178 | 2.26896639 | 2.39099206 | 1.4214936 | 1.2975503 | 0.03001079 | 0.03335891 | 0.06135484 | 0.05591765 | 0.02439356 | 0.02715603 | 0.05491532 | 0.04914154 |
| Percentage of built up area within green space | % | 4.70794854 | 4.97588728 | 1.98145779 | 3.47044909 | 4.56427454 | 4.81878772 | 1.97368661 | 3.6012133 | 0.04582959 | 0.0508663 | 0.06414154 | 0.11792881 | 0.03823757 | 0.04253521 | 0.05445131 | 0.10712062 |
| Percentage of open grass within green space | % | 25.2080558 | 24.2136735 | 31.6965711 | 34.5189752 | 25.6710062 | 24.6774512 | 31.8738751 | 34.4812968 | 0.12519475 | 0.13212946 | 0.48777244 | 0.55047971 | 0.11077604 | 0.11715554 | 0.42622399 | 0.47618586 |
| Percentage of other vegetation within green space | % | 15.905578 | 15.534903 | 18.2323567 | 19.4958987 | 16.0203104 | 15.6574803 | 18.1712916 | 19.3852063 | 0.07065614 | 0.07584425 | 0.25121264 | 0.27208695 | 0.06178058 | 0.06648555 | 0.21665887 | 0.23548227 |
| Percentage of roads/paths within green space | % | 3.28715961 | 2.60434845 | 9.32670428 | 7.62175054 | 3.36310109 | 2.64850412 | 9.45929361 | 7.5879514 | 0.03583217 | 0.03525799 | 0.18531956 | 0.14823152 | 0.03159374 | 0.03085738 | 0.16005874 | 0.12970985 |
| Percentage of shadow within green space | % | 1.08777828 | 1.22506423 | 0.01438271 | 0.03311085 | 1.04983548 | 1.18897998 | 0.0143657 | 0.03140547 | 0.01627969 | 0.01822333 | 0.00113696 | 0.00320319 | 0.01263588 | 0.01418106 | 0.00095253 | 0.00269817 |
| Percentage of swimming pools within green space | % | 0.11725807 | 0.12180577 | 0.07447112 | 0.09171849 | 0.12249088 | 0.12782227 | 0.08201449 | 0.08450496 | 0.00385008 | 0.0042215 | 0.00739827 | 0.0156866 | 0.00346921 | 0.00382581 | 0.00694577 | 0.01300408 |
| Percentage of tree canopy within green space | % | 34.9062516 | 36.2257436 | 26.3777741 | 22.4453159 | 34.406313 | 35.7397286 | 26.3049037 | 22.2939601 | 0.14775165 | 0.15898369 | 0.46939926 | 0.51752854 | 0.129951 | 0.14011137 | 0.41001728 | 0.45243 |
| Percentage of water within green space | % | 0.65861644 | 0.52336401 | 1.6232556 | 1.81835329 | 0.67726339 | 0.54168716 | 1.58388658 | 1.80170868 | 0.01781097 | 0.01708162 | 0.09764714 | 0.13409062 | 0.01579474 | 0.01520507 | 0.08336923 | 0.11418461 |
| **Land use indices – original sourced** | | | | | | | | | | | | | | | | | |
| Area of commercial land use surrounding green space (within 100m doughnut buffer) | m2 | 1810.37764 | 1942.6388 | 1044.26309 | 451.835055 | 3114.70861 | 3355.78996 | 1716.26609 | 845.160065 | 40.4119651 | 44.8110685 | 105.07452 | 56.1668614 | 58.8422905 | 65.5949347 | 145.266636 | 83.1677926 |
| Area of education land use surrounding green space (within 100m doughnut buffer) | m2 | 820.754394 | 874.084043 | 330.013044 | 509.374394 | 1430.21532 | 1525.92111 | 604.452883 | 879.180003 | 16.8045404 | 18.6467441 | 27.4688213 | 48.6962427 | 23.7601951 | 26.4658358 | 41.8209411 | 65.6606529 |
| Area of hospital/medical land use surrounding green space (within 100m doughnut buffer) | m2 | 118.910625 | 125.111715 | 111.543352 | 18.093837 | 207.759232 | 219.869216 | 178.686317 | 40.5095922 | 7.60210355 | 8.4299897 | 20.4265561 | 10.98823 | 10.5880424 | 11.8292767 | 24.7890207 | 16.8178248 |
| Area of industrial land use surrounding green space (within 100m doughnut buffer) | m2 | 339.989953 | 344.659318 | 280.462675 | 334.256063 | 606.884348 | 619.177932 | 482.404214 | 559.909995 | 13.2107382 | 14.3816829 | 33.8721207 | 55.1072834 | 19.1025822 | 20.8837993 | 49.3767333 | 75.923895 |
| Area of other land use surrounding green space (within 100m doughnut buffer) | m2 | 24.1045047 | 22.377726 | 21.8434599 | 57.7851756 | 40.9583894 | 37.0191401 | 42.0966258 | 106.120923 | 2.80817115 | 3.04585216 | 5.18889516 | 14.1590261 | 4.13415908 | 4.3475595 | 7.77064229 | 28.1990826 |
| Area of parkland land use surrounding green space (within 100m doughnut buffer) | m2 | 4456.3202 | 4675.90993 | 2725.57761 | 2797.24039 | 7841.91186 | 8282.55891 | 4491.92418 | 4720.34885 | 62.8453305 | 69.8787558 | 118.41981 | 142.415241 | 90.9799675 | 101.706312 | 157.524863 | 198.703528 |
| Area of primary production land use surrounding green space (within 100m doughnut buffer) | m2 | 28.9472675 | 26.2008058 | 11.9217411 | 99.9764915 | 54.3695102 | 51.5108251 | 21.3776131 | 145.390946 | 3.25301669 | 3.49700385 | 3.87175817 | 19.2926235 | 5.27964687 | 5.81734665 | 5.69452756 | 23.304905 |
| Area of residential land use surrounding green space (within 100m doughnut buffer) | m2 | 31852.6876 | 34297.9006 | 12174.2701 | 13906.014 | 52216.1408 | 56410.4542 | 20457.8446 | 22337.0409 | 251.121691 | 277.528492 | 386.508938 | 508.782899 | 353.413177 | 391.848766 | 526.397778 | 690.43475 |
| Area of transport land use surrounding green space (within 100m doughnut buffer) | m2 | 387.315219 | 437.164357 | 0.91594259 | 2.23985774 | 665.347203 | 755.143773 | 2.70100922 | 3.32690306 | 16.5179337 | 18.6321948 | 0.56946568 | 1.17840704 | 23.58005 | 26.7401722 | 1.59328371 | 1.38617932 |
| Area of water land use surrounding green space (within 100m doughnut buffer) | m2 | 22.5034188 | 25.1567345 | 3.48031669 | 0 | 32.9141156 | 37.0302694 | 4.52507016 | 0 | 3.75301208 | 4.23620094 | 1.52740003 | 0 | 4.44425643 | 5.04489964 | 1.58721286 | 0 |
| Area of tree canopy surrounding green space (within 100m doughnut buffer) | m2 | 2667.19748 | 2959.82006 | 369.670047 | 340.36851 | 4359.82822 | 4857.82952 | 636.772361 | 551.308892 | 32.5942645 | 36.3389582 | 16.6063936 | 14.5354453 | 46.9219852 | 52.5164721 | 22.668502 | 20.0748337 |
| Area of tree canopy ≤4m surrounding green space (within 100m doughnut buffer) | m2 | 199.872842 | 217.92159 | 49.1142956 | 68.1696386 | 329.726951 | 360.940639 | 83.2850014 | 108.071436 | 1.84737423 | 2.04273952 | 2.10255215 | 3.51810897 | 2.61566665 | 2.90158301 | 2.76968363 | 4.61587694 |
| Area of tree canopy >4m surrounding green space (within 100m doughnut buffer) | m2 | 2390.75311 | 2657.37235 | 308.198275 | 256.583438 | 3903.9137 | 4356.96766 | 532.710336 | 418.512308 | 30.5725884 | 34.1094679 | 14.4362888 | 10.8776397 | 43.9727865 | 49.2544963 | 19.8091046 | 15.2599221 |
| **Land use indices – calculated** | | | | | | | | | | | | | | | | | |
| Percentage of commercial land use surrounding green space (within 100m doughnut buffer) | % | 3.36385763 | 3.33482352 | 4.19739728 | 2.79703638 | 3.45085032 | 3.42554722 | 4.08622821 | 3.0571925 | 0.05107921 | 0.05332525 | 0.24421978 | 0.22731081 | 0.04621584 | 0.04838906 | 0.21114755 | 0.20800201 |
| Percentage of education land use surrounding green space (within 100m doughnut buffer) | % | 2.2418855 | 2.18223879 | 2.6372573 | 2.78971715 | 2.3112131 | 2.23946618 | 2.76187557 | 2.94205195 | 0.03114512 | 0.03254975 | 0.13416613 | 0.15961796 | 0.02815974 | 0.02944257 | 0.12045299 | 0.14120048 |
| Percentage of hospital/medical land use surrounding green space (within 100m doughnut buffer) | % | 0.28994831 | 0.2863058 | 0.47108935 | 0.11928894 | 0.29350436 | 0.29091281 | 0.46170804 | 0.11982016 | 0.01237545 | 0.0133057 | 0.0540633 | 0.02828748 | 0.01087737 | 0.01177388 | 0.0448859 | 0.02430197 |
| Percentage of industrial land use surrounding green space (within 100m doughnut buffer) | % | 1.1846159 | 1.08052855 | 1.97072389 | 2.01559936 | 1.25986755 | 1.14110827 | 2.16069828 | 2.10378639 | 0.03110017 | 0.03156971 | 0.15917886 | 0.17661445 | 0.028712 | 0.02890562 | 0.1469046 | 0.16752202 |
| Percentage of other land use surrounding green space (within 100m doughnut buffer) | % | 0.23458463 | 0.15635825 | 0.60096542 | 1.15087324 | 0.24934293 | 0.1651084 | 0.69059583 | 1.10358505 | 0.01527769 | 0.01345839 | 0.09567527 | 0.1441769 | 0.01457236 | 0.01288618 | 0.09335498 | 0.12546802 |
| Percentage of parkland land use surrounding green space (within 100m doughnut buffer) | % | 12.9870078 | 12.5194297 | 17.2567775 | 15.7598683 | 12.9479284 | 12.4912225 | 16.893721 | 15.5706378 | 0.08890216 | 0.09402821 | 0.35993184 | 0.38961833 | 0.079431 | 0.08437091 | 0.31239234 | 0.33647656 |
| Percentage of primary production land use surrounding green space (within 100m doughnut buffer) | % | 0.2038695 | 0.14986425 | 0.24277129 | 1.11472294 | 0.20541266 | 0.1517561 | 0.27874634 | 1.01820675 | 0.01430981 | 0.01312532 | 0.05504098 | 0.15004482 | 0.01292857 | 0.01199515 | 0.0540718 | 0.12359159 |
| Percentage of residential land use surrounding green space (within 100m doughnut buffer) | % | 78.7777841 | 79.4865941 | 72.5593393 | 74.2438976 | 78.5533145 | 79.2721698 | 72.6148427 | 74.0731477 | 0.1088372 | 0.11454153 | 0.43780036 | 0.50702973 | 0.0977912 | 0.10323491 | 0.38287152 | 0.44496262 |
| Percentage of transport land use surrounding green space (within 100m doughnut buffer) | % | 0.67336347 | 0.7593742 | 0.00662421 | 0.00899599 | 0.6889489 | 0.78088179 | 0.01023398 | 0.01157171 | 0.02027411 | 0.02284339 | 0.00304107 | 0.00413911 | 0.01807606 | 0.02047439 | 0.00306932 | 0.00398038 |
| Percentage of water land use surrounding green space (within 100m doughnut buffer) | % | 0.04308314 | 0.04448287 | 0.05705449 | 0 | 0.03961733 | 0.04182694 | 0.04135002 | 0 | 0.00512602 | 0.00548157 | 0.0254498 | 0 | 0.00422433 | 0.00464417 | 0.0157006 | 0 |
| Percentage of tree canopy to roads surrounding green space (within 100m doughnut buffer) | % | 17.7279407 | 18.867113 | 8.55762763 | 8.9648434 | 17.5780928 | 18.7410167 | 8.66490538 | 8.97009094 | 0.05929515 | 0.06233104 | 0.14037225 | 0.16533999 | 0.05353275 | 0.05630499 | 0.13422676 | 0.14497447 |
| Percentage of tree canopy ≤4m to roads surrounding green space (within 100m doughnut buffer) | % | 1.69949371 | 1.73606215 | 1.31281535 | 1.53870074 | 1.69918269 | 1.73881279 | 1.29148697 | 1.5412907 | 0.00503389 | 0.00502223 | 0.03197727 | 0.0201508 | 0.00448273 | 0.00449131 | 0.02702285 | 0.01784563 |
| Percentage of tree canopy >4m to roads surrounding green space (within 100m doughnut buffer) | % | 15.4241735 | 16.4878798 | 6.99066341 | 7.07278342 | 15.275999 | 16.3589645 | 7.11773629 | 7.07470998 | 0.05692377 | 0.06017995 | 0.12360468 | 0.15184063 | 0.05131107 | 0.05428438 | 0.1204874 | 0.13282915 |
| Percentage of tree canopy surrounding green space (within 100m doughnut buffer) | % | 4.92701684 | 5.30293943 | 1.99693144 | 1.90978212 | 4.85369383 | 5.23540649 | 2.02838694 | 1.897531 | 0.0197412 | 0.021035 | 0.0339924 | 0.03347239 | 0.01732702 | 0.01846718 | 0.03052271 | 0.0287455 |
| Percentage of tree canopy ≤4m surrounding green space (within 100m doughnut buffer) | % | 0.454129 | 0.47264384 | 0.28145636 | 0.34255243 | 0.45354896 | 0.47306836 | 0.28034385 | 0.33981589 | 0.0014371 | 0.00149995 | 0.0056516 | 0.00574177 | 0.00126929 | 0.00132532 | 0.00483788 | 0.0049434 |
| Percentage of tree canopy >4m surrounding green space (within 100m doughnut buffer) | % | 4.30803923 | 4.65326693 | 1.65420271 | 1.48885995 | 4.23603096 | 4.5856381 | 1.68569721 | 1.47985521 | 0.01876647 | 0.02008522 | 0.03091031 | 0.02887969 | 0.01645095 | 0.01761992 | 0.02777634 | 0.0248054 |

IRSD = Index of Relative Social Disadvantage; when reversed, the higher the score the greater the disadvantage.

IRSD scores were reversed calculating Max - X + Min, where Max and Min are scores in the original NSW-wise data, and X is a score to be reversed.

m=metres | sq m=square metres | number = number of items

Table S11. Mean values and standard errors of aggregated green space features and SEIFA IRSD score (reversed) – 800m and 1600m buffers.

|  |  | **Buffer 800m (Mean)** | | | | **Buffer 1600m (Mean)** | | | | **Buffer 800m (Standard Error)** | | | | **Buffer 1600m (Standard Error)** | | | |
| --- | --- | --- | --- | --- | --- | --- | --- | --- | --- | --- | --- | --- | --- | --- | --- | --- | --- |
| **Index description** | **Unit** | **Full set** | **Syd** | **New** | **Wol** | **Full set** | **Syd** | **New** | **Wol** | **Full set** | **Syd** | **New** | **Wol** | **Full set** | **Syd** | **New** | **Wol** |
| **SEIFA (IRSD) and general indices** | | | | | | | | | | | | | | | | | |
| Score IRSD reversed | NA | 475.889162 | 473.968919 | 479.498111 | 499.610717 | 475.889162 | 473.968919 | 479.498111 | 499.610717 | 0.49270387 | 0.54087343 | 1.42704435 | 1.74698755 | 0.49270387 | 0.54087343 | 1.42704435 | 1.74698755 |
| Quintile IRSD reversed | NA | 2.77500105 | 2.72218319 | 2.99505958 | 3.27998581 | 2.77500105 | 2.72218319 | 2.99505958 | 3.27998581 | 0.00667168 | 0.00726674 | 0.02026874 | 0.02445657 | 0.00667168 | 0.00726674 | 0.02026874 | 0.02445657 |
| N green spaces in MB buffer | # | 9.78464699 | 10.4954724 | 4.81315396 | 5.36417469 | 36.2497717 | 39.3206208 | 15.4213891 | 16.9461594 | 0.03488976 | 0.03849255 | 0.04668993 | 0.06296463 | 0.1053868 | 0.11319097 | 0.12022072 | 0.13773607 |
| Intersect of buffer with green space | m2 | 31117.5119 | 30190.7573 | 41717.9722 | 31781.886 | 211260.567 | 205976.019 | 279959.574 | 203899.138 | 146.996248 | 153.251501 | 675.5597 | 613.122063 | 680.779469 | 718.430124 | 2899.62614 | 2507.64806 |
| Green space area (A) | m2 | 319110.764 | 296459.71 | 570511.465 | 344866.401 | 948816.053 | 877099.763 | 1731948.57 | 1033164.54 | 4112.71202 | 4549.21569 | 13142.7637 | 8806.67629 | 6058.7812 | 6502.76201 | 23955.2301 | 14623.5927 |
| Green space perimeter (P) | m | 1570.22645 | 1662.86147 | 974.751757 | 929.264315 | 8680.26912 | 9167.74374 | 5585.99503 | 5354.02514 | 6.42867145 | 7.07388427 | 15.4940247 | 17.3584174 | 23.2103897 | 25.2007344 | 52.3475595 | 60.3469189 |
| **Access indices – original sourced** | | | | | | | | | | | | | | | | | |
| Length of pathways within green space | m | 271.059636 | 281.653534 | 232.327171 | 161.402568 | 1404.74926 | 1478.5937 | 1043.67493 | 767.909154 | 2.2645915 | 2.47687325 | 8.30326019 | 5.74751799 | 8.20456868 | 9.16374095 | 21.1233272 | 15.9466835 |
| Length of bikeways within green space | m | 228.916827 | 220.003507 | 315.743996 | 254.031847 | 1386.33053 | 1350.05614 | 1818.77446 | 1384.12005 | 1.92881445 | 2.05131707 | 8.05856595 | 7.40611156 | 7.37535761 | 7.95114036 | 28.8038747 | 24.1862019 |
| Length of pathways covered by trees within green space | m | 85.1381968 | 92.3508081 | 38.472125 | 30.7712536 | 460.909751 | 498.054204 | 207.734533 | 148.823275 | 0.87906747 | 0.98791189 | 1.44576491 | 1.67553602 | 3.32039894 | 3.69574389 | 4.84948189 | 4.22668279 |
| Length of bikeways covered by trees within green space | m | 64.400444 | 69.0548408 | 23.6382123 | 42.6088451 | 401.871784 | 429.375897 | 172.940615 | 222.06402 | 0.6830636 | 0.76259341 | 0.98383126 | 2.10771632 | 2.60668502 | 2.88206009 | 4.43878897 | 6.35244795 |
| Length of pathways greater than 6 degrees gradient within green space | m | 68.1197243 | 70.2361529 | 66.2926331 | 38.8753373 | 366.348478 | 382.338195 | 328.230704 | 175.095253 | 0.73902406 | 0.80373849 | 2.93193626 | 1.86788508 | 2.50269265 | 2.761658 | 8.39513913 | 4.78513862 |
| Length of bikeways greater than 6 degrees gradient within green space | m | 43.8720964 | 42.6061518 | 54.5422732 | 49.5104449 | 267.286598 | 265.203719 | 312.039245 | 242.666443 | 0.49544331 | 0.51241123 | 2.36522339 | 2.27025582 | 1.74032164 | 1.83205238 | 7.83889845 | 6.99200773 |
| Number of Railway Stations within and surrounding a green space (within 100m buffer) | # | 0.15580454 | 0.17582126 | 0.0269224 | 0.01756565 | 0.66540989 | 0.75249459 | 0.10562498 | 0.07985204 | 0.00287232 | 0.00327726 | 0.00344998 | 0.00210422 | 0.00725959 | 0.00825053 | 0.0076728 | 0.0046322 |
| Number of Bus Stops within and surrounding a green space (within 100m buffer) | # | 7.5659802 | 8.18372133 | 3.44592411 | 3.47627984 | 35.0632048 | 37.7222421 | 17.5438221 | 17.7111611 | 0.0443829 | 0.04967621 | 0.07233173 | 0.08221437 | 1.36E-01 | 0.15090589 | 0.21597539 | 0.23978249 |
| Number of ferry wharves within and surrounding a green space (within 100m buffer) | # | 0.04228153 | 0.04810013 | 0.00651138 | 0 | 0.14701873 | 0.16618933 | 0.03845633 | 0 | 0.00164918 | 0.00189166 | 0.00146098 | 0 | 3.39E-03 | 3.89E-03 | 0.00375581 | 0 |
| Area of GS which is greater than 6 degrees of Slope | m2 | 7776.38391 | 7421.80913 | 12259.0575 | 7506.31009 | 54705.7972 | 51763.2521 | 94277.7349 | 49369.2451 | 55.3197412 | 56.6058741 | 302.174087 | 193.693625 | 273.299598 | 270.640229 | 1673.40252 | 770.655007 |
| Length of Main Roads within 100 metres of the Green Space | m | 864.000952 | 966.137503 | 222.753081 | 138.349232 | 4262.81849 | 4732.27617 | 1350.6071 | 975.896149 | 9.38085581 | 10.6791299 | 6.52801732 | 5.52760365 | 3.09E+01 | 3.50E+01 | 23.2434195 | 20.3046044 |
| **Access indices – calculated** | | | | | | | | | | | | | | | | | |
| Ratio of Area(A) to Perimeter (P) | m2 | 33.3082791 | 29.8004193 | 64.1826102 | 47.2736057 | 43.897226 | 40.4934487 | 76.1264073 | 54.0014965 | 0.13770977 | 0.12811486 | 0.80662351 | 0.59064592 | 0.14970965 | 0.148518 | 0.75797817 | 0.47251531 |
| Ratio of Perimeter (P) to perimeter standardised by area (A) | m | 1.6199 | 1.65662606 | 1.38496195 | 1.36436838 | 1.6395362 | 1.67995919 | 1.37669424 | 1.37143831 | 0.00217946 | 0.00243476 | 0.0030533 | 0.00286375 | 0.00165229 | 0.00180805 | 0.00226083 | 0.00197009 |
| Total number of transport stops/stations/wharves within and surrounding a green space (within 100m buffer) | # | 7.76406627 | 8.40764272 | 3.47935789 | 3.49384549 | 35.8756334 | 38.640926 | 17.6879034 | 17.7910131 | 0.04541431 | 0.05082991 | 0.07275353 | 0.08221803 | 0.13927606 | 0.15410083 | 0.21680496 | 0.23939045 |
| Pathway Length per Area of Green Space | Ratio | 0.00702661 | 0.00751737 | 0.00373848 | 0.00379623 | 0.00613744 | 0.00656217 | 0.0033653 | 0.00333323 | 4.0182E-05 | 4.4483E-05 | 0.00010271 | 9.3281E-05 | 2.3421E-05 | 2.5605E-05 | 5.8592E-05 | 5.7156E-05 |
| Bikeway Length per Area of Green Space | Ratio | 0.00589958 | 0.00581119 | 0.00633646 | 0.00667358 | 0.00601365 | 0.00597848 | 0.00617987 | 0.00632411 | 3.6843E-05 | 0.00004015 | 0.00012754 | 0.00012476 | 2.3595E-05 | 2.5914E-05 | 7.7479E-05 | 7.3359E-05 |
| Trees over Pathway per Area of Green Space | Ratio | 0.00224246 | 0.00246113 | 0.000699 | 0.00075477 | 0.0020059 | 0.00218474 | 0.00067781 | 0.00063819 | 1.5597E-05 | 1.736E-05 | 2.2833E-05 | 3.1509E-05 | 9.94E-06 | 1.0845E-05 | 1.3805E-05 | 1.7698E-05 |
| Trees over Bikeway per Area of Green Space | Ratio | 0.00167315 | 0.00180772 | 0.00050203 | 0.00103386 | 0.00174267 | 0.00187746 | 0.00057311 | 0.00092051 | 1.3542E-05 | 1.5124E-05 | 1.6878E-05 | 3.6767E-05 | 8.71E-06 | 9.52E-06 | 1.2868E-05 | 2.2402E-05 |
| Percentage of GS which is greater than 6 degrees of Slope | % | 25.9377966 | 25.3109862 | 31.2561116 | 28.6860044 | 26.8065473 | 26.2535445 | 33.0500011 | 27.2894111 | 0.11132074 | 0.11857176 | 0.43148499 | 0.46392446 | 0.09189639 | 0.09829555 | 0.355464 | 0.34939654 |
| Pathway Length > 6 degrees per Area of GS | Ratio | 0.00178338 | 0.00190982 | 0.00097285 | 0.00090435 | 0.00167587 | 0.00179765 | 0.00095543 | 0.00075475 | 1.7179E-05 | 1.932E-05 | 3.293E-05 | 3.8448E-05 | 1.1906E-05 | 1.3407E-05 | 2.215E-05 | 1.899E-05 |
| Bikeway Length > 6 degrees slope per Area of GS | Ratio | 0.00118132 | 0.00120697 | 0.00088769 | 0.00116307 | 0.00117377 | 0.00120466 | 0.00092972 | 0.00101631 | 1.4463E-05 | 1.622E-05 | 2.9553E-05 | 4.0668E-05 | 6.50E-06 | 7.08E-06 | 2.1089E-05 | 2.3533E-05 |
| **Amenities/Activities indices – original sourced** | | | | | | | | | | | | | | | | | |
| Number of public toilet facilities within and surrounding a green space (within 100m buffer) | # | 1.45172075 | 1.59121255 | 0.47046533 | 0.59125002 | 6.97442381 | 7.63856704 | 2.2086558 | 3.122101 | 0.01645861 | 0.01873375 | 0.0156938 | 0.02177694 | 0.04627639 | 0.05220947 | 0.05153256 | 0.06056212 |
| Number of cafes within and surrounding a green space (within 100m buffer) | # | 1.6496601 | 1.86074362 | 0.36746993 | 0.096659 | 6.38119551 | 7.21107606 | 1.23240443 | 0.57168717 | 0.02856911 | 0.03266362 | 0.02580496 | 0.00599089 | 0.08918362 | 0.10205197 | 0.05172951 | 0.01993179 |
| Number of hotels/bars within and surrounding a green space (within 100m buffer) | # | 0.50327217 | 0.56928597 | 0.11646049 | 3.8329E-05 | 1.82870851 | 2.06034204 | 0.55902708 | 0.00038147 | 0.01314699 | 0.01508555 | 0.00702567 | 1.72E-05 | 0.04100607 | 0.04710963 | 0.02269656 | 7.7217E-05 |
| Number of restaurants/takeaways within and surrounding a green space (within 100m buffer) | # | 6.20291684 | 7.04811015 | 0.66489703 | 0.48478059 | 22.3656152 | 25.3675794 | 2.97594855 | 2.29512047 | 0.11187788 | 0.12813001 | 0.03531519 | 0.02923527 | 0.31217892 | 0.35727267 | 0.10615046 | 0.09095312 |
| Number of restaurants within and surrounding a green space (within 100m buffer) | # | 4.26125664 | 4.84480878 | 0.43020425 | 0.32241179 | 15.0615907 | 17.1038052 | 1.78964157 | 1.50819094 | 0.08191705 | 0.09385222 | 0.02861635 | 0.02340811 | 0.22854998 | 0.26177515 | 0.08165336 | 0.07720288 |
| Number of takeaways within and surrounding a green space (within 100m buffer) | # | 1.94166021 | 2.20330137 | 0.23469278 | 0.1623688 | 7.30402457 | 8.26377423 | 1.18630698 | 0.78692953 | 0.03136008 | 0.03587849 | 0.01148419 | 0.00927981 | 0.08568495 | 0.09782961 | 0.03247919 | 0.0208324 |
| Number of supermarkets/greengrocers within and surrounding a green space (within 100m buffer) | # | 0.82092283 | 0.92108818 | 0.1667185 | 0.14063661 | 3.0328372 | 3.38276917 | 0.72267009 | 0.75497083 | 0.01116431 | 0.01272082 | 0.01020747 | 0.01065587 | 0.02644057 | 0.02998851 | 0.02227143 | 0.02533397 |
| Presence of parkrun event | 1/0 | 0.04967023 | 0.0465446 | 0.07623318 | 0.06328645 | 0.13611164 | 0.13284539 | 0.17988957 | 0.12993539 | 0.00101032 | 0.00105072 | 0.00458902 | 0.00468486 | 0.0015804 | 0.00167927 | 0.00654877 | 0.00637128 |
| **Beach/Coast indices – original sourced** | | | | | | | | | | | | | | | | | |
| Presence of coastline | 1/0 | 0.21915883 | 0.20632867 | 0.28340807 | 0.33049593 | 0.32929756 | 0.31102516 | 0.41063644 | 0.49676956 | 0.00192368 | 0.00201838 | 0.00779308 | 0.00905102 | 0.00216596 | 0.00229033 | 0.00838767 | 0.00947433 |
| Area of beach within and surrounding a green space (within 100m buffer) | m2 | 1700.7993 | 1635.63305 | 1644.35106 | 2740.16725 | 6562.66498 | 6184.30929 | 6846.96001 | 11759.4802 | 46.3679993 | 50.4291628 | 122.224702 | 208.882297 | 116.046704 | 124.340336 | 341.016147 | 577.599191 |
| **Biodiversity indices – original sourced** | | | | | | | | | | | | | | | | | |
| Number of bird species/species habitat likely to occur within green space | # | 32.3681479 | 31.948281 | 36.4657698 | 33.5418209 | 36.2725631 | 35.8023842 | 40.4957861 | 37.9508256 | 0.06744883 | 0.07189729 | 0.26142689 | 0.27325699 | 0.07343684 | 0.07939925 | 0.26050838 | 0.26637082 |
| Number of fish species/species habitat likely to occur within green space | # | 2.06928317 | 2.03639485 | 1.83198804 | 2.85233161 | 2.72087343 | 2.67641731 | 2.31793083 | 3.87042355 | 0.01577123 | 0.01710413 | 0.04706502 | 0.06677764 | 0.01672928 | 0.01815361 | 0.05075262 | 0.0676528 |
| Number of flora species/species habitat likely to occur within green space | # | 8.24309655 | 7.96340614 | 10.1850523 | 10 | 8.99558189 | 8.72965828 | 10.7776809 | 10.6938263 | 0.01322204 | 0.0142185 | 0.02963925 | 0.04264773 | 0.01378552 | 0.01507432 | 0.02630861 | 0.0392247 |
| Number of frog species/species habitat likely to occur within green space | # | 1.73179803 | 1.7455346 | 1.14618834 | 2.25240563 | 1.79838144 | 1.80546852 | 1.20226678 | 2.43072505 | 0.00252721 | 0.00253406 | 0.00852808 | 0.00892903 | 0.00241591 | 0.00228128 | 0.01006534 | 0.00958697 |
| Number of mammal species/species habitat likely to occur within green space | # | 7.71240134 | 7.6210508 | 8.09058296 | 8.60325685 | 7.96323201 | 7.83095075 | 8.43214182 | 9.32376167 | 0.00966205 | 0.01029471 | 0.03104339 | 0.04481698 | 0.01046024 | 0.01112745 | 0.03387901 | 0.04492256 |
| Number of other animal species/species habitat likely to occur within green space | # | 0.2513569 | 0.28916862 | 0 | 0 | 0.33804881 | 0.38957701 | 0 | 0 | 0.00212669 | 0.00239037 | 0 | 0 | 0.00234202 | 0.00260639 | 0 | 0 |
| Number of reptile species/species habitat likely to occur within green space | # | 1.67940318 | 1.59356187 | 1.91330344 | 2.6669134 | 2.21370462 | 2.11360521 | 2.32054635 | 3.54953338 | 0.01083859 | 0.01148371 | 0.04202481 | 0.04573927 | 0.011387 | 0.01219388 | 0.04251469 | 0.04002134 |
| Area of land with biodiversity value within green space | m2 | 2368.60324 | 2378.87836 | 3255.27661 | 1118.06228 | 19436.7201 | 20142.404 | 20995.2922 | 7164.05589 | 31.2471302 | 33.4821725 | 148.398213 | 56.6377543 | 164.691048 | 178.307369 | 719.165261 | 250.388442 |
| Area of land with biodiversity value outside of green space within 100m | m2 | 6578.94786 | 6975.61621 | 6003.23735 | 1390.37553 | 45541.3257 | 49449.4606 | 29917.4395 | 7532.23277 | 90.9951646 | 101.169808 | 306.778799 | 70.1761862 | 403.770686 | 454.333333 | 962.357212 | 228.898745 |
| **Biodiversity indices – calculated** | | | | | | | | | | | | | | | | | |
| Total number of species/species habitat likely to occur within green space | # | 54.0554871 | 53.1973979 | 59.6328849 | 59.9167283 | 60.3023854 | 59.3480613 | 65.5463528 | 67.8190955 | 0.10092494 | 0.10827535 | 0.35365442 | 0.4078732 | 0.108275 | 0.11747156 | 0.35410241 | 0.39012942 |
| Shannon biodiversity index | unit | 1.12985851 | 1.12617774 | 1.08946618 | 1.23462236 | 1.15243083 | 1.14884435 | 1.09524678 | 1.2756487 | 0.00060066 | 0.00064236 | 0.00200709 | 0.0018175 | 0.000593 | 0.00062383 | 0.00222307 | 0.00149604 |
| Simpson biodiversity index (range 0 to 1=most diverse) | unit | 0.58152197 | 0.57923921 | 0.57216908 | 0.62706143 | 0.58297055 | 0.58088909 | 0.56719469 | 0.6329764 | 0.0002482 | 0.0002618 | 0.00098364 | 0.00074623 | 0.00024491 | 0.00025485 | 0.0010303 | 0.00071633 |
| Percentage of land with biodiversity value within green space | % | 8.14319712 | 8.43693983 | 8.12809591 | 3.79184399 | 9.74490719 | 10.1754526 | 9.21336799 | 4.08818975 | 0.07472178 | 0.08143562 | 0.29233771 | 0.17117931 | 0.06638309 | 0.07215342 | 0.26362847 | 0.14288489 |
| Percentage of land with biodiversity value outside of green space within 100m | % | 2.78938986 | 2.7311206 | 4.4599787 | 1.58812709 | 3.38746203 | 3.41027251 | 4.70463205 | 1.42614273 | 0.02854703 | 0.02925814 | 0.16125277 | 0.08707205 | 0.02520562 | 0.02619414 | 0.13629534 | 0.05841291 |
| **Incivilities indices – original sourced** | | | | | | | | | | | | | | | | | |
| Density of alcohol-related assaults | Score 0-3 | 0.82041302 | 0.86031643 | 0.6813154 | 0.39896373 | 1.36829584 | 1.42090963 | 1.11537344 | 0.9091888 | 0.00550715 | 0.00600714 | 0.01905697 | 0.01604961 | 0.00603745 | 0.00653616 | 0.02149514 | 0.02000136 |
| Density of malicious damage | Score 0-3 | 1.74418856 | 1.76483905 | 1.63318386 | 1.57438934 | 2.30903375 | 2.32637325 | 2.132229 | 2.27315147 | 0.00577474 | 0.00617886 | 0.02224459 | 0.02327788 | 0.00474533 | 0.00504939 | 0.01965922 | 0.01851953 |
| Density of non-domestic violence assaults | Score 0-3 | 0.95982268 | 1.00134335 | 0.84215247 | 0.48778682 | 1.56838505 | 1.61901498 | 1.39465272 | 1.04055994 | 0.00576865 | 0.00626855 | 0.02081994 | 0.01698365 | 0.00603269 | 0.00649288 | 0.02231064 | 0.02102666 |
| Density of robberies | Score 0-3 | 0.80847659 | 0.83971839 | 0.72705531 | 0.44448557 | 1.44701459 | 1.48780965 | 1.28451032 | 1.04953338 | 0.00527899 | 0.0057066 | 0.01961074 | 0.01757863 | 0.00580655 | 0.00619306 | 0.02218991 | 0.02352262 |
| Density of steal from persons | Score 0-3 | 0.56166072 | 0.59759192 | 0.44155456 | 0.17579571 | 1.00484292 | 1.06095173 | 0.83870968 | 0.38729361 | 0.00477564 | 0.0052369 | 0.01669264 | 0.01072518 | 0.00567964 | 0.00613099 | 0.02139722 | 0.01629849 |
| **Incivilities indices – calculated** | | | | | | | | | | | | | | | | | |
| Mean level of incivilities | Score 0-3 | 0.97891231 | 1.01276183 | 0.86505232 | 0.61628423 | 1.53951443 | 1.58301185 | 1.35309503 | 1.13194544 | 0.00460809 | 0.00501979 | 0.01634579 | 0.01314707 | 0.00485068 | 0.00524794 | 0.01756922 | 0.01554399 |
| **Land cover indices – original sourced** | | | | | | | | | | | | | | | | | |
| Area of bare earth within green space | m2 | 3890.38956 | 3867.66711 | 4502.44208 | 3476.59939 | 25084.8685 | 25423.374 | 24993.2997 | 20263.9904 | 35.2548192 | 37.5352789 | 150.985499 | 131.217144 | 166.712779 | 184.211879 | 496.273262 | 503.19104 |
| Area of buildings within green space | m2 | 691.223924 | 699.717643 | 715.643998 | 535.240064 | 4639.47945 | 4687.27517 | 4965.13747 | 3548.33492 | 6.70826131 | 7.24465549 | 26.2099361 | 22.4943329 | 34.0154576 | 37.2590634 | 126.893378 | 83.8062538 |
| Area of built up area within green space | m2 | 1165.49826 | 1148.79203 | 1072.20297 | 1527.83562 | 6926.03665 | 6825.46298 | 6495.34632 | 8913.45983 | 8.85806585 | 9.05210382 | 34.5704616 | 54.3817674 | 37.4442018 | 38.0075728 | 173.734984 | 205.660572 |
| Area of open grass within green space | m2 | 9108.92579 | 8580.03672 | 13379.0634 | 11717.1174 | 64509.1656 | 61679.4025 | 86550.5203 | 79017.8814 | 56.3804399 | 57.4959953 | 271.409189 | 273.158864 | 271.561586 | 278.226331 | 1315.54229 | 1206.97745 |
| Area of other vegetation within green space | m2 | 4785.82029 | 4523.00799 | 7150.09401 | 5784.48677 | 33015.9792 | 30965.1325 | 54459.1775 | 36901.327 | 29.3641607 | 30.2327546 | 140.032543 | 130.570188 | 152.747536 | 153.749497 | 839.429695 | 532.893729 |
| Area of roads/paths within green space | m2 | 1202.04602 | 843.17798 | 4364.71588 | 2646.35675 | 7800.61415 | 5918.06844 | 23989.9289 | 15602.9349 | 11.7935825 | 8.45114576 | 99.7502844 | 61.1843995 | 52.294792 | 41.4866277 | 360.25294 | 235.300029 |
| Area of shadow within green space | m2 | 314.513006 | 360.63606 | 6.89534169 | 7.59295265 | 2023.9113 | 2323.42944 | 61.3804147 | 39.1457422 | 4.18341071 | 4.76826168 | 0.4835251 | 0.54911636 | 20.0399116 | 22.704849 | 2.65835688 | 1.75537251 |
| Area of swimming pools within green space | m2 | 41.6235483 | 42.2955513 | 33.4218196 | 41.7156179 | 260.227415 | 267.807364 | 225.953344 | 191.339636 | 0.75422228 | 0.79491073 | 2.63116558 | 4.01128807 | 2.4029342 | 2.53034994 | 11.0953807 | 9.14493046 |
| Area of tree canopy within green space | m2 | 9132.52253 | 9399.10482 | 9212.49388 | 5079.07546 | 62221.8563 | 63092.3168 | 72737.9171 | 36791.4571 | 52.7868247 | 57.8712689 | 190.669048 | 113.459436 | 262.840735 | 281.370099 | 1198.16142 | 499.791306 |
| Area of water within green space | m2 | 418.234598 | 359.638884 | 861.552698 | 743.722894 | 2853.74462 | 2613.3486 | 4768.6945 | 4034.80584 | 8.86434433 | 9.21260014 | 43.9036692 | 33.8788795 | 37.4487009 | 40.5164027 | 131.398023 | 136.647242 |
| **Land cover indices - calculated** | | | | | | | | | | | | | | | | | |
| Percentage of bare earth within green space | % | 11.0725063 | 11.4535793 | 8.35656669 | 8.75064364 | 11.2215232 | 11.604139 | 8.42654671 | 9.03519626 | 0.06124606 | 0.06767029 | 0.14823977 | 0.21196099 | 0.0529354 | 0.05853755 | 0.12674242 | 0.179316 |
| Percentage of buildings within green space | % | 2.13306327 | 2.22771771 | 1.53288134 | 1.46502123 | 2.08463511 | 2.15618025 | 1.67379994 | 1.54028948 | 0.01408969 | 0.01553224 | 0.04329605 | 0.03867056 | 0.00886165 | 0.00948041 | 0.03781006 | 0.02692529 |
| Percentage of built up area within green space | % | 3.91018372 | 4.07050627 | 2.03882659 | 3.82741688 | 3.34095227 | 3.42098825 | 1.98884576 | 3.81395984 | 0.0221567 | 0.02454575 | 0.04107341 | 0.07766254 | 0.01331686 | 0.01432332 | 0.03526308 | 0.06028274 |
| Percentage of open grass within green space | % | 27.770331 | 27.0021972 | 31.109607 | 35.0703149 | 29.9632104 | 29.5793702 | 29.5473757 | 36.0596998 | 0.08688305 | 0.09251803 | 0.3335151 | 0.34374738 | 0.07193554 | 0.07716125 | 0.27966163 | 0.25307952 |
| Percentage of other vegetation within green space | % | 15.8331794 | 15.4256843 | 18.3749991 | 18.7606988 | 15.5642654 | 15.039465 | 19.6540309 | 18.2525482 | 0.04487236 | 0.04787846 | 0.17290667 | 0.17210602 | 0.03675511 | 0.03869676 | 0.16004366 | 0.11549991 |
| Percentage of roads/paths within green space | % | 3.43309614 | 2.63841739 | 9.576859 | 7.68610698 | 3.34168288 | 2.64703203 | 8.40083044 | 7.32947694 | 0.02237179 | 0.02022454 | 0.11712491 | 0.08697622 | 0.01533095 | 0.01266111 | 0.07723076 | 0.05903176 |
| Percentage of shadow within green space | % | 0.99950637 | 1.1463556 | 0.01497489 | 0.02859927 | 0.94464829 | 1.08475642 | 0.01807438 | 0.02657552 | 0.00835307 | 0.00938738 | 0.00070218 | 0.00178319 | 0.00673409 | 0.00750853 | 0.00060551 | 0.0015507 |
| Percentage of swimming pools within green space | % | 0.13437839 | 0.14140733 | 0.0847532 | 0.09097402 | 0.13399791 | 0.14294341 | 0.0704162 | 0.08074301 | 0.00237474 | 0.00261949 | 0.00550181 | 0.00916074 | 0.00145467 | 0.00162705 | 0.00296564 | 0.00421882 |
| Percentage of tree canopy within green space | % | 32.8340301 | 34.1041266 | 26.578631 | 21.6641257 | 31.4447935 | 32.4588045 | 27.6766232 | 21.2390619 | 0.10119915 | 0.10959247 | 0.32190157 | 0.32997825 | 0.08475895 | 0.09206801 | 0.27783771 | 0.24536373 |
| Percentage of water within green space | % | 0.84073801 | 0.71697488 | 1.56803972 | 1.78470765 | 1.05938513 | 0.94939459 | 1.783384 | 1.78422974 | 0.0130806 | 0.01317727 | 0.0626748 | 0.07091251 | 0.01158891 | 0.01193134 | 0.05343102 | 0.05402328 |
| **Land use indices – original sourced** | | | | | | | | | | | | | | | | | |
| Area of commercial land use surrounding green space (within 100m doughnut buffer) | m2 | 14722.4528 | 15963.2031 | 7876.98696 | 4738.14592 | 71667.1933 | 77487.9648 | 38280.0048 | 27552.0308 | 197.796696 | 223.853502 | 391.871672 | 259.580631 | 746.582583 | 848.072338 | 1242.21349 | 893.598371 |
| Area of education land use surrounding green space (within 100m doughnut buffer) | m2 | 7387.30593 | 7939.65873 | 3288.15353 | 4244.51799 | 40468.137 | 42912.1251 | 23610.9833 | 25451.5064 | 69.4943322 | 78.1030333 | 127.687536 | 162.948363 | 209.98206 | 233.434415 | 499.076131 | 506.359719 |
| Area of hospital/medical land use surrounding green space (within 100m doughnut buffer) | m2 | 1143.65613 | 1216.9737 | 906.90665 | 345.990551 | 6165.90425 | 6533.64861 | 5092.84722 | 2098.88938 | 33.976509 | 38.337836 | 67.6465285 | 74.4948938 | 95.9175402 | 107.719683 | 227.254139 | 215.326461 |
| Area of industrial land use surrounding green space (within 100m doughnut buffer) | m2 | 3473.58338 | 3626.16402 | 2464.25487 | 2453.14189 | 24496.9239 | 25915.3155 | 16597.6212 | 13455.0598 | 62.7935469 | 70.0938587 | 157.922125 | 168.578076 | 246.359932 | 277.288654 | 583.014216 | 446.039566 |
| Area of other land use surrounding green space (within 100m doughnut buffer) | m2 | 243.86313 | 229.470504 | 366.345833 | 306.353919 | 2682.49396 | 2673.81016 | 3238.91799 | 2122.58574 | 14.0213374 | 15.417442 | 48.8212492 | 36.3138627 | 159.038661 | 181.663864 | 269.192696 | 127.003886 |
| Area of parkland land use surrounding green space (within 100m doughnut buffer) | m2 | 42612.6893 | 45873.2379 | 20636.9247 | 21310.4682 | 265752.399 | 285979.404 | 137958.502 | 126996.195 | 300.019394 | 337.712637 | 458.620565 | 595.374236 | 1183.68623 | 1318.47543 | 1963.08722 | 2035.81876 |
| Area of primary production land use surrounding green space (within 100m doughnut buffer) | m2 | 279.384083 | 266.375763 | 195.517348 | 576.735175 | 1940.56294 | 1878.02043 | 1149.76454 | 3834.36196 | 15.5397128 | 17.2319147 | 27.9718885 | 61.4930918 | 82.0321943 | 92.6403784 | 102.693684 | 242.190187 |
| Area of residential land use surrounding green space (within 100m doughnut buffer) | m2 | 235330.751 | 256708.032 | 89620.0754 | 97683.9222 | 1148223.99 | 1245039.75 | 489407.726 | 542290.439 | 1061.97423 | 1172.61237 | 1428.40303 | 1909.89564 | 3513.32648 | 3776.14496 | 5126.92494 | 6724.6858 |
| Area of transport land use surrounding green space (within 100m doughnut buffer) | m2 | 3193.7191 | 3671.4456 | 20.9494104 | 14.3203622 | 15818.5615 | 18209.1666 | 163.509372 | 99.9812421 | 70.7088011 | 81.0773213 | 4.22616948 | 3.14551145 | 227.850061 | 260.554966 | 12.9910549 | 15.6548757 |
| Area of water land use surrounding green space (within 100m doughnut buffer) | m2 | 157.121281 | 179.141967 | 19.4095143 | 0 | 1080.26471 | 1218.78163 | 310.408325 | 0 | 11.7150235 | 13.4668542 | 5.23820955 | 0 | 46.4082749 | 53.0695995 | 75.4084246 | 0 |
| Area of tree canopy surrounding green space (within 100m doughnut buffer) | m2 | 19053.5411 | 21405.9833 | 2884.71321 | 2505.26882 | 90805.6272 | 100799.752 | 16822.5135 | 14087.0518 | 149.415777 | 167.546808 | 60.2409407 | 54.1359596 | 529.989876 | 581.74342 | 248.839373 | 184.143131 |
| Area of tree canopy ≤4m surrounding green space (within 100m doughnut buffer) | m2 | 1498.01024 | 1653.43437 | 369.72348 | 479.607741 | 7466.44509 | 8149.13265 | 2119.70702 | 2588.29456 | 7.97064865 | 8.79817398 | 7.12201138 | 11.6513839 | 27.4118263 | 29.2223834 | 28.8456379 | 37.217017 |
| Area of tree canopy >4m surrounding green space (within 100m doughnut buffer) | m2 | 16984.5131 | 19113.9213 | 2421.32714 | 1914.44364 | 80520.687 | 89530.7687 | 14190.7577 | 10900.0618 | 140.14516 | 157.382893 | 52.9153915 | 41.6879221 | 497.438421 | 547.410902 | 223.118936 | 143.965224 |
| **Land use indices – calculated** | | | | | | | | | | | | | | | | | |
| Percentage of commercial land use surrounding green space (within 100m doughnut buffer) | % | 3.75546046 | 3.70176773 | 4.51900116 | 3.60901229 | 3.7315274 | 3.63237092 | 4.63633353 | 4.06796064 | 0.03732137 | 0.03845665 | 0.18083777 | 0.17407621 | 0.02595542 | 0.02641907 | 0.12827238 | 0.12946238 |
| Percentage of education land use surrounding green space (within 100m doughnut buffer) | % | 2.35080167 | 2.27048156 | 2.7234563 | 3.08439831 | 2.52590777 | 2.42473054 | 3.16210684 | 3.22372954 | 0.01891675 | 0.01939052 | 0.08716016 | 0.09833431 | 0.01155791 | 0.01156029 | 0.06141125 | 0.05703756 |
| Percentage of hospital/medical land use surrounding green space (within 100m doughnut buffer) | % | 0.27662206 | 0.26373953 | 0.49956212 | 0.19228366 | 0.29910313 | 0.28331342 | 0.56081465 | 0.20739163 | 0.00653879 | 0.00678543 | 0.03250929 | 0.02624055 | 0.00428236 | 0.00429011 | 0.02481379 | 0.01748718 |
| Percentage of industrial land use surrounding green space (within 100m doughnut buffer) | % | 1.41538728 | 1.24923218 | 2.65032953 | 2.3584742 | 1.71414369 | 1.58043187 | 2.83732804 | 2.28755292 | 0.02308838 | 0.02240651 | 0.13354014 | 0.12859318 | 0.01752187 | 0.01724332 | 0.1022032 | 0.08391261 |
| Percentage of other land use surrounding green space (within 100m doughnut buffer) | % | 0.27900766 | 0.19441146 | 0.82564306 | 0.86083698 | 0.3461148 | 0.28101964 | 0.87065155 | 0.65276826 | 0.01297808 | 0.01204334 | 0.07980225 | 0.08473143 | 0.01247692 | 0.01273946 | 0.06477266 | 0.05461656 |
| Percentage of parkland land use surrounding green space (within 100m doughnut buffer) | % | 13.7199651 | 13.3458401 | 16.9886466 | 15.2393344 | 16.7807879 | 16.5587771 | 19.7276396 | 16.3965342 | 0.06251852 | 0.06660481 | 0.2483846 | 0.24908208 | 0.05057199 | 0.0541867 | 0.20365355 | 0.17775442 |
| Percentage of primary production land use surrounding green space (within 100m doughnut buffer) | % | 0.22331956 | 0.18641524 | 0.25263802 | 0.73605439 | 0.28206411 | 0.24702518 | 0.21051042 | 0.88422733 | 0.01166848 | 0.01202769 | 0.03936689 | 0.07339426 | 0.01128247 | 0.01195123 | 0.02126693 | 0.06936896 |
| Percentage of residential land use surrounding green space (within 100m doughnut buffer) | % | 77.2038077 | 77.9001746 | 71.4970112 | 73.9087212 | 73.4929238 | 74.0483754 | 67.8904572 | 72.2677903 | 0.07683583 | 0.08065506 | 0.30885724 | 0.34906696 | 0.05966656 | 0.06280938 | 0.22772804 | 0.27062778 |
| Percentage of transport land use surrounding green space (within 100m doughnut buffer) | % | 0.7317113 | 0.8397323 | 0.01585048 | 0.01088452 | 0.75623877 | 0.86866036 | 0.02409011 | 0.0120452 | 0.01360224 | 0.01557473 | 0.00281214 | 0.00246575 | 0.00938034 | 0.01069946 | 0.00199666 | 0.00174792 |
| Percentage of water land use surrounding green space (within 100m doughnut buffer) | % | 0.04391726 | 0.0482053 | 0.02786155 | 0 | 0.07118859 | 0.07529555 | 0.08006804 | 0 | 0.00317216 | 0.00360735 | 0.00658986 | 0 | 0.00314273 | 0.00333931 | 0.01661149 | 0 |
| Percentage of tree canopy to roads surrounding green space (within 100m doughnut buffer) | % | 17.2563026 | 18.4642648 | 8.72754103 | 9.04123495 | 17.2183622 | 18.355064 | 8.81233511 | 8.48199568 | 0.04483592 | 0.04711857 | 0.10453558 | 0.10834644 | 0.03997762 | 0.04147366 | 0.08032871 | 0.06848988 |
| Percentage of tree canopy ≤4m to roads surrounding green space (within 100m doughnut buffer) | % | 1.67876381 | 1.71667669 | 1.29323152 | 1.56804243 | 1.65951021 | 1.69357153 | 1.30512038 | 1.52449521 | 0.003472 | 0.00336368 | 0.02266566 | 0.0141705 | 0.00260397 | 0.00249088 | 0.01807284 | 0.01121707 |
| Percentage of tree canopy >4m to roads surrounding green space (within 100m doughnut buffer) | % | 14.9837413 | 16.1122107 | 7.17509257 | 7.11098026 | 14.9700794 | 16.0344315 | 7.246155 | 6.60788432 | 0.04300044 | 0.04549144 | 0.09318836 | 0.09814807 | 0.03837485 | 0.04003925 | 0.07362967 | 0.05852646 |
| Percentage of tree canopy surrounding green space (within 100m doughnut buffer) | % | 4.73546959 | 5.12740647 | 2.06468452 | 1.94957647 | 4.63888299 | 4.99295036 | 2.06319805 | 1.86483817 | 0.014184 | 0.01504731 | 0.0251753 | 0.0221002 | 0.01260733 | 0.01317452 | 0.02052314 | 0.0167249 |
| Percentage of tree canopy ≤4m surrounding green space (within 100m doughnut buffer) | % | 0.44758205 | 0.46704396 | 0.28272835 | 0.34948494 | 0.43590318 | 0.45282179 | 0.28543794 | 0.33722379 | 0.00095548 | 0.0009767 | 0.00386277 | 0.00399782 | 0.00073138 | 0.00072929 | 0.00329849 | 0.00300006 |
| Percentage of tree canopy >4m surrounding green space (within 100m doughnut buffer) | % | 4.126779 | 4.48623884 | 1.71896851 | 1.51972081 | 4.04616079 | 4.37160824 | 1.71586039 | 1.4503557 | 0.01350404 | 0.01441623 | 0.02310949 | 0.01890594 | 0.01202934 | 0.01265234 | 0.01911381 | 0.01390837 |

IRSD = Index of Relative Social Disadvantage; when reversed, the higher the score the greater the disadvantage.

IRSD scores were reversed calculating Max - X + Min, where Max and Min are scores in the original NSW-wise data, and X is a score to be reversed.

m=metres | sq m=square metres | number = number of items

Table S12. Analysis of green space qualities associations with IRSD adjusted for population density and random effects of geographic areas (buffer size=1600m).

|  | **Output variables negatively correlated with IRSD** | | | | | | **Output variables positively correlated with IRSD** | | | |
| --- | --- | --- | --- | --- | --- | --- | --- | --- | --- | --- |
| **Model structure, variables** | **Trees/roads (%)** | **Trees (%)** | **Slope >6 degrees (%)** | **Mammal species, n** | **All species, n** | **Shannon index** | **Mean incivilities** | **Open grass (sq m)** | **Open grass (%)** | **Bare earth (%)** |
| **Spearman's rho with reversed IRSD** | -0.52 | -0.46 | -0.49 | -0.47 | -0.39 | -0.39 | 0.30 | 0.30 | 0.38 | 0.33 |
| **Distribution** | Normal | Normal | Log-normal | Bi-modal | Bi-modal | Normal | U-shape | Log-normal | Normal | Log-normal |
| **Single-level models** | | | | | | | | | | |
| **Model 1: Intercept only** | | | | | | | | | | |
| **Intercept** | 17.2 (17.1, 17.3) * | 31.4 (31.3, 31.6) * | 26.8 (26.6, 27.0) * | 8.0 (7.9, 8.0) * | 60.3 (60.1, 60.5) * | 1.152 (1.151, 1.154) * | 1.539 (1.530, 1.549) * | 64507 (63972, 65036) * | 30.0 (29.8, 30.1) * | 11.2 (11.1, 11.3) * |
| **Var(MB, level 1)** | 71.3 (70.4, 72.3) | 326.6 (322.4, 330.9) | 395.5 (390.5, 400.6) | 5.2 (5.1, 5.2) | 552.0 (545.0, 559.0) | 0.017 (0.016, 0.017) | 1.110 (1.090, 1.120) | 3.35E9 (3.31E9, 3.4E9) | 235 (232, 238) | 127 (126, 129) |
| **DIC:** | 317,026 | 392,187 | 412,978 | 210,781 | 430,837 | -59,466 | 138,424 | 1,126,137 | 377,271 | 349,385 |
| **ΔDIC:** | 94,715 | 65,367 | 56,672 | 117,567 | 100,997 | 87,829 | 66,185 | 34,875 | 52,247 | 67,453 |
| **Model 2: Intercept + IRSD** | | | | | | | | | | |
| **Intercept** | 23.5 (23.4, 23.6) * | 43.0 (42.7, 43.3) * | 41.1 (40.8, 41.4) * | 9.1 (9.1, 9.2) * | 70.9 (70.5, 71.3) * | 1.224 (1.222, 1.226) * | 1.177 (1.159, 1.195) * | 47668 (46656, 48681) * | 22.7 (22.4, 22.9) * | 8.4 (8.2, 8.6) * |
| **IRSD quintile (ref=1, least disadvantaged)** |  |  |  |  |  |  |  |  |  |  |
| **2** | -4.8 (-5.0, -4.6) * | -7.9 (-8.3, -7.5) * | -11.5 (-12.0, -11.1) * | -0.8 (-0.9, -0.7) * | -6.8 (-7.3, -6.2) * | -0.059 (-0.062, -0.056) * | 0.234 (0.207, 0.260) * | 12373 (10834, 13893) * | 4.5 (4.1, 4.9) * | 1.4 (1.1, 1.7) * |
| **3** | -8.1 (-8.3, -7.9) * | -15.0 (-15.4, -14.5) * | -19.1 (-19.6, -18.6) * | -1.4 (-1.4, -1.3) * | -12.5 (-13.1, -11.9) * | -0.091 (-0.094, -0.088) * | 0.396 (0.368, 0.424) * | 22746 (21142, 24339) * | 8.8 (8.4, 9.2) * | 3.4 (3.0, 3.7) * |
| **4** | -10.0 (-10.2, -9.8) * | -19.0 (-19.5, -18.5) * | -22.4 (-22.9, -21.9) * | -1.8 (-1.9, -1.8) * | -16.3 (-16.9, -15.6) * | -0.109 (-0.112, -0.105) * | 0.563 (0.534, 0.591) * | 27118 (25473, 28754) * | 11.8 (11.4, 12.2) * | 4.4 (4.1, 4.7) * |
| **5 (most disadvantaged)** | -11.3 (-11.6, -11.1) * | -22.1 (-22.5, -21.6) * | -26.3 (-26.8, -25.9) * | -2.4 (-2.5, -2.4) * | -23.7 (-24.3, -23.1) * | -0.137 (-0.140, -0.133) * | 0.828 (0.800, 0.856) * | 31135 (29534, 32721) * | 15.5 (15.1, 15.9) * | 6.7 (6.4, 7.0) * |
| **Var(MB, level 1)** | 52.9 (52.2, 53.6) | 257.0 (253.7, 260.4) | 298.4 (294.5, 302.2) | 4.4 (4.3, 4.4) | 481.8 (475.7, 488.1) | 0.014 (0.014, 0.014) | 1.020 (1.010, 1.040) | 3.21E9 (3.17E9, 3.26E9) | 204 (201, 206) | 122 (120, 123) |
| **DIC:** | 303,710 | 381,293 | 399,777 | 203,206 | 424,443 | -67,029 | 134,694 | 1,124,203 | 370,695 | 347,279 |
| **ΔDIC:** | 81,399 | 54,473 | 43,471 | 109,992 | 94,603 | 80,266 | 62,455 | 32,941 | 45,671 | 65,347 |
| **Multi-level models (two level intercepts)** | | | | | | | | | | |
| **Model 3: Intercept + RE(SA2)** | | | | | | | | | | |
| **Intercept** | 16.7 (16.1, 17.6) * | 30.7 (29.3, 32.4) * | 25.7 (24.1, 27.3) * | 7.8 (7.6, 8.0) * | 58.5 (56.6, 60.8) * | 1.141 (1.131, 1.153) * | 1.439 (1.353, 1.534) * | 61330 (57204, 65484) * | 30.0 (28.8, 31.3) * | 11.2 (10.3, 12.2) * |
| **Variance of random effects** |  |  |  |  |  |  |  |  |  |  |
| **Var (SA2, level 2)** | 62.8 (54.2, 72.6) | 261.0 (225.3, 302.0) | 279.2 (241.2, 323.0) | 4.5 (3.9, 5.2) | 495.4 (428.2, 572.9) | 0.014 (0.012, 0.016) | 0.884 (0.764, 1.023) | 1.78E9 (1.54E9, 2.06E9) | 166 (143, 192) | 98 (85, 114) |
| **Var (MB, level 1)** | 8.6 (8.5, 8.7) | 79.7 (78.7, 80.7) | 123.2 (121.6, 124.8) | 0.4 (0.4, 0.4) | 64.1 (63.3, 65.0) | 0.003 (0.003, 0.003) | 0.291 (0.287, 0.294) | 1.57E9 (1.55E9, 1.59E9) | 76 (75, 77) | 29 (28, 29) |
| **DIC:** | 222,912 | 328,418 | 358,713 | 93,215 | 329,862 | -147,221 | 75,766 | 1,092,023 | 326,023 | 282,169 |
| **ΔDIC:** | 601 | 1,598 | 2,407 | 1 | 22 | 74 | 3,527 | 761 | 999 | 237 |
| **Model 4: Intercept + IRSD + RE(SA2)** | | | | | | | | | | |
| **Intercept** | 17.8 (16.8, 18.5) * | 34.7 (33.0, 36.2) * | 31.9 (30.3, 33.4) * | 7.8 (7.6, 8.1) * | 58.9 (56.6, 61.3) * | 1.147 (1.135, 1.159) * | 1.125 (1.032, 1.216) * | 50309 (45921, 54447) * | 27.2 (25.9, 28.4) * | 10.6 (9.5, 11.6) * |
| **IRSD quintile (ref=1, least disadvantaged)** |  |  |  |  |  |  |  |  |  |  |
| **2** | -0.8 (-0.9, -0.7) * | -2.0 (-2.2, -1.7) * | -4.5 (-4.8, -4.2) * | 0.0 (-0.0, 0.0) | -0.4 (-0.6, -0.2) x | -0.005 (-0.006, -0.003) * | 0.196 (0.180, 0.211) * | 5765 (4571, 6954) * | 1.7 (1.4, 1.9) * | 0.2 (0.1, 0.4) x |
| **3** | -1.3 (-1.4, -1.2) * | -4.6 (-4.9, -4.3) * | -8.0 (-8.4, -7.7) * | 0.0 (-0.0, 0.0) | -0.3 (-0.6, -0.0) + | -0.006 (-0.007, -0.004) * | 0.368 (0.350, 0.386) * | 14520 (13101, 15937) * | 4.0 (3.7, 4.3) * | 0.8 (0.6, 1.0) * |
| **4** | -1.2 (-1.4, -1.1) * | -6.1 (-6.4, -5.7) * | -9.7 (-10.1, -9.3) * | -0.0 (-0.0, 0.0) | 0.1 (-0.2, 0.5) | -0.009 (-0.011, -0.007) * | 0.530 (0.509, 0.551) * | 18199 (16586, 19804) * | 4.7 (4.3, 5.0) * | 1.4 (1.2, 1.6) * |
| **5 (most disadvantaged)** | -1.2 (-1.3, -1.0) * | -8.2 (-8.6, -7.7) * | -11.4 (-11.9, -10.9) * | -0.0 (-0.1, 0.0) | 0.3 (-0.1, 0.7) ^ | -0.005 (-0.008, -0.003) * | 0.715 (0.690, 0.739) * | 24911 (23021, 26785) * | 6.2 (5.7, 6.6) * | 1.9 (1.6, 2.1) * |
| **Variance of random effects** |  |  |  |  |  |  |  |  |  |  |
| **Var (SA2, level 2)** | 59.2 (51.1, 68.6) | 225.0 (194.1, 260.5) | 222.2 (191.7, 257.2) | 4.4 (3.8, 5.1) | 497.5 (429.9, 575.7) | 0.013 (0.011, 0.015) | 0.797 (0.688, 0.923) | 1.65E9 (1.42E9, 1.92E9) | 147 (127, 171) | 96 (83, 111) |
| **Var (MB, level 1)** | 8.5 (8.4, 8.6) | 76.9 (76.0, 78.0) | 117.0 (115.5, 118.6) | 0.4 (0.4, 0.4) | 64.1 (63.3, 64.9) | 0.000 (0.000, 0.000) | 0.270 (0.270, 0.270) | 1.54E9 (1.52E9, 1.57E9) | 74 (73, 75) | 29 (28, 29) |
| **DIC:** | 222,314 | 326,822 | 356,309 | 93,218 | 329,841 | -147,293 | 72,241 | 1,091,266 | 325,025 | 281,933 |
| **ΔDIC:** | 3 | 2 | 3 | 4 | 1 | 2 | 2 | 4 | 1 | 1 |
| **Model 5: Intercept + IRSD + Pop (SA2)+ RE(SA2)** | | | | | | | | | | |
| **Intercept** | 12.6 (10.7, 13.9) * | 34.9 (31.8, 38.0) * | 38.8 (35.8, 41.8) * | 6.9 (6.6, 7.3) * | 48.2 (44.5, 52.4) * | 1.136 (1.115, 1.158) * | -0.023 (-0.159, 0.118) | 29860 (21205, 38491) * | 25.4 (22.8, 27.9) * | 10.6 (8.6, 12.6) * |
| **IRSD quintile (ref=1, least disadvantaged)** |  |  |  |  |  |  |  |  |  |  |
| **2** | -0.8 (-0.9, -0.7) * | -2.0 (-2.2, -1.7) * | -4.5 (-4.8, -4.2) * | 0.0 (-0.0, 0.0) | -0.4 (-0.6, -0.2) * | -0.005 (-0.006, -0.003) * | 0.196 (0.180, 0.211) * | 5761 (4558, 6954) * | 1.7 (1.4, 1.9) * | 0.2 (0.1, 0.4) x |
| **3** | -1.3 (-1.4, -1.2) * | -4.6 (-4.9, -4.3) * | -8.0 (-8.4, -7.7) * | 0.0 (-0.0, 0.0) | -0.3 (-0.6, -0.0) + | -0.006 (-0.008, -0.004) * | 0.368 (0.350, 0.386) * | 14515 (13086, 15939) * | 4.0 (3.7, 4.3) * | 0.8 (0.6, 1.0) * |
| **4** | -1.2 (-1.4, -1.1) * | -6.1 (-6.4, -5.7) * | -9.7 (-10.1, -9.3) * | -0.0 (-0.0, 0.0) | 0.1 (-0.2, 0.4) | -0.009 (-0.011, -0.007) * | 0.530 (0.509, 0.551) * | 18175 (16573, 19802) * | 4.7 (4.3, 5.0) * | 1.4 (1.2, 1.6) * |
| **5 (most disadvantaged)** | -1.2 (-1.3, -1.0) * | -8.2 (-8.6, -7.7) * | -11.4 (-11.9, -10.9) * | -0.0 (-0.1, 0.0) | 0.3 (-0.1, 0.7) | -0.005 (-0.008, -0.003) * | 0.713 (0.689, 0.738) * | 24832 (22937, 26730) * | 6.1 (5.7, 6.6) * | 1.9 (1.6, 2.1) * |
| **SA2 population density (deciles)** | 0.9 (0.7, 1.2) * | -0.1 (-0.6, 0.4) | -1.3 (-1.8, -0.8) * | 0.2 (0.1, 0.2) * | 1.9 (1.2, 2.5) * | 0.002 (-0.002, 0.005) | 0.208 (0.185, 0.229) * | 3678 (2274, 5008) * | 0.3 (-0.1, 0.7) | -0.0 (-0.4, 0.3) |
| **Variance of random effects** |  |  |  |  |  |  |  |  |  |  |
| **Var (SA2, level 2)** | 53.4 (46.2, 61.9) | 225.3 (194.7, 261.0) | 208.8 (180.3, 241.9) | 4.3 (3.7, 4.9) | 470.3 (406.6, 543.9) | 0.013 (0.012, 0.015) | 0.443 (0.383, 0.513) | 1.55E9 (1.34E9, 1.8E9) | 147 (127, 170) | 96 (83, 111) |
| **Var (MB, level 1)** | 8.5 (8.4, 8.6) | 76.9 (76.0, 78.0) | 117.0 (115.5, 118.5) | 0.4 (0.4, 0.4) | 64.1 (63.3, 64.9) | 0.003 (0.003, 0.003) | 0.270 (0.266, 0.273) | 1.54E9 (1.52E9, 1.56E9) | 74 (73, 75) | 29 (28, 29) |
| **DIC:** | 222,314 | 326,822 | 356,309 | 93,218 | 329,841 | -147,293 | 72,240 | 1,091,265 | 325,025 | 281,933 |
| **ΔDIC:** | 3 | 2 | 3 | 4 | 1 | 2 | 1 | 3 | 1 | 1 |
| **Multi-level models (four level intercepts)** | | | | | | | | | | |
| **Model 6: Intercept + RE (SA2, 3, 4)** | | | | | | | | | | |
| **Intercept** | 16.0 (15.0, 17.0) * | 32.4 (28.1, 39.3) * | 29.0 (23.2, 43.2) * | 7.8 (7.3, 9.1) * | 59.5 (53.8, 73.7) * | 1.153 (1.117, 1.239) * | 1.389 (1.146, 1.890) * | 58033 (47593, 69326) * | 30.2 (25.7, 41.2) * | 11.1 (8.8, 15.2) * |
| **Var (SA4, level 4)** | 45.3 (19.8, 96.0) | 33.0 (0.0, 157.4) | 136.9 (34.6, 411.2) | 3.3 (1.4, 7.3) | 263.7 (104.6, 614.3) | 0.010 (0.004, 0.023) | 0.170 (0.002, 0.592) | 3.72E8 (0, 1.21E9) | 83 (17, 262) | 11 (0, 40) |
| **Var (SA3, level 3)** | 11.9 (6.4, 20.4) | 196.2 (92.8, 333.5) | 100.2 (48.3, 180.9) | 0.8 (0.4, 1.3) | 114.6 (58.4, 193.7) | 0.003 (0.002, 0.005) | 0.320 (0.158, 0.568) | 5.2E8 (1.87E8, 1.11E9) | 46 (19, 99) | 24 (4, 49) |
| **Var (SA2, level 2)** | 11.8 (10.0, 13.9) | 91.5 (78.0, 107.4) | 110.8 (94.4, 130.0) | 1.1 (0.9, 1.3) | 183.9 (156.8, 215.6) | 0.004 (0.003, 0.005) | 0.455 (0.389, 0.533) | 9.94E8 (8.46E8, 1.16E9) | 82 (69, 96) | 66 (56, 77) |
| **Var (MB, level 1)** | 8.6 (8.5, 8.7) | 79.7 (78.7, 80.8) | 123.2 (121.6, 124.8) | 0.4 (0.4, 0.4) | 64.1 (63.3, 65.0) | 0.003 (0.003, 0.003) | 0.291 (0.287, 0.294) | 1.57E9 (1.55E9, 1.59E9) | 76 (75, 77) | 29 (28, 29) |
| **DIC:** | 222,909 | 328,416 | 358,712 | 93,214 | 329,861 | -147,223 | 75,765 | 1,092,020 | 326,022 | 282,168 |
| **ΔDIC:** | 598 | 1,596 | 2,406 | 0 | 21 | 72 | 3,526 | 758 | 998 | 236 |
| **Model 7: Intercept + IRSD + RE (SA2, 3, 4)** | | | | | | | | | | |
| **Intercept** | 16.4 (13.8, 20.0) * | 36.1 (31.9, 39.9) * | 33.2 (28.2, 38.6) * | 7.7 (7.0, 8.4) * | 57.9 (49.3, 65.2) * | 1.145 (1.092, 1.189) * | 1.034 (0.716, 1.325) * | 45762 (29435, 57044) * | 25.4 (20.2, 30.0) * | 10.0 (7.5, 11.9) * |
| **IRSD quintile (ref=1, least disadvantaged)** |  |  |  |  |  |  |  |  |  |  |
| **2** | -0.8 (-0.8, -0.7) * | -1.9 (-2.2, -1.7) * | -4.5 (-4.8, -4.2) * | 0.0 (-0.0, 0.0) | -0.4 (-0.6, -0.1) x | -0.004 (-0.006, -0.003) * | 0.196 (0.181, 0.212) * | 5729 (4526, 6923) * | 1.7 (1.4, 1.9) * | 0.2 (0.1, 0.4) x |
| **3** | -1.3 (-1.4, -1.2) * | -4.6 (-4.9, -4.3) * | -8.0 (-8.4, -7.6) * | 0.0 (-0.0, 0.0) | -0.3 (-0.6, -0.0) + | -0.006 (-0.007, -0.004) * | 0.369 (0.350, 0.387) * | 14465 (13046, 15885) * | 4.0 (3.7, 4.3) * | 0.8 (0.6, 1.0) * |
| **4** | -1.2 (-1.4, -1.1) * | -6.0 (-6.4, -5.7) * | -9.6 (-10.1, -9.2) * | -0.0 (-0.0, 0.0) | 0.2 (-0.2, 0.5) | -0.009 (-0.011, -0.007) * | 0.532 (0.511, 0.553) * | 18159 (16530, 19789) * | 4.6 (4.3, 5.0) * | 1.4 (1.2, 1.6) * |
| **5 (most disadvantaged)** | -1.2 (-1.3, -1.0) * | -8.1 (-8.5, -7.6) * | -11.3 (-11.8, -10.8) * | -0.0 (-0.0, 0.0) | 0.4 (-0.0, 0.7) ^ | -0.005 (-0.007, -0.003) * | 0.719 (0.694, 0.743) * | 24954 (23044, 26848) * | 6.1 (5.7, 6.5) * | 1.8 (1.6, 2.1) * |
| **Variance of random effects** |  |  |  |  |  |  |  |  |  |  |
| **Var (SA4, level 4)** | 46.2 (19.2, 103.4) | 24.1 (0.0, 89.5) | 73.9 (14.8, 190.4) | 3.3 (1.4, 7.4) | 267.3 (103.1, 635.9) | 0.010 (0.004, 0.022) | 0.214 (0.043, 0.668) | 3.83E8 (37, 1.2E9) | 63 (21, 149) | 8 (0, 29) |
| **Var (SA3, level 3)** | 11.9 (6.2, 20.5) | 159.9 (86.2, 270.1) | 86.2 (41.3, 154.9) | 0.8 (0.4, 1.4) | 121.9 (65.7, 203.9) | 0.003 (0.002, 0.005) | 0.270 (0.134, 0.477) | 4.35E8 (1.68E8, 8.81E8) | 36 (14, 66) | 25 (8, 50) |
| **Var (SA2, level 2)** | 11.4 (9.7, 13.4) | 86.9 (74.1, 101.9) | 98.9 (84.2, 116.2) | 1.1 (0.9, 1.3) | 183.0 (156.3, 214.3) | 0.004 (0.003, 0.004) | 0.375 (0.321, 0.438) | 9.37E8 (7.99E8, 1.1E9) | 79 (67, 93) | 65 (55, 76) |
| **Var (MB, level 1)** | 8.5 (8.4, 8.6) | 76.9 (75.9, 77.9) | 117.0 (115.5, 118.5) | 0.4 (0.4, 0.4) | 64.1 (63.3, 64.9) | 0.003 (0.003, 0.003) | 0.270 (0.266, 0.273) | 1.54E9 (1.52E9, 1.56E9) | 74 (73, 75) | 29 (28, 29) |
| **DIC:** | 222,311 | 326,820 | 356,306 | 93,216 | 329,840 | -147,295 | 72,239 | 1,091,262 | 325,024 | 281,932 |
| **ΔDIC:** | 0 | 0 | 0 | 2 | 0 | 0 | 0 | 0.9 | 0 | 0 |

Variables with some of the most extreme (rho ≥0.30 or rho ≤-0.39) coefficients of correlation were selected to represent six "concepts", trees, open grass, bare earth, slope, biodiversity and incivility. Output variables were considered to be continuous normally distributed variables, though some of them were log-normal, bi-modal and U-shaped as assessed visually.

MB=Mesh block, level 1, the lowest, n= 44620 to 47079 due to different number of missing values|| SA=Statistical area || SA2, n=362; SA3, n=48; SA4, n=16.

RE= Random effects, are random intercepts associated with different geographical areas.

Var = Variance of the random intercepts of different levels.

IRSD = Index of relative socioeconomic disadvantage; here is reversed, the higher the index, the higher the disadvantage.

IRSD is used as a categorical variable; population densities are continuous.

Trees/roads (%) = Percentage of tree canopy to roads surrounding green space (within 100m doughnut buffer).

Slope >6 degrees (%) = Percentage of green space which is greater than 6 degrees of slope.

Trees (%) = Percentage of tree canopy within green space.

Open grass (sq m) = Area of open grass within green space.

Open grass (%) = Percentage of open grass within green space.

Bare earth (%) = Percentage of bare earth within green space.

Mean incivilities = Mean level of incivilities.

Mammal species, All species = Number of species/species habitat likely to occur within green space.

Population density is per square km, converted to deciles.

DIC= Deviance information criterion, the smaller the better fit of the model.

Fits are produced using MCMC procedure in MLwiN v3.05 software with 5000-50000 burn-in and sample iterations. Confidence intervals are credible intervals.

* P≤0.001, x P≤0.01, + P≤0.05, ^ P≤0.1.

Table S13. Correlations between qualities with the most extreme correlations with SEIFA IRSD.

| **Variable** | **Desc** | Incivility_eq | AreaSGrass_w0sum | MalDamage_Max | PerSEarth_eq | Robbery_Max | PerSGrass_eq | CanPerTR_eq | CanPerTG4_eq | PerSlope6_eq | Mammals_Max | PerSTrees_eq | Can100TG4P_eq | Can100TrPe_eq | Total number of species | Shannon biodiversity index |
| --- | --- | --- | --- | --- | --- | --- | --- | --- | --- | --- | --- | --- | --- | --- | --- | --- |
| Incivility_eq | Mean level of incivilities |  | 0.37 | 0.82 | 0.20 | 0.84 | 0.19 | 0.07 | 0.09 | -0.29 | 0.11 | -0.24 | 0.21 | 0.20 | 0.12 | -0.07 |
| AreaSGrass_w0sum | Area of open grass within green space | 0.37 |  | 0.35 | 0.12 | 0.32 | 0.66 | -0.23 | -0.21 | -0.32 | -0.04 | -0.41 | -0.12 | -0.12 | -0.03 | -0.15 |
| MalDamage_Max | Density of malicious damage | 0.82 | 0.35 |  | 0.16 | 0.62 | 0.23 | 0.01 | 0.02 | -0.30 | 0.11 | -0.23 | 0.12 | 0.11 | 0.09 | -0.06 |
| PerSEarth_eq | Percentage of bare earth within green space | 0.20 | 0.12 | 0.16 |  | 0.20 | 0.05 | -0.41 | -0.41 | -0.40 | -0.25 | -0.65 | -0.39 | -0.39 | -0.12 | -0.20 |
| Robbery_Max | Density of robberies | 0.84 | 0.32 | 0.62 | 0.20 |  | 0.21 | -0.04 | -0.02 | -0.26 | -0.03 | -0.23 | 0.08 | 0.08 | -0.03 | -0.14 |
| PerSGrass_eq | Percentage of open grass within green space | 0.19 | 0.66 | 0.23 | 0.05 | 0.21 |  | -0.43 | -0.43 | -0.48 | -0.27 | -0.56 | -0.33 | -0.32 | -0.30 | -0.30 |
| CanPerTR_eq | Percentage of tree canopy to roads surrounding green space (within 100m doughnut buffer) | 0.07 | -0.23 | 0.01 | -0.41 | -0.04 | -0.43 |  | 1.00 | 0.36 | 0.50 | 0.63 | 0.95 | 0.94 | 0.45 | 0.35 |
| CanPerTG4_eq | Percentage of tree canopy >4m to roads surrounding green space (within 100m doughnut buffer) | 0.09 | -0.21 | 0.02 | -0.41 | -0.02 | -0.43 | 1.00 |  | 0.35 | 0.51 | 0.63 | 0.95 | 0.94 | 0.46 | 0.35 |
| PerSlope6_eq | Percentage of park which is greater than 6 degrees of Slope | -0.29 | -0.32 | -0.30 | -0.40 | -0.26 | -0.48 | 0.36 | 0.35 |  | 0.39 | 0.57 | 0.23 | 0.23 | 0.26 | 0.43 |
| Mammals_Max | Number of mammal species/species habitat likely to occur within green space | 0.11 | -0.04 | 0.11 | -0.25 | -0.03 | -0.27 | 0.50 | 0.51 | 0.39 |  | 0.33 | 0.46 | 0.45 | 0.75 | 0.63 |
| PerSTrees_eq | Percentage of tree canopy within green space | -0.24 | -0.41 | -0.23 | -0.65 | -0.23 | -0.56 | 0.63 | 0.63 | 0.57 | 0.33 |  | 0.56 | 0.55 | 0.24 | 0.36 |
| Can100TG4P_eq | Percentage of tree canopy >4m surrounding green space (within 100m doughnut buffer) | 0.21 | -0.12 | 0.12 | -0.39 | 0.08 | -0.33 | 0.95 | 0.95 | 0.23 | 0.46 | 0.56 |  | 1.00 | 0.42 | 0.27 |
| Can100TrPe_eq | Percentage of tree canopy surrounding green space (within 100m doughnut buffer) | 0.20 | -0.12 | 0.11 | -0.39 | 0.08 | -0.32 | 0.94 | 0.94 | 0.23 | 0.45 | 0.55 | 1.00 |  | 0.40 | 0.26 |
| Nspecies_eq | Total number of species/species habitat likely to occur within green space | 0.12 | -0.03 | 0.09 | -0.12 | -0.03 | -0.30 | 0.45 | 0.46 | 0.26 | 0.75 | 0.24 | 0.42 | 0.40 |  | 0.57 |
| Shannon_Index_eq | Shannon biodiversity index | -0.07 | -0.15 | -0.06 | -0.20 | -0.14 | -0.30 | 0.35 | 0.35 | 0.43 | 0.63 | 0.36 | 0.27 | 0.26 | 0.57 |  |

### S5. Maps

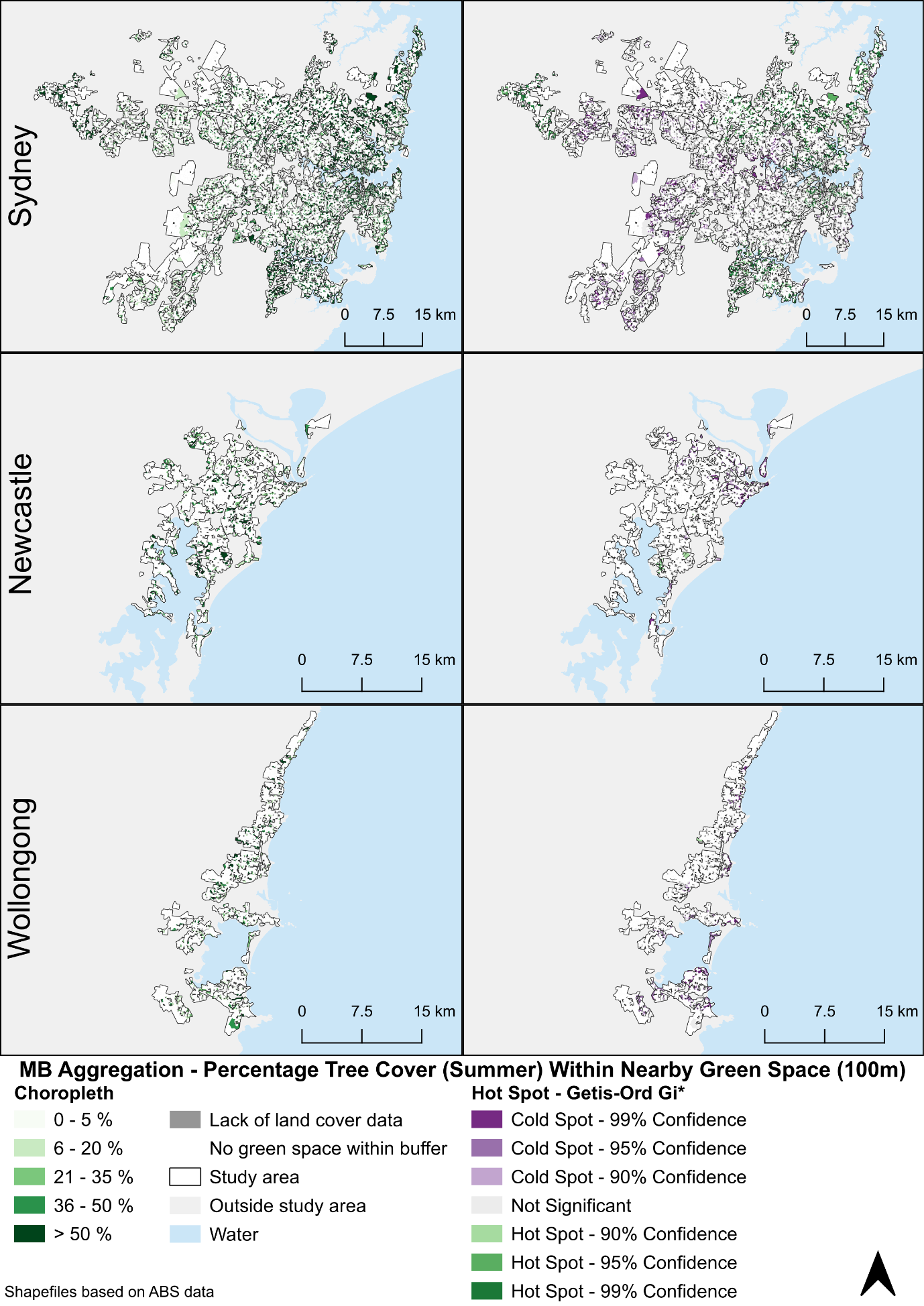

Figure S5. Percentage Tree Cover Within Nearby Green Space (100m network distance) – Aggregation for residential MBs that contain SEIFA data. Shapefiles based on ABS data.

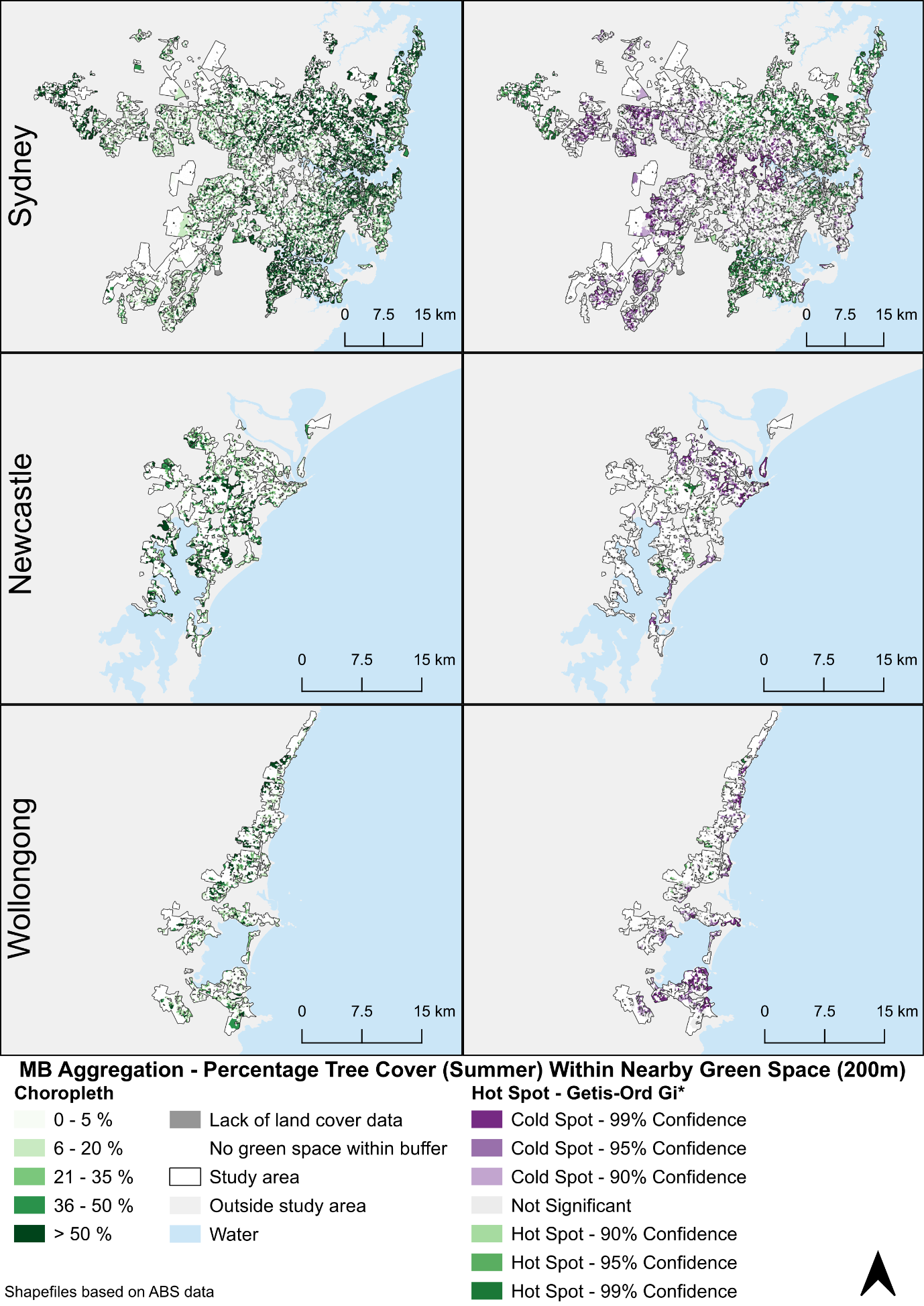

Figure S6. Percentage Tree Cover Within Nearby Green Space (200m network distance) – Aggregation for residential MBs that contain SEIFA data. Shapefiles based on ABS data.

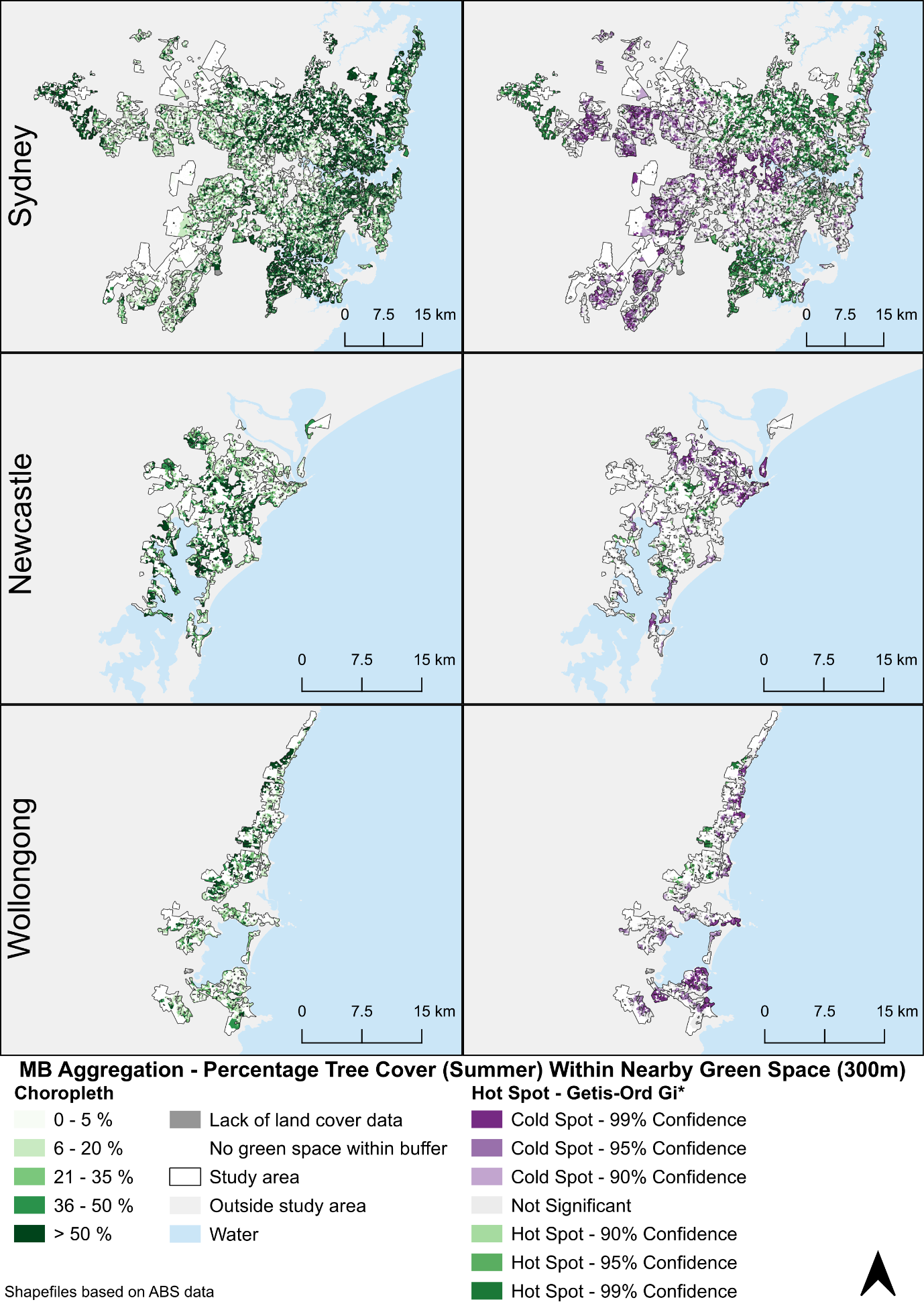

Figure S7. Percentage Tree Cover Within Nearby Green Space (300m network distance) – Aggregation for residential MBs that contain SEIFA data. Shapefiles based on ABS data.

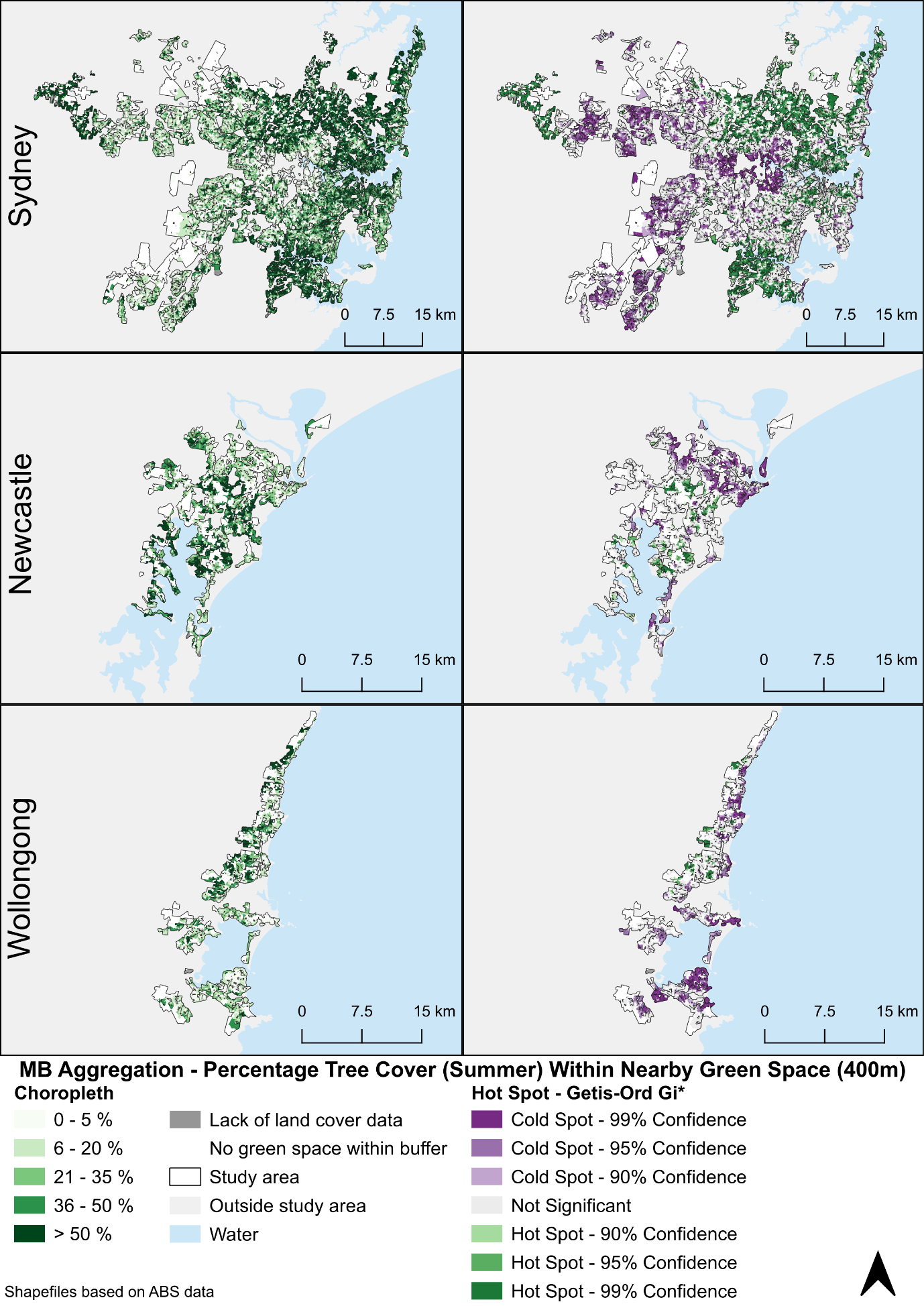

Figure S8. Percentage Tree Cover Within Nearby Green Space (400m network distance) – Aggregation for residential MBs that contain SEIFA data. Shapefiles based on ABS data.

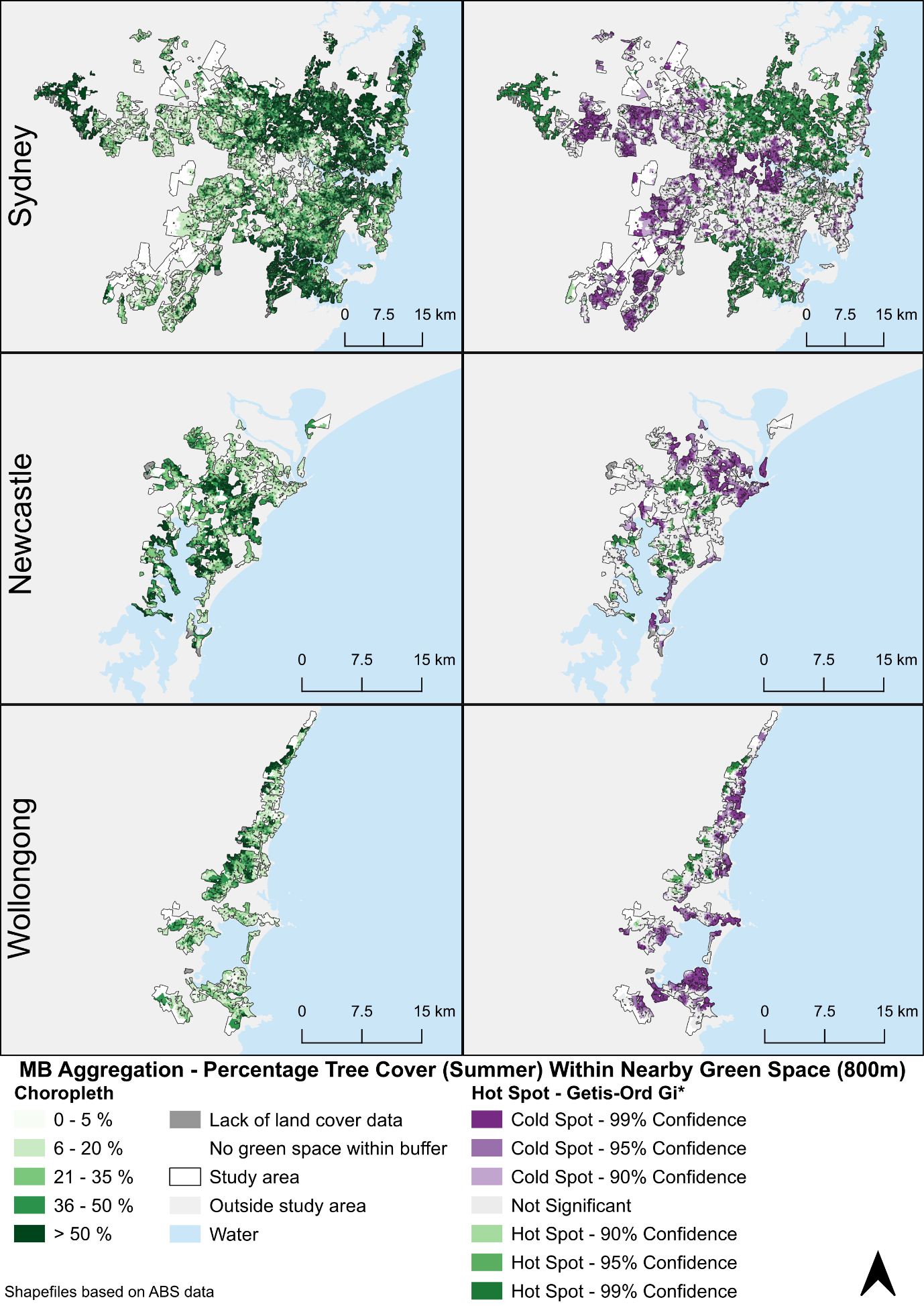

Figure S9. Percentage Tree Cover Within Nearby Green Space (800m network distance) – Aggregation for residential MBs that contain SEIFA data. Shapefiles based on ABS data.

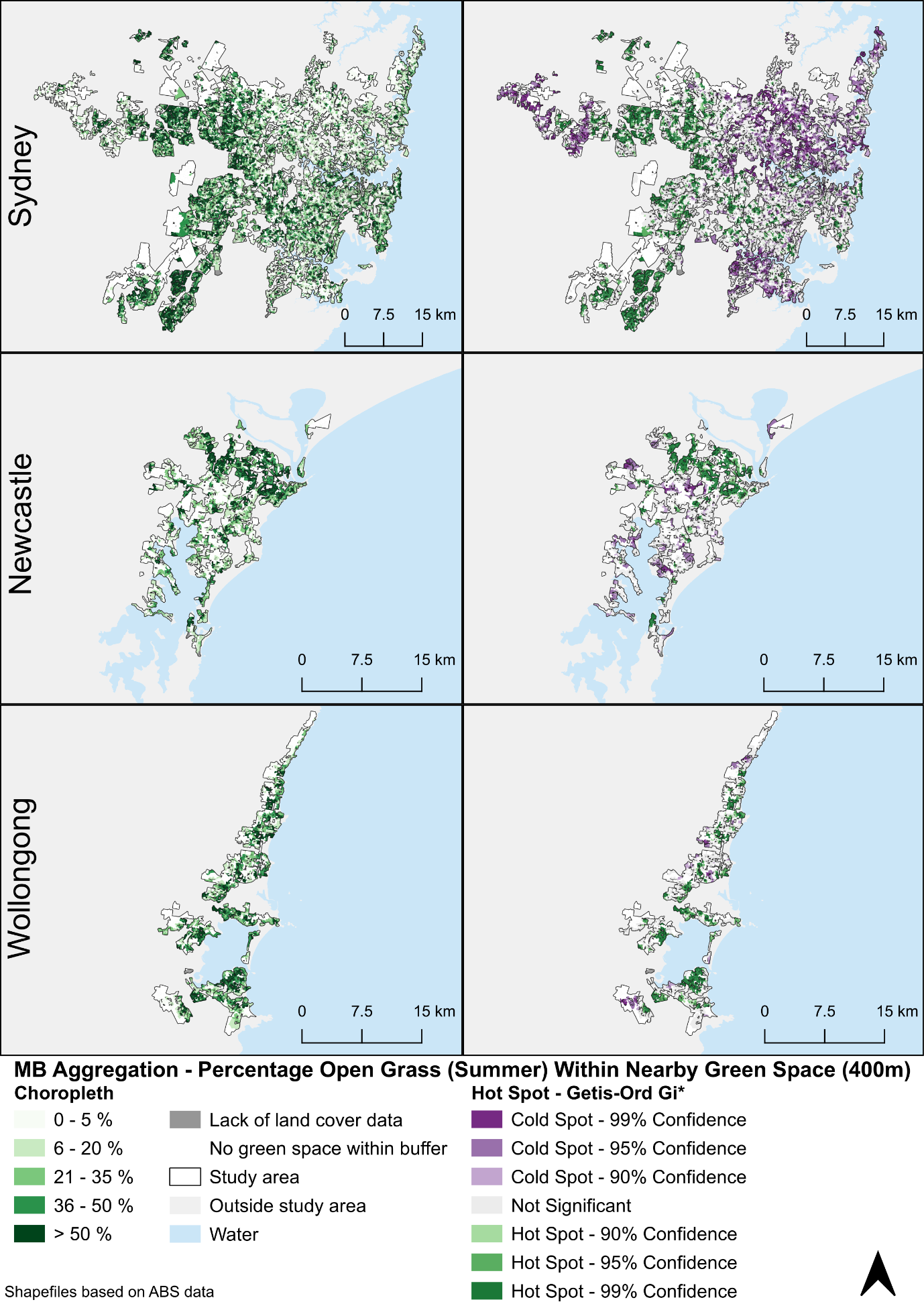

Figure S10. Percentage Open Grass Within Nearby Green Space (400m network distance) – Aggregation for residential MBs that contain SEIFA data. Shapefiles based on ABS data.

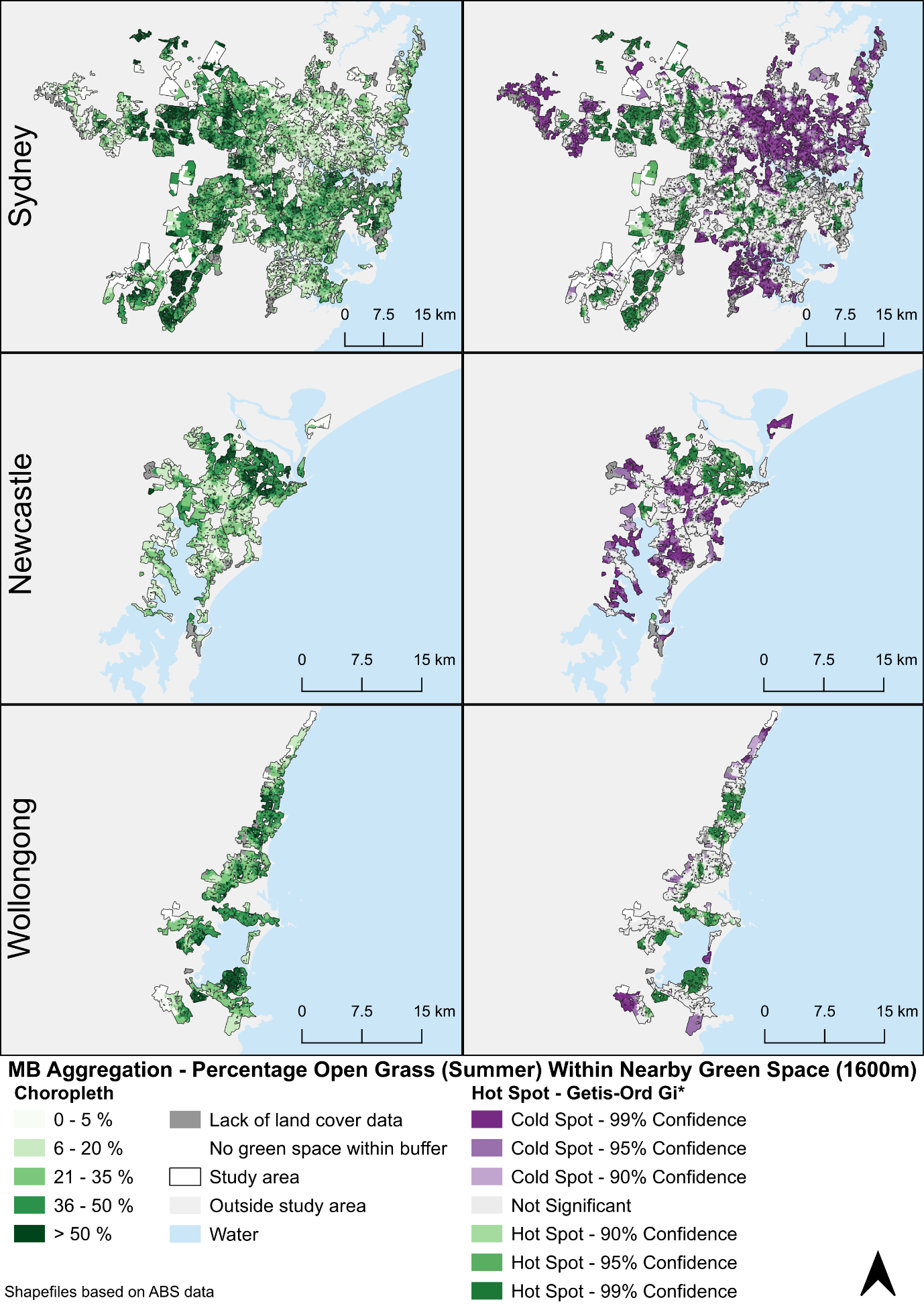

Figure S11. Percentage Open Grass Within Nearby Green Space (1600m network distance) – Aggregation for residential MBs that contain SEIFA data. Shapefiles based on ABS data.

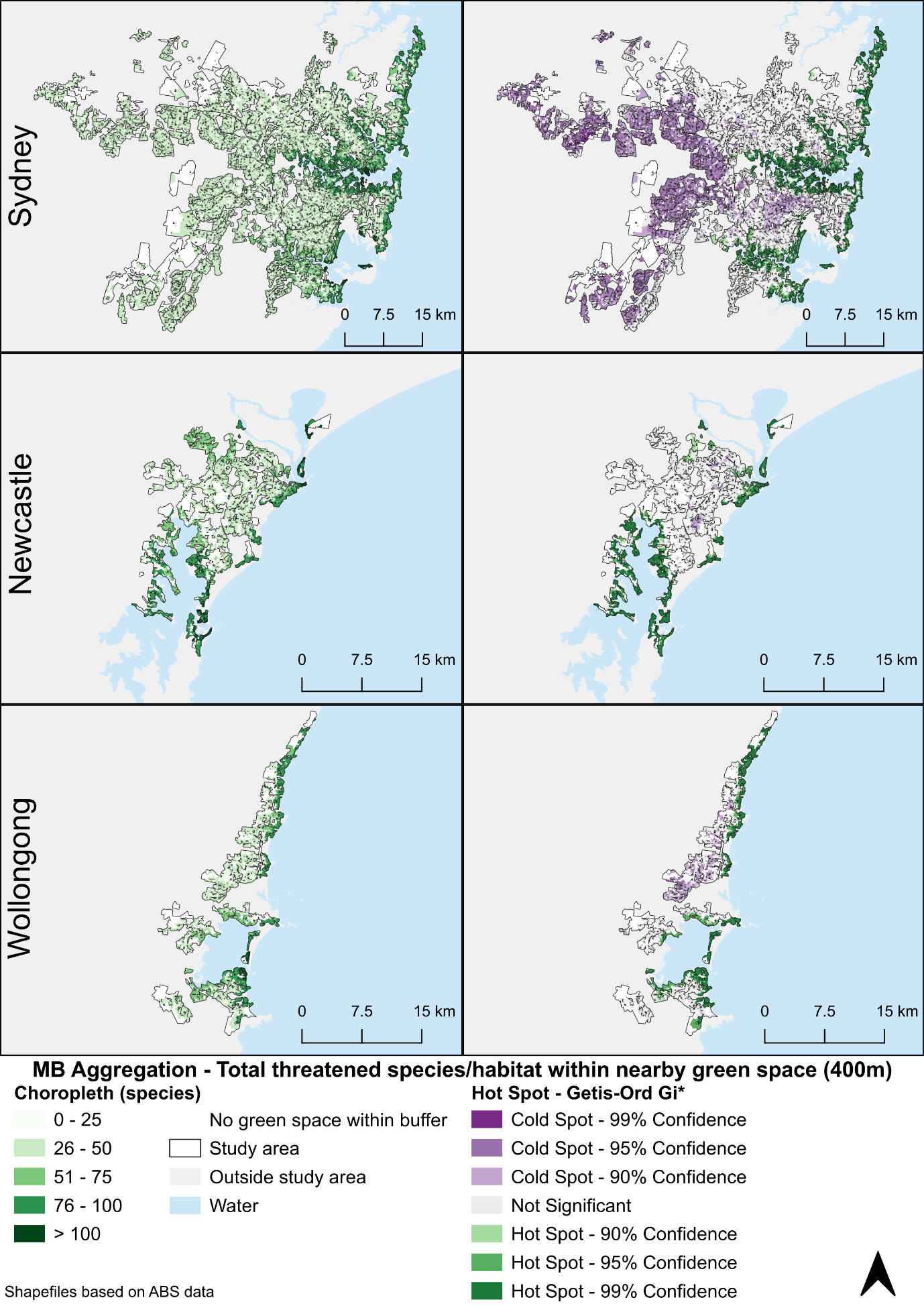

Figure S12. Total likely threatened species/species habitat within nearby green space (400m network distance) – Aggregation for residential MBs that contain SEIFA data. Shapefiles based on ABS data.

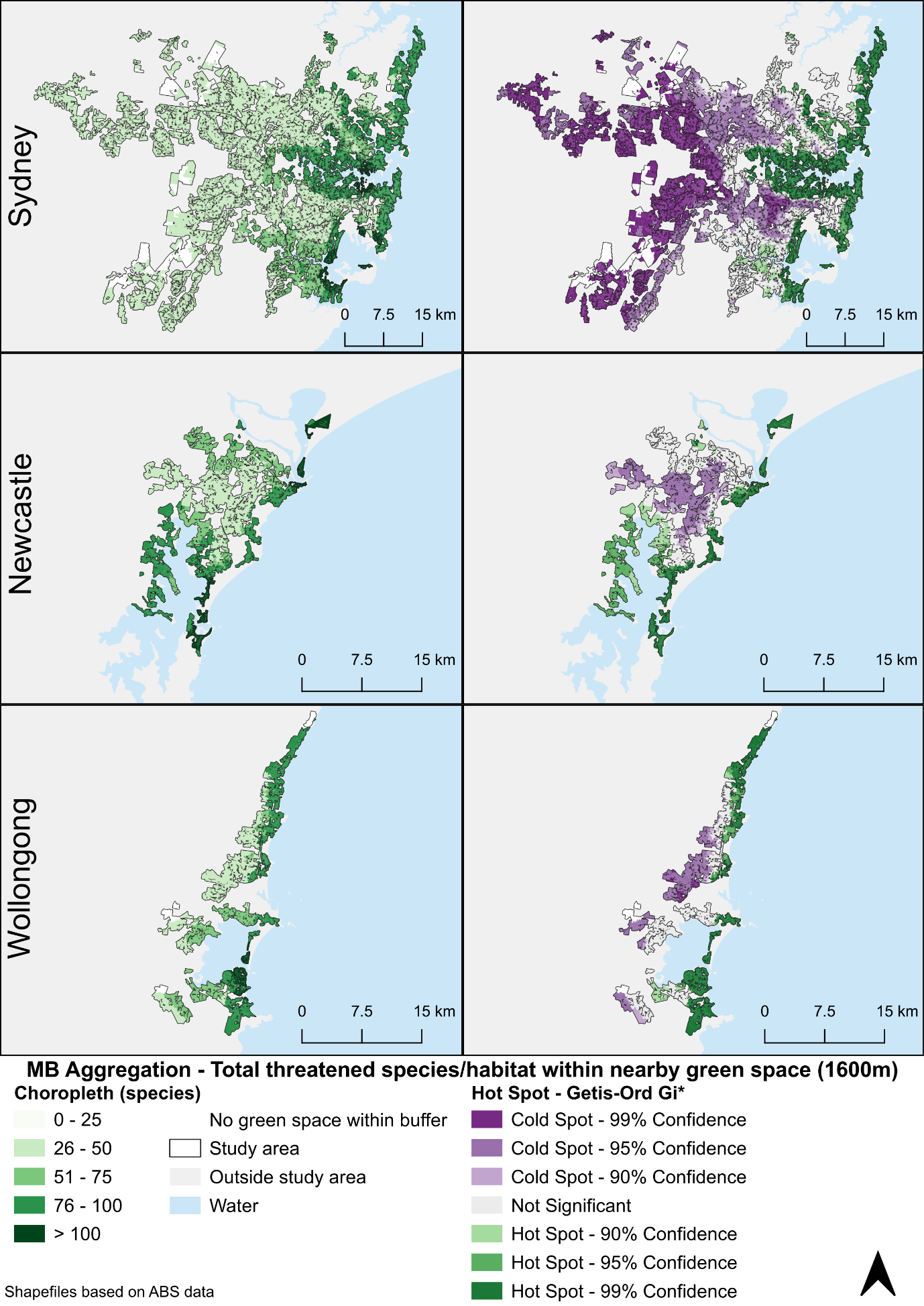

Figure S13. Total likely threatened species/species habitat within nearby green space (1600m network distance) – Aggregation for residential MBs that contain SEIFA data. Shapefiles based on ABS data.

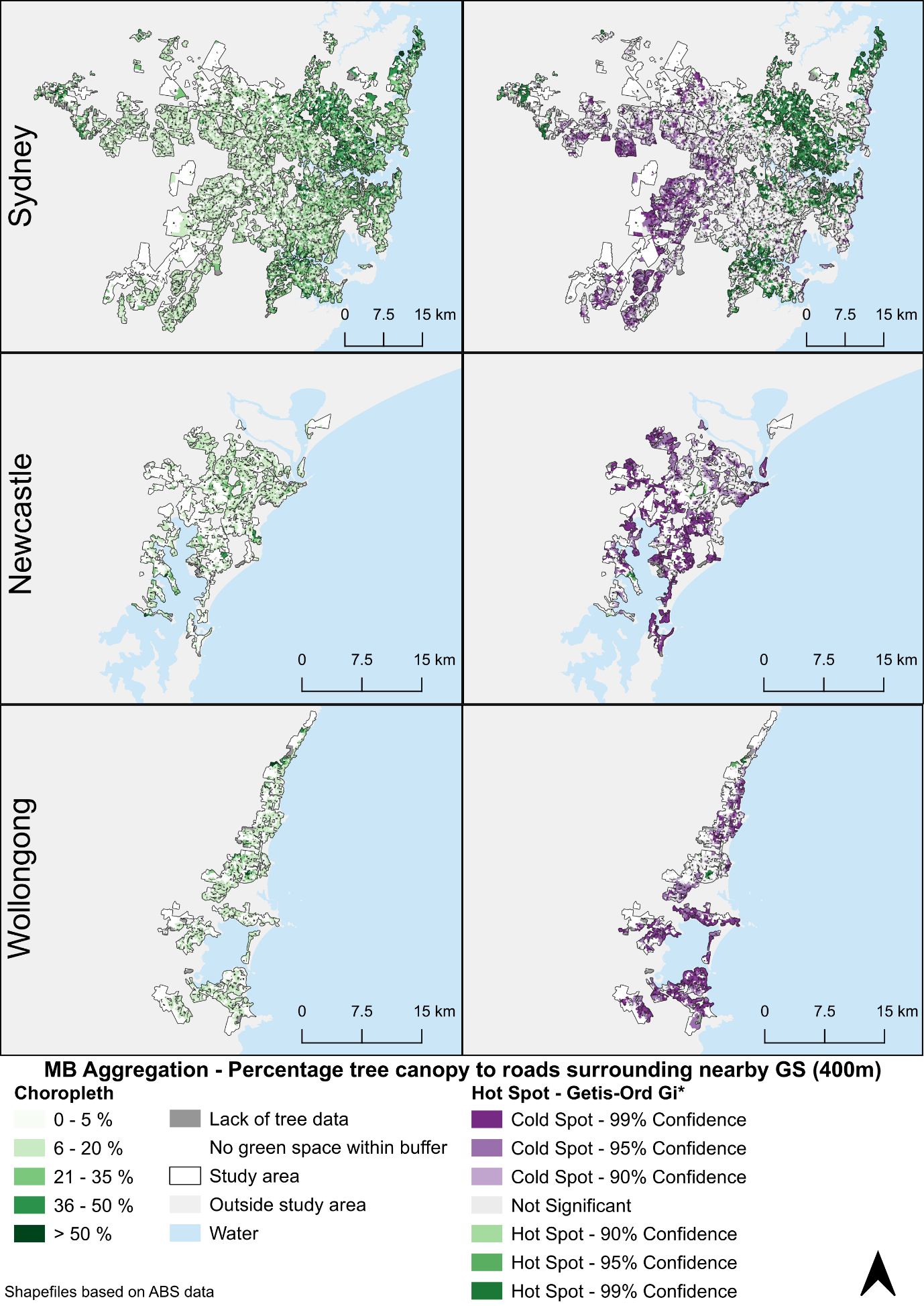

Figure S14. Percentage tree canopy to roads surrounding nearby green space (400m network distance) – Aggregation for residential MBs that contain SEIFA data. Shapefiles based on ABS data.

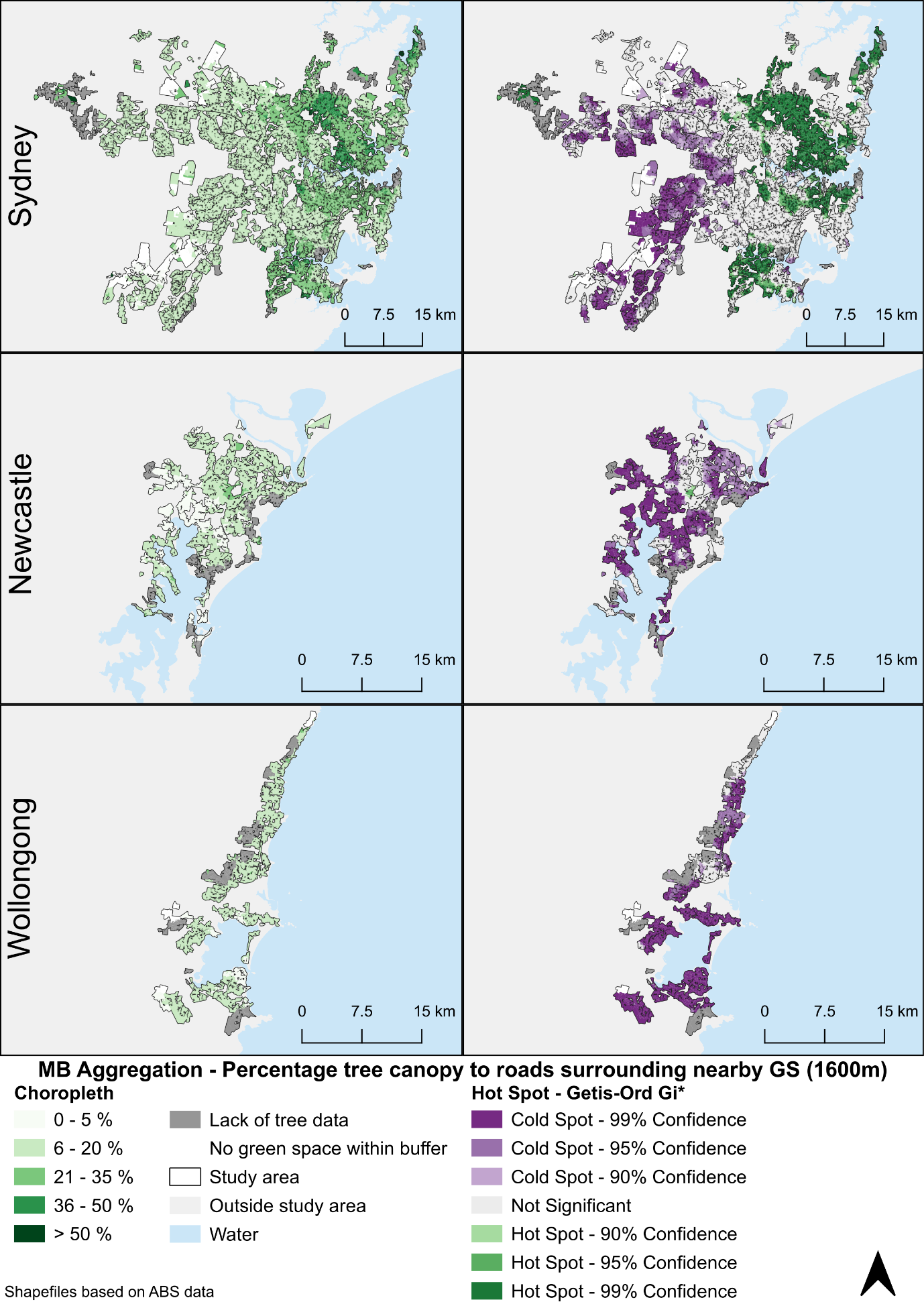

Figure S15. Percentage tree canopy to roads surrounding nearby green space (1600m network distance) – Aggregation for residential MBs that contain SEIFA data. Shapefiles based on ABS data.

### S6. Notes to tables

Notes for Table 1:

Method of aggregating intersects over mesh blocks: Count=unweighted count | Sum=unweighted sum | w0 Sum = sum weighted by the ratio of the green space intersect with the buffer area to the green space area | w1 Mean = mean weighted by the ratio of the intersect area to the sum of all intersect areas | Max = maximum value over the intersects within the buffer | Quality = Value 0/1, aggregated to 1 if any intersects equal 1 (technically Quality = Max) | Formula = calculation of the index from other qualities that have already been aggregated as weighted sums or weighted means.

P-value signs: * ≤0.001, × ≤0.01, + ≤0.05

400m: Full set: n = 39,570; Sydney: n = 34,846; Newcastle: n = 2664; Wollongong: n = 2060.

1600m: Full set: n = 47,079; Sydney: n = 40,852; Newcastle: n = 3441; Wollongong: n = 2786.

GS = green space

Notes for Table 2:

Model 2: Intercept + IRSD; Model 4: Intercept + IRSD + RE(SA2); Model 5: Intercept + IRSD + Pop (SA2)+ RE(SA2).

Qualities with some of the most extreme (rho ≥0.30 or rho ≤-0.39) coefficients of correlation were selected to represent six "concepts", trees, open grass, bare earth, slope, biodiversity and incivility. Output variables were considered to be continuous normally distributed variables, though some of them were log-normal, bi-modal and U-shaped as assessed visually.

MB=Mesh block, level 1, the lowest, n= 44620 to 47079 due to different number of missing values|| SA=Statistical area || SA2, n=362; SA3, n=48; SA4, n=16.

IRSD = Index of relative socioeconomic disadvantage; here is reversed, the higher the index, the higher the disadvantage.

IRSD is used as a categorical variable; population densities are continuous.

Trees/roads (%) = Percentage of tree canopy to roads surrounding green space (within 100m doughnut buffer); Slope >6° (%) = Percentage of green space which is greater than >6°; Trees (%) = Percentage of tree canopy within green space; Open grass (sq m) = Area of open grass within green space; Open grass (%) = Percentage of open grass within green space; Bare earth (%) = Percentage of bare earth within green space; Mean incivilities = Mean level of incivilities; Mammal species, All species = Number of species/species habitat likely to occur within green space.

Population density is per square km, converted to deciles.

DIC= Deviance information criterion, the smaller the better fit of the model.

Fits are produced using MCMC procedure in MLwiN v3.05 software with 5000-50000 burn-in and sample iterations. Confidence intervals are credible intervals.

Green shading indicates qualities negatively correlated with IRSD. Blue shading indicates qualities positively correlated with IRSD.

* P≤0.001, x P≤0.01, + P≤0.05, ^ P≤0.1.

1. Note we do not analyse levels of noise in this research, but this is used to demonstrate the method for weighted means over qualities.
2. Tree data was checked against a data set derived from GeoVision December 2020 2m landcover data (Precisely) and World Imagery basemap (Esri, Maxar, Earthstar Geographics, and the GIS User Community). Note there were many minor occurrences where there was no instance of trees in the GeoVision November 2021 data but the data set derived from GeoVision December 2020 had trees. These cases were not necessarily coded as inadequate data unless the extent of the data likely did not properly overlap the green spaces/study area.
